# Impacts on labour force and healthcare services related to mental-health issues following an acute SARS-CoV-2 infection: rapid review

**DOI:** 10.1101/2024.08.09.24311746

**Authors:** Liza Bialy, Jennifer Pillay, Sabrina Saba, Samantha Guitard, Sholeh Rahman, Maria Tan, Lisa Hartling

**Affiliations:** Alberta Research Centre for Health Evidence, Department of Pediatrics, University of Alberta Faculty of Medicine and Dentistry, Edmonton, Alberta, Canada

**Keywords:** mental health, post-COVID-19, healthcare services, labor outcomes, rapid review

## Abstract

**Purpose:** The impact on the labour force, including healthcare services, from the emergence of mental health symptoms after COVID-19 is uncertain. This rapid review examined the impacts on the labour force and healthcare services and costs related to mental health issues following an acute SARS-CoV-2 infection.

**Methods:** We searched Medline, Embase, and PsycInfo in January 2024, conducted forward citation searches in Scopus, and searched reference lists for studies reporting labour force outcomes (among those with mental health symptoms after COVID-19) and mental health services use among people of any age at least 4 weeks after confirmed/suspected SARS-CoV-2 infection. Titles/abstracts required one reviewer to include but two to exclude; we switched to single reviewer screening after 50% of citations were screened. Selection of full texts used two independent reviewers. Data extraction and risk of bias assessments by one reviewer were verified. Studies were sorted into categories based on the population and outcomes, including timing of outcome assessment, and, if suitable, study proportions were pooled using Freeman-Tukey transformation with assessment of heterogeneity using predetermined subgroups.

**Results:** 45 studies were included with 20 reporting labour force and 28 mental healthcare services use outcomes. 60% were rated as high risk of bias, mainly due to difficulty attributing the outcomes to COVID-19 from potential confounding from employment status or mental healthcare services use prior to infection. Studies on labour force outcomes mostly (85%) reported on populations with symptoms after acute infection that was cared for in outpatient/mixed care settings. Among studies reporting mental healthcare use, 50% were among those hospitalized for acute care and 43% assessed outcomes among populations with post-acute or prolonged symptoms.

Across 13 studies (N=3,106), on average 25% (95% CI 14%, 38%) of participants with symptoms after COVID-19 had mental health symptoms and were unable to work for some duration of time. It was difficult to associate inability to work with having any mental health symptom, because studies often focused on one or a couple of symptoms. The proportion of participants unable to work ranged from 4% to 71%, with heterogeneity being very high across studies (I^2^ >98%) and not explained by subgroup analyses. Most of these studies focused on people infected with pre-Omicron strains. There was scarce data to inform duration of inability to work. For outcomes related to work capacity and productivity, there was conceptual variability between studies and often only single studies reporting on an outcome among a narrowly focused mental health symptom.

On average across 21 studies (N=445,994), 10% (95% CI 6%, 14%) of participants reported seeing a mental healthcare professional of any type (psychiatrist, psychologist, or unspecified). Heterogeneity was very high and not explained after investigation. There was very limited information on the number of sessions attended. Among seven studies, mainly reporting on populations with post-COVID-19 symptoms, participant referrals to mental health services ranged from 4.2% to 45.3% for a variety of types of mental health symptoms including neuropsychology, psychiatric, and psychological. Though at high risk of bias, findings from one large study suggested 1-2% of those hospitalized during their acute infection may be re-hospitalized due to mental health symptoms attributed to COVID-19.

**Conclusions:** A large minority of people (possibly 25%) who experience persisting symptoms after COVID-19 may not be able to work for some period of time because of mental health symptoms. About 10% of people experiencing COVID-19 may have use for mental health care services after the acute phase, though this rate may be most applicable for those hospitalized for COVID-19. A small minority (possibly 1-2%) may require re-hospitalization for mental health issues. There is limited applicability of the results in most cases to populations with post-COVID-19 symptoms rather than more broadly post-COVID-19 or general populations. Overall, this rapid review highlights the variability of measurement, definition of outcomes and difficulty attributing the outcomes to mental health symptoms after COVID-19 infection.

**PROSPERO:** CRD42024504369

## BACKGROUND

The global pandemic caused by coronavirus disease 2019 (COVID-19) affected over 775 million people and resulted in more than 5 million deaths worldwide.^1^ While most people infected with SARS-CoV-2 typically recover within a few weeks,^2^ some may experience persistent symptoms lasting for several weeks or even months after the initial infection.^3, 4^ Evidence indicates varying estimates, ranging from 10% to more than 80% of infected individuals suffering from on-going symptoms for weeks or months after the initial infection, with most commonly reported symptoms being fatigue, weakness, and breathlessness among others including “brain fog”, anxiety and depression.^5–9^ Some people inflicted with symptoms after infection develop post-COVID-condition. The term “post COVID-19 condition (PCC)” was established by the WHO on October 6, 2021, to refer to new or ongoing symptoms in individuals with a history of probable or confirmed SARS-CoV-2 infection, occurring usually 3 months from the onset of the infection and lasting for at least 2 months following initial recovery that cannot be explained by an alternative diagnosis.^10^ The exact frequency and nature of PCC remains largely unknown, partly due to the variability in source populations (e.g., differing severities of acute COVID-19) and lack of a consistent definition and ascertainment criteria prior to October 2021.^11^ Estimates from general populations in Canada and the US suggests that 19%^12^ and 18-23%,^13, 14^ respectively, of adults who experience COVID-19 will develop PCC. Many other individuals will experience one or more symptoms for a shorter duration of time.

Many local governments and healthcare systems are now facing the challenge of providing services to people with PCC and the impact it has on healthcare use and resource management.^15^ There is a clear need for better understanding this emerging threat and indeed, it has been urged to prioritize research in this area.^16–18^ The economic impact of PCC is substantial for the workforce and individual ability to work is compromised for many with PCC.^19^

Mental health (MH) symptoms may affect 20-70% of infected people at 1-2 months after symptom onset.^20, 21^ In one meta-analysis, it was estimated that one fifth of recovered COVID-19 patients had psychiatric symptoms 12 months after recovering.^22^ Further, mental health symptoms may be perpetuated by other symptoms, including fatigue, shortness of breath, joint pain and chest pain. The emergence of MH symptoms after COVID-19 may impact work related outcomes, yet a previous review found no evidence to support estimates about the impact on the labour force specific to MH issues arising due to PCC.^23^ We are unaware of any synthesis on the burden on mental healthcare services after COVID-19. Because MH issues after COVID-19 may affect people not (yet/ever) diagnosed with PCC, this review focused on labour force outcomes and healthcare service use of patients experiencing MH issues at any time following the acute phase of the infection. This review was conducted for the Public Health Agency of Canada to help inform economic modeling related to MH issues following COVID-19.

## OBJECTIVE

The objective of this review was to answer the following question: What are the impacts on the (a) labour force and (b) healthcare services and costs related to mental health issues following an acute SARS-CoV-2 infection?

## METHODS

We undertook a rapid review following a protocol which was developed in collaboration with knowledge users at the Public Health Agency of Canada (PHAC) and prospectively registered on PROSPERO (CRD42024504369).

### Eligibility criteria

**Supplementary Table S1.** details our eligibility criteria. The population of interest included people of any age after confirmed (e.g., by laboratory testing) or suspected (e.g., physician diagnosed or self-reported) SARS-CoV-2 infection at least 4 weeks previous to outcome assessment. The population examined in the study could have been specific to those having new-onset/worsened or recurring MH symptoms or conditions, or broader (i.e., post-Covd-19 population, general population) so long as the outcomes of interest were examined. Primary research and modelling studies/economic analyses using secondary data sources with full texts reported in English or French were included. If reported, we included data for concurrent control groups, either without previous confirmed/suspected SARS-CoV-2 infection (ideally but not required to have negative testing at baseline and during follow-up) (“healthy control”) or with previous infection but no MH symptoms. We were interested in capturing data on how outcomes varied among several pre-defined specific populations: age (5-17 vs. 18-65 vs. 65+ years), sex, race/ethnicity, socioeconomic status (via income or if not reported educational level), reinfection status, vaccination status (0 vs. full dosing vs. 1+ boosters), virus variant (i.e. Omicron vs. earlier, differences among pre-Omicron variants), severity of acute infection, pre and/or co-existing conditions.

For labour force outcomes and costs, populations with a broad range of MH conditions were included, such as cognition, memory, neurobehavioural issues (exception of intellectual disorders), and sleep disturbances. Studies needed to include at least 30 people with MH symptoms after COVID-19 infection. Outcomes of interest included those related to economic burden (e.g., labor and healthcare costs), return to work (e.g., loss of income, proportion of individuals returning to work full time/part time/not returning, sick days, long-term disability claims), and productivity loss (e.g., presenteeism, leaveism, annual wages/income lost, impairment in the ability to perform job duties/work capacity).

For the mental healthcare services use outcome(s), we focused on those most related to common MH conditions/symptoms such as psychological counselling. Our search did not include terms related to sleep or memory providers/clinics. Any type of service provider was eligible. If all participants were identified to have a MH issue we considered including outcomes related to use of post-COVID-19/multidisciplinary clinics, assuming these were focused on the MH symptoms. Referrals to MH services were eligible to serve as a proxy for services use. Participants could have been enrolled or participated in a treatment study (that may or may not target their MH symptoms), though the effects of different treatments were not considered.

There were no limits on date, country or setting. Pre-prints were eligible as were non-peer reviewed papers and reports.

### Literature search and study selection

The search strategies were developed by a research librarian and peer-reviewed by a second librarian using the PRESS 2015 checklist.^24^ Searches were run on January 11, 2024 in MEDLINE ALL, Embase (via OVID) (EMBASE now contains preprint posted in sites such as MedRxiv) and PsycInfo (via Ebsco) using a combination of controlled vocabulary and key words for two main combinations: i) after/post - SARS-CoV-2/COVID-19, new-onset/worsening/recurring MH issues, and labour force impacts (return to work, work leave/absenteeism/productivity, lost wages/income) and ii) after/post-COVID-19 and MH services use and costs (searches located in **Supplement**). No limits were applied for date, language or study design. Vocabulary and syntax were adjusted across databases. We also performed a forward citation search in Scopus for studies included with labour force outcomes. Initially we had planned to forward search all studies included in this rapid review, but due to a one MH service use study of an international cohort^3^ retrieving over 1,200 results in Scopus, we deemed this to be unmanageable to screen for a rapid review and therefore limited the forward search to the 20 studies with labour force outcomes. The CADTH Grey Literature checklist was consulted for other governmental or professional organization websites to consider (primarily economic sites). One reviewer scanned the reference lists of the included studies and applicable systematic reviews. Results of the searches were uploaded to an EndNote (v. X9, Clarivate Analytics, Philadelphia, PA) library.

We selected records in DistillerSR using a two-step process, first by title and abstract (screening) and then by full text (selection). Using standardized forms, all reviewers involved in screening and selection piloted a random sample of records on the screening form (n=100) and selection criteria (n=25) prior to beginning each stage. During screening DistillerSR’s AI feature was utilized to re-prioritize records during screening, and we applied the liberal accelerated method whereby each title and abstract required one reviewer to include but two to exclude. Once over 50% of the citations were screened in duplicate we switched to single reviewer screening; this process has been found to enable over 95% recall of relevant studies.^25^

Any potentially relevant record from screening was retained for full-text review. For selection, two reviewers independently reviewed all full-text records and came to consensus, using input from the review leads (LB, JP) for decisions when necessary. Study authors were contacted by email with one reminder if additional information was needed to come to a final decision on inclusion. We documented the screening process using a PRISMA flow-diagram,^26^ and recorded the reason for all full-text exclusions.

### Data extraction and management

We developed standardized data extraction forms in Microsoft Office Excel (v. 2016, Microsoft Corporation, Redmond, WA) to collect relevant information from the included studies. Reviewers piloted the form on a sample of six studies and made adjustments where necessary. Following the pilot phase, one reviewer independently extracted data from the included studies, with verification for accuracy and completeness by another reviewer. Disagreements were resolved by discussion or by consulting a third reviewer. We used figures to extract data if necessary. If more than one report of a study was available, we used the report of the study’s primary outcome first and add any other missing data from associated publications.

We extracted the following information from each study: study characteristics (i.e., author, year, country, funding source, registration/protocol, design), population characteristics i.e., inclusion and exclusion criteria, sample size, population demographic information [age, sex, major comorbidities], COVID-19 confirmation method and timing [including variant if reported], re-infections, vaccinations, diagnosis of PCC [if applicable]), timing and definitions of MH issue, setting and type of care for acute phase (out-patient, hospitalization, ICU, mixed with out- and in-patients), comparator(s) and any relevant subgroup variables with their definitions, outcomes (methods/sources of data collection, timing, definitions) and data related to the outcomes and any subgroup analyses. For the purpose of this review, we defined PCC as any new or persisting symptoms present ≥12 weeks after a positive COVID-19 test or symptom onset. When the variant of infection was not reported, we attempted to define this as pre-versus post-Omicron by comparing the reported dates of infection against the Centres for Disease Control timeline (https://www.cdc.gov/museum/timeline/covid19.html). When multiple analyses were reported in comparative observational studies we extracted results from the most adjusted analysis.

### Risk of bias assessment

We assessed risk of bias by outcome (e.g., labour force and/or MH service use) for included studies using the JBI critical appraisal checklist for cohort studies.^27^ For comparative studies, we looked for adjustment related to vaccination status, age, sex, severity of infection, and reinfection status. For labour force outcomes, a high risk of bias was assigned if employment status prior to COVID-19 infection could not be clearly determined from population characteristics, or if data was collected amongst a group that was clearly not all employed pre-infection. Similarly, for MH service use outcomes if we could not determine if the service use was new after infection we allocated a high risk of bias. Before performing the assessments, reviewers piloted each tool on a sample of six studies. After piloting, one reviewer independently assessed the risk of bias for each study, with verification by another. Disagreements were resolved by discussion, or the involvement of a third reviewer.

### Data synthesis

With input from knowledge users at PHAC, studies were sorted into categories based on the population (severity of infection, source population serving as denominator [e.g., previous COVID-19, all with post-COVID symptoms, PCC, or all experiencing MH symptoms; some studies had two or more possible source populations]) and outcomes, including timing of outcome measurements. Severity of infection was presented in two categories based on the number of hospitalized participants, inpatients (≥90% hospitalized) or mix of inpatients and outpatients (<90% hospitalized). Timing of outcome measurement after COVID-19 diagnosis was classified into three categories: short (≤3 months), mid (3-6 months) and long-term (≥6 months).

Although many of the studies reporting labour outcomes had a comparator group without the specific MH symptoms (at times measured using a few select symptoms), it was not specified in many cases if they were free of all MH symptoms. Further, the studies mainly recruited populations with varied post-COVID-19 symptoms. It became apparent that control groups having no post-COVID-19 symptoms would be ideal, and because we lacked these our synthesis focused on rates among the populations with MH symptoms (though our data tables have results from within-study comparisons). For our syntheses, we used the entire study population as the denominator and for the numerator in labour force outcomes we used the number with MH symptoms experiencing the outcome, and for MH services, we used the number using/referred to the services or experiencing the outcome (e.g., hospitalization for MH issues).

We pooled proportions, using the Freeman-Tukey transformation^28^, for return to work and MH service use outcomes. For return to work we included several definitions (e.g., unable to work or sick leave) related to an inability to work and when necessary used the inverse of the event, for example, if it was presented as those having returned to work. MH services use included attendance at one or more visits to a MH professional; where the type of service use overlapped (e.g., visit to psychiatrist and visit psychologist) we used the highest number in the main analysis (but used both for our subgroup analysis by type of provider). Likewise, when the data were presented separately based on participants with potentially overlapping MH issues (e.g., depression, brain fog) we used the highest event rate rather than combining the events to avoid overestimating.

We first pooled all studies reporting on the outcome, then explored heterogeneity using subgroup/stratified analyses, as able, using the following pre-defined variables: severity of COVID-19, timing of outcome measurement, variant of concern (pre-Omicron vs. Omicron), country (OECD or US vs. not), and risk of bias (high vs. low/moderate). We added variables after data extraction, realizing some unanticipated variations in populations or outcomes. For return to work, we added variables about whether the MH symptoms captured were narrowly versus broadly defined as well as whether the study sample all had MH symptoms or not. For MH service use we also examined if there were differences by type of professional (psychologist vs. psychiatrist vs. unspecified). For other outcomes, such as those related to work capacity and productivity, there was too much conceptual variability between studies and we therefore undertook a descriptive synthesis. For this, we prioritized findings from low/moderate risk of bias studies from high-income countries with similar healthcare systems and workforce and employment systems as Canada (e.g., OECD countries and United States).

## RESULTS

We identified 4,185 records from searching databases and 434 records from other sources; 45 (1%) unique records met the eligibility criteria and were included (**Figure 1**; see **Supplement** for the lists of included and excluded studies). Three studies provided both labour force and MH service use outcomes.^29–31^ A summary of main study characteristics is included in **Table 1**. Studies originated from USA (n=11, 24%), Italy (n=5, 11%), UK (n=3, 1%), Switzerland (n=3, 1%), France (n=3, 1%), Australia (n=3, 1%), Spain (n=3, 1%), Germany (n=3, 1%), and one each from Canada, China, India, Sweden, Argentina, Ireland, Bangladesh, Japan, Netherlands, and Brazil. Almost half (n=22, 49%) assessed outcomes 6 months after a positive COVID-19 test or symptom onset; 9 (20%) assessed outcomes at 1 to ≤3 months and 14 (31%) between 3 and 6 months. The studies included a median of 261 participants (range: 30 to 388,980), with lab-confirmed COVID-19 in a majority (n=29, 64%). Across all outcomes just over half were assessed at high risk of bias (n=37, 60%) (**Supplementary Table S2**). The main reason for a high risk of bias assessment was due to potential confounding from employment status or MH services use prior to infection (i.e., difficulty attributing the outcomes to COVID-19).

**Figure 1:**
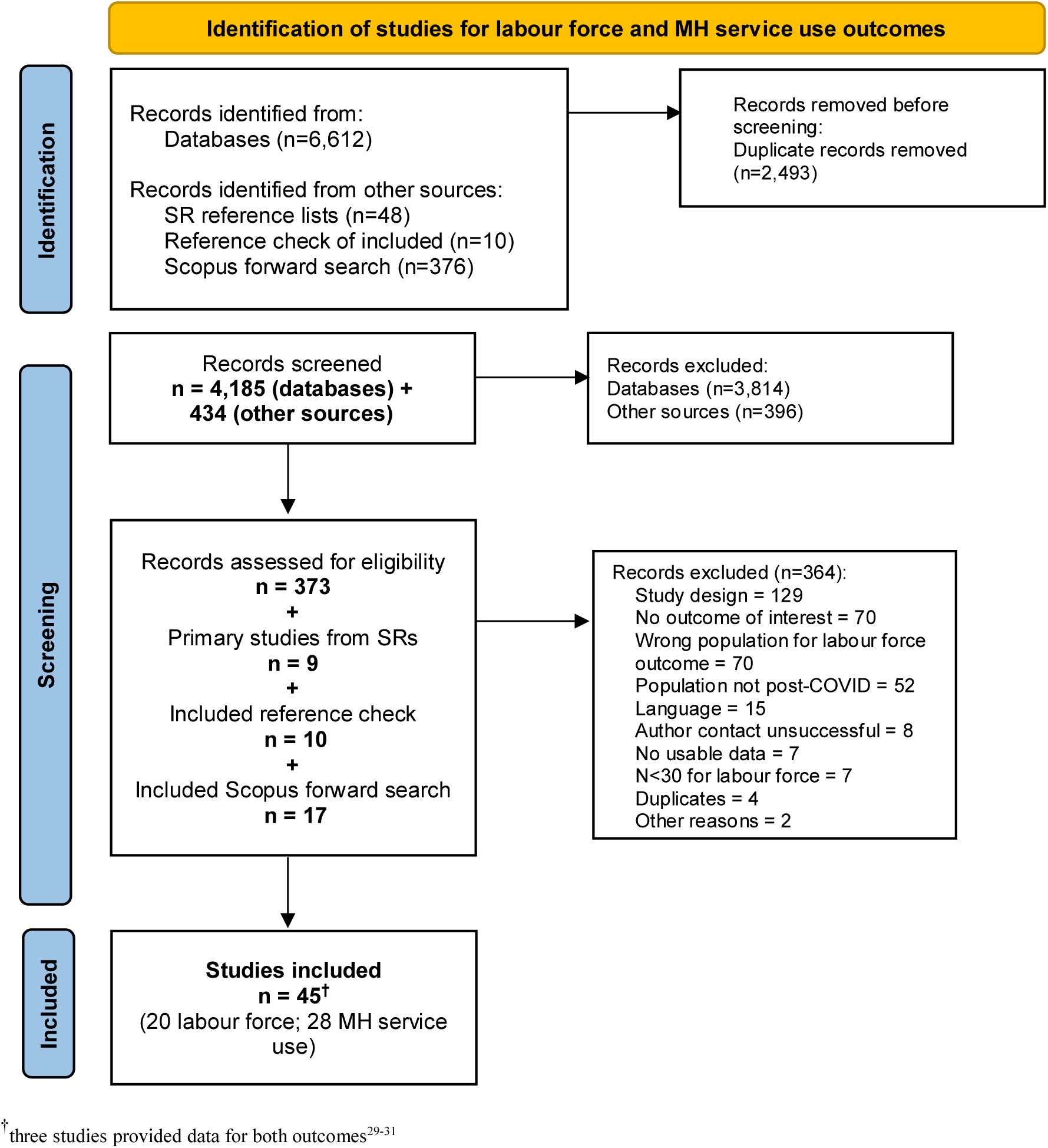
PRISMA 2020 flow diagram for impacts on labour force and healthcare services related to mental health issues following an acute SARS-CoV-2 infection.

**Table 1:** Summary of characteristics of included studies.

| Characteristics | Labour force (n=20)<sup>†</sup> | MH services (n=28)<sup>†</sup> |
| --- | --- | --- |
| **Study design (n)** |  |  |
| With control group | 12 | 3 |
| Without control group | 8 | 25 |
| **Country (n)** | USA: 5<br>UK: 4<br>Spain: 3<br>Germany: 3<br>Switzerland: 2<br>Bangladesh: 1<br>Japan: 1<br>Brazil: 1 | USA: 8<br>Italy: 5<br>France: 3<br>Australia: 3<br>Switzerland: 2<br>Canada: 1<br>China: 1<br>India: 1<br>Sweden: 1<br>Argentina: 1<br>Ireland: 1<br>Netherlands: 1 |
| **Population** | Sample size, median (range): 218 (30 to 11,710)<br>Age, median (range): 46.2 (36.6 to 53.6)<br>Female %, median (range): 68.5 (13.5 to 88.0) | Sample size, median (range): 285 (30 to 388,980)<br>Age, median (range): 53.0 (33.2 to 65.0)<br>Female %, median (range): 49.8 (12.7 to 87.0) |
| **COVID-19 ascertainment (n)** | ≥90% lab-confirmed: 11<br><90% lab confirmed: 2<br>NR: 7 | ≥90% lab-confirmed: 13<br><90% lab confirmed: 3<br>NR: 12 |
| **Severity (n)** | Inpatients (≥ 90% hospitalized): 1<br>Mix/Out (0% - 89% hospitalized): 19 | Inpatients (≥ 90% hospitalized): 14<br>Mix/Out (0% - 89% hospitalized): 14 |
| **Outcome assessment timing (n)** | Short-term (≤3 months): 3<br>Mid-term (3-6 months): 4<br>Long-term (≥6 months): 13 | Short-term (≤3 months): 6<br>Mid-term (3-6 months): 10<br>Long-term (≥6 months): 12 |
| **COVID population** | All PCC: 11<br>(100% MH symptoms = 2)<br>Post-COVID with symptoms: 6<br>(100% MH symptoms = 0)<br>COVID infected: 3<br>(100% MH symptoms = 1) | All PCC: 6<br>Post-COVID with symptoms: 6<br>COVID infected: 13<br>Mix: 3 |
| **Risk of bias** | Low/Moderate: 16<br>High: 13 | Low/Moderate: 9<br>High: 24 |

<sup>†</sup> three studies provided data for both outcomes<sup>29-31</sup>

### Labour force outcomes

Twenty studies (44%) reported labour force outcomes among post-COVID populations, with 12 (60%) comparing those with MH symptoms to those without MH symptoms or with different MH symptom clusters (**Supplementary Table 3a** and **3b**). These tables also contain any scarce within study subgroups of interest. The studies included a median of 218 participants (range: 30 to 11,710), with lab-confirmed COVID-19 in just over half of the study populations (n=11, 55%), and a large majority (n=17, 85%) having entire populations with either PCC (n=11) or post-COVID-19 symptoms (but not necessarily PCC) (n=6). Based on hospitalizations, the COVID-19 severity varied across studies with one study only including inpatients,^32^ and others ranging from 1.5% to 46.3% participants hospitalized. Ten (50%) studies reported ICU admission of patients ranging from 0.1% to 50.9%. The median age was 46.2 years (range: 36.6 to 53.6) and the median female proportion across the studies was 68.5%. A wide range of outcomes were reported with relation to labour force with most ill-defined. None of the studies included outcomes related to economic burden such as lost labour and/or healthcare costs. Outcomes were grouped into three main categories: inability to work, productivity loss, and work ability/capacity (**Table 2**).

**Table 2:** Outcome groups.

| **Labour force outcomes** | **MH services outcomes** |
| --- | --- |
| **INABILITY TO WORK**<br>- Unable to work<sup>29, 30, 33, 34, 58</sup><br>- Temporarily unable to work<sup>35</sup><br>- Taking time off beyond required quarantine period<sup>40</sup><br>- Return to work in any capacity*<sup>36</sup><br>- Sick leave<sup>39, 41, 42</sup><br>- Off work due to symptoms<sup>38</sup><br>- Return to work obstacle<sup>37</sup><br>- Mean duration incapacity to work<sup>29</sup><br>- Mean lost work days for those returning to work<sup>31</sup><br>**PRODUCTIVITY LOSS**<br>- Decrease in working hours<sup>32</sup><br>- Decrease in income<sup>32</sup><br>- Medically downgraded to perform duties<sup>43</sup><br>- Change in job description<sup>42</sup><br>- Reduced work quota<sup>34</sup><br>**WORK ABILITY / CAPACITY**<br>- Occupational performance suffered upon return to work<sup>40</sup><br>- Workability<sup>3, 45</sup><br>- At work but struggling with symptoms<sup>38</sup><br>- Experiencing significant interruptions in professional lives<sup>30</sup><br>- Work capacity and professional career influenced by MH<sup>44</sup><br>- Psychological workability<sup>37</sup><br>- Mean percentage loss of current work capacity<sup>46</sup> | **VISITS TO PSYCHIATRIST / PSYCHIATRY**<sup>29, 52, 59, 60</sup><br>**VISITS TO PSYCHOLOGIST / PSYCHOTHERAPY**<sup>29, 47, 48, 52, 61-69</sup><br>**VISIT TO OTHER MH PROFESSIONAL**<br>- Healthcare use related to MH<sup>70</sup><br>- Current MH professional use<sup>71</sup><br>- Visit to MH specialist<sup>49</sup><br>- Treatment for anxiety/depression<sup>72</sup><br>- MH clinic encounters<sup>73</sup><br>- Attended MH treatment<sup>31</sup><br>**MH HOSPITALIZATIONS**<br>- Readmitted for confusion<sup>51</sup><br>- Subsequent re-hospitalization for various MH conditions<sup>50</sup><br>- Hospitalized psychiatrically<sup>52</sup><br>**MH SERVICE REFERRALS**<br>- Referral to neuropsychology<sup>56</sup><br>- Referred to further psychiatric consultation<sup>54</sup><br>- Referred for psychologist consultation<sup>48</sup><br>- Referrals to psychology<sup>55</sup><br>- Referral to psychiatrists<sup>53</sup><br>- Recommended psychotherapeutic treatment<sup>30</sup><br>- Referred to MH treatment<sup>31</sup> |

\* events inverted for analysis

#### Inability to work

The outcomes comprising inability to work included taking time off, current sick leave and off work due to symptoms. (**Table 3**) Thirteen studies were analyzed with all including a mix of inpatient and outpatient populations ranging from 7% to 46% hospitalized. Timing since COVID-19 ranged from ≤3 months (n=2), 3-6 months (n=3), to ≥6 months (n=8). An assortment of MH symptoms was captured across studies, including cognitive dysfunction,^33–37^ brain fog,^38^ sleep disturbances,^29, 36^ PTSD,^39^ depression^40^ and three studies presenting a broad spectrum of MH symptom clusters.^30, 41, 42^ Just over half of the outcomes were low/moderate risk of bias (n=7, 54%).

**Table 3:** Labour Force – Inability to work by COVID-19 timing and severity.

| OUTCOME | OUTCOME DESCRIPTION | SEVERITY | MH SYMPTOMS | EVENT RATE |
| --- | --- | --- | --- | --- |
| **COVID TIMING: Short (<=3 months)** |  |  |  |  |
| **SEVERITY: Mix inpatient and outpatient (<90% hospitalized)** |  |  |  |  |
| Danesh, 2021<sup>33</sup><br>Unable to work<br>Control: outcome in those without cognitive dysfunction<br>RoB: High | Unable to return to work due to the severity and persistence of sequelae | ICU: NR<br>Home O2: 13%<br>Hospitalized: 30% | Cognitive dysfunction | With MH = 4.5% (9/200)<br>Without MH = 5.5% (11/200) |
| Thompson, 2023<sup>40</sup><br>Taking time off beyond required quarantine period<br>Control: without MH symptoms<br>RoB: Low/Moderate | NR | ICU: NR<br>Hospitalized: 6.7% | Clinically significant depression, determined by scoring ≥10 on the Patient Health Questionnaire (PHQ-9) | With MH = 37.3% (22/59)<br>Without MH = 10.2% (6/59) |
| **COVID TIMING: Mid (3-6 months)** |  |  |  |  |
| **SEVERITY: Mix inpatient and outpatient (<90% hospitalized)** |  |  |  |  |
| Lemhofer, 2023<sup>58</sup><br>Unable to work<br>Control: Without MH symptoms<br>RoB: Low/Moderate | NR | ICU: NR<br>Hospitalized: 32.6% | Deficits in the mental component summary score of the SF-36 | With MH = 23.4% (74/316)<br>Without MH = 16.5% (52/316) |
| Romero-Rodrigue, 2023<sup>39</sup><br>Some sick leave<br>Control: without MH symptoms<br>RoB: High | Some sick leave due to PASC. Other option was no sick leave. | ICU: 3.6%<br>Hospitalized: 23% | Mental confusion and PTSD as part of persistent COVID signs and symptoms | With mental confusion = 47.8% (285/596)<br>Without mental confusion = 30.5% (182/596)<br>With PTSD = 20.1% (120/596)<br>Without PTSD = 58.2% (347/596) |
| Vanichkachorn, 2021<sup>36</sup><br>Return to work in any capacity<br>Control: Without MH symptoms<br>RoB: High | Included reduced hours or accommodated functional activities, at their usual work site or an alternative location such as home | ICU: NR<br>Hospitalized: 25% | Cognitive symptoms defined as any subjective reports of concentration difficulty, memory loss, word finding difficulty, and inability to effectively multitask.<br>Presenting symptom of sleep disturbance. | With cognitive impairment = 29% (29/100)<br>Without cognitive impairment = 34% (34/100)<br>With sleep disturbance = 18% (18/100)<br>Without sleep disturbance = 45% (45/100) |
| **COVID TIMING: Long (>=6 months)** |  |  |  |  |
| **SEVERITY: Mix inpatient and outpatient (<90% hospitalized)** |  |  |  |  |
| Garcia-Molina, 2022<sup>35</sup><br>Temporarily unable to work<br>Control: without cognitive alterations<br>RoB: Low/Moderate | If patient working: Has the patient returned to work as normal? (yes/no) Follow-up questions: Does the patient work in the same conditions as before COVID-19? | ICU: 50.9%<br>Hospitalized: 46.3% | Cognitive alterations | With MH = 34.8% (24/69)<br>Without MH = 10.1% (7/69) |
| Green, 2023<sup>38</sup><br>Off work due to symptoms<br>Control: Without MH symptoms<br>RoB: High | NR | ICU (not intubated): 1%<br>ICU (intubated): 2%<br>Hospitalized: 12% | Brain fog | With MH = 25.7% (55/214)<br>Without MH = 9.3% (20/214) |
| Diem, 2022<sup>29</sup><br>Incapacity to work<br>Control: outcome in those without cognitive dysfunction<br>RoB: Low/Moderate | NR | ICU (with intubation): 2%<br>ICU without intubation: 2.6%<br>Hospitalized: 10.7% | Insomnia and excessive daytime sleepiness (EDS) | With insomnia = 29.2% (69/236)<br>Without insomnia = 39.4% (93/236)<br>With EDS = 41.2% (91/221)<br>Without EDS = 28.1% (62/221) |
| Diem, 2022<sup>29</sup><br>Mean duration incapacity to work<br>Control: outcome in those without cognitive dysfunction<br>RoB: Low/Moderate | NR | ICU (with intubation): 2%<br>ICU without intubation: 2.6%<br>Hospitalized: 10.7% | Insomnia and excessive daytime sleepiness (EDS) | Among those with vs. without insomnia<br>29.7 (24.3-35.1) weeks; n=69 vs. 25.2 (21.5-30.0) weeks; n=93<br>Among those with vs. without EDS<br>26.3 (22.4-30.1); n=91 vs. 27.9 (22.6-33.4); n=62 |
| Tsuchida, 2023<sup>42</sup><br>Current employment status:<br>Leave of absence<br>Control: without MH symptoms (clusters 1, 2, 4, and 5)<br>RoB: High | • Patient visiting outpatient clinic | ICU: NR<br>Hospitalized: NR | Post-COVID psychological symptom cluster 3 (psychological symptoms, including anxiety, depressed mood, forgetfulness, insomnia in addition to fatigue, headache, and lack of motivation) | With MH = 8.3% (41/496)<br>Without MH = 14.7% (73/496) |
| Farooqi, 2022<sup>30</sup><br>Unable to return to work<br>Control: None<br>RoB: Low/Moderate | • NR | Significant illness, intubated: 3.3%<br>Hospitalized: 36.7% | Neuropsychiatric symptoms (anxiety, depression, fatigue, and cognitive problems) | With MH = 20% (6/30) |
| Braga, 2022<sup>41</sup><br>Sick leave<br>Control: None<br>RoB: Low/Moderate | • From outpatient hospital unit | ICU: 17.3%<br>Hospitalized: 33.3% | Treatment for cognitive issues (attention, concentration, and memory difficulties, slow reasoning, black-outs, etc.) | With MH = 4.2% (26/614) |
| Lunt, 2022<sup>37</sup><br>Return to work obstacle<br>Control: None<br>RoB: Low/Moderate | • Most frequently portrayed as obstacles at the individual level. The relapsing nature of symptoms was also widely attributed to hampering 'return-to-work planning' | ICU: NR<br>Hospitalized: 12% | Poor concentration<br>Symptom interaction with cognitive job demands of a given job | With poor concentration= 13.8% (20/145)<br>With symptom interaction = 8.3% (12/145) |
| Dressing, 2022<sup>34</sup><br>Inability to work<br>Control: None<br>RoB: Low/Moderate | • Inability to work and restriction of daily life activities | Endotracheal ventilation at ICU: 13%<br>Hospitalized: NR | All admitted to outpatient clinic due to lasting neurocognitive symptoms | With MH = 29% (9/31) |
| LeGoff, 2023<sup>31</sup> [1135]<br>Mean lost work days of those RTW<br>Control: None<br>RoB: High | • RTW at time of initial referral with or without restrictions, since being placed on leave by their primary treating physicians. All were determined to be temporarily disabled | ICU: NR<br>Hospitalized: NR | "Brain fog" including mental fatigue, forgetfulness/impaired short-term memory, difficulties concentrating, inattentiveness, diminished task focus, confusion were affecting returned to work at time of initial referral | 459.4 days<br>(15 months: from 19 weeks to 2.5 yrs [range 134-903 days [SD 126.6 days]]; median 425.5 days) |

Overall, it was difficult to associate inability to work with having *any* MH symptom; in some studies only a single or narrow range of symptoms were measured (e.g., depression, anxiety, brain fog, etc.) with no accounting for other MH symptoms that could have been present. Moreover, the populations recruited for these studies where generally highly symptomatic with other indicators (e.g., respiratory, gastrointestinal, etc.) commonly seen in those with post COVID-19 sequelae. The entire sample in two studies^34, 41^ recruited populations reporting one or more MH symptoms. One study^36^ provided numbers for those returning to work so these were inversed to be used in the analysis.

When pooling proportions across all 13 studies (N=3,106), 25% (95% CI 14%, 38%) of participants with symptoms post COVID-19 had MH symptoms and were unable to work for some duration of time (**Figure 2**). Nine studies (69%) in this analysis focused on participants with PCC, four determined by standardized criteria (WHO and NICE), four by symptom evaluation at ≥12 weeks, and one by means unreported. The proportion of participants unable to work ranged from 4% to 71%, with heterogeneity being very high across studies (I^2^ > 98%). Proportions in six studies fell within the overall CI. The high variability among these studies was not explained (i.e. heterogeneity remained high in each strata) when looking at various subgroups including timing since infection (**Figure 3**), risk of bias alone (**Supplementary Figure S1**) and among the long-term follow-up studies (**Supplementary Figure S2**), study samples with only MH symptoms versus not (**Supplementary Figure S3**), and study use of a narrow versus broad spectrum of MH symptoms (**Supplementary Figure S4**). With 12 of the 13 studies taking place in OECD countries we did not look at subgroups by country. Similarly, 12 studies included participants infected during the Alpha/Delta variant phase.

**Figure 2:**
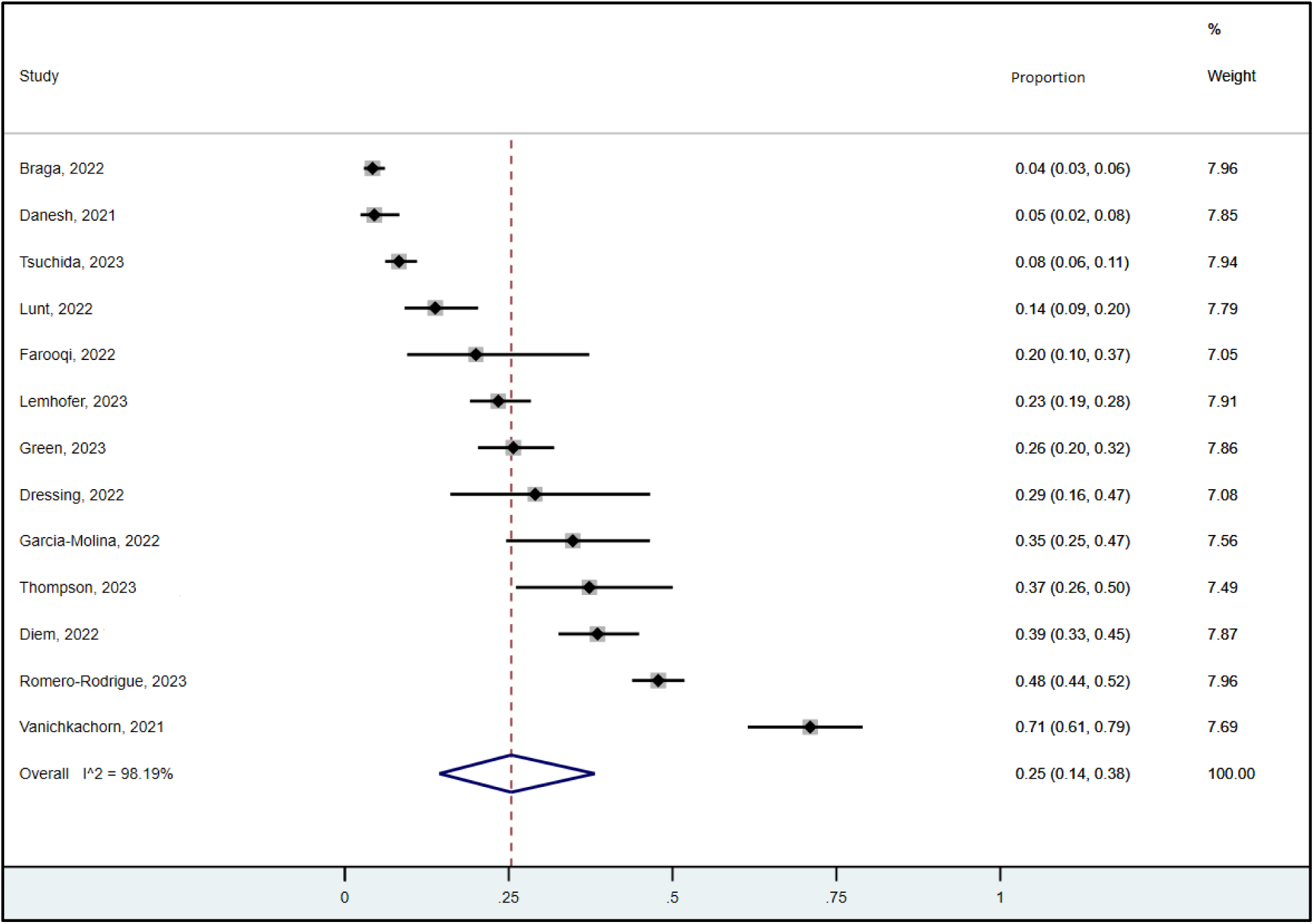
Inability to work (labour force), all studies.

**Figure 3:**
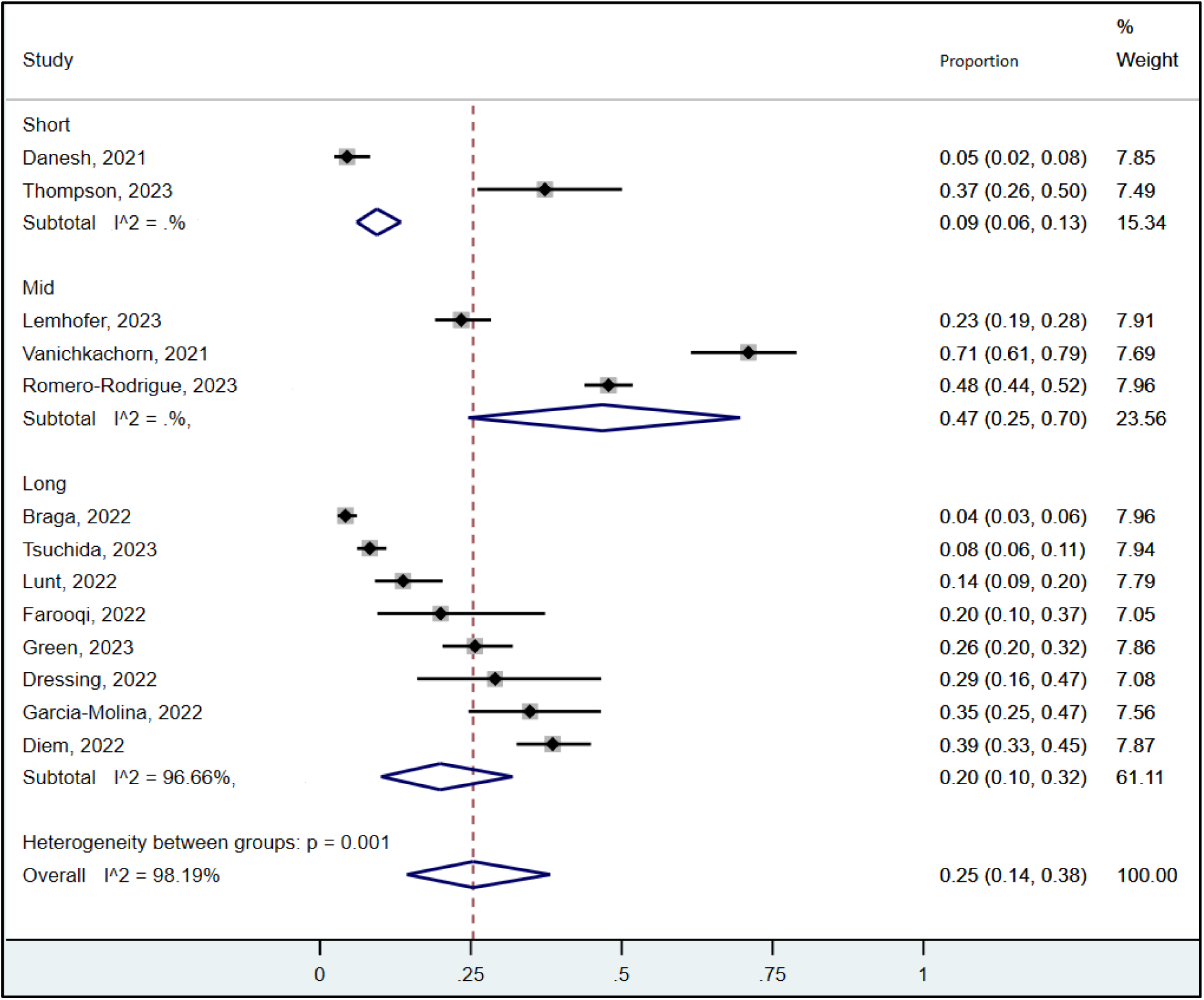
Inability to work, subgroups on timing of outcome measurement (labour force)

Two studies provided data about duration of inability to work.^29, 31^ Among a sample of 309 people with PCC at 13 months post-COVID-19, the mean duration of incapacity to work was 29.7 weeks for those with insomnia compared to 25.5 weeks for those without, and was 26.3 weeks for those with excessive daytime sleepiness compared to 27.9 weeks for those without this symptom.^29^ In the other study, authors enrolled 64 essential frontline workers referred by their primary treating occupational physicians to an outpatient mental health provider panel for assessment and treatment due to their being off work. During follow-up, these participants had a mean duration of work leave of 15 months (range 134 to 903 days), between a doctor’s first report of illness to the date of either full duty or modified duty release to work.^31^

#### Productivity loss

Productivity loss outcomes included decreases in working hours or income, reduced work quota, and changes in job description (**Table 4**). Four studies were included with one consisting of only inpatients^32^ and others a combination of in and outpatients.^34, 42, 43^ With the exception of one study^32^ outcomes were assessed 6 months after infection. One study included an exclusively symptomatic population with 31 participants having neurocognitive issues.^34^ The remaining studies included 45% of participants with self-described depression, anxiety or stress,^32^ 6% with cognitive impairments,^43^ and 42% with a cluster of various psychological symptoms.^42^

**Table 4:** Labour Force – Productivity loss by COVID-19 timing and severity.

| STUDY / OUTCOME | OUTCOME DESCRIPTION | SEVERITY | MH SYMPTOMS | FINDINGS |
| --- | --- | --- | --- | --- |
| **COVID TIMING: Mid (3-6 months)** |  |  |  |  |
| **SEVERITY: Inpatient (≥ 90% hospitalized)** |  |  |  |  |
| Anik, 2023<sup>32</sup><br>Decrease in working hours<br>Control: without MH symptoms<br>RoB: High | Changes in working hours having increased or decreased since being discharged from the hospital | ICU: NR<br>Hospitalized: 100% | Depression, anxiety and/or stress | With MH = 14.6% (70/481)<br>Without MH = 10.8% (52/481) |
| Anik, 2023<sup>32</sup><br>Decrease in income<br>Control: without MH symptoms<br>RoB: High | Those who reported changes in their income and expenditure | ICU: NR<br>Hospitalized: 100% | Depression, anxiety and/or stress | With MH = 23.9% (115/481)<br>Without MH = 24.1% (116/481) |
| **COVID TIMING: Long (≥6 months)** |  |  |  |  |
| **SEVERITY: Mix inpatient and outpatient (<90% hospitalized)** |  |  |  |  |
| Dressing, 2022<sup>34</sup><br>Reduced work quota<br>Control: None<br>RoB: Low/Moderate | Work quota defined as reduction of work quota necessary | Endotracheal ventilation at ICU: 13%<br>Hospitalized: NR | All admitted to outpatient clinic due to lasting neurocognitive symptoms | With MH = 9.7% (3/31) |
| O'Sullivan, 2023<sup>43</sup><br>Medically downgraded to perform duties<br>Control: without MH symptoms<br>RoB: Low/Moderate | Based on medical deployment status to determine ability to perform job role without restrictions (fully deployable) or with limitations (medically downgraded) | ICU: NR<br>Hospitalized: 36.9% | Cognitive impairment based on presence of 'any' cognitive symptom<br>Individuals medically downgraded to perform duties due to MH conditions for poor memory, concentration, attention and confusion | Cognitive symptoms<br>With = 36.0% (80/222)<br>Without = 34.2% (76/222)<br>Poor memory<br>With = 20.7% (46/222)<br>Without = 49.5% (110/222)<br>Poor concentration<br>With = 24.8% (55/222)<br>Without = 45.4% (101/222)<br>Poor attention<br>With = 31.5% (70/222)<br>Without = 38.7% (86/222)<br>Confusion<br>With = 21.6% (48/222)<br>Without = 48.6% (108/222) |
| Tsuchida, 2023<sup>42</sup><br>Current employment status:<br>Change in job description<br>Control: without MH symptoms (clusters 1, 2, 4, and 5)<br>RoB: High | • Patient visiting outpatient clinic | ICU: NR<br>Hospitalized: NR | Post-COVID psychological symptom cluster 3 (psychological symptoms, including anxiety, depressed mood, forgetfulness, insomnia in addition to fatigue, headache, and lack of motivation) | With MH = 2.0% (10/496)<br>With other symptoms = 9.3% (46/496) |

One study^43^ with a low/moderate risk of bias described the medical downgrading of USA military personnel, mean age 37 years, due to various cognitive symptoms.^43^ All sampled employees (n=222) had ongoing symptomatic or initially severe COVID-19 infection. Among those with any cognitive symptom (n=97) the number medically downgraded was very similar to those downgraded without cognitive symptoms, but potentially with other non-cognitive MH symptoms (36% vs. 34%, respectively). Interestingly when poor memory, concentration, attention and confusion were assessed separately, those without symptoms were more likely to be medically downgraded. The other study in this group with low/moderate risk of bias found that 10% of participants (n=31) admitted to an outpatient clinic due to lasting neurocognitive symptoms had a reduced work quota.^34^

#### Work ability/capacity

Eight studies included work ability/capacity outcomes,^3, 30, 37, 38, 40, 44–46^ among those having a range of MH symptoms such as cognitive dysfunction, depression, neurocognitive and brain fog (**Table 5**). Most of the studies (n=6, 75%) included those with both inpatient and outpatient populations ranging from 7% to 37% hospitalized,^3, 30, 37, 38, 40, 44^ and two studies (25%) included only non-hospitalized participants.^45, 46^ The majority (n=6, 75%) of studies assessed outcomes at 6 months or longer after COVID-19 infection,^30, 37, 38, 44–46^ with the remaining studies measuring at 1 to ≤3 months.^3, 40^ More than half of the outcomes were assessed as low/moderate risk of bias (n=5, 63%).

**Table 5:**
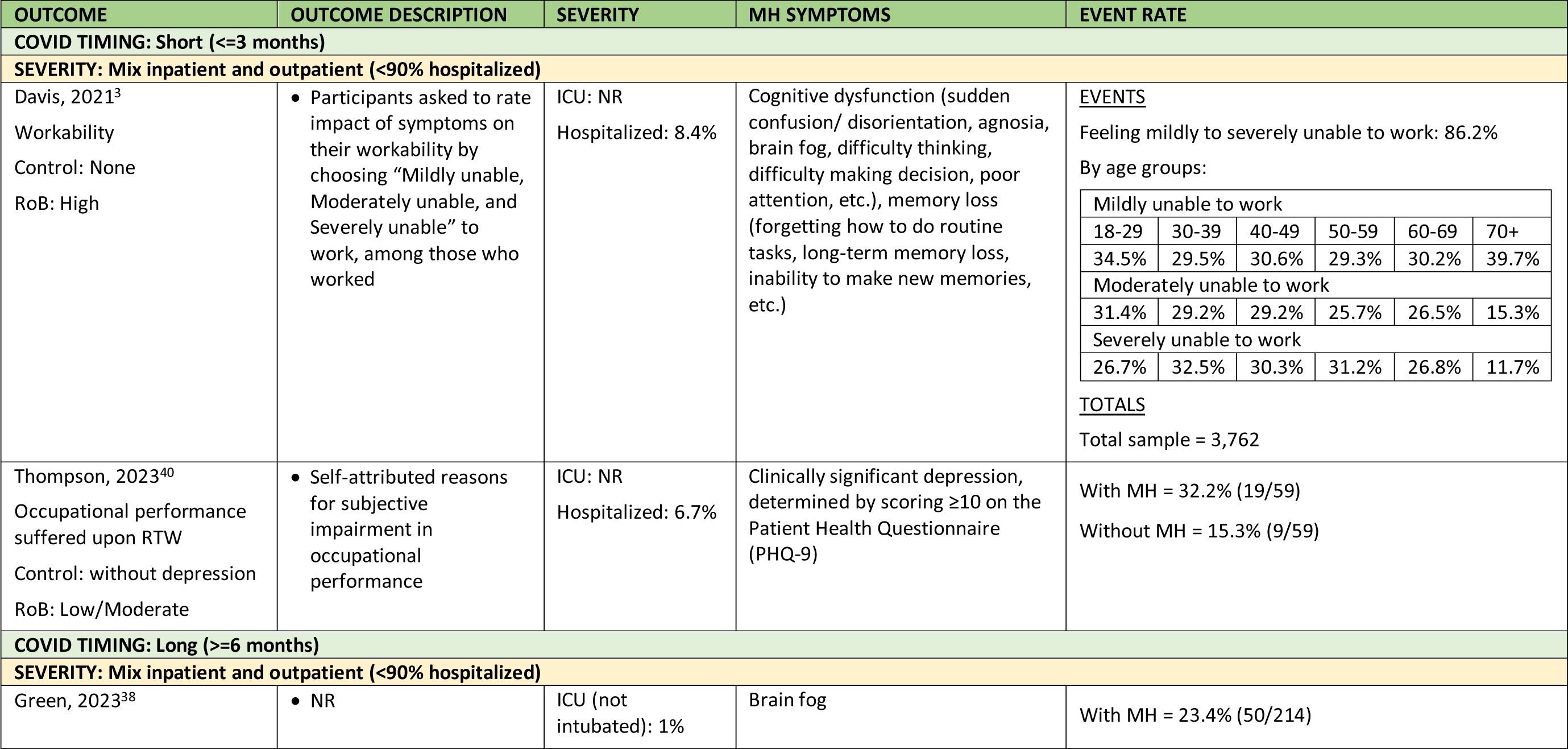
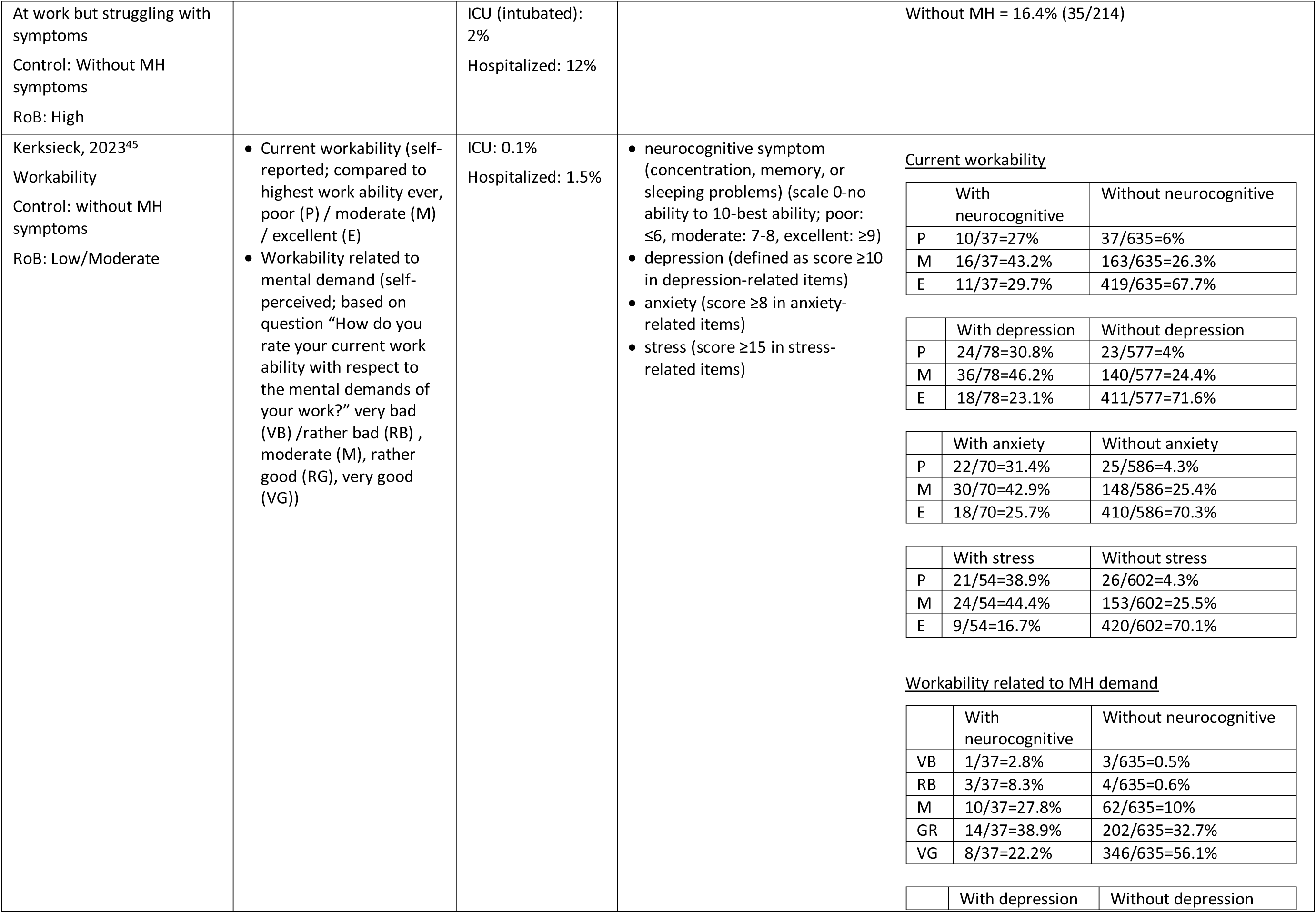
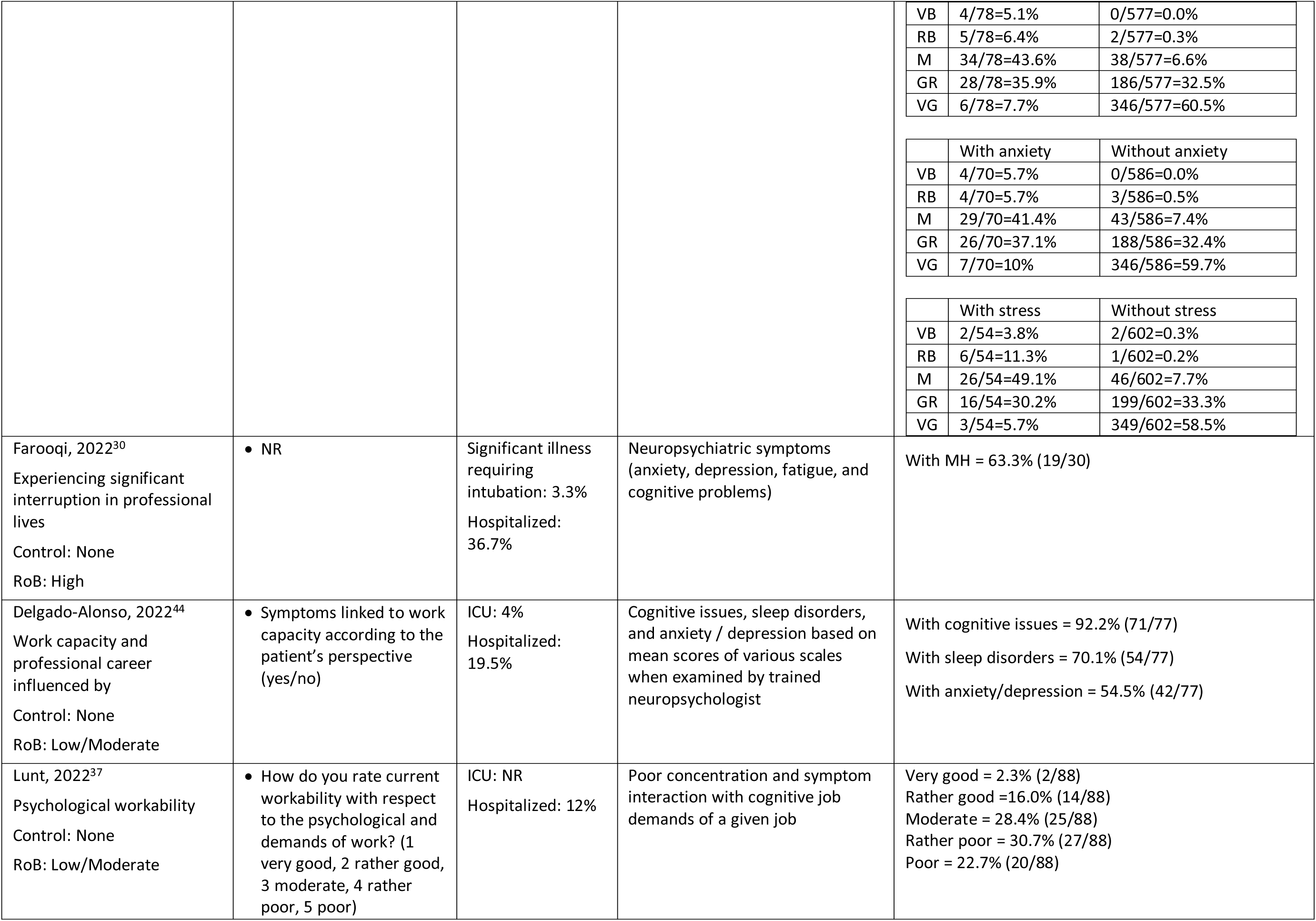
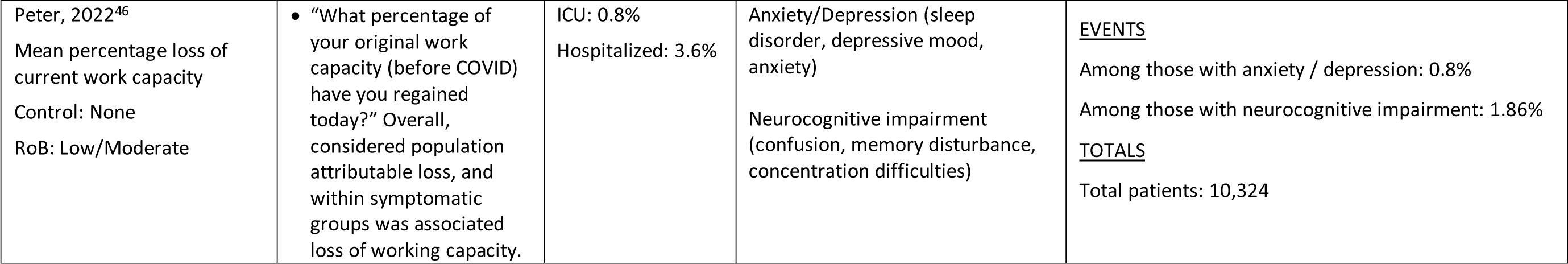
Labour Force – Work ability/capacity by COVID-19 timing and severity.

Three studies measured workability on different Linkert scales. One study assessed at low/moderate risk of bias found that among a general post-COVID-19 population (n=672; 18% reporting one or more post-COVID symptoms), few reported poor workability with neurocognitive (1.5%), depressive (3.7%), anxiety (3.3%), or stress symptoms (3.2%).^45^ Despite this, there were higher proportions with poor workability among those with (27-39%) versus without (4-6%) these symptoms. Similarly, workability with respect to mental demands was rated very bad or rather bad more often for those with the MH symptoms (about 11% for each symptom) versus without (about 1%). Another study at low/moderate risk of bias measured psychological workability among 145 workers with previous (majority more than 6 months) COVID-19, of which 53% reported this was rather poor or poor.^37^ Further, 14% reported poor concentration leading to obstacles. In one high risk of bias study, 86% of working survey participants with post-COVID-19 symptoms felt mildly to severely unable to work due to memory or cognitive dysfunction.^3^

Three studies included participants with PCC.^38, 40, 44^ One study (n=59) with low/moderate risk of bias measuring clinically significant depression (scoring ≥10 on the Patient Health Questionnaire) among people employed before COVID-19 found that 33 (60%) had clinical depression and among these 19 (32% of study population) suffered occupational performance upon return to work; 9 (15%) of the study population suffered occupational performance but not clinical depression.^40^ Another study (n=77) with low/moderate risk of bias including people actively working prior to COVID-19 diagnosis observed that most participants (92%) had cognitive issues influencing their work capacity and professional career.^44^ The remaining study with high risk of bias found that 23% (50 of 214) of participants had brain fog and were at work but struggling with symptoms compared to 16% struggling but without brain fog.^38^

One large population-based study of over 11,000 participants with lab-confirmed COVID-19 measured work capacity (i.e., % of original work capacity that had been regained after infection; 10-point scale from 0% to 100%) and estimated the population attributable loss in work capacity associated with 13 symptom clusters (% loss multiplied by the symptom cluster’s prevalence).^46^ There was a 10.7% overall loss of work capacity, with less than 1% (0.8%) attributed to anxiety/depression and 1.86% to neurocognitive impairment.

### Mental health services use

Twenty-eight studies (62%) reported outcomes related to MH service use after COVID-19 infections with only three (11%) of these providing a comparison to populations without COVID-19 (**Supplementary Table 4a** and **4b**). The studies included a median of 285 participants (range: 30 to 388,980), with lab-confirmed COVID-19 in just under half of the studies (n=13, 46%). Six (21%) studies examined populations having PCC (3 confirming PCC through validated guidelines, two by symptom evaluation, and one through neurocognitive screening) and in another six the population had post-COVID-19 symptoms. Half of the studies reporting these outcomes focused on inpatient populations (n=14, 50%), and the other half reporting between 0% and 87% hospitalized. The median age was 53.0 years (range: 33.2 to 65.0) and the median female proportion across the studies was 49.8%. A wide range of MH services were reported and grouped into three main categories, with pooled proportions among all and for each: visits to psychiatrists, psychologists, and unspecified MH professionals (**Table 6**). Outcomes for MH hospitalizations and referrals have been described separately (**Table 7** and **8**).

**Table 6:** Mental Health Services – Utilization of professional mental health services by COVID-19 timing and severity.

| OUTCOME | OUTCOME DESCRIPTION | SEVERITY | POPULATION DESCRIPTION | EVENT RATE |
| --- | --- | --- | --- | --- |
| **COVID TIMING: Short (<=3 months)** |  |  |  |  |
| **SEVERITY: Inpatient (≥ 90% hospitalized)** |  |  |  |  |
| Benzakour, 2021<sup>60</sup><br>Psychiatric follow-up<br>Control: None<br>RoB: High | Psychiatrist provided intervention depending on patients' mental status at assessment. Patients whose questionnaire scores exceeded established cut-off were referred to a psychiatric consultation by liaison psychiatry team | ICU: 16.5%<br>Hospitalized: 100% | Hospitalized for COVID and attending CoviCare program (a multidisciplinary strategy, including specialists in internal medicine, pulmonary disease, primary care, infectious diseases and psychiatry) | With MH service use = 23.9% (26/109) |
| Bonazza, 2020<sup>61</sup><br>Psychological service use<br>Control: None<br>RoB: Low/Moderate | Psychologist service use, patients decided to begin a cycle of 8 other sessions with the hospital psychologist | ICU: 7.5%<br>Hospitalized: 100% | Recovered COVID patients previously hospitalized and offered a multidisciplinary follow-up screening program within 2 months | With MH service use = 6.1% (16/261) |
| Chopra, 2021<sup>70</sup><br>Health care use related to MH<br>Control: None<br>RoB: High | NR | ICU: 13.2%<br>Hospitalized: 100% | Hospitalized for COVID | With MH service use = 2.2% (28/1250) |
| **SEVERITY: Mix inpatient and outpatient (<90% hospitalized)** |  |  |  |  |
| Tajer, 2023<sup>67</sup> | Required new consultation for psychotherapy | ICU: NR | Latin-American health professionals with COVID; some reported that the | With MH service use = 14.4% (672/4673) |
| Requirement for psychotherapy<br>Control: None<br>RoB: High |  | Hospitalized: 11.7% | symptoms had not lasted more than one month since COVID episode |  |
| Van Wambeke, 2023<sup>69</sup><br>Psychological support therapy attendance<br>Control: None<br>RoB: High | Patients contacted the post-COVID coordination team and received phone consultations with an advanced practice nurse (45-65 minutes), who proposed therapeutic follow-up focused on patients' main symptoms. Outcome reported as those who had the opportunity to benefit from the psychological support therapy | ICU: NR<br>Hospitalized: 0% | COVID patients receiving consultations with study team, including follow-up through the first and second phone call. | With MH service use = 24.4% (11/45) |
| **COVID TIMING: Mid (3-6 months)** |  |  |  |  |
| **SEVERITY: Inpatient (≥ 90% hospitalized)** |  |  |  |  |
| Lazzaroni, 2022<sup>48</sup><br>Psychologist consultation<br>Control: None<br>RoB: Low/Moderate | Psychologist consultation; 4-6 visits among patients after COVID (previously hospitalized) that screened positive for PTSD or depression<br>Trauma-focused psychological support intervention was proposed to patients with positive scores aimed to facilitating the resumption of the daily life by reducing the impact of this recent traumatic experience | ICU: 32%<br>Hospitalized: 100% | Previously hospitalized with COVID, attending a COVID outpatient clinic after hospital discharge | With MH service use = 11.1% (58/523) |
| Bek, 2023<sup>47</sup><br>Psychologist consultation<br>Control: None<br>RoB: Low/Moderate | Attended psychologist session for anxiety or depression | ICU: 41%<br>Hospitalized: 100% | Patients attending the COVID follow-up care path (CO-FLOW) after hospital discharge | With MH service use = 11.5% (71/617) |
| Ahmad, 2022<sup>59</sup><br>Psychiatry service use<br>Control: None | At the post-COVID clinic when presenting with depressed mood, hopelessness, anxiety, and/or fatigue | ICU: 7.9%<br>Hospitalized: 87.7% | COVID positive patients attending post-COVID clinic for initial evaluation of long-term sequelae, | With MH service use = 1.5% (22/1511) |
| RoB: High |  |  | early rehabilitation, and targeted specialist referrals |  |
| **SEVERITY: Mix inpatient and outpatient (<90% hospitalized)** |  |  |  |  |
| Staples, 2023<sup>71</sup><br>Current MH professional use<br>Control: without COVID completing assessment at outpatient service<br>RoB: Low/Moderate | NR | ICU: NR<br>Hospitalized: NR | Patient with assessment at MindSpot clinic, a national digital MH service | With COVID = 20% (1862/9341)<br>Without COVID = 14.1% (1319/9341) |
| Formoso, 2023<sup>49</sup><br>Visit to MH specialist<br>Control: without COVID; matched from local health authority database with equal distribution of sex, age, and comorbidities<br>RoB: High | Outpatient MH specialist visits (e.g., anxiety, depression, sleep problems) | ICU: NR<br>Hospitalized: 2% | Diagnosed with COVID using outpatient service use | With COVID, at least one visit = 0.5% (174/36036)<br>Without COVID, at least one visit = 0.5% (175/36036)<br>With COVID, total first visits = 0.3% (117/36036)<br>Without COVID, total first visits = 0.3% (104/36036) |
| Brehon, 2022<sup>62</sup><br>Receiving psychology services<br>Control: None<br>RoB: High | Prior to beginning the post-COVID rehabilitation program | ICU: NR<br>Hospitalized: 17% | Workers discharged from post-COVID occupational rehabilitation program | With MH service use = 32% (26/81) |
| **COVID TIMING: Long (>=6 months)** |  |  |  |  |
| **SEVERITY: Inpatient (≥ 90% hospitalized)** |  |  |  |  |
| Frontera, 2022 [640]<br>Psychological talk therapy<br>Control: None<br>RoB: Low/Moderate | to deal with prolonged COVID symptoms | ICU: NR<br>Invasive mechanical ventilation: 34%<br>Hospitalized: 100% | Hospitalized with COVID | With MH service use = 4.5% (11/242) |
| Rival, 2023<sup>66</sup><br>Psychological follow-up<br>Control: None<br>RoB: High | Psychological or psychiatric consultation was offered to patients with a positive Post-Traumatic Stress Checklist-5 (PCL-5) screening result (score<br>of ≥33/80 or 4/4 positive clusters) | ICU: 45%<br>Hospitalized: 100% | Hospitalized with COVID | With MH service use = 15% (6/40) |
| Umbrello, 2022<sup>68</sup><br>Need for psychologist<br>Control: None<br>RoB: High | based on information on health service utilization and need for care after ICU discharge | ICU: 100%<br>Hospitalized: 100% | Italian-speaking patients admitted to ICU | With MH service use = 13.9% (11/79) |
| Huang, 2022<sup>64</sup><br>Use of psychological help<br>Control: None<br>RoB: Low/Moderate | Accessed after discharge | ICU: 4%<br>Hospitalized: 100% | COVID survivors who were previously hospitalized and completed 3 follow-up visits | With MH service use = 0.25% (3/1187) |
| Hodgson, 2021<sup>72</sup><br>Treatment for anxiety/depression<br>Control: None<br>RoB: High | NR | ICU: 100%<br>ICU mortality: 18.4%<br>Hospitalized: 100% | Hospitalized with COVID | With MH service use = 7.5% (16/212) |
| **SEVERITY: Mix inpatient and outpatient (<90% hospitalized)** |  |  |  |  |
| Lynch, 2024<sup>52</sup><br>Received neurocognitive rehabilitation<br>Started seeing a psychiatrist<br>Started psychotherapy<br>Control: None<br>RoB: High | NR | ICU: 0%<br>Hospitalized: 12% | Undergoing neuropsychological, psychiatric and medical assessments. More than half referred from outpatient post-COVID recovery program offering treatment for various medical symptoms of PCC | Neurocognitive rehab = 2.7% (2/75)<br>Seeing psychiatrist = 5.3% (4/75)<br>Psychotherapy = 16% (12/75) |
| Diem, 2022<sup>29</sup><br>Consulted a psychiatrist<br>Consulted a psychologist<br>Control: None<br>RoB: High | NR | ICU with intubation: 2%<br>ICU without intubation: 2.6%<br>Hospitalized: 10.7% | Patients with PCC and persistent symptoms over 3 months | With psychiatrist consult = 19.7% (61/309)<br>With psychologist consult = 16.8% (52/309) |
| Wander, 2023<sup>73</sup> | NR | ICU: NR | With COVID in the national US Veterans Affairs health care system | With MH service use = 0.2% (804/388980) |
| MH clinic encounters (U09.9)<br>Control: None<br>RoB: High |  | Mechanical ventilation: 1%<br>Hospitalized: 10.3% |  |  |
| Kisiel, 2023<sup>65</sup><br>Newly introduced treatment for anxiety or depression<br>Control: None<br>RoB: Low/Moderate | Including psychological treatment or medication | ICU: 0%<br>Hospitalized: 0% | Non-hospitalized healthcare workers and other occupational groups, receiving outpatient follow-up care | With MH service use = 4.7% (17/359) |
| LeGoff, 2023<sup>31</sup><br>Attended MH treatment<br>Control: None<br>RoB: High | Attended MH treatment after assessment by neurocognitive screening evaluation with an average number of therapy session of 2.4 (range 0-42) | ICU: NR<br>Hospitalized: NR | California employees referred by occupational physician's outpatient MH provider panel for assessment and treatment due to diagnosis of PCC with neurocognitive complaints of "brain fog" | With MH service use = 14.1% (9/64) |

**Table 7:** Mental Health Services – Hospitalizations by COVID-19 timing and severity.

| OUTCOME | OUTCOME DESCRIPTION | SEVERITY | POPULATION DESCRIPTION | EVENT RATE |
| --- | --- | --- | --- | --- |
| **COVID TIMING: Mid (3-6 months)** |  |  |  |  |
| **SEVERITY: Inpatient (≥ 90% hospitalized)** |  |  |  |  |
| Kalyani, 2022<sup>51</sup><br>Readmitted for confusion<br>Control: None<br>RoB: High | Previously hospitalized patients being readmitted to hospital with complaint of confusion | ICU: 22.1%<br>Hospitalized: 100% | COVID infected reporting back with persistent symptoms (various complaints related to medical problems and ENT) | MH readmissions = 1.3% (34/2654) |
| **COVID TIMING: Long (≥6 months)** |  |  |  |  |
| **SEVERITY: Inpatient (≥ 90% hospitalized)** |  |  |  |  |
| Decio, 2022<sup>50</sup><br>Subsequent re-hospitalization<br>Control: previously hospitalized for another | At least one subsequent hospitalization following discharge for the reference hospital stay for COVID or hospitalization for another reason | ICU: 10.4%<br>Hospitalized: 100% | Hospitalized for COVID or another reason | Psychiatric disorder<br>With COVID = 0.4% (10685/2,894,088)<br>Without COVID = 8.9% (258,566/2,894,088)<br>Psychotic disorder<br>With COVID = 0.04% (1019/2,894,088) |
| medical reason other than COVID infection<br>RoB: High |  |  |  | Without COVID = 0.9% (26595/2,894,088)<br>Mood disorder<br>With COVID = 0.1% (4115/2,894,088)<br>Without COVID = 3.7% (106,861/2,894,088)<br>Anxiety disorder<br>With COVID = 0.2% (5834/2,894,088)<br>Without COVID = 4.9% (140,819/2,894,088)<br>Personality disorder<br>With COVID = 0.03% (766/2,894,088)<br>Without COVID = 0.9%(27,182/2,894,088) |
| **SEVERITY: Mix inpatient and outpatient (<90% hospitalized)** |  |  |  |  |
| Lynch, 2024<sup>52</sup><br>Hospitalized psychiatrically<br>Control: None<br>RoB: High | • NR | ICU: 0%<br>Hospitalized: 12% | Undergoing neuropsychological, psychiatric and medical assessments. More than half referred from outpatient post-COVID recovery program offering treatment for various medical symptoms of PCC | MH hospitalization = 0% (0/75) |

#### Visits to MH professionals

Twenty-one studies provided outcomes on visits to various types of MH professionals. When pooling the proportions for all studies reporting visits to a MH professional (21 studies, N=455,994), 10% (95% CI 6%, 14%) of participants reported seeing a MH professional of any type (psychiatrist, psychologist, or unspecified) (**Figure 4**). The heterogeneity was very high (I^2^=99.8%) suggesting a wide variation across studies. Proportions ranged from <1% to 37% with two falling within the overall CI.^47, 48^ Subgroup analysis for type of MH professional (**Supplementary Figure S5**), severity (inpatient vs. mixed; across all time points)) (**Supplementary Figure S6**), severity by COVID-19 variant (post-vs. preOmicron) (**Supplementary Figure S7**), and risk of bias (**Supplementary Figure S8**) did not explain the variability well. The majority of the studies (n=19, 90%) were done in OECD countries therefore we did not look at subgroups by country. Though interaction effects were not statistically significant and heterogeneity remained high within subgroups, there was a signal for more MH visits among mixed compared with inpatient populations, when looking across all studies (**Supplementary Figure S6**; p=0.10) and within each of the three categories of follow-up (**Figure 5**).

**Figure 4:**
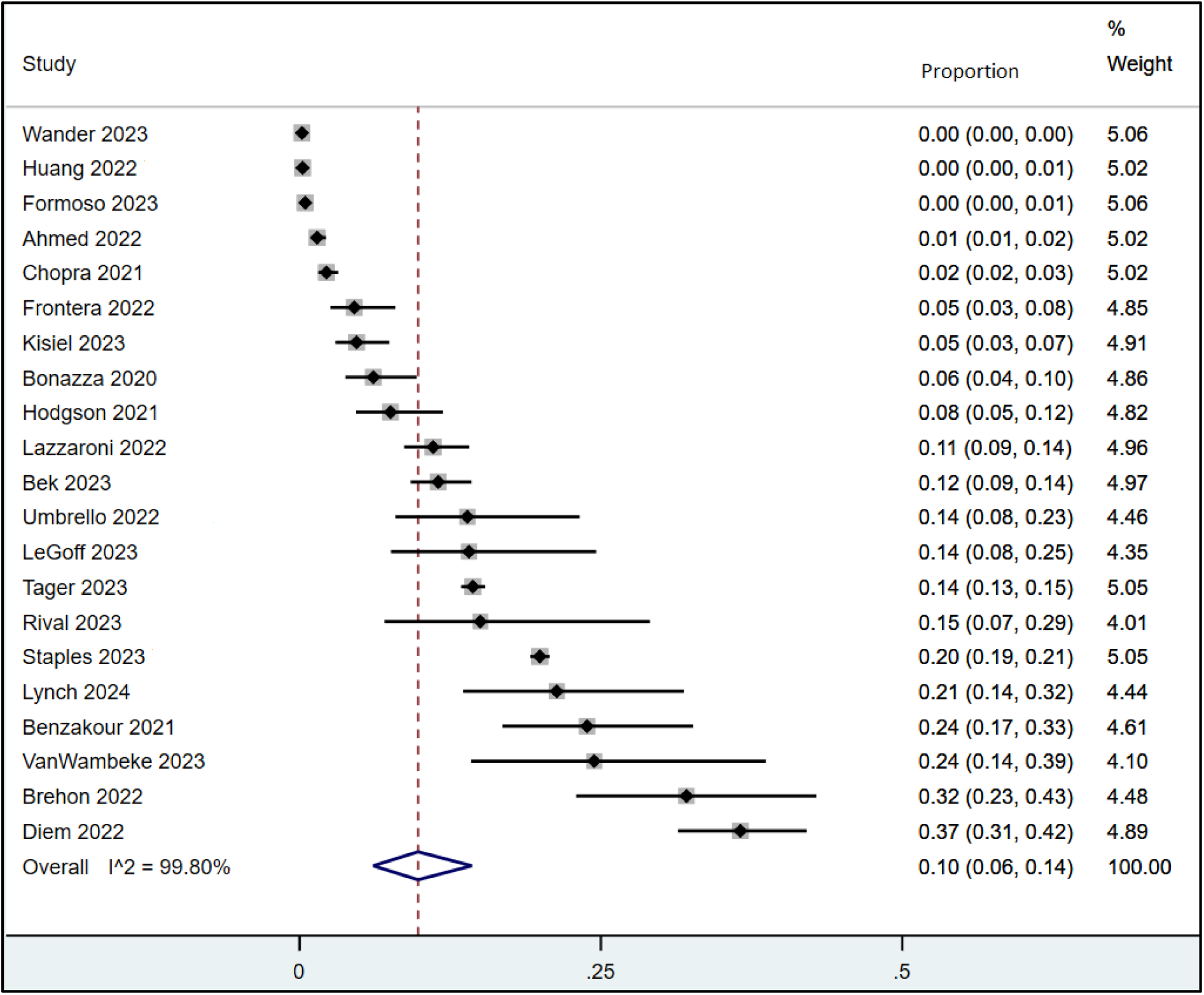
Mental health service use, all studies.

**Figure 5:**
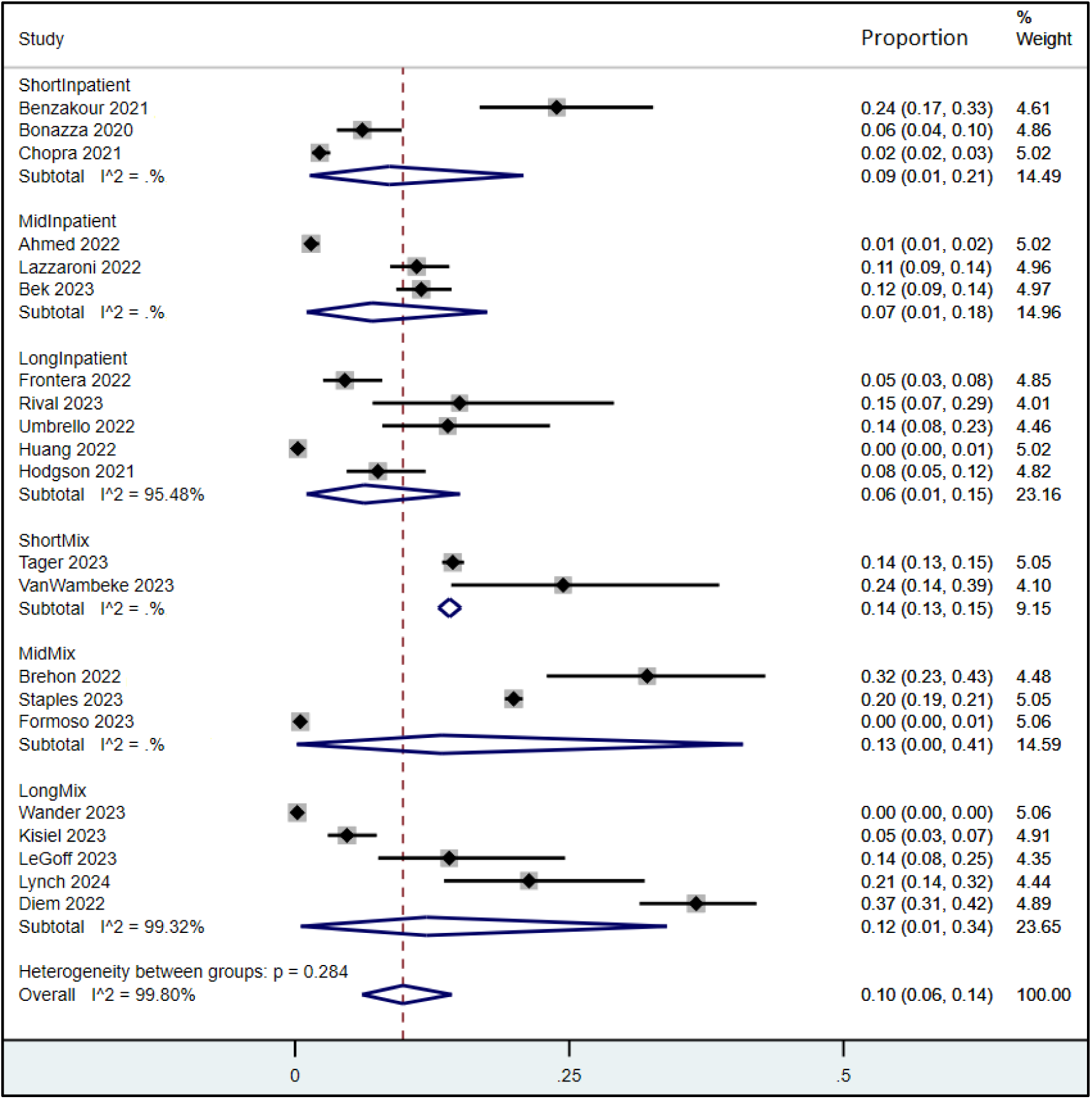
Mental health service use, by severity and timing.

There was limited information on the number of sessions attended, with the exception of Formoso et al.^49^ with a total of 117 first visits and 190 total visits, LeGoff et al.^31^ averaging 2.4 therapy sessions and Lazzaroni et al.^48^ averaging between 4-6 visits.

#### Hospitalizations for MH

Three studies reported on hospitalizations specifically for MH symptoms related to a previous COVID-19 infection (**Table 7**).^50–52^ All were assessed as high risk of bias. A nationwide study comparing hospitalizations for COVID-19 (n=96,313) and for other reasons (n=2,797,775) found that hospitalization for COVID-19 was associated with a higher risk of subsequent (12-month) hospitalization for psychiatric disorders.^50^ Eleven percent of patients initially hospitalized for COVID-19 compared with 9% of those initially hospitalized for another medical reason were re-hospitalized at least once for a psychiatric disorder. One study of 2,654 hospitalized patients, of which 22% were admitted to ICU and fewer than 5% had PCC, 1.3% were readmitted within 3 months for confusion.^51^ In the final study of 75 patients undergoing neuropsychological, psychiatric and medical assessments (12% were hospitalized and none admitted to ICU), none of the participants had been hospitalized psychiatrically over 6-month follow-up.^52^

#### Referrals to MH services

Seven studies reported on referrals to various MH services, most were assessed at high risk of bias (n=5, 71%) and just under half reported only on inpatient populations (n=3, 43%) (**Table 8**). Three studies included participants with PCC,^30, 31, 53^ two with post-COVID-19 symptoms^54, 55^ and two with COVID-19 infected populations.^48, 56^ These studies were put in a separate group since we could not determine if the referrals resulted in actual visits to MH services, with the exception of one study that did report that 62% (n=58/93 visits) of visits were based on referrals.^48^ Two studies (29%)^48, 54^ were assessed at low risk of bias with the remaining assessed at high risk of bias.^30, 31, 53, 55, 56^ Overall, participant referrals to MH services ranged from 4.2% to 45.3% for a variety of types of MH symptoms including neuropsychology,^31, 56^ psychiatric,^53, 54^ and psychological.^30, 48, 55^

**Table 8:**
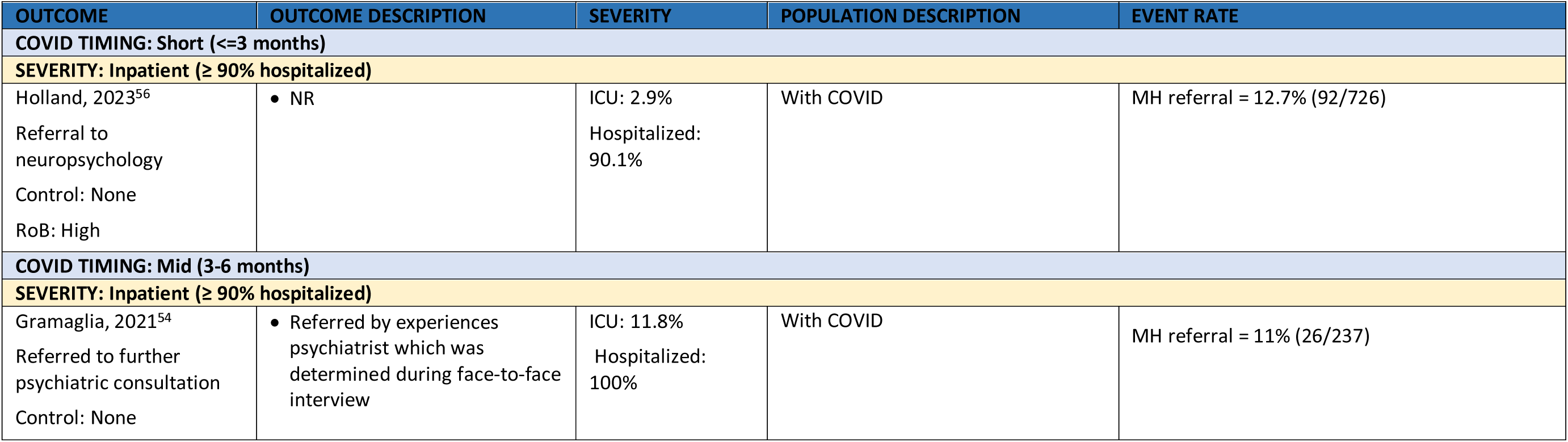
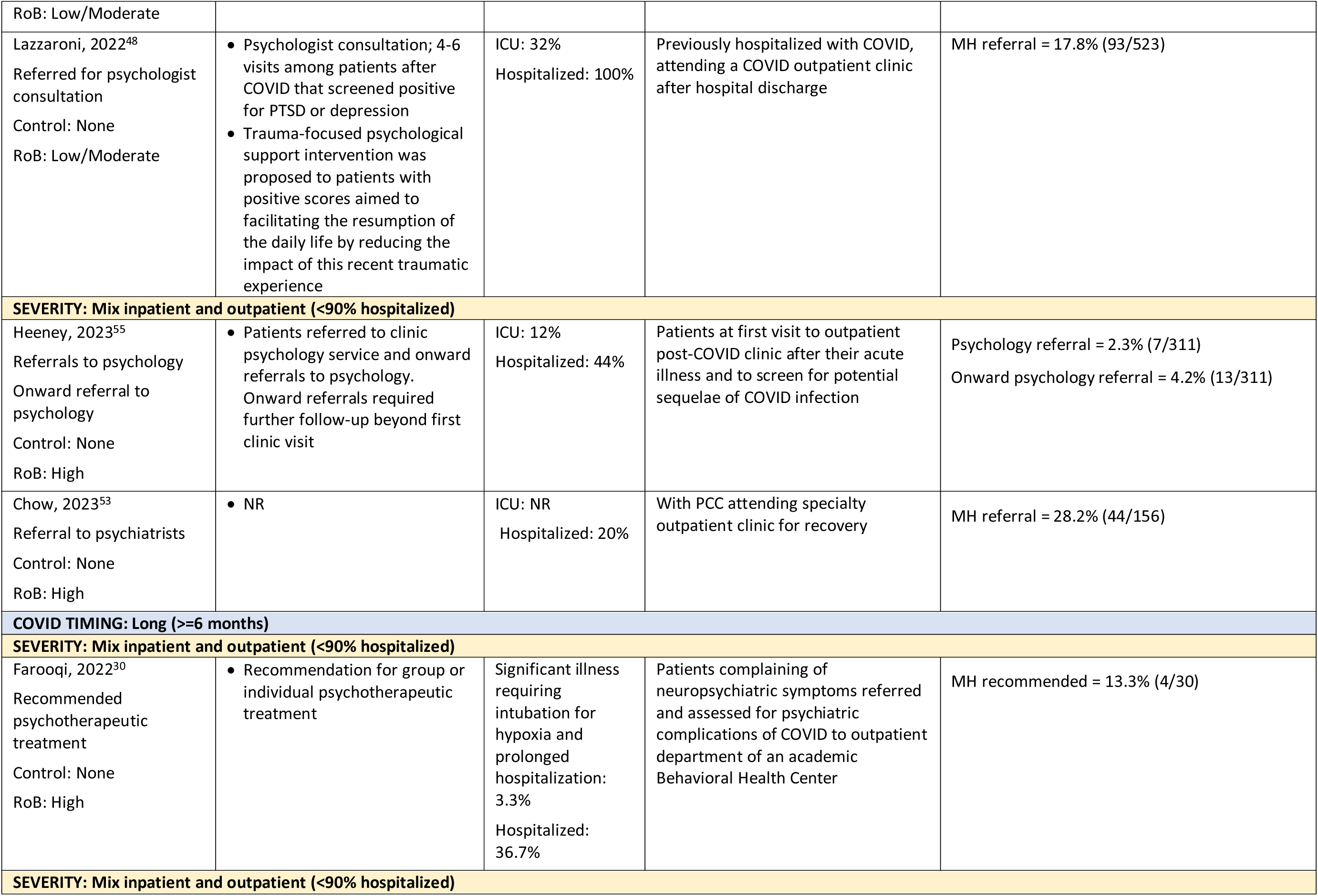
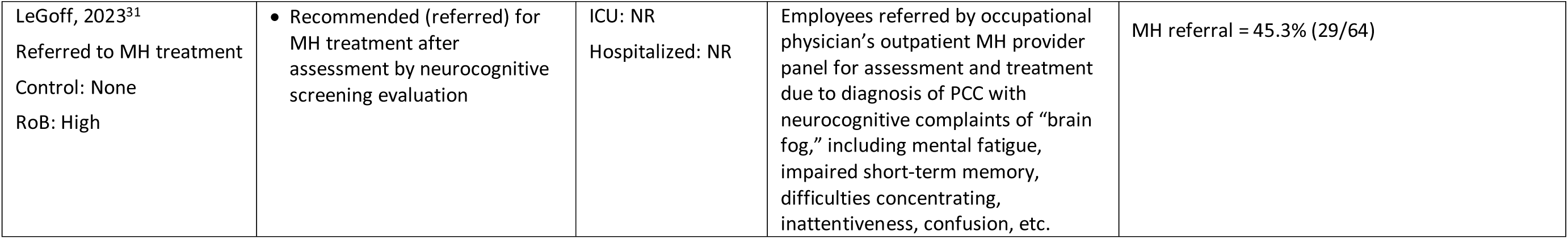
Mental Health Services – Referrals by COVID-19 timing and severity.

## DISCUSSION

This rapid review was conducted in response to the increasing concern of prolonged MH symptoms arising from COVID-19 infections on employment and the use of MH services. Specifically, this review focused on populations with new-onset, worsening, or recurring MH symptoms after acute-phase COVID-19 infection resulting in disruptions in employment and/or mental health service use and cost. Forty-five observational studies were included, 20 studies with labour force and 28 studies with MH service use outcomes. Overall, the findings across all pooled study results were highly heterogenous without any consistent pattern to explain these findings.

Although we had originally considered analyzing those without MH symptoms for comparison as control groups we found that most studies included participants that were highly symptomatic with a combination of other indicators (e.g., respiratory, gastrointestinal, pain, etc.) common with post-COVID-19 infections. Therefore, we did not prioritize finding from comparative data using a group without MH symptoms and only analyzed those having MH symptoms.

Proportions were pooled for inability to work and mental health service use, and for both outcomes the heterogeneity was very high among main analyses (I^2^ > 98%) and within subgroup strata. We attempted to explain the high variability through various subgroup analysis including risk of bias, timing since infection, severity of COVID-19, variant, populations with only MH symptoms, narrow or broad MH symptom profiles, and risk of bias. All of these analyses had equally high variability and we caution the use of the pooled estimates as representing anything apart from an average effect in the studies. For inability to work, the data appears to be most applicable to populations having post-COVID-19 symptoms/sequelae and infected with pre-Omicron strains. There was scarce data to inform duration of inability to work. For MH professional service use (i.e., psychiatrist, psychologist, or unspecified) there was limited information on the number of sessions attended. We were able to observe that a trend for more MH services use may exist for mixed versus inpatient populations, but caution against making strong conclusions to this effect. Though at high risk of bias, findings from one large study suggested that 1-2% of those hospitalized during their acute infection may be re-hospitalized due to mental health symptoms attributed to COVID-19. Overall, this rapid review highlights the variability of measurement, definition of outcomes and difficulty attributing the outcomes to MH symptoms after COVID-19 infection.

### Limitations of review

The methodology of a rapid review was undertaken to provide evidence in a timely and resource-efficient manner following recent guidelines.^57^ We employed AI software to prioritize records during title/abstract screening and did not use dual reviewer screening for our entire search; we may have missed a few studies from using this approach though our hand-searching of reference lists and reviews as well as the Scopus searches would help circumvent this. We included only English language articles and may have not included some studies published in other languages. Our risk of bias assessments relied on verification rather than two independent reviewers, though we piloted the forms and discussed all results to avoid any biases or inaccuracies. We did not formally assess the certainty of evidence for each outcome, but undertook subgroup analyses and report clearly the limitations across the studies in terms of risk of bias, heterogeneity in ascertainment of MH symptoms and study findings, and limited applicability of the results in most cases to populations with post-COVID-19 symptoms rather than more broadly post-COVID-19 or general populations.

## Conclusions

A large minority of people (possibly 25%) who experience persisting symptoms after COVID-9 may not be able to work for some period of time because of mental health symptoms. About 10% of people experiencing COVID-19 may have use for mental health care services after the acute phase, though this rate may be most applicable for those hospitalized for COVID-19. A small minority (possibly 1-2%) may require re-hospitalization for mental health issues. There is limited applicability of the results in most cases to populations with post-COVID-19 symptoms rather than more broadly post-COVID-19 or general populations. Overall, this rapid review highlights the variability of measurement, definition of outcomes and difficulty attributing the outcomes to mental health symptoms after COVID-19 infection.

## Data Availability

The data generated during this study are available within the manuscript or its supplementary files.

## DECLARATIONS

### Ethics approval and consent to participate

Not applicable.

### Consent for publication

Not applicable.

## Acknowledgments

The authors would like to thank Becky Skidmore (MLS, Independent Information Specialist) for peer-reviewing the search and creating a subsequent search on socioeconomic impact from which some search concepts were borrowed (see https://sporevidencealliance.ca/wp-content/uploads/2023/02/pcc-socioecon-impacts_les11.1_2023-01-31.pdf). They would also like to thank biostatistician Ben Vandermeer (University of Alberta) for running the analyses.

## REFERENCES

1. World Health Organization. WHO Coronavirus (COVID-19) Dashboard: WHO; 2021 [cited 2021 December 10]. Available from: https://covid19.who.int/

2. Tenforde MW, Kim SS, Lindsell CJ, Billig Rose E, Shapiro NI, Files DC, et al. Symptom duration and risk factors for delayed return to usual health among outpatients with COVID-19 in a Multistate Health Care Systems Network - United States, March-June 2020. MMWR Morb Mortal Wkly Rep. 2020;69(30):993–8.

3. Davis HE, Assaf GS, McCorkell L, Wei H, Low RJ, Re’em Y, et al. Characterizing long COVID in an international cohort: 7 months of symptoms and their impact. EClinicalMedicine. 2021;38:101019.

4. Kayaaslan B, Eser F, Kalem AK, Kaya G, Kaplan B, Kacar D, et al. Post-COVID syndrome: A single-center questionnaire study on 1007 participants recovered from COVID-19. J Med Virol. 2021;93(12):6566–74.

5. Ayoubkhani D, Khunti K, Nafilyan V, Maddox T, Humberstone B, Diamond I, et al. Post-COVID syndrome in individuals admitted to hospital with COVID-19: retrospective cohort study. BMJ (Clinical research ED). 2021;372:n693.

6. Domingo FR, Waddell LA, Cheung AM, Cooper CL, Belcourt VJ, Zuckermann AME, et al. Prevalence of long-term effects in individuals diagnosed with COVID-19: an updated living systematic review. medRxiv. 2021:2021.06.03.21258317.

7. Government of Canada Task Force on Post COVID-19 Condition. Post COVID-19 condition (long COVID). 2023.

8. Huang C, Huang L, Wang Y, Li X, Ren L, Gu X, et al. 6-month consequences of COVID-19 in patients discharged from hospital: a cohort study. Lancet. 2021;397(10270):220–32.

9. Michelen M, Manoharan L, Elkheir N, Cheng V, Dagens A, Hastie C, et al. Characterising long COVID: a living systematic review. BMJ Glob Health. 2021;6(9).

10. World Health Organization. A clinical case definition of post COVID-19 condition by a Delphi consensus, 6 October 2021.

11. Akbarialiabad H, Taghrir MH, Abdollahi A, Ghahramani N, Kumar M, Paydar S, et al. Long COVID, a comprehensive systematic scoping review. Infection. 2021;49(6):1163–86.

12. Government of Canada. COVID-19: Longer-term symptoms among Canadian adults. 2024.

13. Bonner C, Ghouralal SL. Long COVID and chronic conditions in the US workforce: prevalence, productivity loss, and disability. J Occup Environ Med. 2024;66(3):e80–e6.

14. Hejazian SS, Sadr AV, Shahjouei S, Vemuri A, Abedi V, Zand R. Prevalence and determinants of long-term post-COVID conditions in the United States: 2022 Behavioral Risk Factor Surveillance System. Am J Med. 2024.

15. Manhas KP, Horlick S, Krysa J, Kovacs Burns K, Brehon K, Laur C, et al. Implementation of a provincial long COVID care pathway in Alberta, Canada: provider perceptions. Healthcare. 2024;12(7).

16. Carson G. Research priorities for long COVID: refined through an international multi-stakeholder forum. BMC Med. 2021;19(1):84.

17. National Institutes of Health. NIH launches new initiative to study “Long COVID”: NIH; 2021 [cited 2021 December 15]. Available from: https://www.nih.gov/about-nih/who-we-are/nih-director/statements/nih-launches-new-initiative-study-long-covid.

18. Wise J. Long COVID: WHO calls on countries to offer patients more rehabilitation. BMJ. 2021;372(n405).

19. Smith N CA, Kim D, Wong G, Mitton C. COVID-19 living evidence synthesis #11.1: Socioeconomic impact of post COVID-19 condition: effects on return to work. Final report. Vancouver, Canada: Centre for Clinical Epidemiology and Evaluation, 22 January 2024.

20. Carfì A, Bernabei R, Landi F. Persistent symptoms in patients after acute COVID-19. JAMA. 2020;324(6):603–5.

21. Carvalho-Schneider C, Laurent E, Lemaignen A, Beaufils E, Bourbao-Tournois C, Laribi S, et al. Follow-up of adults with noncritical COVID-19 two months after symptom onset. Clin Microbiol Infect. 2021;27(2):258–63.

22. Zeng N, Zhao YM, Yan W, Li C, Lu QD, Liu L, et al. A systematic review and meta-analysis of long term physical and mental sequelae of COVID-19 pandemic: call for research priority and action. Mol Psychiatry. 2023;28(1):423–33.

23. Smith N DM, Wong G, Conte T, Lakzadeh P, Bunka M, Ceka A, et al. COVID-19 living evidence synthesis #11.1: Socioeconomic impact of post COVID-19 condition: Final report. Vancouver, Canada: Centre for Clinical Epidemiology and Evaluation, 31 January 2023.

24. McGowan J, Sampson M, Salzwedel DM, Cogo E, Foerster V, Lefebvre C. PRESS Peer Review of Electronic Search Strategies: 2015 Guideline Statement. J Clin Epidemiol. 2016;75:40–6.

25. Hamel C, Kelly SE, Thavorn K, Rice DB, Wells GA, Hutton B. An evaluation of DistillerSR’s machine learning-based prioritization tool for title/abstract screening - impact on reviewer-relevant outcomes. BMC Med Res Methodol. 2020;20(1):256.

26. Page MJ, McKenzie JE, Bossuyt PM, Boutron I, Hoffmann TC, Mulrow CD, et al. The PRISMA 2020 statement: an updated guideline for reporting systematic reviews. Bmj. 2021;372:n71.

27. Tufanaru C MZ, Aromataris E, Campbell J, Hopp L. Chapter 3: Systematic reviews of effectiveness. In: Aromataris E MZ, editor. Joanna Briggs Institute Reviewer’s Manual: The Joanna Briggs Institute. 2017.

28. Freeman MF, Tukey JW. Transformations related to the angular and the square root. Ann Math Stat. 1950;21:607–11.

29. Diem L, Schwarzwald A, Friedli C, Hammer H, Gomes-Fregolente L, Warncke J, et al. Multidimensional phenotyping of the post-COVID-19 syndrome: A Swiss survey study. CNS Neurosci Ther. 2022;28(12):1953–63.

30. Farooqi M, Khan A, Jacobs A, D’Souza V, Consiglio F, Karmen CL, et al. Examining the long-term sequelae of SARS-cov2 infection in patients seen in an outpatient psychiatric department. Neuropsychiatr Dis Treat. 2022;18:1259–68.

31. LeGoff DB, Lazarovic J, Kofeldt M, Peters A. Neurocognitive and symptom validity testing for post-COVID-19 condition in a workers compensation context. J Occup Environ Med. 2023;65(10):803–12.

32. Anik AI, Ahmed T, Nandonik AJ, Parvez A, Das Pooja S, Kabir ZN. Evidence of mental health-related morbidities and its association with socio-economic status among previously hospitalized patients with symptoms of COVID-19 in Bangladesh. Front Public Health. 2023;11:1132136.

33. Danesh V, Arroliga AC, Bourgeois JA, Widmer AJ, McNeal MJ, McNeal TM. Post-acute sequelae of COVID-19 in adults referred to COVID recovery clinic services in an integrated health system in Texas. Proceedings (Baylor University Medical Center). 2021;34(6):645–8.

34. Dressing A, Bormann T, Blazhenets G, Schroeter N, Walter LI, Thurow J, et al. Neuropsychologic profiles and cerebral glucose metabolism in neurocognitive long COVID Syndrome. J Nucl Med. 2022 Jul;63(7):1058–63.

35. Garcia-Molina A, Garcia-Carmona S, Espina-Bou M, Rodriguez-Rajo P, Sanchez-Carrion R, Ensenat-Cantallops A. Neuropsychological rehabilitation for post-COVID-19 syndrome: results of a clinical programme and six-month follow up. Neurologia. 2022.

36. Vanichkachorn G, Newcomb R, Cowl CT, Murad MH, Breeher L, Miller S, et al. Post-COVID-19 syndrome (long haul syndrome): description of a multidisciplinary clinic at mayo clinic and characteristics of the initial patient cohort. Mayo Clin Proc. 2021;96(7):1782–91.

37. Lunt J, Hemming S, Burton K, Elander J, Baraniak A. What workers can tell us about post-COVID workability. Occup Med (Lond). 2024;74(1):15–23.

38. Green CE, Leeds JS, Leeds CM. Occupational effects in patients with post-COVID-19 syndrome. Occup Med (Lond). 2024 Feb 19;74(1):86–92.

39. Romero-Rodriguez E, Perula-de Torres LA, Monserrat-Villatoro J, Gonzalez-Lama J, Carmona-Casado AB, Ranchal-Sanchez A. Sociodemographic and Clinical Profile of Long COVID-19 Patients, and Its Correlation with Medical Leave: A Comprehensive Descriptive and Multicenter Study. Healthcare (Basel). 2023;11(19).

40. Thompson M, Ferrando SJ, Dornbush R, Lynch S, Shahar S, Klepacz L, et al. Impact of COVID-19 on employment: sociodemographic, medical, psychiatric and neuropsychological correlates. Front Rehabil Sci. 2023;4:1150734.

41. Braga LW, Oliveira SB, Moreira AS, Pereira ME, Carneiro VS, Serio AS, et al. Neuropsychological manifestations of long COVID in hospitalized and non-hospitalized Brazilian Patients. NeuroRehabilitation. 2022;50(4):391–400.

42. Tsuchida T, Yoshimura N, Ishizuka K, Katayama K, Inoue Y, Hirose M, et al. Five cluster classifications of long COVID and their background factors: A cross-sectional study in Japan. Clin Exp Med. 2023;23(7):3663–70.

43. O’Sullivan O, Houston A, Ladlow P, Barker-Davies RM, Chamley R, Bennett AN, et al. Factors influencing medium- and long-term occupational impact following COVID-19. Occup Med (Lond). 2023.

44. Delgado-Alonso C, Cuevas C, Oliver-Mas S, Diez-Cirarda M, Delgado-Alvarez A, Gil-Moreno MJ, et al. Fatigue and cognitive dysfunction are associated with occupational status in post-COVID syndrome. Int J Environ Res Public Health. 2022;19(20).

45. Kerksieck P, Ballouz T, Haile SR, Schumacher C, Lacy J, Domenghino A, et al. Post COVID-19 condition, work ability and occupational changes in a population-based cohort. Lancet Reg Health Eur. 2023 Jun 23;31:100671.

46. Peter RS, Nieters A, Krausslich H-G, Brockmann SO, Gopel S, Kindle G, et al. Post-acute sequelae of covid-19 six to 12 months after infection: population based study. BMJ (Clinical research ED). 2022;379:e071050.

47. Bek LM, Hellemons ME, Berentschot JC, Visser MM, Huijts SM, van Bommel J, et al. Cognitive and psychological recovery patterns across different care pathways 12 months after hospitalization for COVID-19: A multicenter cohort study (CO-FLOW). Ann Phys Rehabil Med. 2023;66(5):101737.

48. Lazzaroni E, Tosi D, Pontiggia S, Ermolli R, Borghesi L, Rigamonti V, et al. Early psychological intervention in adult patients after hospitalization during COVID-19 pandemia. A single center observational study. Front Psychol. 2022;13:1059134.

49. Formoso G, Marino M, Formisano D, Grilli R. Patterns of utilisation of specialist care after SARS-Cov-2 infection: a retrospective cohort study. BMJ Open. 2023;13(3):e063493.

50. Decio V, Pirard P, Pignon B, Bouaziz O, Perduca V, Chin F, et al. Hospitalization for COVID-19 is associated with a higher risk of subsequent hospitalization for psychiatric disorders: A French nationwide longitudinal study comparing hospitalizations for COVID-19 and for other reasons. Eur Psychiatry. 2022;65(1):e70.

51. Kalyani T, Kumar BV. A study to analyze potential long-term post-COVID clinical conditions and their management in a tertiary care hospital. Int J Pharm Clin Res. 2022;14(8):353–60.

52. Lynch ST, Dornbush R, Shahar S, Mansour R, Klepacz L, Primavera LH, et al. Change in neuropsychological test performance seen in a longitudinal study of patients with post-acute sequelae of COVID-19: a 6-month follow-up study. J Acad Consult Liaison Psychiatry. 2024.

53. Chow CM, Schleyer W, DeLisi LE. The prevalence of psychiatric symptoms and their correlates as part of the long-COVID syndrome. Psychiatry Res. 2023;323:115166.

54. Gramaglia C, Gambaro E, Bellan M, Balbo PE, Baricich A, Sainaghi PP, et al. Mid-term psychiatric outcomes of patients recovered from covid-19 from an italian cohort of hospitalized patients. Front Psychol. 2021;12:667385.

55. Heeney A, Connolly SP, Dillon R, O’Donnell A, McSweeney T, O’Kelly B, et al. Post-COVID care delivery: The experience from an Irish tertiary centre’s post-COVID clinic. PLoS ONE. 2023;18(8 August):e0289245.

56. Holland AE, Fineberg D, Marceau T, Chong M, Beaman J, Wilson L, et al. The Alfred Health post-COVID-19 service, Melbourne, 2020-2022: an observational cohort study. Med J Aust. 2024 Feb 5;220(2):91–96.

57. Garritty C, Hamel C, Trivella M, Gartlehner G, Nussbaumer-Streit B, Devane D, et al. Updated recommendations for the Cochrane rapid review methods guidance for rapid reviews of effectiveness. BMJ. 2024;384:e076335.

58. Lemhofer C, Sturm C, Loudovici-Krug D, Guntenbrunner C, Bulow M, Reuken P, et al. Quality of life and ability to work of patients with Post-COVID syndrome in relation to the number of existing symptoms and the duration since infection up to 12 months: a cross-sectional study. Qual Life Res. 2023;32(7):1991–2002.

59. Ahmad M, Kim K, Indorato D, Petrenko I, Diaz K, Rotatori F, et al. Post-COVID care center to address rehabilitation needs in COVID-19 survivors: a model of care. Am J Med Qual. 2022;37(3):266–71.

60. Benzakour L, Braillard O, Mazzola V, Gex D, Nehme M, Perone SA, et al. Impact of peritraumatic dissociation in hospitalized patients with COVID-19 pneumonia: A longitudinal study. J Psychiatr Res. 2021;140:53–9.

61. Bonazza F, Borghi L, di San Marco EC, Piscopo K, Bai F, Monforte AD, et al. Psychological outcomes after hospitalization for COVID-19: data from a multidisciplinary follow-up screening program for recovered patients. Res Psychother. 2021 Jan 14;23(3):491.

62. Brehon K, Niemelainen R, Hall M, Bostick GP, Brown CA, Wieler M, et al. Return-to-Work following occupational rehabilitation for long COVID: descriptive cohort study. JMIR Rehabil Assist Technol. 2022;9(3):e39883.

63. Frontera JA, Thorpe LE, Simon NM, de Havenon A, Yaghi S, Sabadia SB, et al. Post-acute sequelae of COVID-19 symptom phenotypes and therapeutic strategies: A prospective, observational study. PloS One. 2022;17(9):e0275274.

64. Huang L, Li X, Gu X, Zhang H, Ren L, Guo L, et al. Health outcomes in people 2 years after surviving hospitalisation with COVID-19: a longitudinal cohort study. Lancet Respir Med. 2022;10(9):863–76.

65. Kisiel MA, Lee S, Janols H, Faramarzi A. Absenteeism costs due to COVID-19 and their predictors in non-hospitalized patients in sweden: a poisson regression analysis. Int J Environ Res Public Health. 2023;20(22).

66. Rival G, Chalbet S, Dupont C, Brun P, Letranchant L, Reynaud C, et al. Post-traumatic stress among COVID-19 survivors: A descriptive study of hospitalized first-wave survivors. Can J Respir Ther. 2023;59:20–5.

67. Tajer C, Martínez MJ, Mariani J, De Abreu M, Antonietti L. Post COVID-19 syndrome. Severity and evolution in 4673 health care workers. Medicina (B Aires). 2023;83(5):669–682.

68. Umbrello M, Miori S, Sanna A, Lassola S, Baruzzo E, Penzo D, et al. High rates of impaired quality of life and social and economic problems at 6 months after COVID-19-related ARDS. J Anesth Analg Crit Care. 2022;2(1):20.

69. Van Wambeke E, Bezler C, Kasprowicz A-M, Charles A-L, Andres E, Geny B. Two-years follow-up of symptoms and return to work in complex post-COVID-19 patients. J Clin Med. 2023;12(3).

70. Chopra V, Flanders SA, O’Malley M, Malani AN, Prescott HC. Sixty-day outcomes among patients hospitalized with COVID-19. Ann Intern Med. 2021;174(4):576–8.

71. Staples LG, Nielssen O, Dear BF, Bisby MA, Fisher A, Kayrouz R, et al. Prevalence and predictors of long covid in patients accessing a national digital mental health service. Int J Environ Res Public Health. 2023;20(18).

72. Hodgson CL, Higgins AM, Bailey MJ, Mather AM, Beach L, Bellomo R, et al. The impact of COVID-19 critical illness on new disability, functional outcomes and return to work at 6 months: a prospective cohort study. Crit Care. 2021;25(1):382.

73. Wander PL, Baraff A, Fox A, Cho K, Maripuri M, Honerlaw JP, et al. Rates of ICD-10 code U09.9 documentation and clinical characteristics of va patients with post-COVID-19 condition. JAMA Netw Open. 2023;6(12):e2346783.

