## Supplement for "Impacts on labour force and healthcare services related to mental-health issues following an acute SARS-CoV-2 infection: rapid review"

### Supplemental file


| | Page number |
|---|---|
| Table S1: Eligibility Criteria | 2 |
| Search Strategy | 3 |
| Included Studies | 24 |
| Excluded Studies | 28 |
| Table S2. Risk of Bias Assessments | 54 |
| Table S3a: Study Characteristics for Labour Force Outcomes with Control Groups | 58 |
| Table S3b: Study Characteristics for Labour Force Outcomes without Control Groups | 73 |
| Table S4a: Study Characteristics for Mental Health Service Use with Control Groups | 80 |
| Table S4b. Study Characteristics for Mental Health Service Use without Control Groups | 84 |
| Supplementary Figures | 102 |

### Table S1. Eligibility Criteria

| Domain | Inclusion criteria | Exclusion criteria |
|---|---|---|
| Population | For a) impacts on the labour force and healthcare costs: people of any age experiencing new/worsening/recurring MH symptoms/disorder at least 4 weeks after a SARS-CoV-2 infection. The population examined in the study may be broader, including the general population or people with previous SARS-CoV-2 infection.<br>For b) impact on healthcare services use related to MH issues: people of any age with confirmed (e.g., by laboratory testing) or suspected (e.g., physician diagnosed or self-reported) SARS-CoV-2 infection at least 4 weeks previous to outcome assessment. The population examined in the study may be broader, including the general population or those tested for SARs-CoV-2 infection.<br><br>Specific populations: age (5-17 vs. 18-65 vs. 65+ years), sex, race/ethnicity, socioeconomic status (via income or if not reported educational level), reinfection status, vaccination status (0 vs. full dosing vs. 1+ boosters), virus variant (i.e. Omicron vs. earlier, differences among pre-Omicron variants), severity of acute infection, pre and/or co-existing conditions | |
| Exposure & Comparison(s) | None required<br><br>Notes: Participants may be enrolled or have participated in a treatment study (that may or may not target their MH symptoms), though the effects of different treatments will not be considered.<br><br>We will include data for a concurrent control group without previous confirmed/suspected SARS-CoV-2 infection (ideally but not required to have negative testing at baseline and during follow-up) (“healthy control”) | |
| Outcomes & analyses | a) Among those with new-onset/worsened/recurring MH symptoms/disorders after infection:<br>• Economic burden (e.g., labor and healthcare costs)<br>• Return to work (e.g., loss of income due to inability to return to work/change in job responsibilities, proportion of individuals returning to work full time/part time/not returning, sick days, long-term disability claims)<br>• Productivity loss (e.g., presenteeism, leaveism, annual wages/income lost, impairment in the ability to perform job duties/work capacity)<br>b) After-infection MH services use, including referrals as a proxy. Any type of service provider is eligible. If all participants are identified to have a MH issue we will consider including outcomes related to use of post-COVID/multidisciplinary clinics. | |
| Timing of events/outcome assessment | Outcomes measured ≥4 weeks after a SARS-CoV-2 infection | |
| Study design, analyses & publication type | Primary research and modelling studies/economic analyses using secondary data sources with full texts. Studies must include n≥30 people with MH symptoms after COVID-19. | Conference abstracts, reviews and qualitative studies. |
| Country | Any | |
| Language | English or French | |

### Search Strategy

Ovid Medline(R) ALL

| # | Searches | Results |
|---|---|---|
| 1 | ("after covid*" or post-covid* or postcovid* or post-coronavir* or postcoronavir* or post-sarscov* or post-sars-cov*).ti,ab,kf. | 15763 |
| 2 | ((convalesc* or post-acute or postacute or "after acute" or "after discharg*" or "after hospital discharg*" or "after recover*" or "following acute" or post-infect* or postrecovery or post-recovery or post-viral or postviral or post-discharg* or postdischarg* or survivor$1) adj3 (COVID or COVID-19 or COVID19 or coronavirus* or corona virus* or 2019-nCoV or 19nCoV or 2019nCoV or nCoV or n-CoV or SARS-CoV-2 or SARS-CoV2 or SARSCoV-2 or SARSCoV2 or 2019-novel CoV or sarscov* or "sars cov 2" or "sars cov2" or Sars-coronavirus2 or "novel CoV" or "severe acute respiratory syndrome")).ti,ab,kf. | 6547 |
| 3 | (("after" or "at least" or beyond or exceeding or follow* or "greater than" or "in excess of" or later* or longer or "more than" or surpassing or upward* or longitud*) adj3 (week? or wk or wks or month? or mo or mos or year? or yr or yrs)).ti,ab,kf. | 1786889 |
| 4 | (COVID or COVID-19 or COVID19 or coronavirus* or corona virus* or 2019-nCoV or 19nCoV or 2019nCoV or nCoV or n-CoV or SARS-CoV-2 or SARS-CoV2 or SARSCoV-2 or SARSCoV2 or 2019-novel CoV or sarscov* or "sars cov 2" or "sars cov2" or Sars-coronavirus2 or "novel CoV" or "severe acute respiratory syndrome").ti,ab,kf. | 412347 |
| 5 | 3 and 4 | 24298 |
| 6 | or/1-2,5 [AFTER COVID] | 40289 |
| 7 | Post-Acute COVID-19 Syndrome/ | 2864 |
| 8 | Covid-19/rh | 214 |
| 9 | ("postcovid condition*" or "post-covid condition*" or "postcovid syndrome*" or "post-covid syndrome*").ti,ab,kf. | 761 |
| 10 | ((longhaul or long-haul or long or longterm or long-term) adj (COVID or COVID-19 or COVID19 or coronavirus* or corona virus*)).ti,ab,kf. | 4529 |
| 11 | (PASC or ((postacute or post-acute) adj2 (COVID or COVID-19 or COVID19 or coronavirus* or corona virus* or sarscov* or "sars cov 2" or "sars cov2" or "severe acute respiratory syndrome"))).ti,ab,kf. | 1678 |

| | | |
|---|---|---|
| 12 | ((COVID* or coronavirus* or "corona virus*" or ICD or "International Classification of Diseases" or "international statistical classification of diseases") and (B94* or U09* or RA02*)).ti,ab,kf. | 25 |
| 13 | ((late-effect* or late-onset or persist* or postrecovery or post-recovery or "after initial infection") adj3 (COVID or COVID-19 or COVID19 or coronavirus* or corona virus* or 2019-nCoV or 19nCoV or 2019nCoV or nCoV or n-CoV or SARS-CoV-2 or SARS-CoV2 or SARSCoV-2 or SARSCoV2 or 2019-novel CoV or sarscov* or "sars cov 2" or "sars cov2" or Sars-coronavirus2 or "novel CoV" or "severe acute respiratory syndrome")).ti,ab,kf. | 2252 |
| 14 | (("covid* care" or longcovid* or long-covid* or longhaul* or long-haul* or PASC or "postacute covid*" or "post-acute covid*" or "postacute sars*" or "post-acute sars*" or "post severe acute respiratory syndrome" or postcovid* or post-covid*) adj3 (centre or centres or center or centers or clinic or clinics or integrative or interdisciplinary or inter-disciplinary or multidisciplinary or multi-disciplinary or program or programs or programme or programmes or rehab or rehabilitation)).ti,ab,kf. | 550 |
| 15 | or/7-14 [PCC] | 8364 |
| 16 | Mental health/ | 64736 |
| 17 | ((emotional or mental) adj (health or hygiene or wellbeing or well-being or wellness)).ti,ab,kf. | 253450 |
| 18 | exp Mental Disorders/ | 1458134 |
| 19 | exp Adaptation, psychological/ | 140812 |
| 20 | exp Emotions/ | 419966 |
| 21 | Affective symptoms/ or Delusions/ or Depersonalization/ or Mental fatigue/ or Obsessive behavior/ or Paranoid behavior/ or Problem behavior/ or exp Self-injurious behavior/ or exp Stress, Psychological/ or Occupational stress/ | 264715 |
| 22 | (anxious* or Anxiet* or "attempted suicide*" or bereavement or bipolar* or compulsive* or compulsion* or Coping or Coronaphobia or delusion* or depressed or depressi* or dysthymi* or "Emotional regulation" or "emotional distress" or "Emotional disregulation" or "Emotional dysregulation" or fear* or grief or grieving or Hypochondriasis or hypomania or hypomanic or hypo-mania or hypo-manic or mania or manic or neuros#s or neurotic or obsession* or obsessive* or panic* or phobia* or phobic or post-trauma* or posttrauma* or PTSD or psychosis or psychoses or psychotic or selfharm* or self-harm* or self-injur* or selfinjur* or Suicidal* or worried or worries or worry*).ti,ab,kf. | 1132262 |

| | | |
|---|---|---|
| 23 | ((adjustment or affective or alcohol-induced or amnestic* or attention-deficit or ADHD or behaviour* or behavior* or conduct or compulsi* or conversion or dissociative or eating or emotional or hyperactiv* or hyper-activ* or impulse or impulsive* or mental or mood or neuro-behavio?ral or neuro-cognitive or neuro-psych* or neurobehavio?ral or neurocognitive or neuropsych* or obsessi* or personality or psychiatric or psychological or stress or trauma or somatoform or sleep or tic or tics or drug-induced or social functioning) adj1 (condition* or diagnos#s or disorder* or disturbed or disturbance* or episod* or ill or illness* or impact* or phenomena or phenomenon or sequela* or symptom*)).ti,ab,kf. | 520071 |
| 24 | ((alcoholism or (alcohol or anxiolytic* or drug? or hypnotic* or multi-drug? or multidrug? or nicotine or tobacco or opiate* or opioid* or poly-drug? or polydrug? or polysubstance? or poly-substance? or psychoactive* or medication or psycho-active * or sedative* or stimulant? or substance?)) adj3 (abus* or addict* or mis-us* or misus* or over-us* or overus* or "problem* use" or "use disorder*")).ti,ab,kf. | 157360 |
| 25 | Mental fatigue/ | 1885 |
| 26 | Cognitive dysfunction/ | 38121 |
| 27 | exp Neurobehavioral Manifestations/ not Intellectual disability/ | 251187 |
| 28 | ((brain or cognitive or mental or mind or thinking) adj (confusion or fatigue or fog or fogg* or cloud* or fuzz*)).ti,ab,kf. | 3695 |
| 29 | ((cognitive or cognition) adj1 (disorder* or dysfunction or impact* or impair*)).ti,ab,kf. | 122372 |
| 30 | or/16-29 [MENTAL HEALTH ISSUES] | 2921221 |
| 31 | *Incidence/ or *Prevalence/ or *Risk/ or *Odds Ratio/ | 6034 |
| 32 | (Incidence* or prevalen* or probabilit* or predict* or risk or risks or "odds ratio*").ti,ab,kf. | 6020600 |
| 33 | ((new or newly or recent) adj3 (onset or diagnos$2 or problem$1)).ti,ab,kf. | 130591 |
| 34 | (declin* or deteriorat* or provok* or provocation or recur* or trigger* or worsen*).ti,ab,kf. | 1843951 |
| 35 | (rate or rates).ti,kf. | 278875 |
| 36 | (incident adj3 (anxious* or anxiety or cogniti* or depress* or "mental health" or "mental disorder*" or "mental illness*" or mood or neurocogniti* or neuro-cogniti* or neuropsych* or neuro-psych* or psych*)).ti,ab,kf. | 2352 |
| 37 | or/31-36 [NEW ONSET] | 7555687 |

| | | |
|---|---|---|
| 38 | 30 and 37 [NEW ONSET MH ISSUES] | 925114 |
| 39 | 6 and 38 [NEW ONSET MH ISSUES AFTER COVID] | 4385 |
| 40 | Post-Acute COVID-19 Syndrome/px [Psychology] | 5 |
| 41 | (15 and 30) or 40 [PCC + MH ISSUES] | 2013 |
| 42 | 39 or 41 [PCC + MH ISSUES OR NEW ONSET MH ISSUES AFTER COVID] | 5549 |
| 43 | Employment/ | 51055 |
| 44 | Return to Work/ | 3736 |
| 45 | Return to school/ | 62 |
| 46 | ((assum* or reenter* or re-enter* or reentry or re-entry or restart* or resum* or return* or start*) adj3 (employ* or job or jobs or work* or college or school or university)).ti,ab,kf. | 28046 |
| 47 | (back-to-work or employabilit* or workabilit*).ti,ab,kf. | 3304 |
| 48 | ((able or abilit* or capabl* or incapabl* or capacit* or incapacit* or disable* or disabilit* or inabilit* or unable* or limit*) adj5 (employ* or job or jobs or work* or college or school or university)).ti,ab,kf. | 83594 |
| 49 | Sick Leave/ | 6850 |
| 50 | ((disabilt* or sick* or ill or illness* or unwell or "not well") adj5 (days or leave or "time off" or absenc* or absent* or presenc* or present)).ti,ab,kf. | 27226 |
| 51 | ((shortterm or short-term or longterm or long-term) adj3 (disabl* or disabilit*)).ti,ab,kf. | 6188 |
| 52 | Absenteeism/ | 9815 |
| 53 | Presenteeism/ | 608 |
| 54 | (absence* or absentee* or presentee* or leaveism or leavism).ti,ab,kf. | 719536 |
| 55 | "labo?r participation".ti,ab,kf. | 177 |
| 56 | Occupational Medicine/ | 23488 |
| 57 | ((industrial* or occupational*) adj medicine).ti,ab,kf. | 9579 |

| | | |
|---|---|---|
| 58 | Efficiency/ | 15738 |
| 59 | ((efficien* or productiv*) adj3 (declin* or decreas* or diminish* or less* or lose or losing or loss or losses or lost or low or lower* or reduc*)).ti,ab,kf. | 110833 |
| 60 | ((labo?r or staff*) adj1 (impact* or issue or issues or shortage*)).ti,ab,kf. | 2732 |
| 61 | ("work ability index" or "work productivity and impairment questionnaire" or "occupational status item" or "symptom burden questionnaire for long covid").ti,ab,kf. | 588 |
| 62 | or/43-61 [WORK PRODUCTIVITY - ADAPTED FROM COVID-END] | 1040057 |
| 63 | exp health care costs/ | 72511 |
| 64 | exp "cost of illness"/ | 33817 |
| 65 | ((disease or diseases or disabilit* or sick* or ill or illness* or unwell or "not well") adj5 (cost or costs or costly or costing or economic* or expenditure? or expens* or financ*)).ti,ab,kf. | 42847 |
| 66 | (burden* adj1 (health or healthcare or "health care")).ti,ab,kf. | 12232 |
| 67 | Financial Stress/ | 1161 |
| 68 | ((econom* or financ*) adj3 (challeng* or difficult* or distress* or hardship* or pressur* or problem* or strain*)).ti,ab,kf. | 23322 |
| 69 | ((econom* or financ*) adj burden*).ti,ab,kf. | 26652 |
| 70 | or/63-69 [COST OF ILLNESS] | 184676 |
| 71 | Economics/ | 27522 |
| 72 | exp "Costs and Cost Analysis"/ | 268109 |
| 73 | Economics, Nursing/ | 4013 |
| 74 | Economics, Medical/ | 9262 |
| 75 | Economics, Pharmaceutical/ | 3122 |
| 76 | exp Economics, Hospital/ | 25784 |

| | | |
|---|---|---|
| 77 | Economics, Dental/ | 1921 |
| 78 | exp "Fees and Charges"/ | 31441 |
| 79 | exp Budgets/ | 14170 |
| 80 | budget*.ti,ab,kf. | 37169 |
| 81 | (economic* or cost or costs or costly or costing or price or prices or pricing or pharmacoeconomic* or pharmaco-economic* or expenditure or expenditures or expense or expenses or financial or finance or finances or financed).ti,kf. | 289839 |
| 82 | (economic* or cost or costs or costly or costing or price or prices or pricing or pharmacoeconomic* or pharmaco-economic* or expenditure or expenditures or expense or expenses or financial or finance or finances or financed).ab. /freq=2 | 395970 |
| 83 | (cost* adj2 (effective* or utilit* or benefit* or minimi* or analy* or outcome or outcomes)).ab,kf. | 218173 |
| 84 | (value adj2 (money or monetary)).ti,ab,kf. | 3144 |
| 85 | exp models, economic/ | 16256 |
| 86 | economic model*.ab,kf. | 4357 |
| 87 | markov chains/ | 16069 |
| 88 | markov.ti,ab,kf. | 30075 |
| 89 | monte carlo method/ | 32599 |
| 90 | monte carlo.ti,ab,kf. | 61874 |
| 91 | exp Decision Theory/ | 13531 |
| 92 | (decision* adj2 (tree* or analy* or model*)).ti,ab,kf. | 40573 |
| 93 | or/71-92 | 925885 |
| 94 | ((socioeconomic* or socio-economic* or economic or economical or economies or economy or societ* or labo?r or productivity or "mental health care" or "mental healthcare" or "mental health service*") adj2 (impact$1 or effect$1 or burden$1 or cost$1 or loss or losses)).ti,ab,kf. | 90718 |

| | | |
|---|---|---|
| 95 | 62 or 70 or 93 or 94 [SOCIOECONOMIC IMPACT] | 1989256 |
| 96 | 42 and 95 [COVID MH ISSUES SocEc IMPACT] | 622 |
| 97 | 6 or 15 [AFTER COVID OR PCC] | 43618 |
| 98 | exp Mental health services/ | 106543 |
| 99 | Psychiatric rehabilitation/ | 741 |
| 100 | exp Psychotherapy/ | 221046 |
| 101 | Psychosocial support systems/ | 1009 |
| 102 | or/98-101 | 316912 |
| 103 | Help-Seeking Behavior/ or Patient acceptance of health care/ or "Referral and consultation"/ | 131170 |
| 104 | 102 and 103 | 13095 |
| 105 | exp Mental Health Services/ec, sd, sn [Statistics and numerical data, Supply & distribution, Economic aspects] | 17739 |
| 106 | Psychiatric Rehabilitation/ec, sd, sn | 45 |
| 107 | Psychotherapy/ec, sd, sn | 1527 |
| 108 | or/105-107 | 19066 |
| 109 | ((cogniti* or "e-mental health" or "e-mentalhealth" or "emental health" or "ementalhealth" or helpline* or help-line* or hotline or hot-line or "mental health" or psycholog* or psychosocial or psycho-social or psychiatr* or suicid* or telepsychiatr* or tele-psych*) adj5 (assessment* or burden or consults or consultation* or counsel?ing or evaluation* or impact* or referral* or shortage* or seek or seeking or sought or strain* or therapy or therapies or treatment* or "use" or usage* or utilis* or utiliz* or visit or visits)).ti,ab,kf. | 271916 |
| 110 | 104 or 108 or 109 [MH SERVICES USE] | 291674 |
| 111 | 97 and 110 [AFTER COVID/PCC + MH SERVICES USE] | 1900 |
| 112 | 96 or 111 [AFTER COVID/PCC - SOCIOEC IMPACT OR MH SERVICES USE] | 2369 |

| | | |
|---|---|---|
| 113 | (Case Reports.pt. or (case report? or case study or case studies).ti.) not (review* or trial*).ti,ab,kf,hw. | 2171816 |
| 114 | (comment or editorial or news or newspaper article).pt. | 1719976 |
| 115 | 112 not (113 or 114) [Remove editorials, etc] | 2287 |
| 116 | remove duplicates from 115 | 2270 |

### Ovid Embase

| # | Searches | Results |
|---|---|---|
| 1 | ("after covid*" or post-covid* or postcovid* or post-coronavir* or postcoronavir* or post-sarscov* or post-sars-cov*).ti,ab. | 20778 |
| 2 | ((convalesc* or post-acute or postacute or "after acute" or "after discharg*" or "after hospital discharg*" or "after recover*" or "following acute" or post-infect* or postrecovery or post-recovery or post-viral or postviral or post-discharg* or postdischarg* or survivor$1) adj3 (COVID or COVID-19 or COVID19 or coronavirus* or corona virus* or 2019-nCoV or 19nCoV or 2019nCoV or nCoV or n-CoV or SARS-CoV-2 or SARS-CoV2 or SARSCoV-2 or SARSCoV2 or 2019-novel CoV or sarscov* or "sars cov 2" or "sars cov2" or Sars-coronavirus2 or "novel CoV" or "severe acute respiratory syndrome")).ti,ab. | 8202 |
| 3 | (("after" or "at least" or beyond or exceeding or follow* or "greater than" or "in excess of" or later* or longer or "more than" or surpassing or upward* or longitud*) adj3 (week? or wk or wks or month? or mo or mos or year? or yr or yrs)).ti,ab. | 2672418 |
| 4 | (COVID or COVID-19 or COVID19 or coronavirus* or corona virus* or 2019-nCoV or 19nCoV or 2019nCoV or nCoV or n-CoV or SARS-CoV-2 or SARS-CoV2 or SARSCoV-2 or SARSCoV2 or 2019-novel CoV or sarscov* or "sars cov 2" or "sars cov2" or Sars-coronavirus2 or "novel CoV" or "severe acute respiratory syndrome").ti,ab. | 462412 |
| 5 | 3 and 4 | 35465 |
| 6 | or/1-2,5 [AFTER COVID] | 55072 |
| 7 | long COVID/ | 6572 |
| 8 | coronavirus disease 2019/rh | 328 |

| | | |
|---|---|---|
| 9 | ("postcovid condition*" or "post-covid condition*" or "postcovid syndrome*" or "post-covid syndrome*").ti,ab. | 915 |
| 10 | ((longhaul or long-haul or long or longterm or long-term) adj (COVID or COVID-19 or COVID19 or coronavirus* or corona virus*)).ti,ab. | 4791 |
| 11 | (PASC or ((postacute or post-acute) adj2 (COVID or COVID-19 or COVID19 or coronavirus* or corona virus* or sarscov* or "sars cov 2" or "sars cov2" or "severe acute respiratory syndrome"))).ti,ab. | 1763 |
| 12 | ((COVID* or coronavirus* or "corona virus*" or ICD or "International Classification of Diseases" or "international statistical classification of diseases") and (B94* or U09* or RA02*)).ti,ab. | 48 |
| 13 | ((late-effect* or late-onset or persist* or postrecovery or post-recovery or "after initial infection") adj3 (COVID or COVID-19 or COVID19 or coronavirus* or corona virus* or 2019-nCoV or 19nCoV or 2019nCoV or nCoV or n-CoV or SARS-CoV-2 or SARS-CoV2 or SARSCoV-2 or SARSCoV2 or 2019-novel CoV or sarscov* or "sars cov 2" or "sars cov2" or Sars-coronavirus2 or "novel CoV" or "severe acute respiratory syndrome")).ti,ab. | 2815 |
| 14 | (("covid* care" or longcovid* or long-covid* or longhaul* or long-haul* or PASC or "postacute covid*" or "post-acute covid*" or "postacute sars*" or "post-acute sars*" or "post severe acute respiratory syndrome" or postcovid* or post-covid*) adj3 (centre or centres or center or centers or clinic or clinics or integrative or interdisciplinary or inter-disciplinary or multidisciplinary or multi-disciplinary or program or programs or programme or programmes or rehab or rehabilitation)).ti,ab. | 890 |
| 15 | or/7-14 [PCC] | 11678 |
| 16 | mental health/ or psychological well-being/ | 242479 |
| 17 | ((emotional or mental) adj (health or hygiene or wellbeing or well-being or wellness)).ti,ab. | 292070 |
| 18 | exp mental disease/ | 2748391 |
| 19 | exp psychological adjustment/ | 10912 |
| 20 | exp emotion/ or sadness/ | 794151 |
| 21 | exp mental stress/ | 206269 |

| | | |
|---|---|---|
| 22 | (anxious* or Anxiet* or "attempted suicide*" or bereavement or bipolar* or compulsive* or compulsion* or Coping or Coronaphobia or delusion* or depressed or depressi* or dysthymi* or "Emotional regulation" or "emotional distress" or "Emotional disregulation" or "Emotional dysregulation" or fear* or grief or grieving or Hypochondriasis or hypomania or hypomanic or hypo-mania or hypo-manic or mania or manic or neuros#s or neurotic or obsession* or obsessive* or panic* or phobia* or phobic or post-trauma* or posttrauma* or PTSD or psychosis or psychoses or psychotic or selfharm* or self-harm* or self-injur* or selfinjur* or Suicidal* or worried or worries or worry*).ti,ab. | 1472940 |
| 23 | ((adjustment or affective or alcohol-induced or amnestic* or attention-deficit or ADHD or behaviour* or behavior* or conduct or compulsi* or conversion or dissociative or eating or emotional or hyperactiv* or hyper-activ* or impulse or impulsive* or mental or mood or neuro-behavio?ral or neuro-cognitive or neuro-psych* or neurobehavio?ral or neurocognitive or neuropsych* or obsessi* or personality or psychiatric or psychological or stress or trauma or somatoform or sleep or tic or tics or drug-induced or social functioning) adj1 (condition* or diagnos#s or disorder* or disturbed or disturbance* or episod* or ill or illness* or impact* or phenomena or phenomenon or sequela* or symptom*)).ti,ab. | 680507 |
| 24 | ((alcoholism or (alcohol or anxiolytic* or drug? or hypnotic* or multi-drug? or multidrug? or nicotine or tobacco or opiate* or opioid* or poly-drug? or polydrug? or polysubstance? or poly-substance? or psychoactive* or medication or psycho-active * or sedative* or stimulant? or substance?)) adj3 (abus* or addict* or "mis-us*" or misus* or "over-us*" or overus* or "problem* use" or "use disorder*")).ti,ab. | 211722 |
| 25 | Mental fatigue/ | 769 |
| 26 | cognitive defect/ or mild cognitive impairment/ | 251759 |
| 27 | (exp disorders of higher cerebral function/ or perception disorder/) not mental deficiency/ | 944041 |
| 28 | ((brain or cognitive or mental or mind or thinking) adj (confusion or fatigue or fog or fogg* or cloud* or fuzz*)).ti,ab. | 5278 |
| 29 | ((cognitive or cognition) adj1 (disorder* or dysfunction or impact* or impair*)).ti,ab. | 178098 |
| 30 | or/16-29 [MENTAL HEALTH ISSUES] | 4075310 |
| 31 | *incidence/ or *prevalence/ or *risk/ or *odds ratio/ | 200594 |

| | | |
|---|---|---|
| 32 | (Incidence* or prevalen* or probabilit* or predict* or risk or risks or "odds ratio*").ti,ab. | 8257118 |
| 33 | ((new or newly or recent) adj3 (onset or diagnos$2 or problem$1)).ti,ab. | 230157 |
| 34 | (declin* or deteriorat* or provok* or provocation or recur* or trigger* or worsen*).ti,ab. | 2583392 |
| 35 | (rate or rates).ti,kf. | 372357 |
| 36 | (incident adj3 (anxious* or anxiety or cogniti* or depress* or "mental health" or "mental disorder*" or "mental illness*" or mood or neurocogniti* or neuro-cogniti* or neuropsych* or neuro-psych* or psych*)).ti,ab. | 3195 |
| 37 | or/31-36 [NEW ONSET] | 10296807 |
| 38 | 30 and 37 [NEW ONSET MH ISSUES] | 1431111 |
| 39 | 6 and 38 [NEW ONSET MH ISSUES AFTER COVID] | 7181 |
| 40 | 15 and 30 [PCC + MH Issues] | 3923 |
| 41 | 39 or 40 [PCC + MH ISSUES OR NEW ONSET MH ISSUES AFTER COVID] | 9600 |
| 42 | employment/ | 78981 |
| 43 | return to work/ or work resumption/ | 13573 |
| 44 | return to school/ or school reentry/ | 721 |
| 45 | ((assum* or reenter* or re-enter* or reentry or re-entry or restart* or resum* or return* or start*) adj3 (employ* or job or jobs or work* or college or school or university)).ti,ab. | 37488 |
| 46 | (back-to-work or employabilit* or workabilit*).ti,ab. | 3669 |
| 47 | ((able or abilit* or capabl* or incapabl* or capacit* or incapacit* or disable* or disabilit* or inabilit* or unable* or limit*) adj5 (employ* or job or jobs or work* or college or school or university)).ti,ab. | 106159 |
| 48 | medical leave/ | 8849 |
| 49 | ((disabilt* or sick* or ill or illness* or unwell or "not well") adj5 (days or leave or "time off" or absenc* or absent* or presenc* or present)).ti,ab. | 36461 |
| 50 | ((shortterm or short-term or longterm or long-term) adj3 (disabl* or disabilit*)).ti,ab. | 9256 |

| | | |
|---|---|---|
| 51 | absenteeism/ | 20180 |
| 52 | presenteeism/ | 2471 |
| 53 | (absence* or absentee* or presentee* or leaveism or leavism).ti,ab. | 938870 |
| 54 | "labo?r participation".ti,ab. | 173 |
| 55 | occupational medicine/ | 12457 |
| 56 | ((industrial* or occupational*) adj medicine).ti,ab. | 6960 |
| 57 | productivity/ | 50857 |
| 58 | ((efficien* or productiv*) adj3 (declin* or decreas* or diminish* or less* or lose or losing or loss or losses or lost or low or lower* or reduc*)).ti,ab. | 129955 |
| 59 | ((labo?r or staff*) adj1 (impact* or issue or issues or shortage*)).ti,ab. | 3526 |
| 60 | ("work ability index" or "work productivity and impairment questionnaire" or "occupational status item" or "symptom burden questionnaire for long covid").ti,ab. | 750 |
| 61 | or/42-60 [WORK PRODUCTIVITY] | 1360388 |
| 62 | exp health care cost/ | 347261 |
| 63 | exp "cost of illness"/ | 21518 |
| 64 | ((disease or diseases or disabilit* or sick* or ill or illness* or unwell or "not well") adj5 (cost or costs or costly or costing or economic* or expenditure? or expens* or financ*)).ti,ab. | 54827 |
| 65 | (burden* adj1 (health or healthcare or "health care")).ti,ab. | 16317 |
| 66 | financial stress/ or financial distress/ | 4090 |
| 67 | ((econom* or financ*) adj3 (challeng* or difficult* or distress* or hardship* or pressur* or problem* or strain*)).ti,ab. | 29750 |
| 68 | ((econom* or financ*) adj burden*).ti,ab. | 41737 |
| 69 | or/62-68 [COST OF ILLNESS] | 464094 |
| 70 | Economics/ | 245119 |

| | | |
|---|---|---|
| 71 | Cost/ | 63791 |
| 72 | exp Health Economics/ | 1052441 |
| 73 | Budget/ | 34164 |
| 74 | budget*.ti,ab,kf. | 49153 |
| 75 | (economic* or cost or costs or costly or costing or price or prices or pricing or pharmacoeconomic* or pharmaco-economic* or expenditure or expenditures or expense or expenses or financial or finance or finances or financed).ti,kf. | 356219 |
| 76 | (economic* or cost or costs or costly or costing or price or prices or pricing or pharmacoeconomic* or pharmaco-economic* or expenditure or expenditures or expense or expenses or financial or finance or finances or financed).ab. /freq=2 | 551423 |
| 77 | (cost* adj2 (effective* or utilit* or benefit* or minimi* or analy* or outcome or outcomes)).ab,kf. | 301323 |
| 78 | (value adj2 (money or monetary)).ti,ab,kf. | 4228 |
| 79 | Statistical Model/ | 176015 |
| 80 | exp economic model/ | 4002 |
| 81 | economic model*.ab,kf. | 6507 |
| 82 | Probability/ | 150710 |
| 83 | markov.ti,ab,kf. | 39471 |
| 84 | monte carlo method/ | 52272 |
| 85 | monte carlo.ti,ab,kf. | 65216 |
| 86 | Decision Theory/ | 1859 |
| 87 | Decision Tree/ | 23059 |
| 88 | (decision* adj2 (tree* or analy* or model*)).ti,ab,kf. | 54781 |
| 89 | or/70-88 [Economic evaluation & models - CADTH] | 2060146 |

| | | |
|---|---|---|
| 90 | ((socioeconomic* or socio-economic* or economic or economical or economies or economy or societ* or labo?r or productivity or "mental health care" or "mental healthcare" or "mental health service*") adj2 (impact$1 or effect$1 or burden$1 or cost$1 or loss or losses)).ti,ab. | 119083 |
| 91 | 61 or 69 or 89 or 90 [SOCIOECONOMIC IMPACT] | 3402473 |
| 92 | 41 and 91 [COVID MH ISSUES SocEc IMPACT] | 1455 |
| 93 | 6 or 15 [AFTER COVID OR PCC] | 59662 |
| 94 | exp mental health service/ | 66831 |
| 95 | psychosocial rehabilitation/ or mental health recovery/ | 2782 |
| 96 | exp psychotherapy/ | 300861 |
| 97 | psychosocial care/ | 23764 |
| 98 | or/94-97 | 379921 |
| 99 | help seeking behavior/ or patient referral/ | 177720 |
| 100 | 98 and 99 | 9937 |
| 101 | ((cogniti* or "e-mental health" or "e-mentalhealth" or "emental health" or "ementalhealth" or helpline* or help-line* or hotline or hot-line or "mental health" or psycholog* or psychosocial or psycho-social or psychiatr* or suicid* or telepsychiatr* or tele-psych*) adj5 (assessment* or burden or consults or consultation* or counsel?ing or evaluation* or impact* or referral* or shortage* or seek or seeking or sought or strain* or therapy or therapies or treatment* or "use" or usage* or utilis* or utiliz* or visit or visits)).ti,ab. | 367495 |
| 102 | 100 or 101 [MH SERVICES USE] | 373393 |
| 103 | 93 and 102 [AFTER COVID/PCC + MH SERVICES USE] | 2488 |
| 104 | 92 or 103 [AFTER COVID/PCC - SOCIOEC IMPACT OR MH SERVICES USE] | 3658 |
| 105 | (Case Reports.pt. or (case report? or case study or case studies).ti.) not (review* or trial*).ti,ab,kf,hw. | 361950 |
| 106 | (comment or editorial or news or newspaper article).pt. | 791889 |
| 107 | 104 not (105 or 106) [Remove editorials, opinion pieces etc] | 3573 |

| | | |
|---|---|---|
| 108 | remove duplicates from 107 | 3511 |

### Ovid PsycInfo

| # | Searches | Results |
|---|---|---|
| 1 | ("after covid*" or post-covid* or postcovid* or post-coronavir* or postcoronavir* or post-sarscov* or post-sars-cov*).ti,ab,id. | 1362 |
| 2 | ((convalesc* or post-acute or postacute or "after acute" or "after discharg*" or "after hospital discharg*" or "after recover*" or "following acute" or post-infect* or postrecovery or post-recovery or post-viral or postviral or post-discharg* or postdischarg* or survivor$1) adj3 (COVID or COVID-19 or COVID19 or coronavirus* or corona virus* or 2019-nCoV or 19nCoV or 2019nCoV or nCoV or n-CoV or SARS-CoV-2 or SARS-CoV2 or SARSCoV-2 or SARSCoV2 or 2019-novel CoV or sarscov* or "sars cov 2" or "sars cov2" or Sars-coronavirus2 or "novel CoV" or "severe acute respiratory syndrome")).ti,ab,id. | 391 |
| 3 | (("after" or "at least" or beyond or exceeding or follow* or "greater than" or "in excess of" or later* or longer or "more than" or surpassing or upward* or longitud*) adj3 (week? or wk or wks or month? or mo or mos or year? or yr or yrs)).ti,ab,id. | 265891 |
| 4 | (COVID or COVID-19 or COVID19 or coronavirus* or corona virus* or 2019-nCoV or 19nCoV or 2019nCoV or nCoV or n-CoV or SARS-CoV-2 or SARS-CoV2 or SARSCoV-2 or SARSCoV2 or 2019-novel CoV or sarscov* or "sars cov 2" or "sars cov2" or Sars-coronavirus2 or "novel CoV" or "severe acute respiratory syndrome").ti,ab,id. | 43086 |
| 5 | 3 and 4 | 2592 |
| 6 | or/1-2,5 [AFTER COVID] | 3932 |
| 7 | Post-COVID-19 Conditions/ | 158 |
| 8 | ("postcovid condition*" or "post-covid condition*" or "postcovid syndrome*" or "post-covid syndrome*").ti,ab,id. | 60 |
| 9 | ((longhaul or long-haul or long or longterm or long-term) adj (COVID or COVID-19 or COVID19 or coronavirus* or corona virus*)).ti,ab,id. | 263 |

| | | |
|---|---|---|
| 10 | (PASC or ((postacute or post-acute) adj2 (COVID or COVID-19 or COVID19 or coronavirus* or corona virus* or sarscov* or "sars cov 2" or "sars cov2" or "severe acute respiratory syndrome"))).ti,ab,id. | 88 |
| 11 | ((COVID* or coronavirus* or "corona virus*" or ICD or "International Classification of Diseases" or "international statistical classification of diseases") and (B94* or U09* or RA02*)).ti,ab,id. | 0 |
| 12 | ((late-effect* or late-onset or persist* or postrecovery or post-recovery or "after initial infection") adj3 (COVID or COVID-19 or COVID19 or coronavirus* or corona virus* or 2019-nCoV or 19nCoV or 2019nCoV or nCoV or n-CoV or SARS-CoV-2 or SARS-CoV2 or SARSCoV-2 or SARSCoV2 or 2019-novel CoV or sarscov* or "sars cov 2" or "sars cov2" or Sars-coronavirus2 or "novel CoV" or "severe acute respiratory syndrome")).ti,ab,id. | 117 |
| 13 | (("covid* care" or longcovid* or long-covid* or longhaul* or long-haul* or PASC or "postacute covid*" or "post-acute covid*" or "postacute sars*" or "post-acute sars*" or "post severe acute respiratory syndrome" or postcovid* or post-covid*) adj3 (centre or centres or center or centers or clinic or clinics or integrative or interdisciplinary or inter-disciplinary or multidisciplinary or multi-disciplinary or program or programs or programme or programmes or rehab or rehabilitation)).ti,ab,id. | 34 |
| 14 | or/7-13 [PCC] | 449 |
| 15 | Mental Health/ or Emotional Health/ | 94307 |
| 16 | ((emotional or mental) adj (health or hygiene or wellbeing or well-being or wellness)).ti,ab. | 246075 |
| 17 | exp Mental Disorders/ or Psychiatric Symptoms/ | 1097994 |
| 18 | exp Emotional Adjustment/ | 23216 |
| 19 | exp Emotional States/ | 373632 |
| 20 | Emotional Disturbances/ or exp Psychological Stress/ or Occupational Stress/ | 45883 |
| 21 | (anxious* or Anxiet* or "attempted suicide*" or bereavement or bipolar* or compulsive* or compulsion* or Coping or Coronaphobia or delusion* or depressed or depressi* or dysthymi* or "Emotional regulation" or "emotional distress" or "Emotional disregulation" or "Emotional dysregulation" or fear* or grief or grieving or Hypochondriasis or hypomania or hypomanic or hypo-mania or hypo-manic or mania or manic or neuros#s or neurotic or obsession* or obsessive* or panic* or phobia* or phobic or post-trauma* or posttrauma* or PTSD or psychosis or psychoses or psychotic or selfharm* or self-harm* or self-injur* or selfinjur* or Suicidal* or worried or worries or worry*).ti,ab. | 855479 |

| | | |
|---|---|---|
| 22 | ((adjustment or affective or alcohol-induced or amnestic* or attention-deficit or ADHD or behaviour* or behavior* or conduct or compulsi* or conversion or dissociative or eating or emotional or hyperactiv* or hyper-activ* or impulse or impulsive* or mental or mood or neuro-behavio?ral or neuro-cognitive or neuro-psych* or neurobehavio?ral or neurocognitive or neuropsych* or obsessi* or personality or psychiatric or psychological or stress or trauma or somatoform or sleep or tic or tics or drug-induced or social functioning) adj1 (condition* or diagnos#s or disorder* or disturbed or disturbance* or episod* or ill or illness* or impact* or phenomena or phenomenon or sequela* or symptom*)).ti,ab. | 439546 |
| 23 | ((alcoholism or (alcohol or anxiolytic* or drug? or hypnotic* or multi-drug? or multidrug? or nicotine or tobacco or opiate* or opioid* or poly-drug? or polydrug? or polysubstance? or poly-substance? or psychoactive* or medication or psycho-active * or sedative* or stimulant? or substance?)) adj3 (abus* or addict* or "mis-us*" or misus* or "over-us*" or overus* or "problem* use" or "use disorder*")).ti,ab. | 116076 |
| 24 | Cognitive Impairment/ | 45377 |
| 25 | exp Neurocognitive Disorders/ or Perceptual Disturbances/ | 113502 |
| 26 | ((brain or cognitive or mental or mind or thinking) adj (confusion or fatigue or fog or fogg* or cloud* or fuzz*)).ti,ab. | 1698 |
| 27 | ((cognitive or cognition) adj1 (disorder* or dysfunction or impact* or impair*)).ti,ab. | 58320 |
| 28 | or/15-27 [MENTAL HEALTH ISSUES] | 1899794 |
| 29 | (Incidence* or prevalen* or probabilit* or predict* or risk or risks or "odds ratio*").ti,ab,id. | 1134039 |
| 30 | ((new or newly or recent) adj3 (onset or diagnos$2 or problem$1)).ti,ab,id. | 15163 |
| 31 | (declin* or deteriorat* or provok* or provocation or recur* or trigger* or worsen*).ti,ab,id. | 229991 |
| 32 | (rate or rates).ti,id. | 53754 |
| 33 | (incident adj3 (anxious* or anxiety or cogniti* or depress* or "mental health" or "mental disorder*" or "mental illness*" or mood or neurocogniti* or neuro-cogniti* or neuropsych* or neuro-psych* or psych*)).ti,ab,id. | 1434 |
| 34 | or/29-33 [NEW ONSET] | 1340031 |

| | | |
|---|---|---|
| 35 | 28 and 34 [NEW ONSET MH ISSUES] | 617081 |
| 36 | 6 and 35 [NEW ONSET MH ISSUES AFTER COVID] | 1383 |
| 37 | 14 and 28 [PCC + MH ISSUES] | 305 |
| 38 | 36 or 37 [PCC + MH ISSUES OR NEW ONSET MH ISSUES AFTER COVID] | 1580 |
| 39 | Employment Status/ | 19141 |
| 40 | Employability/ or Job loss/ or Reemployment/ or Unemployment/ | 8825 |
| 41 | ((assum* or reenter* or re-enter* or reentry or re-entry or restart* or resum* or return* or start*) adj3 (employ* or job or jobs or work* or college or school or university)).ti,ab,id. | 14356 |
| 42 | (back-to-work or employabilit* or workabilit*).ti,ab,id. | 3041 |
| 43 | ((able or abilit* or capabl* or incapabl* or capacit* or incapacit* or disable* or disabilit* or inabilit* or unable* or limit*) adj5 (employ* or job or jobs or work* or college or school or university)).ti,ab,id. | 65020 |
| 44 | Employee Leave Benefits/ | 1581 |
| 45 | ((disabilt* or sick* or ill or illness* or unwell or "not well") adj5 (days or leave or "time off" or absenc* or absent* or presenc* or present)).ti,ab,id. | 8016 |
| 46 | ((shortterm or short-term or longterm or long-term) adj3 (disabl* or disabilit*)).ti,ab,id. | 1563 |
| 47 | Employee Absenteeism/ | 2458 |
| 48 | (absence* or absentee* or presentee* or leaveism or leavism).ti,ab,id. | 93470 |
| 49 | "labo?r participation".ti,ab,id. | 131 |
| 50 | ((industrial* or occupational*) adj medicine).ti,ab,id. | 368 |
| 51 | exp Job Performance/ | 24443 |
| 52 | ((efficien* or productiv*) adj3 (declin* or decreas* or diminish* or less* or lose or losing or loss or losses or lost or low or lower* or reduc*)).ti,ab,id. | 11202 |
| 53 | ((labo?r or staff*) adj1 (impact* or issue or issues or shortage*)).ti,ab,id. | 954 |

| | | |
|---|---|---|
| 54 | ("work ability index" or "work productivity and impairment questionnaire" or "occupational status item" or "symptom burden questionnaire for long covid").ti,ab,id. | 179 |
| 55 | or/39-54 [WORK PRODUCTIVITY] | 228924 |
| 56 | exp Health Care Costs/ | 26195 |
| 57 | ((disease or diseases or disabilit* or sick* or ill or illness* or unwell or "not well") adj5 (cost or costs or costly or costing or economic* or expenditure? or expens* or financ*)).ti,ab,id. | 6312 |
| 58 | (burden* adj1 (health or healthcare or "health care")).ti,ab,id. | 1893 |
| 59 | Financial Strain/ | 4601 |
| 60 | ((econom* or financ*) adj3 (challeng* or difficult* or distress* or hardship* or pressur* or problem* or strain*)).ti,ab,id. | 12763 |
| 61 | ((econom* or financ*) adj burden*).ti,ab,id. | 3561 |
| 62 | or/56-61 [COST OF ILLNESS] | 49845 |
| 63 | Economics/ | 28322 |
| 64 | exp "Costs and Cost Analysis"/ | 50239 |
| 65 | exp Health Care Economics/ | 1237 |
| 66 | Budgets/ | 1468 |
| 67 | budget*.ti,ab,id. | 10356 |
| 68 | (economic* or cost or costs or costly or costing or price or prices or pricing or pharmacoeconomic* or pharmaco-economic* or expenditure or expenditures or expense or expenses or financial or finance or finances or financed).ti,id. | 87606 |
| 69 | (economic* or cost or costs or costly or costing or price or prices or pricing or pharmacoeconomic* or pharmaco-economic* or expenditure or expenditures or expense or expenses or financial or finance or finances or financed).ab. /freq=2 | 108818 |
| 70 | (cost* adj2 (effective* or utilit* or benefit* or minimi* or analy* or outcome or outcomes)).ab,id. | 30529 |
| 71 | (value adj2 (money or monetary)).ti,ab,id. | 1178 |

| | | |
|---|---|---|
| 72 | Statistical Probability/ | 8241 |
| 73 | economic model*.ab,id. | 1185 |
| 74 | markov.ti,ab,id. | 4850 |
| 75 | monte carlo.ti,ab,id. | 5627 |
| 76 | Decision Theory/ | 1412 |
| 77 | (decision* adj2 (tree* or analy* or model*)).ti,ab,id. | 11291 |
| 78 | or/63-77 [Economic evaluation & models - adapted from CADTH] | 213469 |
| 79 | ((socioeconomic* or socio-economic* or economic or economical or economies or economy or societ* or labo?r or productivity or "mental health care" or "mental healthcare" or "mental health service*") adj2 (impact$1 or effect$1 or burden$1 or cost$1 or loss or losses)).ti,ab,id. | 17073 |
| 80 | 55 or 62 or 78 or 79 [SOCIOECONOMIC IMPACT] | 443596 |
| 81 | 38 and 80 [COVID MH ISSUES SocEc IMPACT] | 197 |
| 82 | 6 or 14 [AFTER COVID/ PPC] | 4096 |
| 83 | exp Mental Health Services/ or Telepsychology/ or Digital Mental Health Resources/ | 59185 |
| 84 | exp Psychosocial Rehabilitation/ | 14758 |
| 85 | exp Psychotherapy/ | 224743 |
| 86 | or/83-85 | 293768 |
| 87 | Help Seeking Behavior/ or Professional Referral/ or Professional Consultation/ | 20354 |
| 88 | 86 and 87 | 3250 |
| 89 | ((cogniti* or "e-mental health" or "e-mentalhealth" or "emental health" or "ementalhealth" or helpline* or help-line* or hotline or hot-line or "mental health" or psycholog* or psychosocial or psycho-social or psychiatr* or suicid* or telepsychiatr* or tele-psych*) adj5 (assessment* or burden or consults or consultation* or counsel?ing or evaluation* or impact* or referral* or shortage* or seek or seeking or sought or strain* or therapy or therapies or treatment* or "use" or usage* or utilis* or utiliz* or visit or visits)).ti,ab,id. | 304405 |

| | | |
|---|---|---|
| 90 | 88 or 89 [MH SERVICES USE] | 306003 |
| 91 | 82 and 90 [AFTER COVID/PCC + MH SERVICES USE] | 708 |
| 92 | 81 or 91 [AFTER COVID/PCC - SOCIOEC IMPACT OR MH SERVICES USE] | 844 |
| 93 | (case report? or case study or case studies).ti. not (review* or trial*).ti,ab,id,hw. | 37896 |
| 94 | (comment or editorial or opinion).dt. [doc type] | 44474 |
| 95 | 92 not (93 or 94) [Remove editorials, opinion pieces, etc] | 833 |
| 96 | remove duplicates from 95 | 831 |

### Included Studies


1. Ahmad M, Kim K, Indorato D, Petrenko I, Diaz K, Rotatori F, et al. Post-COVID Care Center to Address Rehabilitation Needs in COVID-19 Survivors: A Model of Care. American J Medical Quality. 2022;37(3):266-71.

2. Anik AI, Ahmed T, Nandonik AJ, Parvez A, Das Pooja S, Kabir ZN. Evidence of mental health-related morbidities and its association with socio-economic status among previously hospitalized patients with symptoms of COVID-19 in Bangladesh. Frontiers in Public Health. 2023;11:1132136.

3. Bek LM, Hellemons ME, Berentschot JC, Visser MM, Huijts SM, van Bommel J, et al. Cognitive and psychological recovery patterns across different care pathways 12 months after hospitalization for COVID-19: A multicenter cohort study (CO-FLOW). Annals of Physical and Rehabilitation Medicine. 2023;66(5):101737.

4. Benzakour L, Braillard O, Mazzola V, Gex D, Nehme M, Perone SA, et al. Impact of peritraumatic dissociation in hospitalized patients with COVID-19 pneumonia: A longitudinal study. J Psychiatric Research. 2021;140:53-9.

5. Bonazza F, Borghi L, di San Marco EC, Piscopo K, Bai F, Monforte AA, Vegni E. Psychological outcomes after hospitalization for COVID-19: data from a multidisciplinary follow-up screening program for recovered patients. Research in Psychotherapy (Milano). 2020;23(3):491.

6. Braga LW, Oliveira SB, Moreira AS, Pereira ME, Carneiro VS, Serio AS, et al. Neuropsychological manifestations of long COVID in hospitalized and non-hospitalized Brazilian Patients. NeuroRehabilitation. 2022;50(4):391-400.

7. Brehon K, Niemelainen R, Hall M, Bostick GP, Brown CA, Wieler M, Gross DP. Return-to-Work Following Occupational Rehabilitation for Long COVID: Descriptive Cohort Study. JMIR Rehabilitation and Assistive Ttechnologies. 2022;9(3):e39883.

8. Chopra V, Flanders SA, O'Malley M, Malani AN, Prescott HC. Sixty-Day Outcomes Among Patients Hospitalized With COVID-19. Ann Intern Med. 2021;174(4):576-8.

9. Chow CM, Schleyer W, DeLisi LE. The prevalence of psychiatric symptoms and their correlates as part of the long-COVID syndrome. Psychiatry Research. 2023;323:115166.

10. Danesh V, Arroliga AC, Bourgeois JA, Widmer AJ, McNeal MJ, McNeal TM. Post-acute sequelae of COVID-19 in adults referred to COVID recovery clinic services in an integrated health system in Texas. Proceedings (Baylor University Medical Center). 2021;34(6):645-8.

11. Davis HE, Assaf GS, McCorkell L, Wei H, Low RJ, Re'em Y, et al. Characterizing long COVID in an international cohort: 7 months of symptoms and their impact. EClinicalMedicine. 2021;38:101019.

12. Decio V, Pirard P, Pignon B, Bouaziz O, Perduca V, Chin F, et al. Hospitalization for COVID-19 is associated with a higher risk of subsequent hospitalization for psychiatric disorders: A French nationwide longitudinal study comparing hospitalizations for COVID-19 and for other reasons. European Psychiatry. 2022;65(1):e70.

13. Delgado-Alonso C, Cuevas C, Oliver-Mas S, Diez-Cirarda M, Delgado-Alvarez A, Gil-Moreno MJ, et al. Fatigue and Cognitive Dysfunction Are Associated with Occupational Status in Post-COVID Syndrome. International Journal of Environmental Research And Public Health. 2022;19(20).

14. Diem L, Schwarzwald A, Friedli C, Hammer H, Gomes-Fregolente L, Warncke J, et al. Multidimensional phenotyping of the post-COVID-19 syndrome: A Swiss survey study. CNS Neuroscience & Therapeutics. 2022;28(12):1953-63.

15. Dressing A, Bormann T, Blazhenets G, Schroeter N, Walter LI, Thurow J, et al. Neuropsychologic Profiles and Cerebral Glucose Metabolism in Neurocognitive Long COVID Syndrome. J Nuclear Medicine. 2022;63(7):1058-63.

16. Farooqi M, Khan A, Jacobs A, D'Souza V, Consiglio F, Karmen CL, et al. Examining the Long-Term Sequelae of SARS-CoV2 Infection in Patients Seen in an Outpatient Psychiatric Department. Neuropsychiatric Disease and Treatment. 2022;18:1259-68.

17. Formoso G, Marino M, Formisano D, Grilli R. Patterns of utilisation of specialist care after SARS-Cov-2 infection: a retrospective cohort study. BMJ Open. 2023;13(3):e063493.

18. Frontera JA, Thorpe LE, Simon NM, de Havenon A, Yaghi S, Sabadia SB, et al. Post-acute sequelae of COVID-19 symptom phenotypes and therapeutic strategies: A prospective, observational study. PloS one. 2022;17(9):e0275274.

19. Garcia-Molina A, Garcia-Carmona S, Espina-Bou M, Rodriguez-Rajo P, Sanchez-Carrion R, Ensenat-Cantallops A. Neuropsychological rehabilitation for post-COVID-19 syndrome: results of a clinical programme and six-month follow up. Neurologia. 2022.

20. Gramaglia C, Gambaro E, Bellan M, Balbo PE, Baricich A, Sainaghi PP, et al. Mid-term Psychiatric Outcomes of Patients Recovered From COVID-19 From an Italian Cohort of Hospitalized Patients. Frontiers in Psychiatry. 2021;12:667385.

21. Green CE, Leeds JS, Leeds CM. Occupational effects in patients with post-COVID-19 syndrome. Occupational Medicine (Oxford, England). 2023.

22. Heeney A, Connolly SP, Dillon R, O'Donnell A, McSweeney T, O'Kelly B, et al. Post-COVID care delivery: The experience from an Irish tertiary centre's post-COVID clinic. PLoS ONE. 2023;18(8 August):e0289245.

23. Hodgson CL, Higgins AM, Bailey MJ, Mather AM, Beach L, Bellomo R, et al. The impact of COVID-19 critical illness on new disability, functional outcomes and return to work at 6 months: a prospective cohort study. Crit Care. 2021;25(1):382.

24. Holland AE, Fineberg D, Marceau T, Chong M, Beaman J, Wilson L, et al. The Alfred Health post-COVID-19 service, Melbourne, 2020-2022: an observational cohort study. The Medical Journal of Australia. 2023.

25. Huang L, Li X, Gu X, Zhang H, Ren L, Guo L, et al. Health outcomes in people 2 years after surviving hospitalisation with COVID-19: a longitudinal cohort study. The Lancet Respiratory medicine. 2022;10(9):863-76.

26. Kalyani T, Kumar BV. A Study to Analyze Potential Long-Term Post-COVID Clinical Conditions and Their Management in a Tertiary Care Hospital. International Journal of Pharmaceutical and Clinical Research. 2022;14(8):353-60.

27. Kerksieck P, Ballouz T, Haile SR, Schumacher C, Lacy J, Domenghino A, et al. Post COVID-19 condition, work ability and occupational changes in a population-based cohort. The Lancet Regional Health Europe. 2023:100671.

28. Kisiel MA, Lee S, Janols H, Faramarzi A. Absenteeism Costs Due to COVID-19 and Their Predictors in Non-Hospitalized Patients in Sweden: A Poisson Regression Analysis. International Journal of Environmental Research and Public Health. 2023;20(22).

29. Lazzaroni E, Tosi D, Pontiggia S, Ermolli R, Borghesi L, Rigamonti V, et al. Early psychological intervention in adult patients after hospitalization during COVID-19 pandemia. A single center observational study. Frontiers in Psychology. 2022;13:1059134.

30. LeGoff DB, Lazarovic J, Kofeldt M, Peters A. Neurocognitive and Symptom Validity Testing for Post-COVID-19 Condition in a Workers Compensation Context. Journal of Occupational and Environmental Medicine. 2023;65(10):803-12.

31. Lemhofer C, Sturm C, Loudovici-Krug D, Guntenbrunner C, Bulow M, Reuken P, et al. Quality of life and ability to work of patients with Post-COVID syndrome in relation to the number of existing symptoms and the duration since infection up to 12 months: a cross-sectional study. Quality of Life Research. 2023;32(7):1991-2002.

32. Lunt J, Hemming S, Burton K, Elander J, Baraniak A. What workers can tell us about post-COVID workability. Occup Med (Lond). 2024;74(1):15-23.

33. Lynch ST, Dornbush R, Shahar S, Mansour R, Klepacz L, Primavera LH, Ferrando SJ. Change in Neuropsychological Test Performance Seen in a Longitudinal Study of Patients with Post-Acute Sequelae of COVID-19: A 6-Month Follow-up Study. Journal of the Academy of Consultation-Liaison Psychiatry. 2024.

34. O'Sullivan O, Houston A, Ladlow P, Barker-Davies RM, Chamley R, Bennett AN, et al. Factors influencing medium- and long-term occupational impact following COVID-19. Occupational Medicine (Oxford, England). 2023.

35. Peter RS, Nieters A, Krausslich H-G, Brockmann SO, Gopel S, Kindle G, et al. Post-acute sequelae of covid-19 six to 12 months after infection: population based study. BMJ (Clinical research ed). 2022;379:e071050.

36. Rival G, Chalbet S, Dupont C, Brun P, Letranchant L, Reynaud C, et al. Post-traumatic stress among COVID-19 survivors: A descriptive study of hospitalized first-wave survivors. Canadian Journal of Respiratory Therapy. 2023;59:20-5.

37. Romero-Rodriguez E, Perula-de Torres LA, Monserrat-Villatoro J, Gonzalez-Lama J, Carmona-Casado AB, Ranchal-Sanchez A. Sociodemographic and Clinical Profile of Long COVID-19 Patients, and Its Correlation with Medical Leave: A Comprehensive Descriptive and Multicenter Study. Healthcare (Basel, Switzerland). 2023;11(19).

38. Staples LG, Nielssen O, Dear BF, Bisby MA, Fisher A, Kayrouz R, Titov N. Prevalence and Predictors of Long COVID in Patients Accessing a National Digital Mental Health Service. International journal of environmental research and public health. 2023;20(18).

39. Tajer C, Martinez MJ, Mariani J, De Abreu M, Antonietti L. Post COVID-19 syndrome. Severity and evolution in 4673 health care workers. MEDICINA. 2023;83(5):669-82.

40. Thompson M, Ferrando SJ, Dornbush R, Lynch S, Shahar S, Klepacz L, Smiley A. Impact of COVID-19 on employment: sociodemographic, medical, psychiatric and neuropsychological correlates. Frontiers in rehabilitation sciences. 2023;4:1150734.

41. Tsuchida T, Yoshimura N, Ishizuka K, Katayama K, Inoue Y, Hirose M, et al. Five cluster classifications of long COVID and their background factors: A cross-sectional study in Japan. Clinical and experimental medicine. 2023;23(7):3663-70.

42. Umbrello M, Miori S, Sanna A, Lassola S, Baruzzo E, Penzo D, et al. High rates of impaired quality of life and social and economic problems at 6 months after COVID-19-related ARDS. Journal of Anesthesia, Analgesia and Critical Care (Online). 2022;2(1):20.

43. Van Wambeke E, Bezler C, Kasprowicz A-M, Charles A-L, Andres E, Geny B. Two-Years Follow-Up of Symptoms and Return to Work in Complex Post-COVID-19 Patients. Journal of Clinical Medicine. 2023;12(3).

44. Vanichkachorn G, Newcomb R, Cowl CT, Murad MH, Breeher L, Miller S, et al. Post-COVID-19 Syndrome (Long Haul Syndrome): Description of a Multidisciplinary Clinic at Mayo Clinic and Characteristics of the Initial Patient Cohort. Mayo Clinic Proceedings. 2021;96(7):1782-91.

45. Wander PL, Baraff A, Fox A, Cho K, Maripuri M, Honerlaw JP, et al. Rates of ICD-10 Code U09.9 Documentation and Clinical Characteristics of VA Patients With Post-COVID-19 Condition. JAMA Netw Open. 2023;6(12):e2346783.

### Excluded Studies

**Study design (n=129)**


1. Ahmadi N, Steinberg A, Pynoos R. The effect of reminder focused positive psychiatry suicide safety (RFPP-S) on COVID-19 related PTSD with PGD. Brain, Behavior, and Immunity. 2021;98(Supplement):1.
2. Aitcheson E, Robinson V, FitzMaurice T. Exploring the holistic long-term impact on COVID-19 in a post-hospitalised patient cohort: A service improvement project. Physiotherapy (United Kingdom). 2022;114(Supplement 1):e180.
3. Ajili IME, Haddar Y, Riahi A, Mhamdi S, Daboussi S, Aichaouia C, et al. Does being infected with COVID-19 have a psychological impact on healthcare workers? European Respiratory Journal. 2021;58(SUPPL 65).
4. Aksunger N, Janssens W, Pradhan M. The cost of a peaceful mind: Evidence from Kenya. Tropical Medicine and International Health. 2021;26(SUPPL 1):178.
5. Alhammad A, Aldardeer N, Alqahtani A, Alsaadon A, Alarifi M. Mental Health Assessment Among COVID-19 Patients Survivors of Critical Illness. Intensive Care Medicine Experimental. 2022;10(Supplement 29).
6. Andrade R, Sansone D, Farah D, Garcia R, Tannus G, Fonseca M. PRS17 What IF WE HAD NOT Done Any Isolation? Better Little THAN Nothing? Value in Health. 2020;23(Supplement 2):S719.
7. Andrade R, Sansone D, Farah D, Garcia R, Tannus G, Fonseca M. PMH16 Productivity, Depression, and Measures to Reduce Social IMPACT: The Economic IMPACT of COVID-19 in Brazil. Value in Health. 2020;23(Supplement 2):S587.
8. Andreo Jover J, Vidal-Villegas MP, Mediavilla R, Louzao Rojas I, Cebolla Lorenzo S, Fernandez Jimenez E, et al. Mental health service requirements after hospitalization due to COVID-19: a 1-year follow-up study. European Psychiatry. 2022;65(Supplement 1):S381.
9. Anonymous. A longitudinal tele-follow-up study regarding predictors of post-COVID functional status amongst patients who needed hospitalization. Indian Journal of Psychiatry. 2022;64(SUPPL 3):S667.
10. Araja D, Berkis U, Lunga A, Murovska M. Shadow Burden of Undiagnosed Myalgic Encephalomyelitis/Chronic Fatigue Syndrome (ME/CFS) on Society: Retrospective and Prospective-In Light of COVID-19. Journal of clinical medicine. 2021;10(14).
11. Avelino-Silva T, Garcez F, Dias M, Jacob-Filho W, Aliberti M. Fog ahead: delirium and post-discharge cognitive impairment in severe COVID-19. Journal of the American Geriatrics Society. 2022;70(SUPPL 1):S14-S5.
12. Ayed W, Chebbi S, Ayadi A, Ayari S, Hazem K, Magroun I. Psychological impact of the covid 19 pandemic on health care workers. European Psychiatry. 2023;66(Supplement 1):S458.
13. Ayed W, Chebbi S, Mosbeh M, Ayadi A, Ayari S, Magroun I. Effects of COVID-19 work circumstances on mental health. European Psychiatry. 2023;66(Supplement 1):S814.
14. Azevedo H, Santos N, Lafeta M, Souza V, Menezes T, Penido F, et al. Impact of COVID19 on work return after 6 months of hospitalization. European Respiratory Journal. 2022;60(Supplement 66).
15. Aziz R, Brode WM, Kelley MA, Alvarado NS, Melamed E. Characteristics of long COVID patients presenting to a dedicated post-COVID-19 clinic. Journal of General Internal Medicine. 2023;38(Supplement 2):S108.

16. Babnik K, Staresinic C, Lep Z. Some of the workforce face post COVID after the acute phase of the illness: The employer's supportive role. Human Systems Management. 2022;41(2):257-75.
17. Bajar E, Price O, Leitermann M, Garvin D, Oliveira C, Garavan J, Rooney C. Not for Publication A physiotherapy prospective observational study of post-intensive care COVID-19 patients in Tallaght University Hospital) 8.3 Post-Covid Assessment in a Specialist MDT Clinic: A Retrospective Review of Covid-19 Patients requiring High Dependency Oxygen Requirements or ventilation during Hospital Admission. Irish Journal of Medical Science. 2022;191(Supplement 5):S168-S9.
18. Bannister S. Neurological rehabilitation post Covid-19. Physiotherapy (United Kingdom). 2022;114(Supplement 1):e189-e90.
19. Bates A, Cusack R, Rushbrook S, Shapiro E, Golding H, Pattison N, et al. Can eye movement desensitisation and reprocessing improve psychological recovery following COVID-19 related critical illness? the CovEMERALD feasibility trial. Journal of the Intensive Care Society. 2023;24(1 Supplement):104-5.
20. Biehl M, Sarin K, Bishop E, Veith JM, Holztrager M, Bash K, et al. Aiding the recovery of ICU survivors: Creation and implementation of a post-ICU recovery clinic. American Journal of Respiratory and Critical Care Medicine. 2021;203(9).
21. Boparai S, Channa H, Dhaliwal L, Walter R, Motayar N. Risk factors associated with developing persistent symptoms after COVID-19: A retrospective study. Critical Care Medicine. 2022;50(1 SUPPL):88.
22. Borsi L, Waters A, Bodien Y, Bergin M, Boudreau N, Brown L, et al. Trajectory of Functional Recovery from 6 to 12 Months in Persons Hospitalized for Severe SARS-CoV-2 Illness. Archives of Physical Medicine and Rehabilitation. 2022;103(12):e138.
23. Bradley ER, Baron R, Brij SO, Khan WA. Advanced clinical practitioner (acp) hot airways clinic: a 12 month feasibility study. Thorax. 2022;77(Supplement 1):A140-A1.
24. Brinkley E, Mack C, Petruski-Ivleva N, Reynolds M, Bourke A, Hawaldar K, Dreyer NA. Patients tell all: Using a direct-to-patient community registry to understand patient burden of COVID-19. Pharmacoepidemiology and Drug Safety. 2021;30(SUPPL 1):98.
25. Brown M, Calvert S, James O. ICU follow-up clinic provides the opportunity to recognise and address symptoms post critical illness, which may otherwise have been missed. Journal of the Intensive Care Society. 2023;24(1 Supplement):59-60.
26. Brown M, Sanders S. Similar symptoms at 3 months post discharge in ICU, COVID-19 ICU, and COVID-19 patients that received non-invasive ventilation in high dependency areas. Journal of the Intensive Care Society. 2023;24(1 Supplement):53-4.
27. Chihaoui A, Saadaoui O, Haddad A, Hammouda Z, Maatouk I, Houri F, et al. Post-traumatic stress disorder in survivors of severe covid-19 infections. Annals of Intensive Care. 2023;13(Supplement 1).
28. Chirouze C, Bachelet D, Hulot JS, Cervantes-Gonzalez M, Goehringer F, Lemaignen A, et al. PERSISTENT COVID-19 SYMPTOMS ARE HIGHLY PREVALENT 12 MONTHS after HOSPITALIZATION. Topics in Antiviral Medicine. 2022;30(1 SUPPL):246-7.
29. Chisala E, Bahadur S. Meeting the demands of employment: the impact and rehabilitation of chronic pain and fatigue in the uk armed forces. Rheumatology (United Kingdom). 2022;61(SUPPL 1):i102.

30. Collins B, Humphrys M, Orford R, Ford R, Charles J. Health-related quality of life in long covid (post-covid-19 syndrome) service users in wales is much worse than the general population. Journal of Epidemiology and Community Health. 2023;77(Supplement 1):A102.

31. Diaz-Rodriguez C, Perez-Torres D, Canas-Perez I, Prieto-Lamo AM, Cuenca-Rubio C, Garcia-Garcia MM, et al. Referral to an inpatient rehabilitation facility evaluation of covid-19 patients after icu stay: A prospective cohort study assesing functional status 3 months after hospital discharge. Intensive Care Medicine Experimental. 2021;9(SUPPL 1).

32. Douglas DJ, Wolfe KS, Stutz MS, Pearson SD, Lecompte-Osorio P, Lin J, et al. One-Year Trajectories of Cognitive Impairment and Quality of Life in Critically Ill COVID-19 Survivors. American Journal of Respiratory and Critical Care Medicine. 2022;205(1).

33. Eilam-Stock T, George A, Lustberg M, Wolintz R, Krupp LB, Charvet LE. Telehealth transcranial direct current stimulation for recovery from Post-Acute Sequelae of SARS-CoV-2 (PASC). Brain Stimulation. 2021;14(6):1520-2.

34. Eltarras AM, Pan M, Chi R, Wang S, Czosnowski Q, Kruer R, et al. Post ICU Syndrome, Return to Employment, and Healthcare Utilization Among Critically Ill COVID-19 Survivors. American Journal of Respiratory and Critical Care Medicine. 2023;207(1).

35. Essafi F, Sdiri I, Talik I, Benismail K, Silini A, Moez K, Merhabene T. Three months post COVID: Respiratory and psychological sequelae. Intensive Care Medicine Experimental. 2021;9(SUPPL 1).

36. Fens T, Zon SKR, Rosmalen JGM, Brouwer S, van Asselt ADI. P4 Cost Analysis of Post-COVID-19 Healthcare Consumption in the Netherlands. Value in Health. 2022;25(12 Supplement):S1-S2.

37. Fki A, Sridi C, Hafsia M, Laaroussi R, Ksibi S. Work productivity and activity limitation in healthcare workers after COVID-19 infection. Allergy: European Journal of Allergy and Clinical Immunology. 2023;78(Supplement 111):346.

38. Gallegos M, Morgan ML, Burgos-Videla C, Caycho-Rodriguez T, Martino P, Cervigni M. The impact of long Covid on people's capacity to work. Annals of work exposures and health. 2023;67(7):801-4.

39. Ganesh A, Rosentreter R, Chen Y, Mehta R, McLeod G, Wan M, et al. Frequency, Persistence, and Patient-reported Outcomes of Neurological Symptoms in Mild COVID-19: Results from the ALBERTA HOPE COVID-19 Trial. Neurology. 2022;98(18 SUPPL).

40. Garner O, Kumari Narendra D, Guntupalli K. Characteristics of survivors of COVID-19 seen at a post-acute COVID-19 syndrome clinic. Chest. 2021;160(4 Supplement):A1141-A2.

41. Gatti S, Matteo P, Mauri F, Lucchini A, Bellin V, Bettini F, et al. Three-Months Outcome in COVID-19 Critical Care Patients: Results from a Follow-Up Study. Intensive Care Medicine Experimental. 2021;9(SUPPL 1).

42. Gaur A, Kukreja I, Mishra N, Roy A, Chopra A, Gupta A, et al. Risk of Psychiatric and Neurological Sequelae after the COVID-19 Infection: A Retrospective Cohort Study Using Claim Data. Value in Health. 2022;25(7 Supplement):S294.

43. Gilmartin M, Collins J, Mason S, McDermott G, Baily-Scanlan M, Hevey D, et al. Post-intensive care COVID survivorship clinic: a single centre experience. Critical Care. 2022;26(SUPPL 1).

44. Grewal S, Alismail A, Kim V, Monterroso P, Arias C, Patel P, Jeganathan N. Prevalence and characteristics of memory deficits in COVID-19 survivors. Critical Care Medicine. 2022;50(1 SUPPL):56.

45. Grossi MB, Liyanage-Don N, Nicholas C, Besagar S, Birger M, Freeman N, et al. COVID-19 patient needs after discharge home: A content analysis of medical records from a COVID remote care program. Journal of General Internal Medicine. 2021;36(SUPPL 1):S41.

46. Hamberger J, Jarczok MN, Beschoner P, Buckert D, Kersten J, Gundel H, Weimer K. Patients with functional post-covid symptoms show high utilization of the health care system. Psychosomatic Medicine. 2023;85(4):A124-A5.

47. Herrera E, González-Nosti M. The effect of age on cognitive impairment associated with post COVID-19 syndrome. Psychiatry Research. 2022;318.

48. Johns KN, Igboeli B, Ortiz-Cruzado E, Aase D, Stern S, Boxley L, et al. Ready for the Long Haul: Rapid Creation and Deployment of a Proactive, Modified Collaborative Care Program for COVID-19 Survivors with Behavioral Health Needs. Journal of the Academy of Consultation-Liaison Psychiatry. 2022;63(Supplement):S90-S1.

49. Kannan A, Murad A. Paediatric post covid-19 condition: experience of a northern paediatric post covid-19 assessment centre. Archives of Disease in Childhood. 2022;107(Supplement 2):A121.

50. Karalasingham K, Chew D, Hira R, Siddiqui T, Ranada S, Bourne K, et al. Post-acute sequelae of COVID-19 (PASC) patients experience losses of work productivity. Clinical Autonomic Research. 2022;32(5):365-6.

51. Karim MA, Wadoo O, Reagu SM, Amro R, Abdulla MA. Telepsychiatry in the Arabian Gulf region-Implications beyond the COVID-19 pandemic. Asian Journal of Psychiatry. 2020;54.

52. Kean K, Amato C, Eskin B, Allegra JR. Shifting trends in emergency department pediatric psychiatric visits after the arrival of COVID-19. Academic Emergency Medicine. 2021;28(SUPPL 1):S128-S9.

53. Khan K, Hong S, Larmour K, Hussain S. Neurocognitive manifestations of post-COVID syndrome. Critical Care Medicine. 2022;50(1 SUPPL):97.

54. Kommaraju K, Biehl M, Bishop E, Veith J, Sarin K, O'Brien J, et al. A description of post intensive care syndrome in COVID19 survivors. American Journal of Respiratory and Critical Care Medicine. 2021;203(9).

55. Krotz A, Sosnowsky-Waschek N, Bechtel S, Neumann C, Lohkamp M, Kovacs G, et al. Reducing sick leave, improving work ability, and quality of life in patients with mild to moderate Long COVID through psychosocial, physiotherapeutic, and nutritive supportive digital intervention (MiLoCoDaS): study protocol for a randomized controlled trial. Trials. 2023;24(1).

56. Kumar P, Nyberg S, Jiwan R, Ainley A. Post-COVID and the BHRUT one-stop shop MDT approach to managing the longterm sequale of a novel disease process. European Respiratory Journal. 2021;58(SUPPL 65).

57. Kupferschmitt A, Hinterberger T, Montanari I, Gasche M, Hermann C, Jobges M, et al. Relevance of the post-COVID syndrome within rehabilitation (PoCoRe): study protocol of a multi-centre study with different specialisations. BMC psychology. 2022;10(1):189.

58. Lagali N, Setterud H, Meijer H, Andersson G, Johansson B. Postcovid eye syndrome: ocular symptoms and anterior segment findings. Investigative Ophthalmology and Visual Science. 2023;64(8):1171.

59. Lee J, Sim M, Lee D, Hyun S, Kim CH, Kim W. The outcomes of mental health assessments among COVID-19 patients in South Korea. Asia-Pacific Psychiatry. 2021;13(SUPPL 1).

60. Lemogne C, Pitron V. Shedding light on the work burden of long COVID. The Lancet Regional Health - Europe. 2023;31.

61. Liao Y, Tian H, He X, Chen W, Jiang W. The effect of COVID-19 on the working and psychological status of medical staffs in organ donation and transplantation. Transplantation. 2020;104(SUPPL 3):S114.

62. Liccari M, Furlanis G, Michelutti M, Manganotti P. Neuro-long-covid: a proposal for the management of memory impairment and cognitive deficits in a neurology ambulatory service. Neurological Sciences. 2022;43(Supplement 1):S325.

63. Ling J, Ilchef R. Consultation-liaison psychiatry referrals over 2 years of the covid-19 pandemic. Australian and New Zealand Journal of Psychiatry. 2022;56(SUPPL 1):242.

64. Lugosi S, Sanchez S, Verduzco-Gutierrez M. Letter to the Editor on "Race, ethnicity, and utilization of outpatient rehabilitation for treatment of post COVID-19 condition". PM and R. 2023;15(7):932-3.

65. Lunt J, Hemming S, Elander J, Burton K, Hanney B. Sustaining work ability amongst female professional workers with long COVID. Occupational Medicine. 2024;74(1):104-12.

66. Makam A, Burnfield J, Prettyman E, Votto J. Recovery After Transfer to an LTCH for COVID-19 (RAFT COVID Study): A National Multicenter Cohort. Journal of the American Geriatrics Society. 2022;70(SUPPL 1):S132.

67. Martin A, Thomson K, LePrade A, Bertolli J, Grosse SD. PCR61 Persistent Work Limitations in Long COVID: A Cross-Sectional Analysis of Employed US Respondents to the 2022 National Health and Wellness Survey. Value in Health. 2023;26(6 Supplement):S322.

68. Masserini F, Pomati S, Cucumo V, Boscacci M, Scarpa C, Dell'Osso B, Pantoni L. Persistent cognitive complaints after COVID-19: a proxy for a maladaptive response to an applied stressor in the unique pandemic environment? Neurological Sciences. 2022;43(Supplement 1):S328.

69. Mc Closkey M, Mc Geady M, Laverty R, Khan H, King C, Kelly MG, Sharkey R. Follow up of patients discharged from a Respiratory High Dependency Unit after Covid 19 infection. Irish Journal of Medical Science. 2022;191(Supplement 5):S170.

70. Mera-Cordero F, Bonet-Monne S, Almeda-Ortega J, Garcia-Sangenis A, Cunillera-Puertolas O, Contreras-Martos S, et al. Double-blind placebo-controlled randomized clinical trial to assess the efficacy of montelukast in mild to moderate respiratory symptoms of patients with long COVID: E-SPERANZA COVID Project study protocol. Trials. 2022;23(1):19.

71. Millet C, Jimenez H, Racoosin E, Horani G, Shamoon Y, Narvaneni S, et al. The Racial Divide: A Follow Up Study on Racial Disparity Amongst COVID-19 Survivors in an Urban Community in New Jersey. Open Forum Infectious Diseases. 2021;8(SUPPL 1):S330-S1.

72. Mirin AA, Dimmock ME, Jason LA. Updated ME/CFS prevalence estimates reflecting post-COVID increases and associated economic costs and funding implications. Fatigue: Biomedicine, Health and Behavior. 2022;10(2):83-93.

73. Mohammadi N, Reford E, Spica NR, Tosto J, Tabacof LR, Putrino D, Kellner C. Covid-19 Associated Extreme Fatigue Effects on Activities of Daily Living in Mild and Long-Haul Infections: Patient Perspectives from a Remote Patient Monitoring Program. Neurology. 2022;98(18 SUPPL).

74. Monne SB, Cordero FM, Munoz GA, Ortega JA, Martos SC, Puertolas OC, et al. Double-blind placebo-controlled randomized clinical trial to assess the efficacy of montelukast in mild to moderate respiratory symptoms of patients with long COVID: E-speranza COVID-19 Project study protocol. Basic and Clinical Pharmacology and Toxicology. 2022;130(SUPPL 2):40.

75. Monteith T, Cocores A, Mukhtarzada M, Rincon N, Encarnacion YC, Kursewicz C, et al. Headache as an Early Symptom and Post-Acute Sequela of COVID19 in Hospitalized COVID19 Survivors. Neurology. 2022;98(18 SUPPL).

76. Munblit D, Simpson F, Mabbitt J, Dunn-Galvin A, Semple C, Warner JO. Legacy of COVID-19 infection in children: Long-COVID will have a lifelong health/economic impact. Archives of Disease in Childhood. 2022;107(3):E2.

77. Murdock ME, Kronish IM, Cornelius T, Jurado A, Quispe K, Sullivan A, Liyanage-Don NA. Association between COVID-related psychosocial stressors and mental health outcomes among patients recovering from COVID illness. Psychosomatic Medicine. 2021;83(7):A9-A10.

78. Nucera A, Vazzana MR, Papa M. Post-CoViD neurological sequelae. Italian Journal of Medicine. 2021;15(3):52.

79. Owen R, Ashton RE, Skipper L, Phillips BE, Yates J, Thomas C, et al. Long COVID quality of life and healthcare experiences in the UK: a mixed method online survey. Quality of life research : an international journal of quality of life aspects of treatment, care and rehabilitation. 2023.

80. Padda K, Re'em Y, Faecher D. (PO-071) A Group Psychotherapy for Patients with Post-Acute Sequelae of COVID-19 (PASC) Experiencing Psychological Stress. Journal of the Academy of Consultation-Liaison Psychiatry. 2022;63(Supplement):S34-S5.

81. Palazzo M, Prodi TG, Cerioli M, Conti D, Galbassini A, Nicolini G, Dell'Osso B. Telemedicine in Psychiatry: benefits and challenges of the the Home-Hospital Care system (COD20) project. European Psychiatry. 2023;66(Supplement 1):S526-S7.

82. Papadopoulou A, Efstathiou V, Koliou E, Papazachos K, Barbari A, Kollia N, et al. Clinical and demographic characteristics of hospitalized patients with COVID-19 referred to a Consultation - Liaison Psychiatry Unit. European Psychiatry. 2023;66(Supplement 1):S207.

83. Park J, Brady B. CO195 Incidence and Persistence of Post-COVID-19 Conditions in the Year Following Initial Diagnosis. Value in Health. 2023;26(6 Supplement):S51-S2.

84. Patel G, Dadey E, Gosal E, O'Neill T, Skyllberg EW, Calderwood CJ, et al. Clinical characteristics, mortality and short term follow up of patients admitted with covid-19 in a North East London NHS Trust: A retrospective analysis. Thorax. 2021;76(SUPPL 1):A5.

85. Pauwels S BI, Polli A, Mylle G, De Raeve H, Godderis L. Return to work after long COVID - an evidence review. 2021.

86. Petry Moecke DM, Kwong E, Yao J, Turner J, McLean K, Den Breejen S, et al. Rehabilitation Needs of Adult British Columbians With Long COVID. American Journal of Respiratory and Critical Care Medicine. 2023;207(1).

87. Raut P, Jadhav N, Kale A. Effects of Preventive Health Interventions on COVID Positive Employees of Large Commercial Vehicle Manufacturing Company. Indian Journal of Occupational and Environmental Medicine. 2023;27(1):101.

88. Reford E, Mohammadi N, Spica NR, Tosto J, Tabacof LR, Putrino D, Kellner C. Post Covid-19 Neurological Impairment Effects on Activities of Daily Living: Patient Perspectives from a Remote Patient Monitoring Program. Neurology. 2022;98(18 SUPPL).

89. Rinaldi L, Pellizon F, Rigo S, Minonzio M, Romeo MA, Bisoglio A, et al. Relationships between autonomic symptoms and work ability in post-COVID19 autonomic syndrome. Clinical Autonomic Research. 2022;32(5):367.

90. Roberts L, Kozlowska W. The difficult asthma multi-disciplinary clinic: Silver linings of the COVID cloud. Archives of Disease in Childhood. 2021;106(SUPPL 1):A85.

91. Roesch Ely D, Kramer B, Jaehn A, Merle U, Weisbrod M. Post-Covid Syndrome: A standardized assessment on subjective psychiatric and neuropsychological symptoms. European Psychiatry. 2022;65(Supplement 1):S530-S1.

92. Rohde G, Helseth S, Smastuen MC, Mikkelsen HET, Skarstein S, Haraldstad K. Changes in health-related quality of life in parents of adolescents, and the impact of gender, sociodemographic and psychological factors, and pain on the changes: A two-year longitudinal study. Quality of Life Research. 2022;31(Supplement 2):S89-S90.

93. Roseanne MJ, Thant T, Murray HJ, Dillon J, Golub M. (PO-013) Creating an Effective Clinic Model for Post-COVID Mental Health Treatment. Journal of the Academy of Consultation-Liaison Psychiatry. 2022;63(Supplement):S7.

94. Saigal A, Naidu SB, Shah AJ, Brill SE, Jarvis H, Goldring JG, et al. 'Long-COVID': The need for multi-disciplinary working. Thorax. 2021;76(SUPPL 1):A33-A4.

95. Saigal A, Shah AJ, Naidu SB, Brown J, Goldring JG, Sood T, et al. Ward vs. Emergency department discharge in patients with COVID-19: Does it make a difference to symptom burden and radiological severity at follow up? Thorax. 2021;76(SUPPL 1):A182.

96. Salvador G, Nogueira M, Silva L, Aventurado I, Cendes F, Bandini M, et al. The Analysis of 607 Volunteers Shows a Reduction of Workability in Individuals With LONG-COVID Syndrome Associated With Neuropsychiatric Symptoms. Neurology. 2023;100(17 Supplement 2).

97. Santos RAG, Rodriguez Rodriguez M. Cognitive impairment detected by MoCA (Montreal Cognitive Assessment) test after COVID-19 in Mexico. European Journal of Neurology. 2021;28(SUPPL 1):450-1.

98. Schamess A, Woschkolup K, Rush LJ, McConnell E, Friedberg A, Saigal T, et al. Three-month outcomes for patients treated in an academic medical center long COVID clinic for cognitive symptoms. Journal of General Internal Medicine. 2023;38(Supplement 2):S132-S3.

99. Seyffert SA, Khan SH, Jamil Y, Lindroth HL, Wang S, Khan BA. Beyond the ICU: The association between post intensive care syndrome and survivors of severe COVID-19. American Journal of Respiratory and Critical Care Medicine. 2021;203(9).

100. Shadwell C, Vardy E. Follow-up of older people hospitalised with COVID-19 infection: A single centre study. European Geriatric Medicine. 2022;13(Supplement 1):S172.

101. Shams-Khan H, Nwosu N, Watkins L, Tack G. Employment status of referrals to a regional Long COVID service in the United Kingdom. European Respiratory Journal. 2022;60(Supplement 66).

102. Shamsutdinova N, Lapshina S, Mukhamadieva V, Abdrakipov R, Sukhorukova E, Kupkenova L, Abdulganieva D. Clinical characteristics of post-COVID syndrome in patients with rheumatic diseases in the Republic of Tatarstan. Annals of the Rheumatic Diseases. 2022;81(Supplement 1):1695-6.

103. Shechter A, Abdalla M, Raju D, Kronish I, Liyanage-Don N. COVID-19 related worries and sleep disturbances in patients previously hospitalized with COVID-19 illness. Sleep. 2021;44(SUPPL 2):A93.

104. Shinde V, Master M, Rajesh R. Post COVID Syndrome in patients with COVID -19 : A Cross-Sectional study. Safety and Health at Work. 2022;13(Supplement):S121.

105. Siebler M, Moessinger B, Schmalenbach M, Bonnert J, Waldmann G, Lammers M, et al. Beneficial effects of interdisciplinary stationary rehabilitation in employed and retired patients with the Post-Covid-Syndrome. Neurologie und Rehabilitation. 2022;28(Supplement 1):S43.

106. Stolyarova A, Tyvina N, Golovkina D, Vysokova V, Zhao J, Zhang K, Kinkulkina M. Anxiety and depressive disorders in patients with Covid-19. European Psychiatry. 2023;66(Supplement 1):S339-S40.

107. Stufano A, Bonfrate L, Portincasa P, Dell'Erba A, Lovreglio P. Long covid and gender differences: implications for return to work. European Journal of Clinical Investigation. 2022;52(Supplement 1):175.

108. Tak CR, Landgren K, Hughes P, Ramage M. Long COVID and Mental Health: An Assessment of Health Status and Service Utilization. Journal of Mental Health Policy and Economics. 2023;26(Supplement 1):S31.

109. Thiery G, Laurent R, Correia P, Ezingeard E, Eric D, Lachand R, et al. One-year follow-up of patients and relatives after severe COVID-19 related ARDS treated in the intensive care unit. Annals of Intensive Care. 2022;12(Supplement 1).

110. Thomas S, Frater J, Davies-Cotter D, Watters A, Lane A. Screening and referral patterns for raised edinburgh postnatal depression scores at mater mother's hospital: current practices and impact of the SARS COVID-19 pandemic on routine care. Australian and New Zealand Journal of Psychiatry. 2022;56(SUPPL 1):95-6.

111. Tileubek N, Shamsutdinova N, Mukhamadieva V, Abdulganieva D, Abdrakipov R, Lapshina S. The incidence of post-COVID syndrome in patients with rheumatic diseases. Annals of the Rheumatic Diseases. 2023;82(Supplement 1):1879.

112. Topeli A, Halacli B, Ortac Ersoy E, Kilic S, S OC. Short-term physical and mental health outcome of critically-ill COVID-19 patients after discharge from intensive care unit. Intensive Care Medicine Experimental. 2020;8(SUPPL 2).

113. Van Voorthuizen E, Peters J, Vercoulen J, Van Den Heuvel M, Van Helvoort H, Van Hees J, Van Den Borst B. Recovering after COVID-19: 10- month follow-up data from a multidisciplinary outpatient clinic. European Respiratory Journal. 2021;58(SUPPL 65).

114. Vink M, Vink-Niese F. Is It Useful to Question the Recovery Behaviour of Patients with ME/CFS or Long COVID? Healthcare (Basel, Switzerland). 2022;10(2).

115. Weakley KE, Schikler A, Green JV, Blatt DB, Barton SM, Statler VA, Marshall GS. Clinical Features and Long-Term Follow-up of Children Evaluated for Persistent Unwellness Following Acute COVID-19. Open Forum Infectious Diseases. 2022;9(Supplement 2):S459-S60.

116. Weidemann DK, Redpath AC, Ashoor I. COVID-19 financial ramifications on the pediatric nephrology workforce. Journal of the American Society of Nephrology. 2020;31:808.

117. Weimer K, Hamberger J, Beschoner P, Buckert D, Kersten J, Gundel H, Jarczok MN. Patients with functional post-COVID symptoms show relevant scores and prevalence in psychological screening questionnaires. Psychosomatic Medicine. 2023;85(4):A42.

118. White KR, Osman F, Piscitello G, Gerhart J, Greenberg JA. Associations Between COVID-19, Unexpectedness of Death, and Symptoms of PTSD Among Bereaved ICU Surrogates. American Journal of Respiratory and Critical Care Medicine. 2023;207(1).

119. Wildi K, LiBassi G, McNicholas B, Rainieri F, Forsyth S, Cho S, et al. Long-Term Impact in Intensive Care Survivors of Coronavirus Disease- 19 (AFTERCOR Study): Preliminary 3-Month Analysis After ICU Discharge. American Journal of Respiratory and Critical Care Medicine. 2022;205(1).

120. Williams MK, Crawford CA, Zapolski TC, Hirsh AT, Stewart JC. Longer-term mental health consequences of COVID-19 infection: moderation by race and socioeconomic status. Psychosomatic Medicine. 2023;85(4):A123-A4.

121. Wolfe KS, Douglas DJ, Pearson SD, Stutz MR, Lecompte-Osorio PA, Lin J, et al. Functional and quality of life outcomes of critically ill covid-19 survivors at hospital discharge and six months. American Journal of Respiratory and Critical Care Medicine. 2021;203(9).

122. Womer J, Sarma N, Hauschildt K, Caldwell E, Admon AJ, Hough CL, Iwashyna TJ. Financial Toxicity Is Associated With Impaired Recovery After Severe COVID-19: Results From the BLUE-CORAL Study. American Journal of Respiratory and Critical Care Medicine. 2023;207(1).

123. Wu Q. Understanding the burden of post-covid-19 condition. BMJ. 2023.

124. Wynne-Jones G, Chew-Graham C. Why GPs must not lose their role in supporting people back to work. British Journal of General Practice. 2022;72(717):174.

125. Yus-Fuertes M, Diez-Cirarda M, Matias-Guiu JA, Gil-Martinez L, Gomez-Ruiz N, Polidura-Arruga C, et al. MR assessment of brain alterations in post-COVID cognitive syndrome. Neuroradiology. 2022;64(Supplement 1):S84.

126. Zaidan MF, A PND, Polychronopoulou E, P. Nishi S, G DA, Kuo Yongfang, Sharma G. Health care utilization among patients discharged alive post-COVID-19 hospitalization. Chest. 2022;162(4 Supplement):A519.

127. Zhong Y, Ji Q, Zhou L. Sleep disorders of post-COVID-19 conditions. Sleep and Breathing. 2023;27(6):2435-6.

128. Ziauddeen N, Gurdasani D, O'Hara ME, Hastie C, Roderick P, Yao G, Alwan NA. Characteristics of long COVID: findings from a social media survey. Journal of Epidemiology and Community Health. 2021;75(Supplement 1):A90.

129. Zima BT, Bussing R. The early impact of the COVID-19 pandemic on child mental health service utilization and disparities in care. Journal of the American Academy of Child and Adolescent Psychiatry. 2021;60(10 Supplement):S295-S6.

**No outcome of interest (n=70)**

1. Abd El-Khalik DM, Eltohamy M. Post-COVID-19 depression and anxiety in patients with systemic lupus erythematosus. Lupus. 2023;32(8):974-82.

2. Abramoff BA, Dillingham TR, Brown LA, Caldera F, Caldwell KM, McLarney M, et al. Psychological and Cognitive Functioning Among Patients Receiving Outpatient Rehabilitation for Post-COVID Sequelae: An Observational Study. Archives of Physical Medicine and Rehabilitation. 2023;104(1):11-7.

3. Ali ST, Kang AK, Patel TR, Clark JR, Perez-Giraldo GS, Orban ZS, et al. Evolution of neurologic symptoms in non-hospitalized COVID-19 "long haulers". Annals of clinical and translational neurology. 2022;9(7):950-61.

4. Alqutub S, Mahmoud M, Baksh T. Psychological Impact of COVID-19 on Frontline Healthcare Workers in Saudi Arabia. Cureus. 2021;13(5):e15300.

5. Angeles MR, Wanni Arachchige Dona S, Nguyen HD, Le LK-D, Hensher M. Modelling the potential acute and post-acute burden of COVID-19 under the Australian border re-opening plan. BMC public health. 2022;22(1):757.

6. Ashktorab H, Challa SR, Singh G, Nanduri S, Ibrahim M, Martirosyan Z, et al. Gastrointestinal Manifestations and Their Association with Neurologic and Sleep Problems in Long COVID-19 Minority Patients: A Prospective Follow-Up Study. Digestive diseases and sciences. 2023.

7. Bahmer T, Borzikowsky C, Lieb W, Horn A, Krist L, Fricke J, et al. Severity, predictors and clinical correlates of Post-COVID syndrome (PCS) in Germany: A prospective, multi-centre, population-based cohort study. EClinicalMedicine. 2022;51:101549.

8. Becker JH, Lin JJ, Doernberg M, Stone K, Navis A, Festa JR, Wisnivesky JP. Assessment of Cognitive Function in Patients After COVID-19 Infection. JAMA network open. 2021;4(10):e2130645.

9. Bek LM, Berentschot JC, Hellemons ME, Remerie SC, van Bommel J, Aerts JGJV, et al. Return to work and health-related quality of life up to 1 year in patients hospitalized for COVID-19: the CO-FLOW study. BMC medicine. 2023;21(1):380.

10. Bernacki EJ, Hunt DL, Tsourmas NF, Yuspeh L, Lavin RA, Kalia N, et al. Attributes of Long Duration COVID-19 Workers' Compensation Claims. J Occup Environ Med. 2022;64(5):e327-e32.

11. Boesl F, Audebert H, Endres M, Pruss H, Franke C. A Neurological Outpatient Clinic for Patients With Post-COVID-19 Syndrome - A Report on the Clinical Presentations of the First 100 Patients. Frontiers in neurology. 2021;12:738405.

12. Braig S PR, Nieters A, Kräusslich HG, Brockmann SO, Göpel S, Kindle G, Merle U, Steinacker JM, Kern WV, Rothenbacher D. Post-COVID syndrome and work ability 9-12 months after a SARS-CoV-2 infection among over 9000 employees from the general population. IJID Regions. 2024;10:67-74.

13. Buonsenso D, Gualano MR, Rossi MF, Valz Gris A, Sisti LG, Borrelli I, et al. Post-Acute COVID-19 Sequelae in a Working Population at One Year Follow-Up: A Wide Range of Impacts from an Italian Sample. International journal of environmental research and public health. 2022;19(17).

14. Calabria M, Garcia-Sanchez C, Grunden N, Pons C, Arroyo JA, Gomez-Anson B, et al. Post-COVID-19 fatigue: the contribution of cognitive and neuropsychiatric symptoms. Journal of neurology. 2022;269(8):3990-9.

15. Chieffo DPR, Delle Donne V, Massaroni V, Mastrilli L, Belella D, Monti L, et al. Psychopathological profile in COVID-19 patients including healthcare workers: the implications. European review for medical and pharmacological sciences. 2020;24(22):11964-70.

16. Cohen K, Ren S, Heath K, Dasmarinas MC, Jubilo KG, Guo Y, et al. Risk of persistent and new clinical sequelae among adults aged 65 years and older during the post-acute phase of SARS-CoV-2 infection: Retrospective cohort study. The BMJ. 2022;376:e068414.

17. Crivelli L, Calandri I, Corvalan N, Carello MA, Keller G, Martinez C, et al. Cognitive consequences of COVID-19: results of a cohort study from South America. Arquivos de neuro-psiquiatria. 2022;80(3):240-7.

18. Davisse-Paturet C, Orri M, Legleye S, Florence AM, Hazo JB, Warszawski J, et al. Suicidal ideation following self-reported COVID-19-like symptoms or serology-confirmed SARS-CoV-2 infection in France: A propensity score weighted analysis from a cohort study. PLoS Medicine. 2023;20(2):e1004171.

19. Dawra S, Shrivastava S, Chauhan VS, Asturkar V, Ahmad F, Kumar A, et al. The psychological impact of COVID-19 among newly diagnosed patients: COVID Impact study. Medical journal, Armed Forces India. 2021;77:S333-S7.

20. Diez-Cirarda M, Yus M, Gomez-Ruiz N, Polidura C, Gil-Martinez L, Delgado-Alonso C, et al. Multimodal neuroimaging in post-COVID syndrome and correlation with cognition. Brain : a journal of neurology. 2023;146(5):2142-52.

21. Efgan MG, Cinaroglu OS, Payza U, Kanter E, Bilgin S. Comparison of suicide attempt cases admitted to emergency services during COVID-19 and before Comparison of suicide attempts during and before COVID-19. Annals of Clinical and Analytical Medicine. 2023;14(Supplement 2):S165-S70.

22. Faghy MA, Maden-Wilkinson T, Arena R, Copeland RJ, Owen R, Hodgkins H, Willmott A. COVID-19 patients require multi-disciplinary rehabilitation approaches to address persisting symptom profiles and restore pre-COVID quality of life. Expert review of respiratory medicine. 2022;16(5):595-600.

23. Ferrando SJ, Dornbush R, Lynch S, Shahar S, Klepacz L, Karmen CL, et al. Neuropsychological, Medical, and Psychiatric Findings After Recovery From Acute COVID-19: A Cross-sectional Study. Journal of the Academy of Consultation-Liaison Psychiatry. 2022;63(5):474-84.

24. Frontera JA, Sabadia S, Lalchan R, Fang T, Flusty B, Millar-Vernetti P, et al. A Prospective Study of Neurologic Disorders in Hospitalized Patients With COVID-19 in New York City. Neurology. 2021;96(4):e575-e86.

25. Fung KW, Baye F, Baik SH, Zheng Z, McDonald CJ. Prevalence and characteristics of long COVID in elderly patients: An observational cohort study of over 2 million adults in the US. PLoS Medicine. 2023;20(4):e1004194.

26. Furlanis G, Buoite Stella A, Biaduzzini F, Bellavita G, Frezza NA, Olivo S, et al. Cognitive deficit in post-acute COVID-19: an opportunity for EEG evaluation? Neurological sciences : official journal of the Italian Neurological Society and of the Italian Society of Clinical Neurophysiology. 2023;44(5):1491-8.

27. Gaber TAZK, Ashish A, Unsworth A. Persistent post-covid symptoms in healthcare workers. Occupational medicine (Oxford, England). 2021;71(3):144-6.

28. Ganesh A, Rosentreter RE, Chen Y, Mehta R, McLeod GA, Wan MW, et al. Patient-reported outcomes of neurologic and neuropsychiatric symptoms in mild COVID-19: a prospective cohort study. CMAJ open. 2023;11(4):E696-E705.

29. Garg A, Subramain M, Barlow PB, Garvin L, Hoth KF, Dukes K, et al. Patient experience with healthcare: Feedback for a 'post COVID-19 clinic' at a tertiary care center in rural area. medRxiv. 2021.

30. Goodman ML, Molldrem S, Elliott A, Robertson D, Keiser P. Long COVID and mental health correlates: a new chronic condition fits existing patterns. Health psychology and behavioral medicine. 2023;11(1):2164498.

31. Graham EL, Clark JR, Orban ZS, Lim PH, Szymanski AL, Taylor C, et al. Persistent neurologic symptoms and cognitive dysfunction in non-hospitalized Covid-19 "long haulers". Annals of clinical and translational neurology. 2021;8(5):1073-85.

32. Halpin SJ, McIvor C, Whyatt G, Adams A, Harvey O, McLean L, et al. Postdischarge symptoms and rehabilitation needs in survivors of COVID-19 infection: A cross-sectional evaluation. J Med Virol. 2021;93(2):1013-22.

33. Hirahata K, Nawa N, Fujiwara T. Characteristics of Long COVID: Cases from the First to the Fifth Wave in Greater Tokyo, Japan. Journal of Clinical Medicine. 2022;11(21):6457.

34. Iob E, Steptoe A, Zaninotto P. Mental health, financial, and social outcomes among older adults with probable COVID-19 infection: A longitudinal cohort study. PNAS Proceedings of the National Academy of Sciences of the United States of America. 2022;119(27):1-9.

35. Jacob L, Koyanagi A, Smith L, Tanislav C, Konrad M, van der Beck S, Kostev K. Prevalence of, and factors associated with, long-term COVID-19 sick leave in working-age patients followed in general practices in Germany. International journal of infectious diseases : IJID : official publication of the International Society for Infectious Diseases. 2021;109:203-8.

36. Jain A, Gupta P, Mittal AA, Sengar NS, Chaurasia R, Banoria N, et al. Long-term quality of life and work ability among severe COVID-19 survivors: A multicenter study. Dialogues in health. 2023;2:100124.

37. Jakobsen KD, O'Regan E, Svalgaard IB, Hviid A. Machine learning identifies risk factors associated with long-term sick leave following COVID-19 in Danish population. Communications medicine. 2023;3(1):188.

38. Jaquet P, Legouy C, Le Fevre L, Grinea A, Sinnah F, Franchineau G, et al. Neurologic Outcomes of Survivors of COVID-19-Associated Acute Respiratory Distress Syndrome Requiring Intubation. Critical care medicine. 2022;50(8):e674-e82.

39. Kaplan M, Çamurcu AA, Erol S. Post-Coronavirus Disease-2019 Syndrome in Healthcare Workers COVID-19 Enfeksiyonu Geçiren Sağlık Çalışanlarında Post-COVID-19 Sendromunun İncelenmesi. Mediterranean Journal of Infection, Microbes and Antimicrobials. 2022;11.

40. Khan M, Majeed S, Ain QT, Nawaz A, Sumra KA, Lammi V, et al. Analysing the psychosocial and health impacts of Long COVID in Pakistan: A cross sectional study. medRxiv. 2023.

41. Khanna SK, Khanna N, Malav MK, Bayad HC, Sood A, Abraham L. Profiling Cognitive Impairment in Mild COVID-19 Patients: A Case-Control Study at a Secondary Healthcare Centre in the Hilly Region of North India. Annals of Indian Academy of Neurology. 2022;25(6):1099-103.

42. Kikkenborg Berg S, Dam Nielsen S, Nygaard U, Bundgaard H, Palm P, Rotvig C, Vinggaard Christensen A. Long COVID symptoms in SARS-CoV-2-positive adolescents and matched controls (LongCOVIDKidsDK): a national, cross-sectional study. The Lancet Child and Adolescent Health. 2022;6(4):240-8.

43. Kim S-G, Kwon HC, Kang TK, Kwak MY, Lee S, Lee K, Ko K. COVID-19 Sequelae and Their Implications on Social Services. Journal of Korean medical science. 2022;37(48):e342.

44. Kitsios GD, Blacka S, Jacobs J, Mirza T, Naqvi A, Gentry H, et al. Subphenotypes of Self-Reported Symptoms and Outcomes in Long COVID: a prospective cohort study with latent class analysis. medRxiv. 2023.

45. Kumar NS, Rose D, Amrutha MK, Babu S, Almeda ND, Dileep R. A retrospective observational study to assess the clinical manifestations and functioning status among covid-19 survivors with and without post-covid syndrome. International Journal of Pharmaceutical Sciences and Research. 2023;14(8):3972-7.

46. Kwon J, Milne R, Rayner C, Rocha Lawrence R, Mullard J, Mir G, et al. Impact of Long COVID on productivity and informal caregiving. European Journal of Health Economics. 2023.

47. Lear-Claveras A, Aguilar-Latorre A, Olivan-Blazquez B, Couso-Viana S, Claveria-Fontan A. Evolution of Anxiety and Depression in Men during the First Six Months of the COVID-19 Pandemic and Factors Associated with Worsening of Mental Health: Retrospective Longitudinal Study. Journal of Men's Health. 2022;18(9):jomh1809182.

48. Miraglia Del Giudice M, Klain A, Dinardo G, D'Addio E, Bencivenga CL, Fontanella C, et al. Behavioral and Sleep Disorders in Children and Adolescents following COVID-19 Disease: A Case-Control Study. Children (Basel, Switzerland). 2023;10(7).

49. Nielsen TB, Leth S, Pedersen M, Harbo HD, Nielsen CV, Laursen CH, et al. Mental Fatigue, Activities of Daily Living, Sick Leave and Functional Status among Patients with Long COVID: A Cross-Sectional Study. International journal of environmental research and public health. 2022;19(22).

50. Nishimi K, Neylan TC, Bertenthal D, Seal KH, O'Donovan A. Association of psychiatric disorders with clinical diagnosis of long COVID in US veterans. Psychol Med. 2024:1-9.

51. O'Brien K, Townsend L, Dowds J, Bannan C, Nadarajan P, Kent B, et al. 1-year quality of life and health-outcomes in patients hospitalised with COVID-19: a longitudinal cohort study. Respir Res. 2022;23(1):115.

52. O'Keefe JB, Minton HC, Morrow M, Johnson C, Moore MA, O'Keefe GAD, et al. Postacute Sequelae of SARS-CoV-2 Infection and Impact on Quality of Life 1-6 Months After Illness and Association With Initial Symptom Severity. Open forum infectious diseases. 2021;8(8):ofab352.

53. Palladini M, Bravi B, Colombo F, Caselani E, Di Pasquasio C, D'Orsi G, et al. Cognitive remediation therapy for post-acute persistent cognitive deficits in COVID-19 survivors: A proof-of-concept study. Neuropsychological rehabilitation. 2023;33(7):1207-24.

54. Pihlaja RE, Kauhanen L-LS, Ollila HS, Tuulio-Henriksson AS, Koskinen SK, Tiainen M, et al. Associations of subjective and objective cognitive functioning after COVID-19: A six-month follow-up of ICU, ward, and home-isolated patients. Brain, behavior, & immunity - health. 2023;27:100587.

55. Price E, Hollis N, Salganik J, Lykke M, Paolinelli C, Chamovitz S, et al. Implementing a Multidisciplinary Post-COVID Clinic in a Small Community Environment. Archives of rehabilitation research and clinical translation. 2023;5(3):100270.

56. Rabaiotti P, Ciraci C, Donelli D, Oggioni C, Rizzi B, Savi F, et al. Effects of Multidisciplinary Rehabilitation Enhanced with Neuropsychological Treatment on Post-Acute SARS-CoV-2 Cognitive Impairment (Brain Fog): An Observational Study. Brain sciences. 2023;13(5).

57. Shahar S, Lynch S, Dornbush R, Klepacz L, Smiley A, Ferrando SJ. Frequency and Characteristics of Depression and Its Association with Diminished Quality of Life in a Cohort of Individuals with Post-Acute Sequelae of COVID-19. Neuropsychiatric disease and treatment. 2023;19:2069-79.

58. Shanley JE, Valenciano AF, Timmons G, Miner AE, Kakarla V, Rempe T, et al. Longitudinal evaluation of neurologic-post acute sequelae SARS-CoV-2 infection symptoms. Annals of clinical and translational neurology. 2022;9(7):995-1010.

59. Simonetti A, Bernardi E, Janiri D, Mazza M, Montanari S, Catinari A, et al. Suicide Risk in Post-COVID-19 Syndrome. Journal of personalized medicine. 2022;12(12).

60. Taquet M, Geddes JR, Husain M, Luciano S, Harrison PJ. 6-month neurological and psychiatric outcomes in 236 379 survivors of COVID-19: a retrospective cohort study using electronic health records. Lancet Psychiatry. 2021;8(5):416-27.

61. Taquet M, Luciano S, Geddes JR, Harrison PJ. Bidirectional associations between COVID-19 and psychiatric disorder: retrospective cohort studies of 62 354 COVID-19 cases in the USA. Lancet Psychiatry. 2021;8(2):130-40.

62. Thiruvalluru RK, Sharma MM, Olfson M, Keyes KM, Weissman MM, Pathak J, Xiao Y. Trends in Healthcare Service Disruptions and Associations with COVID-19 Outcomes among Patients with SMI vs. Non-SMI during COVID-19. medRxiv : the preprint server for health sciences. 2023.

63. Van Veenendaal VdMI, Onrust M, Paans W, Dieperink W, Van der Voort P. Long-Term Outcomes in COVID-19 ICU Patients: A Prospective Cohort Study. Research square. 2021.

64. Vlake JH, Van Bommel J, Hellemons ME, Wils E-J, Bienvenu OJ, Schut AFC, et al. Psychologic Distress and Quality of Life After ICU Treatment for Coronavirus Disease 2019: A Multicenter, Observational Cohort Study. Critical care explorations. 2021;3(8):e0497.

65. Vlake JH, van Bommel J, Wils E-J, Bienvenu J, Hellemons ME, Korevaar TI, et al. Intensive Care Unit-Specific Virtual Reality for Critically Ill Patients With COVID-19: Multicenter Randomized Controlled Trial. Journal of medical Internet research. 2022;24(1):e32368.

66. Walker S, Goodfellow H, Pookarnjanamorakot P, Murray E, Bindman J, Blandford A, et al. Impact of fatigue as the primary determinant of functional limitations among patients with post-COVID-19 syndrome: a cross-sectional observational study. BMJ open. 2023;13(6):e069217.

67. Whittaker HR, Gulea C, Koteci A, Kallis C, Morgan AD, Iwundu C, et al. GP consultation rates for sequelae after acute covid-19 in patients managed in the community or hospital in the UK: population-based study. BMJ (Clinical research ed). 2021;375:e065834.

68. Xie Y, Xu E, Al-Aly Z. Risks of mental health outcomes in people with covid-19: cohort study. Bmj. 2022;376:e068993.

69. Yang X, Yang X, Kumar P, Cao B, Ma X, Li T. Social support and clinical improvement in COVID-19 positive patients in China. Nursing outlook. 2020;68(6):830-7.

70. Zhang S, Liu Q, Yang F, Zhang J, Fu Y, Zhu Z, et al. Associations between COVID-19 infection experiences and mental health problems among Chinese adults: A large cross-section study. Journal of Affective Disorders. 2023;340:719-27.

**Wrong population for labour force outcome (n=70)**

1. Admon AJ, Iwashyna TJ, Kamphuis LA, Gundel SJ, Sahetya SK, Peltan ID, et al. Assessment of Symptom, Disability, and Financial Trajectories in Patients Hospitalized for COVID-19 at 6 Months. JAMA network open. 2023;6(2):e2255795.

2. Al Dweik R, Rahman MA, Ahamed FM, Ramada H, Al Sheble Y, ElTaher S, et al. COVID-19: Psychological distress, fear, and coping strategies among community members across the United Arab Emirates. PloS one. 2023;18(3):e0282479.

3. Ariza M, Cano N, Segura B, Bejar J, Barrue C, Cortes CU, et al. Cognitive and emotional predictors of quality of life and functioning after COVID-19. Annals of clinical and translational neurology. 2023.

4. Benoit-Piau J, Tremblay K, Piche A, Dallaire F, Belanger M, d'Entremont M-A, et al. Long-Term Consequences of COVID-19 in Predominantly Immunonaive Patients: A Canadian Prospective Population-Based Study. Journal of clinical medicine. 2023;12(18).

5. Bonner C, Ghouralal S-L. Long COVID and Chronic Conditions in the U.S. Workforce: Prevalence, Productivity Loss, and Disability. Journal of occupational and environmental medicine. 2024.

6. Cheng D, Calderwood C, Skyllberg E, Ainley A. Clinical characteristics and outcomes of adult patients admitted with COVID-19 in East London: a retrospective cohort analysis. BMJ open respiratory research. 2021;8(1).

7. Chommeloux J, Valentin S, Winiszewski H, Adda M, Pineton de Chambrun M, Moyon Q, et al. One-Year Mental and Physical Health Assessment in Survivors after Extracorporeal Membrane Oxygenation for COVID-19-related Acute Respiratory Distress Syndrome. American journal of respiratory and critical care medicine. 2023;207(2):150-9.

8. Damant RW, Rourke L, Cui Y, Lam GY, Smith MP, Fuhr DP, et al. Reliability and validity of the post COVID-19 condition stigma questionnaire: A prospective cohort study. EClinicalMedicine. 2023;55:101755.

9. Datta BK, Coughlin SS, Fazlul I, Pandey A. COVID-19 and health care-related financial toxicity in the United States: Evidence from the 2022 National Health Interview Survey. American Journal of Infection Control. 2023.

10. de Azevedo HMJ, Dos Santos NWF, Lafetá ML, de Albuquerque ALP, Tanni SE, Sperandio PA, Ferreira EVM. Persistence of symptoms and return to work after hospitalization for COVID-19. Jornal Brasileiro de Pneumologia. 2022;48(6).

11. Demoule A, Morawiec E, Decavele M, Ohayon R, Malrin R, Galarza-Jimenez MA, et al. Health-related quality of life of COVID-19 two and 12 months after intensive care unit admission. Annals of Intensive Care. 2022;12(1):16.

12. Ekstrand E, Brogardh C, Axen I, Fange AM, Stigmar K, Hansson EE. Perceived Consequences of Post-COVID-19 and Factors Associated with Low Life Satisfaction. International journal of environmental research and public health. 2022;19(22).

13. Evans RA, McAuley H, Harrison EM, Shikotra A, Singapuri A, Sereno M, et al. Physical, cognitive, and mental health impacts of COVID-19 after hospitalisation (PHOSP-COVID): a UK multicentre, prospective cohort study. The Lancet Respiratory medicine. 2021;9(11):1275-87.

14. Fitzgerald KC, Mecoli CA, Douglas M, Harris S, Aravidis B, Albayda J, et al. Risk factors for infection and health impacts of the COVID-19 pandemic in people with autoimmune diseases. medRxiv : the preprint server for health sciences. 2021.

15. Frontera JA, Yang D, Lewis A, Patel P, Medicherla C, Arena V, et al. A prospective study of long-term outcomes among hospitalized COVID-19 patients with and without neurological complications. Journal of the neurological sciences. 2021;426:117486.

16. Galas FRBG, Fernandes HM, Franci A, Rosario AL, Saretta R, Patore L, Jr., et al. In-hospital and Post-discharge Status in COVID-19 Patients with Acute Respiratory Failure Supported With Extracorporeal Membrane Oxygenation. ASAIO journal (American Society for Artificial Internal Organs : 1992). 2023;69(5):e181-e7.

17. Harari S, Mannucci PM, Nobili A, Galbussera AA, Fortino I, Leoni O, et al. Post-recovery impact of the second and third SARS-CoV-2 infection waves on healthcare resource utilization in Lombardy, Italy. Internal and Emergency Medicine. 2023;18(7):2011-8.

18. Ida FS, Ferreira HP, Vasconcelos AKM, Furtado IAB, Fontenele CJPM, Pereira AC. Post-COVID-19 syndrome: persistent symptoms, functional impact, quality of life, return to work, and indirect costs - a prospective case study 12 months after COVID-19 infection. Cadernos de Saude Publica. 2024;40(2).

19. Iwashyna TJ, Kamphuis LA, Gundel SJ, Hope AA, Jolley S, Admon AJ, et al. Continuing Cardiopulmonary Symptoms, Disability, and Financial Toxicity 1 Month After Hospitalization for

Third-Wave COVID-19: Early Results From a US Nationwide Cohort. Journal of Hospital Medicine. 2021;16(9):531-7.

20. Izadi N, Najafi A, Sadeghniiat-Haghighi K, Mohammadi H. Characterization of Long COVID and Its Contributing Factors among a Population of Health Care Workers in a 6-Month Follow-up. Medical journal of the Islamic Republic of Iran. 2023;37:29.

21. Jaywant A, Gunning FM, Oberlin LE, Santillana M, Ognyanova K, Druckman JN, et al. Cognitive Symptoms of Post-COVID-19 Condition and Daily Functioning. JAMA Network Open. 2024;7(2):E2356098.

22. Jacobsen PA, Andersen MP, Gislason G, Phelps M, Butt JH, Kober L, et al. Return to work after COVID-19 infection - A Danish nationwide registry study. Public Health. 2022;203:116-22.

23. Jamoulle M, Kazeneza-Mugisha G, Zayane A. Follow-Up of a Cohort of Patients with Post-Acute COVID-19 Syndrome in a Belgian Family Practice. Viruses. 2022;14(9).

24. Kisiel MA, Lee S, Malmquist S, Rykatkin O, Holgert S, Janols H, et al. Clustering Analysis Identified Three Long COVID Phenotypes and Their Association with General Health Status and Working Ability. Journal of clinical medicine. 2023;12(11).

25. Kisiel MA, Nordqvist T, Westman G, Svartengren M, Malinovschi A, Janols H. Patterns and predictors of sick leave among Swedish non-hospitalized healthcare and residential care workers with Covid-19 during the early phase of the pandemic. PloS one. 2021;16(12):e0260652.

26. Knight DRT, Munipalli B, Logvinov II, Halkar MG, Mitri G, Dabrh AMA, Hines SL. Perception, Prevalence, and Prediction of Severe Infection and Post-acute Sequelae of COVID-19. The American journal of the medical sciences. 2022;363(4):295-304.

27. Kumar M, Ali K, Sharma N, Sharma A, Jain M, Vats S, et al. Evaluation of Mental Health and Quality of Life among Indian Professionals Embarked as COVID-19 Survivors. Journal of lifestyle medicine. 2023;13(1):66-72.

28. Ladlow P, Holdsworth DA, O'Sullivan O, Barker-Davies RM, Houston A, Chamley R, et al. Exercise tolerance, fatigue, mental health, and employment status at 5 and 12 months following COVID-19 illness in a physically trained population. Journal of applied physiology (Bethesda, Md : 1985). 2023;134(3):622-37.

29. Lemhofer C, Sturm C, Loudovici-Krug D, Best N, Gutenbrunner C. The impact of Post-COVID-Syndrome on functioning - results from a community survey in patients after mild and moderate SARS-CoV-2-infections in Germany. Journal of occupational medicine and toxicology (London, England). 2021;16(1):45.

30. Lemhöfer C, Best N, Gutenbrunner C, Loudovici-Krug D, Teixido L, Sturm C. Perceived and Real Work Capacity of Patients with Post-COVID Symptoms after Mild Acute Course: A Analysis of the Rehabilitation Needs Questionnaire (RehabNeQ). Physikalische Medizin Rehabilitationsmedizin Kurortmedizin. 2022;54(3):151-8.

31. León-Herrera S, Samper-Pardo M, Oliván-Blázquez B, Sánchez-Recio R, Magallón-Botaya R, Sánchez-Arizcuren R. Loss of socioemotional and occupational roles in individuals with Long COVID according to sociodemographic and clinical factors: Secondary data from a randomized clinical trial. PLoS ONE. 2024;19(2 February).

32. Machado FVC, Meys R, Delbressine JM, Vaes AW, Goertz YMJ, van Herck M, et al. Construct validity of the Post-COVID-19 Functional Status Scale in adult subjects with COVID-19. Health and Quality of Life Outcomes. 2021;19(1):40.

33. Magnavita N, Arnesano G, Di Prinzio RR, Gasbarri M, Meraglia I, Merella M, Vacca ME. Post-COVID Symptoms in Occupational Cohorts: Effects on Health and Work Ability. International journal of environmental research and public health. 2023;20(9).

34. Mendola M, Leoni M, Cozzi Y, Manzari A, Tonelli F, Metruccio F, et al. Long-term COVID symptoms, work ability and fitness to work in healthcare workers hospitalized for Sars-CoV-2 infection. La Medicina del lavoro. 2022;113(5):e2022040.

35. Mirin AA. A preliminary estimate of the economic impact of long COVID in the United States. Fatigue: Biomedicine, Health and Behavior. 2022;10(4):190-9.

36. Miskowiak KW, Johnsen S, Sattler SM, Nielsen S, Kunalan K, Rungby J, et al. Cognitive impairments four months after COVID-19 hospital discharge: Pattern, severity and association with illness variables. European neuropsychopharmacology : the journal of the European College of Neuropsychopharmacology. 2021;46:39-48.

37. Miskowiak KW, Pedersen JK, Gunnarsson DV, Roikjer TK, Podlekareva D, Hansen H, et al. Cognitive impairments among patients in a long-COVID clinic: Prevalence, pattern and relation to illness severity, work function and quality of life. Journal of Affective Disorders. 2023;324:162-9.

38. Monti G, Leggieri C, Fominskiy E, Scandroglio AM, Colombo S, Tozzi M, et al. Two-months quality of life of COVID-19 invasively ventilated survivors; an Italian single-center study. Acta anaesthesiologica Scandinavica. 2021;65(7):912-20.

39. Moy FM, Hairi NN, Lim ERJ, Bulgiba A. Long COVID and its associated factors among COVID survivors in the community from a middle-income country-An online crosssectional study. PLoS ONE. 2022;17(8 August):e0273364.

40. Muller K, Poppele I, Ottiger M, Zwingmann K, Berger I, Thomas A, et al. Impact of Rehabilitation on Physical and Neuropsychological Health of Patients Who Acquired COVID-19 in the Workplace. International journal of environmental research and public health. 2023;20(2).

41. Nagata T, Nagata M, Hino A, Tateishi S, Ogami A, Tsuji M, et al. Prospective cohort study of workers diagnosed with COVID-19 and subsequent unemployment. Journal of Occupational Health. 2022;64(1).

42. Nanwani-Nanwani K, Lopez-Perez L, Gimenez-Esparza C, Ruiz-Barranco I, Carrillo E, Arellano MS, et al. Prevalence of post-intensive care syndrome in mechanically ventilated patients with COVID-19. Scientific reports. 2022;12(1):7977.

43. Nehme M, Braillard O, Chappuis F, Courvoisier DS, Kaiser L, Soccal PM, et al. One-year persistent symptoms and functional impairment in SARS-CoV-2 positive and negative individuals. Journal of Internal Medicine. 2022;292(1):103-15.

44. Nowak-Kiczmer M, Kubicka-Baczyk K, Niedziela N, Adamczyk B, Wierzbicki K, Bartman W, Adamczyk-Sowa M. The course of COVID-19 infection in patients with multiple sclerosis-The experience of one center based on the population of Upper Silesia. Multiple sclerosis and related disorders. 2021;52:102984.

45. O' Mahony L, Buwalda T, Blair M, Forde B, Lunjani N, Ambikan A, et al. Impact of Long COVID on health and quality of life. HRB open research. 2022;5:31.

46. O'Regan E, Svalgaard IB, Sorensen AIV, Spiliopoulos L, Bager P, Nielsen NM, et al. A hybrid register and questionnaire study of Covid-19 and post-acute sick leave in Denmark. Nature Communications. 2023;14(1):6266.

47. Okawara M, Hirashima K, Igarashi Y, Mafune K, Muramatsu K, Nagata T, et al. Impact of COVID-19 Infection on Work Functioning in Japanese Workers: A Prospective Cohort Study. Safety and Health at Work. 2023;14(4):445-50.

48. Oravec MJ. Incidence of post-acute COVID-19 sequelae and predictors for post-COVID infection health care utilization in an integrated health system patient population. Dissertation Abstracts International: Section B: The Sciences and Engineering. 2024;85(2-B):No-Specified.

49. Perlis RH, Trujillo KL, Safarpour A, Santillana M, Ognyanova K, Druckman J, Lazer D. Research Letter: Association between long COVID symptoms and employment status. medRxiv. 2022.

50. Peter RS, Nieters A, Brockmann SO, Gopel S, Kindle G, Merle U, et al. Association of BMI with general health, working capacity recovered, and post-acute sequelae of COVID-19. Obesity (Silver Spring, Md). 2023;31(1):43-8.

51. Potter R, Eisenberg E, Mock V, Bose S. Impact of CIVUD-19 pandemic on identified psychosocial needs of specialty social worker embedded in pulmonary practice. Chest. 2023;164(4 Supplement):A3827.

52. Rhead RD, Wels J, Moltrecht B, Shaw RJ, Silverwood RJ, Zhu J, et al. Long COVID and financial outcomes: Evidence from four longitudinal population surveys. medRxiv. 2023.

53. Robinson-Lane SG, Sutton NR, Chubb H, Yeow RY, Mazzara N, DeMarco K, et al. Race, Ethnicity, and 60-Day Outcomes After Hospitalization With COVID-19. Journal of the American Medical Directors Association. 2021;22(11):2245-50.

54. Salmon D, Slama D, Linard F, Dumesges N, Lebaut V, Hakim F, et al. Factors associated with release relief of Long COVID symptoms at 12-Months and their impact on daily life. medRxiv. 2022.

55. Salmon D, Slama D, Linard F, Dumesges N, Lebaut V, Hakim F, et al. Long COVID patients continue to experience significant symptoms at 12 months and factors associated with improvement: a prospective cohort study in France (PERSICOR). International journal of infectious diseases : IJID : official publication of the International Society for Infectious Diseases. 2023.

56. Sansone D, Tassinari A, Valentinotti R, Kontogiannis D, Ronchese F, Centonze S, et al. Persistence of Symptoms 15 Months since COVID-19 Diagnosis: Prevalence, Risk Factors and Residual Work Ability. Life (Basel, Switzerland). 2022;13(1).

57. Schilling C, Nieters A, Schredl M, Peter RS, Rothenbacher D, Brockmann SO, et al. Pre-existing sleep problems as a predictor of post-acute sequelae of COVID-19. Journal of sleep research. 2023:e13949.

58. Singh P, Mohanti BK, Mohapatra SK, Deep A, Harsha B, Pathak M, Patro S. Post-COVID-19 Assessment of Physical, Psychological, and Socio-Economic Impact on a General Population of Patients From Odisha, India. Cureus. 2022;14(10):e30636.

59. Smith JL, Deighton K, Innes AQ, Holl M, Mould L, Liao Z, et al. Improved clinical outcomes in response to a 12-week blended digital and community-based long-COVID-19 rehabilitation programme. Frontiers in medicine. 2023;10:1149922.

60. Sorensen AIV, Spiliopoulos L, Bager P, Nielsen NM, Hansen JV, Koch A, et al. Post-acute symptoms, new onset diagnoses and health problems 6 to 12 months after SARS-CoV-2 infection: a nationwide questionnaire study in the adult Danish population. medRxiv. 2022.

61. Strasburger C, Hieber D, Karthan M, Juster M, Schobel J. Return to work after Post-COVID: describing affected employees' perceptions of personal resources, organizational offerings and care pathways. Frontiers in public health. 2023;11:1282507.
62. Tang S, Horter L, Bosh K, Kassem AM, Kahn EB, Ricaldi JN, et al. Change in unemployment by social vulnerability among United States counties with rapid increases in COVID-19 incidence-July 1-October 31, 2020. PLoS ONE. 2022;17(4 April):e0265888.
63. Vandersmissen G, Verbeeck J, Henckens P, Van Dyck J, Wuytens C, Molenberghs G, Godderis L. Sick leave due to SARS-CoV-2 infection. Occupational medicine (Oxford, England). 2023.
64. Visconti NRGDR, Cailleaux-Cezar M, Capone D, Dos Santos MIV, Graca NP, Loivos LPP, et al. Long-term respiratory outcomes after COVID-19: a Brazilian cohort study. Revista panamericana de salud publica = Pan American journal of public health. 2022;46:e187.
65. Wahlgren C, Divanoglou A, Larsson M, Nilsson E, Ostholm Balkhed A, Niward K, et al. Rehabilitation needs following COVID-19: Five-month post-discharge clinical follow-up of individuals with concerning self-reported symptoms. EClinicalMedicine. 2022;43:101219.
66. Wahlgren C, Forsberg G, Divanoglou A, Ostholm Balkhed A, Niward K, Berg S, Levi R. Two-year follow-up of patients with post-COVID-19 condition in Sweden: a prospective cohort study. The Lancet Regional Health - Europe. 2023;28:100595.
67. Wallin E, Hultstrom M, Lipcsey M, Frithiof R, Rubertsson S, Larsson IM. Intensive care-treated COVID-19 patients' perception of their illness and remaining symptoms. Acta Anaesthesiologica Scandinavica. 2022;66(2):240-7.
68. Wiertz CMH, Hemmen B, Sep SJS, Van Santen S, Van Horn YY, Van Kuijk SMJ, Verbunt JA. Life after COVID-19: the road from intensive care back to living - a prospective cohort study. BMJ Open. 2022;12(11):e062332.
69. Wolff Sagy Y, Feldhamer I, Brammli-Greenberg S, Lavie G. Estimating the economic burden of long-Covid: the additive cost of healthcare utilisation among COVID-19 recoverees in Israel. BMJ global health. 2023;8(7).
70. Ziauddeen N, Gurdasani D, O'Hara ME, Hastie C, Roderick P, Yao G, Alwan NA. Characteristics and impact of Long Covid: Findings from an online survey. PloS one. 2022;17(3):e0264331.

**Population not post-COVID (n=52)**

1. Afsharnejad B, Milbourn B, Brown C, Clifford R, Foley K-R, Logan A, et al. Understanding the utility of "Talk-to-Me" an online suicide prevention program for Australian university students. Suicide & life-threatening behavior. 2023;53(5):725-38.
2. Alyahya MA, Elshaer IA, Sobaih AEE. The Impact of Job Insecurity and Distributive Injustice Post COVID-19 on Social Loafing Behavior among Hotel Workers: Mediating Role of Turnover Intention. International journal of environmental research and public health. 2021;19(1).
3. Ambrosetti J, Macheret L, Folliet A, Wullschleger A, Amerio A, Aguglia A, et al. Psychiatric emergency admissions during and after COVID-19 lockdown: short-term impact and long-term implications on mental health. BMC psychiatry. 2021;21(1):465.
4. Antar AAR, Yu T, Demko ZO, Hu C, Tornheim JA, Blair PW, et al. Long COVID brain fog and muscle pain are associated with longer time to clearance of SARS-CoV-2 RNA from the upper respiratory tract during acute infection. medRxiv : the preprint server for health sciences. 2023.

5. Asiamah N, Muhonja F, Omisore A, Opuni FF, Mensah HK, Danquah E, et al. The association between core job components, physical activity, and mental health in African academics in a post-COVID-19 context. Current psychology (New Brunswick, NJ). 2023;42(9):7235-51.
6. Atalla E, Kalligeros M, Giampaolo G, Mylona EK, Shehadeh F, Mylonakis E. Readmissions among patients with COVID-19. Int J Clin Pract. 2021;75(3):e13700.
7. Aubouin-Bonnaventure J, Chevalier S, Lahiani F-J, Fouquereau E. Preventing workers' need for recovery and turnover intentions: The protective effect of virtuous organizational practices through work ability. Journal of Workplace Behavioral Health. 2023:No-Specified.
8. Baumgartner TJ, Sanborn K, Reta M, Lenards N, Hunzeker A, Zeiler S. Perceptions of burnout in medical dosimetry within a postpandemic work environment. Medical dosimetry : official journal of the American Association of Medical Dosimetrists. 2023;48(2):77-81.
9. Bou Sanayeh E, El Chamieh C, Saade MC, Maalouf RG, Bizri M. Post-traumatic stress symptoms experienced by healthcare workers in Lebanon four months following Beirut's ammonium nitrate explosion: a survey-based study. Archives of public health = Archives belges de sante publique. 2022;80(1):156.
10. Flament J, Scius N, Zdanowicz N, Regnier M, De Canniere L, Thonon H. Influence of post-COVID-19 deconfinement on psychiatric visits to the emergency department. The American journal of emergency medicine. 2021;48:238-42.
11. Goldsmith LP, Anderson K, Clarke G, Crowe C, Jarman H, Johnson S, et al. Service use preceding and following first referral for psychiatric emergency care at a short-stay crisis unit: A cohort study across three cities and one rural area in England. The International journal of social psychiatry. 2023;69(4):928-41.
12. Grandey AA, Sayre GM, French KA. "A blessing and a curse": Work loss during coronavirus lockdown on short-term health changes via threat and recovery. Journal of occupational health psychology. 2021;26(4):261-75.
13. Ham E, Hilton NZ, Crawford J, Kim S. Psychiatric inpatient services in Ontario, 2019-2021: a cross-sectional comparison of admissions, diagnoses and acuity during the COVID-19 prerestriction, restriction and postrestriction periods. CMAJ open. 2023;11(5):E988-E94.
14. Harvey-Dunstan TC, Jenkins AR, Gupta A, Hall IP, Bolton CE. Patient-related outcomes in patients referred to a respiratory clinic with persisting symptoms following non-hospitalised COVID-19. Chronic Respiratory Disease. 2022;19.
15. Hertz-Palmor N, Moore TM, Gothelf D, DiDomenico GE, Dekel I, Greenberg DM, et al. Association among income loss, financial strain and depressive symptoms during COVID-19: Evidence from two longitudinal studies. Journal of affective disorders. 2021;291:1-8.
16. Iasevoli F, Fornaro M, D'Urso G, Galletta D, Casella C, Paternoster M, et al. Psychological distress in patients with serious mental illness during the COVID-19 outbreak and one-month mass quarantine in Italy. Psychological Medicine. 2021;51(6):1054-6.
17. Iverson KM, Stolzmann KL, Brady JE, Adjognon OL, Dichter ME, Lew RA, et al. Integrating Intimate Partner Violence Screening Programs in Primary Care: Results from a Hybrid-II Implementation-Effectiveness RCT. American journal of preventive medicine. 2023;65(2):251-60.
18. Jacquet LH. Major mental health referral quality improvement project for castle high school staff. Dissertation Abstracts International: Section B: The Sciences and Engineering. 2023;84(1-B):No-Specified.

19. Jayawardana D, Gannon B. Use of telehealth mental health services during the COVID-19 pandemic. Australian health review : a publication of the Australian Hospital Association. 2021;45(4):442-6.

20. Kameyama K, Mizutani K, Miyake Y, Iwase T, Mizutani Y, Yamada M, et al. Evaluation of physical and psychological status of health care workers infected with COVID-19 during a hospital outbreak in Japan. Journal of infection and chemotherapy : official journal of the Japan Society of Chemotherapy. 2023;29(2):126-30.

21. Kandiah S, Chamberlain R. Does Implementation of Patient Health Questionnaire Increase Referrals to Behavioral Health Services. Clinical Journal of Sport Medicine. 2023;33(3):328.

22. Kim J, Rim SJ, Jo M, Lee MG, Park S. The Trend of Psychiatric Visits and Psychiatric Medication Prescription Among People Tested for SARS-CoV-2 During the Initial Phase of COVID-19 Pandemic in South Korea. Psychiatry Investig. 2022;19(1):61-71.

23. Kupferschmitt A, Langheim E, Tuter H, Etzrodt F, Loew TH, Kollner V. First results from post-COVID inpatient rehabilitation. Frontiers in rehabilitation sciences. 2022;3:1093871.

24. Lam MH-B, Wing Y-K, Yu MW-M, Leung C-M, Ma RCW, Kong APS, et al. Mental morbidities and chronic fatigue in severe acute respiratory syndrome survivors: long-term follow-up. Archives of internal medicine. 2009;169(22):2142-7.

25. Lee ATC, Mo FYM, Lam LCW. Higher psychogeriatric admissions in COVID-19 than in severe acute respiratory syndrome. International journal of geriatric psychiatry. 2020;35(12):1449-57.

26. Leeds LW, Powers J. Implementation of a bystander intervention model to promote referrals to and enhance utilization of mental health resources by students in a community college setting. Dissertation Abstracts International Section A: Humanities and Social Sciences. 2023;84(3-A):No-Specified.

27. Lerminiaux D, Somers L. [An analysis of child psychiatric emergency department consultations from 2019 to 2022 in the context of COVID-19]. Face a la COVID-19 Une analyse des consultations aux urgences pedopsychiatriques de 2019 a 2022 dans le contexte de la COVID-19. 2023;78(11):614-8.

28. Liu J, Tong Y, Li S, Tian Z, He L, Zheng J. Compliance with COVID-19-preventive behaviours among employees returning to work in the post-epidemic period. BMC public health. 2022;22(1):369.

29. Lynch DA, Stefancic A, Cabassa LJ, Medalia A. Client, clinician, and administrator factors associated with the successful acceptance of a telehealth comprehensive recovery service: A mixed methods study. Psychiatry research. 2021;300:113871.

30. Mantero V, Basilico P, Balgera R, Rigamonti A, Sozzi M, Salmaggi A, Cordano C. Flu-like syndrome due to interferon-beta injections does not increase anxiety, depression, and lost working days in multiple sclerosis patients during the Sars-CoV-2 pandemic. Clinical Neurology and Neurosurgery. 2023;232:107892.

31. McNicholas F, Kelleher I, Hedderman E, Lynch F, Healy E, Thornton T, et al. Referral patterns for specialist child and adolescent mental health services in the Republic of Ireland during the COVID-19 pandemic compared with 2019 and 2018. BJPsych open. 2021;7(3):e91.

32. Meirun T, Bano S, Javaid MU, Arshad MZ, Shah MU, Rehman U, et al. Nuances of COVID-19 and Psychosocial Work Environment on Nurses' Wellbeing: The Mediating Role of Stress and Eustress in Lieu to JD-R Theory. Frontiers in psychology. 2020;11:570236.

33. Modarres MH, Kalafatis C, Apostolou P, Tabet N, Khaligh-Razavi S-M. The use of the integrated cognitive assessment to improve the efficiency of primary care referrals to memory services in the

accelerating dementia pathway technologies study. Frontiers in aging neuroscience. 2023;15:1243316.

34. Monier D, Bonjean P, Carcasset P, Moulin M, Pozzetto B, Botelho-Nevers E, et al. Factors Contributing to Delayed Return to Work among French Healthcare Professionals Afflicted by COVID-19 at a Hospital in the Rhone-Alpes Region, 2021. International journal of environmental research and public health. 2023;20(21).

35. Muthumuni N, Sommer JL, El-Gabalawy R, Reynolds KA, Mota NP. Evaluating the mental health status, help-seeking behaviors, and coping strategies of Canadian essential workers versus non-essential workers during COVID-19: a longitudinal study. Anxiety, stress, and coping. 2023:1-14.

36. Purrington J, Beail N. The impact of Covid-19 on access to psychological services. Advances in Mental Health and Intellectual Disabilities. 2021;15(4):119-31.

37. Puthucheary Z, Brown C, Corner E, Wallace S, Highfield J, Bear D, et al. The Post-ICU presentation screen (PICUPS) and rehabilitation prescription (RP) for intensive care survivors part II: Clinical engagement and future directions for the national Post-Intensive care Rehabilitation Collaborative. Journal of the Intensive Care Society. 2022;23(3):264-72.

38. Qiu X, Lan Y, Miao J, Wang H, Wang H, Wu J, et al. A Comparative Study on the Psychological Health of Frontline Health Workers in Wuhan Under and After the Lockdown. Frontiers in psychiatry. 2021;12:701032.

39. Sawant N, Ingawale S, Lokhande U, Patil S, Ayub EFM, Rathi V. Psychiatric Sequelae and COVID Experiences of Post COVID -19 Recovered Resident Doctors and Interns of a Tertiary General Hospital in Mumbai. The Journal of the Association of Physicians of India. 2021;69(4):22-6.

40. Sonney J, Peck JL. The Cost of Caring During COVID-19: A Clarion Call to Action to Support the Pediatric Advanced Practice Nursing Workforce. Journal of pediatric health care : official publication of National Association of Pediatric Nurse Associates & Practitioners. 2023;37(6):658-72.

41. Takeuchi E, Katanoda K, Cheli S, Goldzweig G, Tabuchi T. Restrictions on healthcare utilization and psychological distress among patients with diseases potentially vulnerable to COVID-19; the JACSIS 2020 study. Health psychology and behavioral medicine. 2022;10(1):229-40.

42. Taylor JL, Adams RE, Pezzimenti F, Zheng S, Bishop SL. Job loss predicts worsening depressive symptoms for young adults with autism: A COVID-19 natural experiment. Autism Research. 2022;15(1):93-102.

43. Turan Ş, Poyraz B, Aksoy Poyraz C, Demirel Ö F, Tanrıöver Aydın E, Uçar Bostan B, et al. Characteristics and outcomes of COVID-19 inpatients who underwent psychiatric consultations. Asian J Psychiatr. 2021;57:102563.

44. Wang CC, Lo J, Saunders R, Adama E, Bulsara C, Etherton-Beer C, Yang AWH. Light acupuncture and five-element music therapy for nurses' mental health and well-being during and post-COVID-19: protocol for a randomised cross-over feasibility study. BMJ open. 2022;12(4):e057106.

45. Wang PR, Anand A, Bena J, Morrison S, Weleff J. Changes in emergency department utilization in vulnerable populations after COVID-19 shelter in place orders. medRxiv. 2023.

46. Werling AM, Walitza S, Drechsler R. Impact of the COVID-19 lockdown on screen media use in patients referred for ADHD to child and adolescent psychiatry: an introduction to problematic use of the internet in ADHD and results of a survey. Journal of neural transmission (Vienna, Austria : 1996). 2021;128(7):1033-43.

47. Werling AM, Walitza S, Grunblatt E, Drechsler R. Media use before, during and after COVID-19 lockdown according to parents in a clinically referred sample in child and adolescent psychiatry: Results of an online survey in Switzerland. Comprehensive psychiatry. 2021;109:152260.
48. Yang Q, Huo J, Li J, Jiang Y. Research on the influence of the COVID-19 epidemic on work stress of returning workers in China: A study based on empirical analyses of industrial enterprises. Work (Reading, Mass). 2020;67(1):67-79.
49. Yu X, Langa KM, Cho T-C, Kobayashi LC. Association of Perceived Job Insecurity With Subsequent Memory Function and Decline Among Adults 55 Years or Older in England and the US, 2006 to 2016. JAMA network open. 2022;5(4):e227060.
50. Zahedifar F, Nejatifar Z, Rafiei S, Hashemi F. The effect of educational intervention on anxiety and quality of life among individuals referring to healthcare centers in the face of covid-19. Acta Medica Iranica. 2021;59(8):484-90.
51. Zhu Z, Xu S, Wang H, Liu Z, Wu J, Li G, et al. COVID-19 in Wuhan: Sociodemographic characteristics and hospital support measures associated with the immediate psychological impact on healthcare workers. EClinicalMedicine. 2020;24:100443.
52. Zielasek J, Lehmann I, Vrinssen J, Gouzoulis-Mayfrank E. Analysis of the utilization, processes, and outcomes of inpatient mental healthcare during the first three waves of the COVID-19 pandemic in the federal state of North Rhine-Westphalia, Germany. Frontiers in psychiatry. 2022;13:957951.

**Language (n=15)**

1. August D, Stete K, Hilger H, Gotz V, Biever P, Hosp J, et al. [Complaints and clinical findings six months after COVID-19: outpatient follow-up at the University Medical Center Freiburg]. Persistierende Beschwerden 6 Monate nach COVID-19 - Erfahrungen aus der COVID-19-Nachsorgeambulanz des Universitatsklinikums Freiburg. 2021;146(17):e65-e73.
2. Bogolepova AN, Osinovskaya NA, Kovalenko EA, Makhnovich EV. Fatigue and cognitive impairment in post-COVID syndrome: possible treatment approaches. Nevrologiya, Neiropsikhiatriya, Psikhosomatika. 2021;13(4):88-93.
3. Delevaux I, Duquenne C, Kokkinakis I, Favrat B. [Neuropsychiatric manifestations of post COVID-19 Syndrome and Disability Insurance (DI)]. Covid long neuropsychiatrique et assurance invalidite (AI). 2023;19(827):992-3.
4. Dresing H, Meyer-Lindenberg A. [Future issues in "long COVID" psychiatric assessment]. Kunftige Aufgaben der psychiatrischen Begutachtung bei Long-COVID". 2022;93(3):309-12.
5. Gulyaev SA. Dynamic electroencephalographic study of persons - mild COVID-19 convalescents. Russkii Zhunal Detskoi Nevrologii. 2022;17(4):44-53.
6. Haller J, Kocalevent R-D, Nienhaus A, Peters C, Bergelt C, Koch-Gromus U. [Persistent fatigue symptoms following COVID-19 infection in healthcare workers: risk factors and impact on quality of life]. Anhaltende Fatigue als Folge einer COVID-19-Infektion bei Beschaftigten im Gesundheitswesen: Risikofaktoren und Auswirkungen auf die Lebensqualitat. 2022;65(4):471-80.
7. Meskina ER, Khadisova MK, Stashko TV, Galkina LA, Tselipanova EE, Shilkina IM. Efficiency of application of sorbed probiotics in the complex therapy of pneumonia caused by SARS-CoV-2. Quality of life in the short term COVID-19. Infectious Diseases: News, Opinions, Training. 2022;11(3):69-80.

8. Petrova NN, Pryanikova EV, Pustotin Yu L, Yakusheva NV, Dorofeikova MV. Post-COVID syndrome in psychiatric practice. Nevrologiya, Neiropsikhiatriya, Psikhosomatika. 2022;14(6):49-54.
9. Rover MM, Trott G, Scolari FL, Silva MMDd, Souza Dd, Santos RdRMD, et al. Health-Related Quality of Life and Long-Term Outcomes after Mildly Symptomatic COVID-19: The Post-COVID Brazil Study 2 Protocol. Qualidade de Vida Relacionada a Saude e Desfechos em Longo Prazo apos COVID-19 Sintomatica Leve: Protocolo do Estudo Pos-COVID Brasil 2. 2023;120(9):e20220835.
10. Rusinova DS, Vasil'eva TM, Bezymyanny AS, Starshinin AV. Analysis of Employee's Working Capacity in Children's City Outpatient's Clinic 133 of Moscow City Health Department after COVID-19. Pediatricheskaya Farmakologiya. 2021;18(6):507-14.
11. Rutsch M, Deck R. [Occupational Stress of Long Covid Rehabilitants and Return to Work After Pneumological Rehabilitation]. Berufliche Belastungen von Long-Covid-Rehabilitand*innen und Ruckkehr zur Arbeit nach einer pneumologischen Rehabilitation. 2023;62(6):369-78.
12. Rutsch M, Frommhold J, Buhr-Schinner H, Gross T, Schuller PO, Deck R. [Pneumological Rehabilitation in Patients with Long Covid - Health Changes at the End of the Inpatient Rehabilitation Measure]. Pneumologische Rehabilitation bei Long Covid - Gesundheitliche Veranderungen am Ende der stationaren Rehabilitationsmasnahme. 2023;62(6):359-68.
13. Tamasi J, Kalabay L. Monitoring the development of post-COVID-19 syndrome. Poszt-COVID-19-szindromas esetek kialakulasanak kovetese. 2022;163(9):335-42.
14. Trott G, Scolari FL, Rover MM, Silva MMDd, Souza Dd, Santos RdRMD, et al. Long-term Health-Related Quality of Life and Outcomes after Hospitalization for COVID-19 in Brazil: Post-COVID Brazil 1 Study Protocol. Qualidade de Vida em Longo Prazo e Desfechos apos Internacao por COVID-19 no Brasil: Protocolo do Estudo Pos-COVID Brasil 1. 2023;120(11):e20230378.
15. Vysokova VO, Tyuvina NA, Maksimova TN, Prokhorova SV. Mental disorders during the pandemic of a new coronavirus infection: clinical features. Nevrologiya, Neiropsikhiatriya, Psikhosomatika. 2023;15(3):60-7.

**Author contact, unsuccessful (n=8)**

1. Adjorlolo S, Adjorlolo P, Andoh-Arthur J, Ahiable EK, Kretchy IA, Osafo J. Post-Traumatic Growth and Resilience among Hospitalized COVID-19 Survivors: A Gendered Analysis. International journal of environmental research and public health. 2022;19(16).
2. Bozzani A, Arici V, Tavazzi G, Ragni F, Mojoli F, Cavallini E, et al. Trends (2020-2022) toward Reduced Prevalence of Postcoronavirus Disease Syndrome and Improved Quality of Life for Hospitalized Coronavirus Disease 2019 Patients with Severe Infection and Venous Thromboembolism. Seminars in thrombosis and hemostasis. 2023.
3. Liu EN, Yang JH, Patel L, Arora J, Gooding A, Ellis R, Graves JS. Longitudinal analysis and treatment of neuropsychiatric symptoms in post-acute sequelae of COVID-19. Journal of neurology. 2023;270(10):4661-72.
4. McLaughlin M, Cerexhe L, Macdonald E, Ingram J, Sanal-Hayes NEM, Hayes LD, et al. A Cross-Sectional Study of Symptom Prevalence, Frequency, Severity, and Impact of Long COVID in Scotland: Part II. The American journal of medicine. 2023.
5. Nehme M, Vieux L, Kaiser L, Chappuis F, Chenaud C, Guessous I. The longitudinal study of subjective wellbeing and absenteeism of healthcare workers considering post-COVID condition and the COVID-19 pandemic toll. Scientific reports. 2023;13(1):10759.

6. Price JK, de Avila L, Stepanova M, Weinstein AA, Pham H, Keo Wo, et al. Severe, Persistent, Disruptive Fatigue Post-SARS-CoV-2 Disproportionately Affects Young Women. International journal of general medicine. 2023;16:4393-404.
7. Romer V, Sivapalan P, Eklof J, Nielsen SD, Harboe ZB, Biering-Sorensen T, et al. SARS-CoV-2 and risk of psychiatric hospital admission and use of psychopharmaceuticals: A nationwide registry study of 4,585,083 adult Danish citizens. European psychiatry : the journal of the Association of European Psychiatrists. 2023;66(1):e50.
8. Simkovich SM, Ahmed N, Chou J, McCullers A, Wisotzky EM, Semel J, et al. Health, social, and economic characteristics of patients enrolled in a COVID-19 recovery program. PLoS ONE. 2022;17(11 November):e0278154.

**No usable data (n=7)**

1. Bonham C, Juarez R, Siegal N. Long COVID and Unemployment in Hawaii. Int J Environ Res Public Health. 2023;20(13).
2. Galanis P, Katsiroumpa A, Vraka I, Kosiara K, Siskou O, Konstantakopoulou O, et al. Post-COVID-19 syndrome and related dysautonomia affect patients' life and work productivity. medRxiv. 2023.
3. Kennedy J, Parker M, Seaborne M, Mhereeg M, Walker A, Walker V, et al. Health care use attributable to COVID-19: A propensity matched national electronic health records cohort study of 249,390 people in Wales, UK. medRxiv. 2022.
4. Miskowiak KW, Fugledalen L, Jespersen AE, Sattler SM, Podlekareva D, Rungby J, et al. Trajectory of cognitive impairments over 1 year after COVID-19 hospitalisation: Pattern, severity, and functional implications. European neuropsychopharmacology : the journal of the European College of Neuropsychopharmacology. 2022;59:82-92.
5. Perlis RH, Lunz Trujillo K, Safarpour A, Santillana M, Ognyanova K, Druckman J, Lazer D. Association of Post-COVID-19 Condition Symptoms and Employment Status. JAMA network open. 2023;6(2):e2256152.
6. Rafferty E, Unsal A, Kirwin E, Kingdom U. Healthcare costs and effects of post-COVID-19 condition in Canada. Can Commun Dis Rep. 2023;49(10):425-32.
7. Yelin D, Margalit I, Nehme M, Bordas-Martinez J, Pistelli F, Yahav D, et al. Patterns of Long COVID Symptoms: A Multi-Center Cross Sectional Study. Journal of Clinical Medicine. 2022;11(4):898.

**N <30 with MH for labour force (n=7)**

1. Dimitrova M, Marinova Y, Dilkov D. Investigation of Cognitive Impairment in the Course of Post-COVID Syndrome. Diagnostics. 2023;13(16).
2. Ghosn J, Bachelet D, Livrozet M, Cervantes-Gonzalez M, Poissy J, Goehringer F, et al. Prevalence of post-acute coronavirus disease 2019 symptoms twelve months after hospitalization in participants retained in follow-up: analyses stratified by gender from a large prospective cohort. Clinical microbiology and infection : the official publication of the European Society of Clinical Microbiology and Infectious Diseases. 2023;29(2):254.e7-.e13.

3. Gilmartin M, Collins J, Mason S, Horgan A, Cuadrado E, Ryberg M, et al. Post-Intensive Care COVID Survivorship Clinic: A Single-Center Experience. Critical care explorations. 2022;4(5):e0700.
4. Larsson I-M, Hultstrom M, Lipcsey M, Frithiof R, Rubertsson S, Wallin E. Poor long-term recovery after critical COVID-19 during 12 months longitudinal follow-up. Intensive & critical care nursing. 2023;74:103311.
5. Park N, Oberlin L, Cherestal S, Bueno Castellano C, Dargis M, Wyka KE, et al. Trajectory and outcomes of psychiatric symptoms in first-wave COVID-19 survivors referred for telepsychotherapy. General Hospital Psychiatry. 2023;81:86-8.
6. Rajajee V, Fung CM-C, Seagly KS, Park PK, Raghavendran K, Machado-Aranda DA, et al. One-Year Functional, Cognitive, and Psychological Outcomes Following the Use of Extracorporeal Membrane Oxygenation in Coronavirus Disease 2019: A Prospective Study. Critical care explorations. 2021;3(9):e0537.
7. Scherlinger M, Felten R, Gallais F, Nazon C, Chatelus E, Pijnenburg L, et al. Refining "Long-COVID" by a Prospective Multimodal Evaluation of Patients with Long-Term Symptoms Attributed to SARS-CoV-2 Infection. Infectious diseases and therapy. 2021;10(3):1747-63.

**Duplicates (n=4)**

1. Benzakour L, Braillard O, Mazzola V, Gex D, Nehme M, Perone SA, et al. P.0805 Impact of peritraumatic dissociation in hospitalized patients with COVID-19 pneumonia: a longitudinal study. European Neuropsychopharmacology. 2021;53(Supplement 1):S589.
2. Bonazza F, Luridiana Battistini C, Fior G, Bergamelli E, Wiedenmann F, D'Agostino A, et al. Recovering from COVID-19: psychological sequelae and post-traumatic growth six months after discharge. European journal of psychotraumatology. 2022;13(1):2095133.
3. Ferrando SJ, Lynch S, Ferrando N, Dornbush R, Shahar S, Klepacz L. Anxiety and posttraumatic stress in post-acute sequelae of COVID-19: prevalence, characteristics, comorbidity, and clinical correlates. Frontiers in psychiatry. 2023;14:1160852.
4. Peter RS, Nieters A, Krausslich HG, Brockmann SO, Gopel S, Kindle G, et al. Prevalence, determinants, and impact on general health and working capacity of post-acute sequelae of COVID-19 six to 12 months after infection: a population-based retrospective cohort study from southern Germany. medRxiv. 2022.

**Other (n=2)**

1. Chen H, Ma Q, Du B, Huang Y, Zhu S-G, Li S-L, et al. Psychotherapy and Follow-Up in Health Care Workers After the COVID-19 Epidemic: A Single Center's Experience. Psychology research and behavior management. 2022;15:2245-58.
2. No authorship. Psychotherapy and follow-up in health care workers after the COVID-19 epidemic: A single center's experience [Retraction]. Psychology Research and Behavior Management. 2022;15:3409.

### Table S2: Risk of Bias Assessments

| Author, year | Were the two groups similar and recruited from the same population? | Were the exposures measured similarly to assign people to both exposed and unexposed groups? | Were the exposures measured in a valid and reliable way? | Were confounding factors identified? | Were strategies to deal with confounding factors stated? | Were the participants free of the outcome at the start of the study or exposure? | Were the outcomes measured in a valid and reliable way? | Was the follow up time reported and sufficient to be long enough for outcomes to occur? | Was follow up complete, and if not, were the reasons to loss to follow up described and explored? | Were strategies to address incomplete follow up utilized? | Was appropriate statistical analysis used (i.e. age, sex, severity)? | Overall assessment* |
|---|---|---|---|---|---|---|---|---|---|---|---|---|
| **Labour Force Outcomes** | | | | | | | | | | | | |
| **Anik, 2023** | + | + | + | ? | - | - | ? | + | + | NA | No | High |
| **Danesh, 2021** | + | + | ? | - | - | ? | ? | + | + | NA | No | High |
| **Diem, 2022** | + | + | + | ? | + | ? | + | + | + | NA | No | Low/ Moderate |
| **Garcia-Molina, 2022** | + | + | + | + | - | + | ? | + | + | NA | - | Low/ Moderate |
| **Green, 2023** | + | + | - | ? | - | ? | ? | + | + | NA | - | High |
| **Kerksieck, 1001** | + | + | + | + | - | + | + | - | + | NA | - | Low/ Moderate |
| **Lemhofer, 2023** | + | + | + | ? | - | + | ? | ? | + | NA | - | Low/ Moderate |
| **O'Sullivan, 2023** | + | + | + | + | ? | + | + | + | + | NA | - | Low/ Moderate |
| **Romero-Rodrique, 2023** | + | + | - | - | - | + | ? | ? | + | NA | - | High |
| **Thompson, 2023** | + | + | + | + | - | + | + | + | + | NA | - | Low/ Moderate |
| **Tsuchida, 2023** | + | + | ? | ? | - | ? | ? | ? | + | NA | - | High |
| **Vanichkachorn, 2021** | + | + | - | - | - | ? | + | + | + | NA | - | High |

| | | | | | | | | | | | | |
|---|---|---|---|---|---|---|---|---|---|---|---|---|
| **Braga, 2022** | NA | NA | + | + | + | - | + | ? | + | NA | - | Low/ Moderate |
| **Davis, 2021** | NA | NA | - | ? | - | ? | + | + | + | NA | - | High |
| **Delgado-Alonso, 2022** | NA | NA | + | ? | - | + | + | + | + | NA | - | Low/ Moderate |
| **Dressing, 2022** | NA | NA | + | + | ? | ? | + | ? | + | NA | - | Low/ Moderate |
| **Farooqi, 2022** | NA | NA | + | - | - | ? | + | + | + | NA | - | High |
| **LeGoff, 2023** | NA | NA | + | ? | - | + | + | + | + | NA | - | High |
| **Lunt, 2022** | NA | NA | - | + | + | ? | + | - | + | NA | - | Low/ Moderate |
| **Peter, 2022** | NA | NA | - | ? | + | + | + | + | ? | - | - | Low/ Moderate |
| **Mental Health Service Use Outcomes** | | | | | | | | | | | | |
| **Decio, 2022** | + | + | + | ? | + | ? | ? | ? | + | NA | - | High |
| **Formoso, 2023** | + | + | + | ? | + | ? | + | + | + | NA | - | High |
| **Staples, 2023** | - | + | + | + | - | ? | + | ? | + | NA | - | Low/ Moderate |
| **Ahmed, 2022** | NA | NA | + | ? | - | ? | + | + | ? | - | - | High |
| **Bek, 2023** | NA | NA | + | + | + | ? | + | + | + | NA | - | Low/ Moderate |
| **Benzakour, 2021** | NA | NA | + | ? | - | ? | + | + | + | NA | - | High |
| **Bonazza, 2020** | NA | NA | + | + | - | ? | + | + | + | NA | - | Low/ Moderate |
| **Brehon, 2022** | NA | NA | + | ? | - | ? | + | + | + | NA | - | High |
| **Chopra, 2021** | NA | NA | + | ? | - | ? | + | + | + | NA | - | High |

| | | | | | | | | | | | | |
|---|---|---|---|---|---|---|---|---|---|---|---|---|
| **Chow, 2023** | NA | NA | + | - | + | - | - | ? | + | NA | - | High |
| **Diem, 2022** | NA | NA | + | - | - | ? | ? | + | + | NA | - | High |
| **Farooqi, 2022** | NA | NA | ? | - | - | - | + | + | + | NA | - | High |
| **Frontera, 2022** | NA | NA | + | + | ? | ? | ? | + | ? | NA | - | Low/ Moderate |
| **Gramaglia, 2021** | NA | NA | + | + | - | + | ? | + | + | NA | - | Low/ Moderate |
| **Heeney, 2023** | NA | NA | + | ? | - | ? | ? | + | + | NA | - | High |
| **Hodgson, 2021** | NA | NA | + | ? | ? | ? | ? | + | + | NA | - | High |
| **Holland, 2023** | NA | NA | + | ? | - | ? | ? | + | ? | - | - | High |
| **Huang, 2022** | NA | NA | + | ? | + | ? | ? | + | + | NA | - | Low/ Moderate |
| **Kalyani, 2022** | NA | NA | + | ? | - | ? | + | + | + | NA | - | High |
| **Kisiel, 2023** | NA | NA | + | ? | - | + | + | + | ? | - | - | Low/ Moderate |
| **Lazzaroni, 2022** | NA | NA | + | + | - | ? | + | ? | + | NA | - | Low/ Moderate |
| **LeGoff, 2023** | NA | NA | + | ? | - | ? | + | + | + | NA | - | High |
| **Lynch, 2024** | NA | NA | + | ? | - | ? | + | + | + | NA | - | High |
| **Rival, 2023** | NA | NA | + | - | - | - | + | + | + | NA | - | High |
| **Umbrello, 2022** | NA | NA | + | ? | - | ? | ? | + | + | NA | - | High |
| **Tager, 2023** | NA | NA | + | ? | - | ? | ? | - | + | NA | - | High |

| | | | | | | | | | | | | |
|---|---|---|---|---|---|---|---|---|---|---|---|---|
| **Van Wambeke, 2023** | NA | NA | ? | ? | - | ? | + | + | ? | - | - | High |
| **Wander, 2023** | NA | NA | + | ? | ? | ? | + | - | + | NA | - | High |

+: Yes/low risk of bias; -: No/high risk of bias; ?: Unclear; NA: Not applicable

#### Table S3a: Study Characteristics for Labour Force Outcomes with Control Groups

| Author, Year, Country<br>Study design<br>Funding | Mean age (SD) yrs<br>Female, %<br>Race/ethnicity<br>Enrollment dates | Comorbidities<br>SES variables<br>Employment status | COVID population description | Exposure and control group details |
|---|---|---|---|---|
| Anik, 2023<br>Bangladesh<br><br>Cohort<br>Industry funded: N<br><br>Sample: hospitalized<br><br>Risk of bias: High | 45.3 (14.2)<br>39.7%<br><br>Race: NR<br><br>April, 2020 to December, 2020 | Comorbidity: NR<br><br>SES:<br>Household SES (n = 382)<br>Poor: 40.8%<br>Middle: 29.3%<br>Rich: 29.8%<br><br>Completed years of education<br>Mean (SD): 12.3 (4.8)<br>0–5: 11.6%<br>6–10: 23.3%<br>11–12: 17.5%<br>More than 12 years: 47.6%<br><br>Monthly income (in BDT) (n = 391)<br>Median (Minimum, Maximum): 32,000 (2,000, 700,000)<br>≤20,000: 24.8%<br>20,001–30,000: 24.5%<br>30,001–40,000: 11.8%<br>40,001–60,000: 20.2%<br>60,001+: 18.7%<br><br>Monthly expenditure (in BDT) (n = 436)<br>Median (Minimum, Maximum): 30,000 (2,000, 600,000)<br>≤15,000: 20.6%<br>15,001–25,000: 24.8%<br>25,001–30,000: 14.7%<br>30,001–50,000: 25.2%<br>50,001+: 14.7%<br><br>Employment:<br>In paid employment: 54.3%<br>Not in paid employment: 18.1% | 481 patients admitted to the hospital with symptoms similar to COVID and discharged after recovery<br><br>Population: COVID infected<br><br>Hospitalized: 100%<br><br>Confirmation method: NR | MH description:<br>Screening positive for any one or more symptoms of depression, anxiety, and stress 6 months after being discharged from the hospital after recovery from symptoms similar to COVID<br><br>MH prevalence:<br>Any one or more symptoms of anxiety, depression or stress: 44.9%<br><br>Previous MH: NR<br><br>Ascertainment:<br>Patient reported telephone survey using an electronic questionnaire<br><br>Control:<br>Participants without MH symptoms (no depression, anxiety, or stress) after being discharged from the hospital with symptoms similar to COVID<br><br>55% reported none of the above symptoms |

| Author, Year, Country<br>Study design<br>Funding | Mean age (SD) yrs<br>Female, %<br>Race/ethnicity<br>Enrollment dates | Comorbidities<br>SES variables<br>Employment status | COVID population description | Exposure and control group details |
|---|---|---|---|---|
| | | Homemaker: 27.6% | | |
| | **Findings**<br>Outcome:<br>Decrease in working hours after discharge from hospital with symptoms similar to COVID<br>1. Among paid employees with MH symptoms vs. Paid employees without MH symptoms<br>2. Among all participants (paid, unpaid and homemakers) vs. Total with and without MH<br>Decrease in income due to MH symptoms (any one or more symptoms of depression, anxiety, and stress) after being discharged from the hospital with symptoms similar to COVID<br>1. Among paid employees with MH symptoms vs. Paid employees without MH symptoms<br>2. Among all participants (paid, unpaid and homemakers) vs. Total with and without MH<br><br>Timing: On average six months since discharge from the hospital (average 186 days)<br>Results:<br>1. 70/216 = 32.4% vs 52/265 = 19.6%<br>2. 70/481 = 14.6% vs 52/481 = 10.8%<br>3. 115/216 = 53.2% vs 116/265 = 43.8%<br>4. 115/481 = 23.9% vs 116/481 = 24.1%<br>Adjustments: None | | | |
| Danesh, 2021<br>USA<br><br>Cohort<br>Industry funded: NR<br><br>Sample: outpatient clinic<br><br>Risk of bias: High | Median: 52, IQR: 41-61<br>71%<br><br>Race:<br>White: 79%<br>Black: 12%<br>Others: 9%<br>Hispanic/Latino: 23%<br><br>November 2020 to February 2021 | Comorbidity: NR<br><br>SES: NR | 200 adults with prolonged symptoms of COVID (>14 days after symptom onset) not resolved following usual primary or specialist care who were referred for COVID-specific follow-up through the COVID recovery clinic<br><br>Population: post-COVID with symptoms<br><br>Hospitalized: 30%<br><br>Confirmation method: NR<br><br>Severity<br>Home oxygen: 13% | MH description:<br>Cognitive dysfunction defined as subjective complaints of “brain fog,” including confusion, poor concentration, memory loss, and/or trouble word-finding. Symptoms of COVID (>14 days after symptom onset) not resolved following usual primary or specialist care who were referred for COVID-specific follow-up through the COVID recovery clinic<br><br>MH prevalence: 26%<br><br>Previous MH: NR<br><br>Ascertainment:<br>Semi-structured interview guide used to incorporate common data elements into evolving standard of care documentation. |

| Author, Year, Country<br>Study design<br>Funding | Mean age (SD) yrs<br>Female, %<br>Race/ethnicity<br>Enrollment dates | Comorbidities<br>SES variables<br>Employment status | COVID population description | Exposure and control group details |
|---|---|---|---|---|
| | | | | Consulting specialists from psychiatry—reviewed case narratives written by the internal medicine physicians conducting the post-COVID telemedicine assessments.<br><br>Control:<br>Without cognitive dysfunction |
| | Outcome: Unable to work due to the severity and persistence of COVID sequelae<br>1. With cognitive dysfunction / All with prolonged COVID symptoms vs. Without cognitive dysfunction<br>2. With cognitive dysfunction / All with prolonged COVID symptoms and cognitive dysfunction vs. Without cognitive dysfunction<br>Timing: Initial onset, Median, days: 37; Persisting sequelae, Median days: 82<br>Results:<br>1. 9 / 200 = 4.5% vs. 11 / 200 = 5.5%<br>2. 9 / 51 = 17.7% vs. 11 / 149 = 7.4%<br>Adjustments: None | | | |
| Diem, 2022<br>Switzerland<br><br>Cohort<br>Industry funded: N<br><br>Sample: mixed (support groups, social media, and our post-COVID consultation)<br><br>Risk of bias: Low/moderate | Overall: (n=309)<br>44.6 (NR); range 19-83<br>80.6%<br><br>Nationality:<br>Swiss: 95.1%<br>German: 1.9%<br>French: 1.9%<br>Belgian: 0.6%<br>Austrian: 0.3%<br><br>Insomnia: (n=91)<br>42.5 (NR); range (42.8-47.6)<br><br>Excessive daytime sleepiness:(n=126)<br>43.5 (NR); range (41.3-45.7)<br><br>October 15, 2021 to December 12, 2021 | Comorbidity:<br>Insomnia: 41.8%<br>Excessive daytime sleepiness: 42.9%<br><br>SES:<br>Overall: Education ≥13 years: 59.5%<br><br>Employment:<br>Overall: Self-employment: 12.0% | 309 with persistent symptoms >3 mos. (excluded if not fulfilling WHO post-COVID criteria)<br><br>Population: PCC (WHO) 100%<br><br>Hospitalized: 10.7%<br><br>Insomnia<br>91/241 (37.8%)<br>Labour force: 89/91 (97.8%)<br>Hospitalized: 7.7%<br>Positive PCR/Antigen Test: 83.5%<br>Vaccinated, Yes: 88.9%<br><br>Intubation: 8.9%<br>ICU without intubation: 1.1%<br>Oxygen requirement: 1.1%<br><br>Excessive daytime sleepiness<br>126/224 (56.3%)<br>Labour force: 125/126 (99%)<br>Hospitalized: 11.1%<br>Positive PCR/Antigen Test: 88.1% | MH description:<br>Insomnia: screening positive on the Insomnia Severity Index (ISI), with a cut-off score of ≥15 for clinical insomnia<br><br>Excessive daytime sleepiness: screening positive on the Epworth Sleepiness Scale (ESS), with a cut-off score of ≥11<br><br>MH prevalence:<br>Insomnia: 37.7%<br>Excessive daytime sleepiness: 56.6%<br><br>Previous MH: NR<br><br>Ascertainment:<br>Self report, online survey integrating validated scale<br><br>Control:<br>Without MH symptom (insomnia or excessive daytime sleepiness) |

<table>
<tr><th>Author, Year, Country<br>Study design<br>Funding</th><th>Mean age (SD) yrs<br>Female, %<br>Race/ethnicity<br>Enrollment dates</th><th>Comorbidities<br>SES variables<br>Employment status</th><th>COVID population description</th><th>Exposure and control group details</th></tr>
<tr><td rowspan="2"></td><td></td><td></td><td>Vaccinated, Yes: 81%<br><br>Intubation: 0.8%<br>ICU without intubation: 2.4%<br>Oxygen requirement: 11.1%</td><td></td></tr>
<tr><td colspan="4">Findings<br><u>Outcome</u>:<br>Incapacity to work<br>1. With insomnia / all with insomnia vs. Without insomnia / all without insomnia<br>2. With insomnia / Total with and without insomnia vs. Without insomnia in total sample / Total with and without insomnia<br>3. With excessive daytime sleepiness / all with EDS vs. Without excessive daytime sleepiness / all without EDS<br>4. With excessive daytime sleepiness / Total with and without EDS vs. Without excessive daytime sleepiness in total sample / Total with and without EDS<br>Mean (95% CI) duration in weeks of incapacity to work<br>5. With insomnia vs. Without insomnia<br>6. With excessive daytime sleepiness vs. Without excessive daytime sleepiness<br><u>Timing</u>: Time between onset of acute infection and questionnaire:<br>Insomnia: mean = 13.7 months, 95%CI (12.7 - 14.7) months<br>Excessive daytime sleepiness: mean = 12.8 months, 95%CI (12.0 - 13.6) months<br><u>Results</u>:<br>1. 69 / 89 = 77.5% vs. 93 / 147 = 63.2%; p-value: 0.03<br>2. 69 / 236 = 29.2% vs. 93 / 236 = 39.4%; p-value: 0.386<br>3. 91 / 125 = 72.8% vs. 62 / 96 = 64.6%; p-value: 0.122<br>4. 91 / 221 = 41.2% vs. 62 / 221 = 28.1%; p-value: 0.753<br>5. 29.7 (24.3-35.1); n=69 vs. 25.2 (21.5-30.0); n=93<br>6. 26.3 (22.4-30.1); n=91 vs. 27.9 (22.6-33.4); n=62<br><u>Adjustments</u>: None</td></tr>
<tr><td>Garcia-Molina, 2022<br>Spain<br><br>Cohort<br>Industry funded: N<br><br><u>Sample</u>: outpatient clinic<br><br><u>Risk of bias</u>: Low/Moderate</td><td>51 (12.4)<br>58.5%<br><br><u>Race</u>: NR<br><br>June 2020 to September 2021</td><td><u>Comorbidity</u>: NR<br><br><u>SES</u>:<br>Level of education:<br>Primary: 14.6%<br>Secondary education: 29.3%<br>Further education:56.1%<br><br><u>Employment</u>: NR</td><td>123 accessing outpatient neurorehabilitation programme aimed at patients with PCC, fulfilling NICE criteria of symptoms persisting beyond 12 weeks post-infection<br><br><u>Population</u>: PCC (NICE) 100%<br><br><u>Hospitalized</u>: 46.3%<br><br><u>Confirmation method</u>: PCR 100%<br><br><u>Severity</u>:</td><td><u>MH description</u>:<br>Cognitive alterations detected by neurological exam also underwent treatment (mean 4.1 sessions/week for 8 weeks) for cognitive symptoms; programme included respiratory therapy, physiotherapy, and neuropsychological rehabilitation<br><br><u>MH prevalence</u>:<br>At follow-up for labour force outcomes: 79.7%</td></tr>
</table>

| Author, Year, Country<br>Study design<br>Funding | Mean age (SD) yrs<br>Female, %<br>Race/ethnicity<br>Enrollment dates | Comorbidities<br>SES variables<br>Employment status | COVID population description | Exposure and control group details |
|---|---|---|---|---|
| | | | ICU: 50.9%<br><br>COVID diagnosis (variant):<br>March 2020: 62.6%<br>April 2020: 13%<br>May-December 2020: 21.1%<br>January 2021: 1.6%<br>February 2021: 0.8%<br>April 2021: 0.8% | Previous MH:<br>None of the participants had neurological and/or psychiatric disease prior to COVID infection<br><br>Ascertainment:<br>Semi-structured interviews<br><br>Control:<br>Without MH (symptoms of cognitive alterations) |
| | **Findings**<br>Outcome:<br>Temporarily unable to work (if employed "Has the patient returned to work as normal?"; "Does the patient work in the same conditions as before COVID?")<br>1. Cognitive alterations among total sample vs. Without cognitive alterations among total sample<br>2. Cognitive alterations among all with cognitive alterations vs. Cognitive alterations among all without cognitive alterations<br>Timing: Outcome evaluated at 6-7 months follow-up: n=69 (56.1%); time between infection and treatment onset, mean (SD) = 7.7 (3.5); range 3 - 17.1 months<br>Results:<br>1. 24 / 69 = 34.8% vs. 7 / 69 = 10.1%<br>2. 24 / 55 = 43.6% vs. 7 / 14 = 50%<br>Adjustments: None | | | |
| Green, 2023<br>UK<br><br>Cohort<br>Industry funded: NR<br><br>Sample: PCC clinic<br><br>Risk of bias: High | 51 (NR)<br>55-59 years: 95%<br>63%<br><br>Race: NR<br><br>January to September 2021 | Comorbidity:<br>≥1 comorbidity: 64%<br><br>SES: NR<br><br>Employment:<br>Employed / self-employed: 90%<br>Unemployed: 3%<br>Retired: 6%<br>Student: 2% | 214 Triaged patients referred to a long COVID clinic. The clinic comprised a multidisciplinary team and accepted referrals from both primary and secondary care<br><br>Population: PCC (persistent symptoms > 12 weeks) 100%<br><br>Hospitalized: 12%<br><br>Confirmation method: NR<br><br>Severity:<br>ICU not intubated: 1%<br>ICU intubated: 2% | MH description:<br>Prevalence of brain fog<br><br>MH prevalence: Brain fog: 61%<br><br>Previous MH: NR<br><br>Ascertainment:<br>Self-reported in symptom assessment questionnaire at PCC clinic (based on yes/no response)<br><br>Control:<br>All patients referred to a PCC clinic who did not have brain fog, including students, unemployed, and retired individuals |

| Author, Year, Country<br>Study design<br>Funding | Mean age (SD) yrs<br>Female, %<br>Race/ethnicity<br>Enrollment dates | Comorbidities<br>SES variables<br>Employment status | COVID population description | Exposure and control group details |
|---|---|---|---|---|
| | **Findings**<br>Outcome:<br>At work but struggling with symptoms (group 2)<br>1. Had brain fog / Struggling with symptoms<br>2. Had brain fog / Brain fog, including students, unemployed, and retired vs. Did not have brain fog, including students, unemployed, and retired individuals<br>3. Had brain fog / All referred to PCC clinic, excluding students, unemployed, and retired individuals<br>4. Had brain fog / All patients referred to PCC clinic<br>Off work due to symptoms (group 3)<br>5. Had brain fog / Struggling with symptoms<br>6. Had brain fog / Brain fog, including students, unemployed, and retired vs. Did not have brain fog, including students, unemployed, and retired individuals<br>7. Had brain fog / All referred to PCC clinic, excluding students, unemployed, and retired individuals<br>8. Had brain fog / All patients referred to PCC clinic<br><br>Timing: >12 weeks after the initial infection; median time from diagnosis of the original infection to the first appointment in clinic:<br>• Group 1 (at work): 258 days<br>• Group 2 (at work but struggling with symptoms): 274 days<br>• Group 3 (off work due to symptoms): 285 days<br>Results:<br>1. 50 / 85 = 58.8%<br>2. 50 / 130 = 38.5% vs. 35 / 84 = 41.7%<br>3. 50 / 199 = 21.5%<br>4. 50 / 214 = 23.4%<br>5. 55 / 75 = 73.3%<br>6. 55 / 130 = 42.3% vs. 20 / 84 = 23.8%<br>7. 55 / 199 = 27.6%<br>8. 55 / 214 = 25.7%<br>Adjustments: None | | | |
| Kerksieck, 2023<br>Switzerland<br><br>Cohort<br>Industry funded: N<br><br>Sample: health database<br><br>Risk of bias: | 42.1 (12.2)<br>54.2%<br><br>Nationality:<br>Swiss: 83.6%<br>Non-Swiss: 16.4%<br><br><br>Enrolled upon or shortly after diagnosis between | Comorbidity:<br>1: 16.8%<br>≥2: 4.0%<br><br>SES:<br>• Income (Swiss Francs)<br>• <6,000: 29.2%<br>• 6,000-12,000: 43.7%<br>• >12,000: 176 (27.2)<br><br>Education | 672 individuals with COVID infection and of working age<br><br>Population: post-COVID with symptoms<br>• self-reporting at least one of 23 common PCC-related symptoms: 17.9%<br>• not recovered at 12-mo: 14.2%<br><br>Hospitalized: 1.5% | MH description:<br>• neurocognitive symptom cluster (concentration,<br>• memory, or sleeping problems) (scale 0-no ability to 10-best ability; poor: ≤6, moderate: 7-8, excellent: ≥9),<br>• depression (defined as score ≥10 in depression-related items),<br>• anxiety (score ≥8 in anxiety-related items) |

| Author, Year, Country<br>Study design<br>Funding | Mean age (SD) yrs<br>Female, %<br>Race/ethnicity<br>Enrollment dates | Comorbidities<br>SES variables<br>Employment status | COVID population description | Exposure and control group details |
|---|---|---|---|---|
| Low/moderate | August 2020 and January 2021 | • None/mandatory school:3.3%<br>• Vocational training or specialized baccalaureate: 37.2%<br>• Higher technical/college: 29%<br>• University: 30.6%<br><br>Employment (at infection):<br>• Employed/self-employed: 87.4%<br>• Student: 6.8%<br>• Housewife/family manager: 1.5%<br>• Unemployed: 2.8%<br>• Disability insurance: 0.6%<br>Other: 0.9% | Confirmation Method: self-report<br><br>Severity:<br>1-5 symptoms: 39.6%<br>≥6 symptoms: 48.7%<br>ICU admission: 0.1%<br><br>Re-infection:<br>0%<br><br>Vaccination:<br>All unvaccinated at time of infection | • stress (score ≥15 in stress-related items)<br><br>MH prevalence: 11.9%<br><br>Previous MH:<br>Entire group: history of psychiatric diagnosis (any condition): 13.2%<br>Neurocognitive: 5.5%<br>Depression: 11.9%<br>Anxiety: 10.7<br>Stress: 8.2%<br><br>Ascertainment:<br>Electronic questionnaire, self-report of symptoms and 21-item Depression, Anxiety and Stress Scale<br><br>Control:<br>Without MH (neurocognitive, depression, anxiety or stress) |
| | **Findings**<br>Outcome:<br>Current work ability (all self-reported; compared to highest work ability ever, poor/moderate/excellent work ability)<br>1. Those with neurocognitive vs. Those without neurocognitive<br>2. Those with neurocognitive among all those with COVID<br>3. Those with depression vs. Those without depression<br>4. Those with depression among all those with COVID<br>5. Those with anxiety vs. Those without anxiety<br>6. Those with anxiety among all those with COVID<br>7. Those with stress vs. Those without stress<br>8. Those with stress among all those with COVID<br>Work ability related to mental demand (all self-perceived; based on question ""How do you rate your current work ability with respect to the mental demands of your work?" very bad/rather bad, moderate, rather good, very good)<br>9. Those with neurocognitive vs. Those without neurocognitive<br>10. Those with neurocognitive among all those with COVID<br>11. Those with depression vs. Those without depression<br>12. Those with depression among all those with COVID<br>13. Those with anxiety vs. Those without anxiety<br>14. Those with anxiety among all those with COVID | | | |

| Author, Year, Country Study design Funding | Mean age (SD) yrs Female, % Race/ethnicity Enrollment dates | Comorbidities SES variables Employment status | COVID population description | Exposure and control group details |
|---|---|---|---|---|

15. Those with stress vs. Those without stress
16. Those with stress among all those with COVID

Timing: Survey conducted one year after the infection, symptoms assessed in the past 7 days

Results:

Current work ability

1.

| | With neurocognitive | Without neurocognitive |
|---|---|---|
| Poor | 10/37=27% | 37/635=6% |
| Moderate | 16/37=43.2% | 163/635=26.3% |
| Excellent | 11/37=29.7% | 419/635=67.7% |

2.

| | Among all with COVID |
|---|---|
| Poor | 10/672=1.5% |
| Moderate | 16/672=2.4% |
| Excellent | 11/672=1.6% |

3.

| | With depression | Without depression |
|---|---|---|
| Poor | 24/78=30.8% | 23/577=4% |
| Moderate | 36/78=46.2% | 140/577=24.4% |
| Excellent | 18/78=23.1% | 411/577=71.6% |

4.

| | Among all with COVID |
|---|---|
| Poor | 24/655=3.7% |
| Moderate | 36/655=5.5% |
| Excellent | 18/655=2.7% |

5.

| | With anxiety | Without anxiety |
|---|---|---|
| Poor | 22/70=31.4% | 25/586=4.3% |
| Moderate | 30/70=42.9% | 148/586=25.4% |
| Excellent | 18/70=25.7% | 410/586=70.3% |

6.

| | Among all with COVID |
|---|---|
| Poor | 22/656=3.3% |
| Moderate | 30/656=4.6% |
| Excellent | 18/656=2.7% |

7.

| | With stress | Without stress |
|---|---|---|
| Poor | 21/54=38.9% | 26/602=4.3% |
| Moderate | 24/54=44.4% | 153/602=25.5% |
| Excellent | 9/54=16.7% | 420/602=70.1% |

8.

| | Among all with COVID |
|---|---|
| Poor | 21/656=3.2% |
| Moderate | 24/656=3.7% |
| Excellent | 9/656=1.4% |

Work ability related to mental demand

9.

| | With neurocognitive | Without neurocognitive |
|---|---|---|
| Very bad | 1/37=2.8% | 3/635=0.5% |
| Rather bad | 3/37=8.3% | 4/635=0.6% |
| Moderate | 10/37=27.8% | 62/635=10% |
| Rather good | 14/37=38.9% | 202/635=32.7% |
| Very good | 8/37=22.2% | 346/635=56.1% |

10.

| | Among all with COVID |
|---|---|
| Very bad | 1/672=0.1% |
| Rather bad | 3/672=0.5% |
| Moderate | 10/672=1.5% |
| Rather good | 14/672=2.1% |
| Very good | 8/672=1.2% |

11.

| | With depression | Without depression |
|---|---|---|
| Very bad | 4/78=5.1% | 0/577=0.0% |

12.

| | Among all with COVID |
|---|---|
| Very bad | 4/655=0.6% |

| Author, Year, Country Study design Funding | Mean age (SD) yrs Female, % Race/ethnicity Enrollment dates | Comorbidities SES variables Employment status | COVID population description | Exposure and control group details |
|---|---|---|---|---|

| | | | | | |
|---|---|---|---|---|---|
| Rather bad | 5/78=6.4% | 2/577=0.3% | | Rather bad | 5/655=0.8% |
| Moderate | 34/78=43.6% | 38/577=6.6% | | Moderate | 34/655=5.2% |
| Rather good | 28/78=35.9% | 186/577=32.5% | | Rather good | 28/655=4.6% |
| Very good | 6/78=7.7% | 346/577=60.5% | | Very good | 6/655=0.9% |

13.

| | With anxiety | Without anxiety | 14. | | Among all with COVID |
|---|---|---|---|---|---|
| Very bad | 4/70=5.7% | 0/586=0.0% | | Very bad | 4/656=0.6% |
| Rather bad | 4/70=5.7% | 3/586=0.5% | | Rather bad | 4/656=0.6% |
| Moderate | 29/70=41.4% | 43/586=7.4% | | Moderate | 29/656=4.4% |
| Rather good | 26/70=37.1% | 188/586=32.4% | | Rather good | 26/656=4.0% |
| Very good | 7/70=10% | 346/586=59.7% | | Very good | 7/656=1.1% |

15.

| | With stress | Without stress | 16. | | Among all with COVID |
|---|---|---|---|---|---|
| Very bad | 2/54=3.8% | 2/602=0.3% | | Very bad | 2/656=0.3% |
| Rather bad | 6/54=11.3% | 1/602=0.2% | | Rather bad | 6/656=0.9% |
| Moderate | 26/54=49.1% | 46/602=7.7% | | Moderate | 26/656=4% |
| Rather good | 16/54=30.2% | 199/602=33.3% | | Rather good | 16/656=2.4% |
| Very good | 3/54=5.7% | 349/602=58.5% | | Very good | 3/656=0.5% |

Adjustments: None

| Author, Year, Country Study design Funding | Mean age (SD) yrs Female, % Race/ethnicity Enrollment dates | Comorbidities SES variables Employment status | COVID population description | Exposure and control group details |
|---|---|---|---|---|
| Lemhofer, 2023<br>Germany<br><br>Cohort<br>Industry funded: N<br><br>Sample: PCC outpatient clinic<br><br>Risk of bias: Low/moderate | 46.9 (10.9)<br>68.9%<br>NR<br><br>March 1, 2021 to October 29, 2021 | Comorbidity: NR<br><br>SES: NR<br><br>Employment: NR | 318 patients attending post-COVID outpatient clinic; with an average of 7.8 symptoms at 12 weeks<br><br>Population: PCC (symptom evaluation) 100%<br><br>Hospitalized: 32.6%<br><br>Confirmation Method: PCR 100% | MH description: Deficits in the mental component summary score of the SF-36<br><br>MH prevalence: 50.9%<br><br>Previous MH: NR<br><br>Ascertainment: Survey completed in person at clinic<br><br>Control: Without MH |

**Findings**
Outcome:
Unable to work
1. With MH deficit / All attending post-COVID clinic vs. All attending post-COVID clinic
2. With MH deficit / All attending post-COVID clinic with MH deficit vs. Attending post-COVID clinic without MH deficit

Timing: majority of sample was 3 to 12+ months since COVID infection (6 participants were <3 months)
Results:
1. 74 / 316 = 23.4% vs. 52 / 316 = 16.5%

| Author, Year, Country<br>Study design<br>Funding | Mean age (SD) yrs<br>Female, %<br>Race/ethnicity<br>Enrollment dates | Comorbidities<br>SES variables<br>Employment status | COVID population description | Exposure and control group details |
|---|---|---|---|---|
| | 2. 74 / 161 = 46% vs. 52 / 155 = 33.5%<br>Adjustments: None | | | |
| O'Sullivan, 2023<br>UK<br><br>Cohort<br>Industry funded: NR<br><br>Sample: outpatient recovery service<br><br>Risk of bias: Low/moderate | 36.6 (10)<br>13.5%<br><br>Race: NR<br><br>August 2020 to December 2021 | Comorbidity: NR<br><br>SES:<br>Military rank as proxy for SES:<br>Junior NCO: 35.6%<br>Senior NCO: 31.5%<br>Officer rank: 30.2%<br><br>Employment: military employees | 222 military employees with ongoing symptomatic or initially severe COVID infection<br><br>Population: post-COVID with symptoms (ongoing symptoms, physical limitations and neurocognitive impact at 6 months post-COVID)<br><br>Hospitalized: 36.9%<br><br>Confirmation method: NR<br><br>Severity:<br>Mean (SD) number of acute symptoms: 3 (1) | MH description: All self-reported:<br>• cognitive impairment based on presence of 'any' cognitive symptoms<br>• individuals medically downgraded to perform duties due to MH conditions for poor memory, poor concentration, poor attention, and confusion<br><br>MH prevalence: 5.8%<br><br>Previous MH:<br>Past MH condition: 7.7%<br><br>Ascertainment:<br>Comprehensive assessment, including symptom questionnaire, patient-reported outcome measures (validated tools for anxiety and PTSD)<br><br>Control: Without MH |
| | **Findings**<br>Outcome: Medically downgraded to perform duties, based on medical deployment status to determine ability to perform job role without restriction ('fully deployable') or with limitations ('medically downgraded')<br>1. Cognitive symptoms / All (medically downgraded and fully deployable) with cognitive symptoms vs. All without cognitive symptoms<br>2. Cognitive symptoms among all (medically downgraded and fully deployable)<br>3. Poor memory / All (medically downgraded and fully deployable) with poor memory vs. All without poor memory<br>4. Poor memory among all (medically downgraded and fully deployable)<br>5. Poor concentration / All (medically downgraded and fully deployable) with poor concentration vs. All without poor concentration<br>6. Poor concentration among all (medically downgraded and fully deployable)<br>7. Poor attention / All (medically downgraded and fully deployable) with poor attention vs. All without poor attention<br>8. Poor attention among all (medically downgraded and fully deployable)<br>9. Confusion /All (medically downgraded and fully deployable) with confusion vs. All without confusion<br>10. Confusion among all (medically downgraded and fully deployable)<br>Timing: 6 months after initial COVID infection<br>Results:<br>1. 80 / 97 = 82.5% vs. 76 / 125 = 60.8% | | | |

| Author, Year, Country<br>Study design<br>Funding | Mean age (SD) yrs<br>Female, %<br>Race/ethnicity<br>Enrollment dates | Comorbidities<br>SES variables<br>Employment status | COVID population description | Exposure and control group details |
|---|---|---|---|---|
| | 2. 80 / 222 = 36%<br>3. 46 / 57 = 80.7% vs. 110 / 165 = 66.7%<br>4. 46 / 222 = 20.7%<br>5. 55 / 63 = 87.3% vs. 101 / 159 = 63.5%<br>6. 55 / 222 = 24.8%<br>7. 70 / 79 = 88.6% vs. 86 / 143 = 60.1%<br>8. 70 / 222 = 31.5%<br>9. 48 / 57 = 84.2% vs. 108 / 165 = 65.5%<br>10. 48 / 222 = 21.6%<br>Adjustments: None | | | |
| Romero-Rodrigue, 2023<br>Spain<br><br>Cohort<br>Industry funded: N<br><br>Sample: community<br><br>Risk of bias: High | Full cohort:<br>(n=689)<br>46 (NR), IQR: 40-52<br>83.2%<br><br>Active workers:<br>(n=596)<br>48 (NR) IQR: 40-52<br>83%<br><br>Race:<br><br>March 2021 to January 2022<br>(Data collection) | Comorbidity: NR<br><br>SES:<br>Region (Autonomous Community) of residence<br>Madrid: 26%<br>Andalusia: 19%<br>Aragon: 12%<br>Catalonia: 11%<br>Valencian: 6.1%<br>Other: 27%<br><br>Employment: 83% employed | 689 with COVID and 596/689 (86.5%) active workers during the course of the disease (not unemployed or retired)<br><br>Population: PCC (WHO) 100%<br>Time since PCC:<br><361 days: 31%<br>≥361 days: 69%<br><br>Hospitalized: 23%<br><br>Confirmation method:<br>PCR: 60%<br>Clinically: 19%<br>Rapid antigen test: 17%<br>Serological test: 12%<br><br>Severity:<br>ICU: 3.6%<br>Pneumonia COVID: 30%<br><br>Vaccination: 87% | MH description:<br>Mental confusion and PTSD as part of persistent COVID signs and symptoms<br><br>MH prevalence:<br>Mental confusion of those able to work before COVID: 58.9%<br>Overall population: 68%<br><br>Previous MH:<br>Depression: 27%,<br>Anxiety: 45%,<br>Mental illness (psychosis, neurosis, Alzheimer's): 4.8%<br><br>Ascertainment:<br>Online questionnaire<br><br>Control:<br>Without MH symptoms |
| | **Findings**<br>Outcome: Some sick leave due to PCC<br>1. Mental confusion for active workers / Active workers vs. Active workers without mental confusion<br>2. Mental confusion for active workers / Active workers with mental confusion vs. Active workers without mental confusion<br>3. PTSD for active workers / Active workers vs. Active workers without PTSD<br>4. PTSD for active workers / All with PTSD vs. Active workers without PTSD<br>Timing: NR (PCC defined as 12 weeks of persistent symptom)<br>Results: | | | |

| Author, Year, Country<br>Study design<br>Funding | Mean age (SD) yrs<br>Female, %<br>Race/ethnicity<br>Enrollment dates | Comorbidities<br>SES variables<br>Employment status | COVID population description | Exposure and control group details |
|---|---|---|---|---|
| | 1. 285 / 596 = 47.8% vs. 182 / 596 = 30.5%<br>2. 285 / 351 = 81.2% vs. 182 / 245 = 74.3%<br>3. 120 / 596 = 20.1% vs. 347 / 596 = 58.2%<br>4. 120 / 141 = 85.1% vs. 347 / 455 = 76.3%<br><u>Adjustments</u>: None | | | |
| Thompson, 2023<br>USA<br><br>Cohort<br>Industry funded: N<br><br><u>Sample</u>: mix (outpatient recovery program and community)<br><br><u>Risk of bias</u>: Low/moderate | 42 (NR)<br>69.5%<br><br><u>Race</u>:<br>White: 71.2%<br>Hispanic: 13.6%<br>African American: 6.8%<br>Asian/South Asian: 6.8%<br>Other: 1.7%<br><br>NR | <u>Comorbidity</u>:<br>Mean (SD) comorbidities, for those with:<br>Time off: 1.6 (1.4)<br>No time off: 1.2 (1.4)<br><br><u>SES</u>:<br>Mean (SD) years education<br>Time off: 15.86 (2.24)<br>No time off: 16.00 (2.13)<br><br><u>Employment</u>: participants falling under broad skill level 3 and 4 which is predominantly professionals: 61% | 59 with PCC symptoms; recruited from Post-COVID recovery program (59.3%) and community (40.7%);<br><br><u>Population</u>: PCC (symptom evaluation) 100%<br><br><u>Hospitalized</u>: 6.7%<br><br><u>Confirmation method</u>: 100% naso, antibody | <u>MH description</u>:<br>Clinically significant depression, determined by scoring ≥10 on the Patient Health Questionnaire (PHQ-9)<br><br><u>MH prevalence</u>:<br>Clinically significant depression: 55.9%<br><br><u>Previous MH</u>:<br>Prior psychiatric history:<br>Time off: 46.4%<br>No time off: 45.2%<br><br><u>Ascertainment</u>:<br>Self-reported questionnaire, supervised by a neuropsychologist<br><br><u>Control</u>:<br>Without clinically significant depression, determined by not meeting criteria on PHQ-9 |
| | **Findings**<br><u>Outcome</u>:<br>Taking time off for PCC symptoms beyond the required quarantine period<br>1. With clinically significant depression / Total sample vs. Without clinically significant depression / Total sample<br>2. With clinically significant depression / All with clinically significant depression vs. Without clinically significant depression / All without clinically significant depression<br>Occupational performance suffered upon RTW from PCC symptoms (all previously employed)<br>3. With clinically significant depression / Total sample vs. Without clinically significant depression / Total sample<br>4. With clinically significant depression / All with clinically significant depression vs. Without clinically significant depression / All without clinically significant depression<br><u>Timing</u>: Survey administered after quarantine, at least 10-20 days after symptom onset<br><u>Results</u>:<br>1. 22 / 59 = 37.3% vs. 6 / 59 = 10.2%<br>2. 22 / 33 = 66.7% vs. 6 / 26 = 23.1% | | | |

| Author, Year, Country<br>Study design<br>Funding | Mean age (SD) yrs<br>Female, %<br>Race/ethnicity<br>Enrollment dates | Comorbidities<br>SES variables<br>Employment status | COVID population description | Exposure and control group details |
|---|---|---|---|---|
| | 3. 19 / 59 = 32.2% vs. 9 / 59 = 15.3%<br>4. 19 / 33 = 57.6% vs. 9 / 26 = 34.6%<br>Adjustments: None | | | |
| Tsuchida, 2023<br>Japan<br><br>Cohort<br>Industry funded: N<br><br>Sample: post-COVID outpatient clinic<br><br>Risk of bias: High | 41.6 (NR)<br>56.9%<br><br>Race: NR<br><br>January 18, 2021 to May 30, 2022 | Comorbidity: 58.1% with comorbidity<br><br>SES: NR<br><br>Employment: NR | 497 post-COVID patients visiting a PCC outpatient clinic treating patients suspected of having COVID sequelae and referred from elsewhere<br><br>Population: post-COVID with symptoms (residual symptoms lasting at least 2 months post-infection)<br><br>Hospitalized: NR<br><br>Confirmation method: PCR 100% | MH description:<br>Post-COVID psychological symptom cluster (cluster 3 comprised of many psychological symptoms, including anxiety, depressed mood, forgetfulness, insomnia in addition to fatigue, headache, and lack of motivation)<br><br>MH prevalence:<br>Anxiety: 42.3%<br>Depressed mood: 40.2%<br>Forgetfulness: 37.8%<br>Insomnia: 29.6%<br><br>Previous MH: NR<br><br>Ascertainment:<br>Cluster classification of post-COVID symptoms was performed based on symptoms described in the questionnaire at the time of the hospital visit<br>Employment status self-reported<br><br>Control:<br>Patients in cluster 1 (fatigue only), cluster 2 (fatigue, dyspnea, chest pain, palpitations, forgetfulness), cluster 4 (hair loss only), and cluster 5 (taste and smell disorders only) |
| | **Findings**<br>Outcome:<br>Current employment status: change in job description<br>1. Outcome in cluster 3 / All in cluster 3 vs. Outcome in other clusters / All in other clusters<br>2. Outcome in cluster 3 / All in cluster 1-5<br>Current employment status: leave of absence | | | |

| Author, Year, Country<br>Study design<br>Funding | Mean age (SD) yrs<br>Female, %<br>Race/ethnicity<br>Enrollment dates | Comorbidities<br>SES variables<br>Employment status | COVID population description | Exposure and control group details |
|---|---|---|---|---|
| | 3. Outcome in cluster 3 / All in cluster 3 vs. Outcome in other clusters / All in other clusters<br>4. Outcome in cluster 3 / All in cluster 1-5<br>Timing: During the 1st year following presentation at the post-COVID outpatient clinic<br>Results:<br>1. 10 / 103 = 9.7% vs. 46 / 393 = 11.7%<br>2. 10 / 496 = 2%<br>3. 41 / 103 = 39.8% vs. 73 / 393 = 18.6%<br>4. 41 / 496 = 8.3%<br>Adjustments: Overall p< 0.001 for current employment status across 5 clusters | | | |
| Vanichkachorn, 2021<br>USA<br><br>Cohort<br>Industry funded: NR<br><br>Sample: outpatient clinic<br><br>Risk of bias: High | 45.4 (14.2)<br>68%<br><br>Race: NR<br><br>June 1, 2020 to December 31, 2020 | Comorbidity: NR<br><br>SES:<br>60,001+: 18.7%<br><br>Employment:<br>Fully employed: 91%<br>Not employed: 9% | 100 patients with PCC participating in COVID Activity Rehabilitation Program-CARP<br><br>Population: PCC (symptom evaluation) 100%<br><br>Hospitalized: 25%<br><br>Confirmation method: PCR 93%; antibody test 8% | MH description:<br>Cognitive symptoms were defined as any subjective reports of concentration difficulty, perceived memory loss, word finding difficulty, and inability to effectively multitask. Presenting symptom of sleep disturbance.<br><br>MH prevalence: Cognitive impairment: 45%<br><br>Previous MH:<br>Pre-existing anxiety/depression: 34%<br><br>Ascertainment:<br>Presenting symptoms were recorded from clinical documentation<br><br>Control:<br>Without MH and employed prior to COVID |
| | **Findings**<br>Outcome:<br>Return to work in any capacity (including reduced hours or accommodated functional activities, at their usual work site or an alternative location such as home)<br>1. Those with cognitive impairment / All with PCC vs. Those without cognitive impairment<br>2. Those with cognitive impairment / All with PCC and cognitive impairment vs. With PCC and without cognitive impairment<br>3. Those with cognitive impairment / All with PCC and employed before COVID infection vs. PCC, employed and without cognitive impairment<br>4. Those with sleep disturbance / All with PCC vs. Those without sleep disturbance<br>5. Those with sleep disturbance / All with PCC and sleep disturbance vs. With PCC and without sleep disturbance | | | |

| Author, Year, Country<br>Study design<br>Funding | Mean age (SD) yrs<br>Female, %<br>Race/ethnicity<br>Enrollment dates | Comorbidities<br>SES variables<br>Employment status | COVID population description | Exposure and control group details |
|---|---|---|---|---|
| | 6. Those with sleep disturbance / All with PCC and employed before COVID infection vs. PCC, employed and without sleep disturbance<br><br><u>Timing</u>: Mean time to presentation: 93.4 ± 65.2 days<br><u>Results</u>:<br>1. 29 / 100 = 29% vs. 34 / 100 = 34%<br>2. 29 / 45 = 64.4% vs. 34 / 55 = 61.8%<br>3. 29 / 91 = 31.9% vs. 34 / 91 = 37.4%<br>4. 18 / 100 = 18% vs. 45 / 100 = 45%<br>5. 18 / 30 = 60% vs. 45 / 70 = 64.3%<br>6. 18 / 91 = 19.8% vs. 45 / 91 = 49.5%<br><u>Adjustments</u>: None | | | |

### Table S3b: Study Characteristics for Labour Force Outcomes without Control Groups

<table>
<tr><th>Author, Year, Country<br>Study design<br>Funding</th><th>Mean age (SD) yrs<br>Female, %<br>Race/ethnicity<br>Enrollment dates</th><th>Comorbidities<br>SES variables<br>Employment status</th><th>COVID population description</th><th>MH details</th></tr>
<tr><td rowspan="2">Braga, 2022<br>Brazil<br><br>Cross-sectional<br>Industry funded: NR<br><br><u>Sample</u>: outpatient hospital unit<br><br><u>Risk of bias</u>: Low/moderate</td><td>47.6 (11.2)<br>73%<br><br><u>Race</u>: NR<br><br>April 2021 to January 2022</td><td><u>Comorbidity</u>: NR<br><br><u>SES</u>:<br>Education (years)<br>5–8: 5%<br>9–11: 6%<br>12–15: 35%<br>16+: 54%<br><br><u>Employment</u>:<br>Active: 74%<br>Retired: 12%<br>Unemployed: 10%<br>Sick Leave/Disability: 4%</td><td>614 patients with a diagnosis of COVID seeking treatment for cognitive issues<br><br><u>Population</u>: COVID infected<br><br><u>Hospitalized</u>: 33.3%<br><br><u>Confirmation method</u>: PCR 100%<br><br><u>Severity</u>:<br>ICU: 17.3%<br>Intubated for oxygen support: 10.6%</td><td><u>MH description</u>:<br>Treatment for cognitive issues (attention, concentration, and memory difficulties, slow reasoning, black-outs, etc.) only diagnosed after the COVID and impacting daily lives following COVID<br><br><u>MH prevalence</u>: 100%, all had memory problems after COVID<br><br><u>Previous MH</u>: NR<br><br><u>Ascertainment</u>: medical charts</td></tr>
<tr><td colspan="4">Findings<br><u>Outcome</u>:<br>Sick leave in patients with MH symptoms impacting daily activities/ All patients with MH symptoms impacting daily activities<br><u>Timing</u>: 8 (4.3) months from COVID diagnosis<br><u>Results</u>: 26/614 = 4.2%<br><u>Adjustments</u>: None</td></tr>
<tr><td>Davis, 2021<br>UK<br><br>Cross-sectional<br>Industry funded: N<br><br><u>Sample</u>: support groups and social media<br><br><u>Risk of bias</u>: High</td><td>NR<br><br>18-29 y: 7.4%<br>30-39 y: 24.1%<br>40-49 y: 31.0%<br>50-59 y: 25.0%<br>60-69 y: 10.1%<br>70-79 y: 2.3%<br>80+ y: 0.3%<br><br>78.9%<br><br><u>Race</u>:<br>White: 85.3%<br>Hispanic, Latino, Spanish: 3.7%<br>Asian, South Asian, SE Asian: 3.3%</td><td><u>Comorbidity</u>:<br>At least one pre-existing condition: 83.0%<br><br><u>SES</u>:<br>Participants in USA earning more than $85,000/year: 51.0%<br>Participants in USA earning more than $150,000/year: 22.5%<br>Participants from elsewhere earning less than €20,000/year: 25%<br>Participants from elsewhere earning less than €40,000/year: 51.1%</td><td>3762 with symptoms<br><br><u>Population</u>: post-COVID with symptoms (lasting longer than 28 days)<br><br><u>Hospitalized</u>: 8.4%<br><br><u>Confirmation method</u>: some received both tests<br>Diagnostic (RT-PCR/antigen): 61.9%<br>Antibody (IgG, IgM or both): 57.6%<br><br><u>Severity</u>:<br>Visited ER or urgent care but not admitted: 34.9%</td><td><u>MH description</u>:<br>Cognitive dysfunction (sudden confusion/disorientation, agnosia, brain fog, difficulty thinking, difficulty making decision, poor attention, etc.), memory loss (forgetting how to do routine tasks, long-term memory loss, inability to make new memories, etc.), or both across all age groups<br><br><u>MH prevalence</u>:<br>Neuropsychiatric - Cognitive Dysfunction: 85.4%<br>Neuropsychiatric - Memory: 72.8%<br><br><u>Previous MH</u>: NR<br><br><u>Ascertainment</u>: Online survey</td></tr>
</table>

<table>
<tr><th>Author, Year, Country<br>Study design<br>Funding</th><th>Mean age (SD) yrs<br>Female, %<br>Race/ethnicity<br>Enrollment dates</th><th>Comorbidities<br>SES variables<br>Employment status</th><th>COVID population description</th><th>MH details</th></tr>
<tr><td></td><td>Black:2.0%<br>Middle Eastern, North African: 1.7%<br>Indigenous Peoples: 1.6%<br>Pacific Islander: 0.1%<br>Other: 2.5%<br><br>September 6, 2020 to November 25, 2020</td><td><u>Employment</u>: NR</td><td></td><td></td></tr>
<tr><td></td><td colspan="4">Findings<br><u>Outcome</u>: Work ability for participants with suspected/confirmed COVID among those who worked<br><u>Timing</u>: Assessment at 1-4 weeks and 2-7 months after the onset of illness<br><u>Results</u>:
<table>
<tr><td colspan="6">Mildly unable to work</td></tr>
<tr><td>18-29 y</td><td>30-39 y</td><td>40-49 y</td><td>50-59 y</td><td>60-69 y</td><td>70+ y</td></tr>
<tr><td>34.5%</td><td>29.5%</td><td>30.6%</td><td>29.3%</td><td>30.2%</td><td>39.7%</td></tr>
<tr><td colspan="6">Moderately unable to work</td></tr>
<tr><td>31.4%</td><td>29.2%</td><td>29.2%</td><td>25.7%</td><td>26.5%</td><td>15.3%</td></tr>
<tr><td colspan="6">Severely unable to work</td></tr>
<tr><td>26.7%</td><td>32.5%</td><td>30.3%</td><td>31.2%</td><td>26.8%</td><td>11.7%</td></tr>
</table>
<u>Adjustments</u>: None</td></tr>
<tr><td>Delgado-Alonso, 2022<br>Spain<br><br>Cross-sectional<br>Industry funded: N<br><br><u>Sample</u>: PCC program<br><br><u>Risk of bias</u>: Low/moderate</td><td>46.3 (8.0)<br>87%<br><br><u>Race</u>: NR<br><br>January to June, 2022</td><td><u>Comorbidity</u>: NR<br><br><u>SES</u>:<br>Mean (SD) years of education: 16.14 (3.03)<br><br>ISCO 08 classification of occupation categories:<br>• Managers professionals: 3.9%<br>• Professionals: 57.1%<br>• Technicians and associate professionals: 19.5%<br>• Service and sales workers: 13.0%<br>• Plant and machine operators and assemblers: 1.3%</td><td>77 with PCC attending program for PCC in the Department of Neurology<br><br><u>Population</u>: PCC (WHO) 100%<br><br><u>Hospitalized</u>: 19.5%<br><br><u>Confirmation method</u>: PCR 100%<br><br><u>Severity</u>:<br>ICU admission: 4%<br>Ventilatory assistance: 5.2%<br><br><u>Re-infection</u>: 29.9%</td><td><u>MH description</u>:<br>Cognitive issues influencing their work capacity and professional career, based on mean scores of each scale<br><br><u>MH prevalence</u>:<br>Excluded those with active psychiatric disorder not explained by PCC<br><br><u>Previous MH</u>: NR<br><br><u>Ascertainment</u>:<br>Examined by a trained neuropsychologist</td></tr>
</table>

<table>
<tr><th>Author, Year, Country<br>Study design<br>Funding</th><th>Mean age (SD) yrs<br>Female, %<br>Race/ethnicity<br>Enrollment dates</th><th>Comorbidities<br>SES variables<br>Employment status</th><th>COVID population description</th><th>MH details</th></tr>
<tr><td rowspan="2"></td><td></td><td>• Elementary occupations: 5.2%<br><br><u>Employment</u>: active employment prior to infection</td><td></td><td></td></tr>
<tr><td colspan="4">Findings<br><u>Outcome</u>:<br>Work capacity and professional career influenced by:<br>1. Cognitive issues and actively working before COVID / All actively working before COVID<br>2. Sleep disorders and actively working before COVID / All actively working before COVID<br>3. Anxiety/depression and actively working before COVID / All actively working before COVID<br><u>Timing</u>: 20.7 ± 6.5 months after clinical onset<br><u>Results</u>:<br>1. 71 / 77 = 92.2%<br>2. 54 / 77 = 70.1%<br>3. 42 / 77 = 54.5%<br><u>Adjustments</u>: None</td></tr>
<tr><td rowspan="2">Dressing, 2022<br>Germany<br><br>Cross-sectional<br>Industry funded: N<br><br><u>Sample</u>: outpatient clinic<br><br><u>Risk of bias</u>: Low/moderate</td><td>53.6 (12)<br>64.5%<br><br><u>Race</u>: NR<br><br><br>June 16, 2020 to January 29, 2021</td><td><u>Comorbidity</u>: NR<br><br><u>SES</u>:<br>Mean (SD) years of education: 13.6 (2.6), range 8-18<br><br><u>Employment</u>: NR</td><td>31 admitted to outpatient clinic with PCC and neurocognitive symptoms in the chronic phase (>3 mo) after COVID<br><br><u>Population</u>: PCC (NR) 100%<br><br><u>Hospitalized</u>: NR<br><br><u>Confirmation method</u>: PCR 100%<br><br><u>Severity</u>:<br>Endotracheal ventilation at ICU: 13%</td><td><u>MH description</u>:<br>All admitted to outpatient clinic due to lasting neurocognitive symptoms<br><br><u>MH prevalence</u>: 100%<br><br><u>Previous MH</u>:<br>None with pre-existing neurodegenerative disorders<br><br><u>Ascertainment</u>:<br>Neurologic deficits were examined in a complete neurologic assessment by a board-certified neurologist</td></tr>
<tr><td colspan="4">Findings<br><u>Outcome</u>:<br>1. Reduced work quota with neurocognitive symptoms persisting after COVID / All with neurocognitive symptoms persisting after COVID<br>2. Inability to work and impairment in daily activities with neurocognitive symptoms persisting after COVID / All with neurocognitive symptoms persisting after COVID<br><u>Timing</u>: Neurologic exam = 202.3 ± 57.5 d after first positive COVID PCR<br><u>Results</u>:<br>1. 3 / 31 = 9.7%</td></tr>
</table>

| Author, Year, Country<br>Study design<br>Funding | Mean age (SD) yrs<br>Female, %<br>Race/ethnicity<br>Enrollment dates | Comorbidities<br>SES variables<br>Employment status | COVID population description | MH details |
|---|---|---|---|---|
| | 2. 9 / 31 = 29%<br>Adjustments: None | | | |
| Farooqi, 2022<br>USA<br><br>Cross-sectional<br>Industry funded: N<br><br>Sample: outpatient clinic<br><br>Risk of bias: High | 51.3 (12.6)<br>87%<br><br>Race:<br>Caucasian: 53%<br>Black: 13%<br>Hispanic: 13%<br>Asian: 3%<br>Other: 20%<br><br>December 1, 2020 to July 26, 2021 | Comorbidity:<br>Mean (SD) number of medical comorbidities: 1.13 (0.96)<br><br>SES: NR<br><br>Employment:<br>Currently employed: 23%<br>Not employed/retired before pandemic: 17%<br>Significant interruption: 60% | 30 patients referred and assessed for psychiatric complications of COVID to the outpatient department of an academic Behavioral Health Center<br><br>Population: PCC (symptom evaluation) 100%<br><br>Hospitalized: 36.7%<br><br>Confirmation method: NR<br><br>Severity:<br>Moderate illness constituting pulmonary symptoms, short-term hospitalization, or significant symptoms while at home: 33.3%<br>Significant illness requiring intubation for hypoxia and prolonged hospitalization: 3.3%<br><br>COVID Diagnosis (variant):<br>February-March 2020: 30%<br>January-February 2021: 30%<br>January 2020-March 2021: 40% | MH description:<br>Adult patients complaining of neuropsychiatric symptoms (anxiety, depression, fatigue, and cognitive problems)<br><br>MH prevalence:<br>Psychiatric diagnosis: 87%<br>Anxiety disorder: 17%<br>Depressive disorder: 47%<br>Anxiety/depressive disorder: 7%<br>Adjustment disorder: 17%<br><br>Previous MH:<br>Past psychiatric history: 56.7%<br><br>Ascertainment: Self-report |
| | **Findings**<br>Outcome:<br>1. Those experiencing a significant interruption in their professional lives / All COVID patients with MH symptoms<br>2. Unable to return to work for all COVID patients with MH symptoms and significant interruptions in professional lives / All COVID patients with MH symptoms<br>Timing: Mean (SD) months from COVID diagnosis to timing of consult: 7.23 (4.89)<br>Results:<br>1. 19 / 30 = 63.3%<br>2. 6 / 30 = 20%<br>Adjustments: None | | | |
| LeGoff, 2023<br>USA | 48.3 (NR)<br>54.7% | Comorbidity: NR<br><br>SES: NR | 67 California employees referred by occupational physician's outpatient MH provider panel for assessment and | MH description:<br>"Brain fog" including mental fatigue, forgetfulness/impaired short-term |

| Author, Year, Country<br>Study design<br>Funding | Mean age (SD) yrs<br>Female, %<br>Race/ethnicity<br>Enrollment dates | Comorbidities<br>SES variables<br>Employment status | COVID population description | MH details |
|---|---|---|---|---|
| Cross-sectional<br>Industry funded: NR<br><br>Sample: outpatient MH clinic<br><br>Risk of bias: High | Race: NR<br><br>January 2021 to December 2022 (referral cases)<br><br>April 2020 to February 2022 (contracted COVID) | <br>Employment:<br>Occupational Category:<br>Healthcare services: 50%<br>Corrections: 21.9%<br>First responders: 14.1%<br>Facilities maintenance: 14.1% | treatment due to diagnosis of PCC with neurocognitive complaints of "brain fog," including mental fatigue, forgetfulness/impaired short-term memory, difficulties concentrating, inattentiveness, diminished task focus, confusion, etc.<br><br>Population: PCC (neurocognitive screening) 100%<br><br>Hospitalized: NR<br><br>Confirmation method: NR | memory, difficulties concentrating, inattentiveness, diminished task focus, confusion were affecting returned to work at time of initial referral for NCSE (study enrollment), with or without restrictions<br><br>MH prevalence:<br>100%, All with neurocognitive complaints<br><br>Previous MH: NR<br><br>Ascertainment:<br>Self-reported neurocognitive complaints after PCC diagnosis |
| | **Findings**<br>Outcome:<br>1. Returned to work at time of initial referral for neurocognitive screening/ Assessed by neurocognitive screening with MH symptoms<br>2. Mean duration of work leave from doctor's first report illness to date of return to full or modified duty<br>Timing: The sample mean total duration of illness, from date of initial diagnosis to date of discharge from all healthcare services due to being at MMI, was 16 months (462.8 days; SD, 187.2 days), with a range of 4 1/2 months (134 days) to 2 1/2 years (903 days)<br>Results:<br>1. 0/64 = 0%<br>2. 459.4 days (15 months: from 19 weeks to 2.5 yrs [range 134-903 days [SD 126.6 days]]; median 425.5 days)<br>Adjustments: None | | | |
| Lunt, 2022<br>UK<br><br>Cross-sectional<br>Industry funded: NR<br><br>Sample: mix (community and COVID support group)<br><br>Risk of bias: | NR, range 25-65<br>88%<br><br>Race: NR<br><br>March 16, 2020 to July 1, 2020 | Comorbidity: NR<br><br>Employment:<br>Nurses: 17%<br>Medics: 15%<br>Allied health: 10%<br>Teachers: 12%<br>Social or support workers: 6% | 145 workers who had either tested positive for COVID or suspected infection<br><br>Population: COVID infected<br><br>Hospitalized: 12%<br><br>Confirmation method: NR<br><br>Severity<br>Mild/moderate at home: 35%<br>Severe at home: 45% | MH description:<br>Poor concentration and symptom interaction with cognitive job demands (applied to having to 'concentrate,' 'word find' and to meta-cognitive task')<br><br>Psychological workability/return-to-work status determined by asking: "How do you rate your current workability with respect to the psychological and demands of your work? |

<table>
<tr><th>Author, Year, Country<br>Study design<br>Funding</th><th>Mean age (SD) yrs<br>Female, %<br>Race/ethnicity<br>Enrollment dates</th><th>Comorbidities<br>SES variables<br>Employment status</th><th>COVID population description</th><th>MH details</th></tr>
<tr><td rowspan="2">Low/moderate</td><td></td><td></td><td></td><td><u>MH prevalence</u>: over half (52%) of participants reported pre-existing mental or physical health conditions<br><u>Previous MH</u>: 6%<br><br><u>Ascertainment</u>: open-ended interviews</td></tr>
<tr><td colspan="4"><u>Outcome</u>:<br>Return to work<br>1. Poor concentration leading to obstacles / Total sample<br>2. Symptom interaction with cognitive job demands / Total sample<br>Psychological workability<br>3. Reporting outcome on scale / All reporting outcome<br><u>Timing</u>: More than 6 months previously with their symptoms continuing for longer than 6 months, 65%<br><u>Results</u>:<br>1. 20 / 145 = 13.8%<br>2. 12 / 145 = 8.3%<br>3.<br><table><tr><td>Very good</td><td>Rather good</td><td>Moderate</td><td>Rather poor</td><td>Poor</td></tr><tr><td>2 / 88 = 2.3%</td><td>14 / 88 = 16%</td><td>25 / 88 = 28.4%</td><td>27 / 88 = 30.7%</td><td>20 / 88 = 22.7%</td></tr></table><br><u>Adjustments</u>: None</td></tr>
<tr><td>Peter, 2022<br>Germany<br><br>Cross-sectional<br>Industry funded: N<br><br><u>Sample</u>: health database<br><br><u>Risk of bias</u>: Low/moderate</td><td>44.1 (13.7)<br>58.8%<br><br><u>Nationality</u>:<br>German: 94.1%<br>Other: 5.9%<br><br>Late August - September 2021 (COVID infected October 1 2020 - April 1 2021)</td><td><u>Comorbidity</u>: NR<br><br><u>SES</u>:<br>University entrance qualification: 51.9%<br><br>Place of birth:<br>Germany: 88.7%<br>Other: 11.3%<br><br><u>Pre-pandemic employment</u>:<br>Full-time: 56.8%<br>Part-time: 27.7%<br>Studying/vocational education: 9.8%<br><br>Current employment:<br>Full-time: 54.4%</td><td>11,710 with COVID 6 to 12 months prior to enrollment<br><br><u>Population</u>: post-COVID with symptoms<br><br><u>Hospitalized</u>: 3.6%<br><br><u>Confirmation method</u>: PCR 100%<br><br><u>Severity</u><br>ICU admission: 0.8%<br><br><u>Vaccination</u>:<br>first dose before positive PCR test: 1.9%</td><td><u>MH description</u>:<br>Symptom clusters 6-12 months after acute infection (not present before acute COVID infection)<br><br><u>MH prevalence</u>:<br>Anxiety/Depression (sleep disorder, depressive mood, anxiety): 21.1%<br><br>Neurocognitive impairment (confusion, memory disturbance, concentration difficulties): 31.3%<br><br><u>Previous MH</u>: 12.8% had pre-existing mental disorders<br><br><u>Ascertainment</u>: Self-report, questionnaires mailed to individuals</td></tr>
</table>

| Author, Year, Country<br>Study design<br>Funding | Mean age (SD) yrs<br>Female, %<br>Race/ethnicity<br>Enrollment dates | Comorbidities<br>SES variables<br>Employment status | COVID population description | MH details |
|---|---|---|---|---|
| | | Part-time: 27.6%<br>Studying/vocational education: 8.8% | | |
| | **Findings**<br>Outcome:<br>Mean percentage loss of current working capacity:<br>1. All patients after COVID infection with anxiety/depression (population attributable loss of working capacity) / All patients after COVID infection<br>2. All patients after COVID infection with anxiety/depression (associated loss of work capacity) / All patients after COVID infection with anxiety/depression<br>3. All patients after COVID infection with neurocognitive impairment (population attributable loss of working capacity) / All patients after COVID infection<br>4. All patients after COVID infection with neurocognitive impairment (associated loss of work capacity) / All patients after COVID infection with neurocognitive impairment<br>Timing: Mean (SD) time since positive PCR test, months: 8.5 (1.6) (range: 6-12 months)<br>Results:<br>1. 0.8% (n=10,324)<br>2. 3.8% (n=2,422)<br>3. 1.86% (n=10,324)<br>4. 5.9% (n=3,561)<br>Adjustments: None | | | |

### Table S4a: Study Characteristics for Mental Health Service Use with Control Groups

| **Author, Year, Country<br>Study design<br>Funding** | **Mean age (SD) yrs<br>Female, %<br>Race/ethnicity, %<br>Enrollment dates** | **Comorbidities<br>SES variables** | **COVID population description** | **MH service description and control group details** |
|---|---|---|---|---|
| Decio, 2022<br>France<br><br>Cohort<br>Industry funded: N<br><br><u>Sample</u>: hospitalized<br><br><u>Risk of bias</u>: High | <u>Overall</u>: n=2894088<br>18–39: 23.1%<br>40–59: 21.2%<br>60–74: 25.9%<br>75+: 29.8%<br><br>Female: 56.1%<br><br><u>Hospitalized for COVID</u>: n=96313 (3.3%)<br>18–39: 8.2%<br>40–59: 21.5%<br>60–74: 27.5%<br>75+: 42.8%<br><br>Female: 47.4%<br><br><u>Hospitalized for other reason</u>: n=2797775 (96.7%)<br>18–39: 23.64%<br>40–59: 21.1%<br>60–74: 25.9%<br>75+: 29.4%<br><br>Female: 56.4%<br><br><u>Race</u>: NR<br><br><u>Enrollment</u>: NR | <u>Comorbidity</u>: NR<br><br><u>SES</u>: Social deprivation index (quintiles)<br><br>*Overall*<br>1 (least deprived): 16.7%<br>2: 19%<br>3: 20.8%<br>4: 21.5%<br>5: (most deprived): 21.9%<br>Unknown: 1.6%<br><br>*Hospitalized for COVID*<br>1 (least deprived): 23.8%<br>2: 17.4%<br>3: 17.2%<br>4: 18%<br>5: (most deprived): 23.7%<br>Unknown: 1.3%<br><br>*Hospitalized for other reason*<br>1 (least deprived): 16.5%<br>2: 19.1%<br>3: 21%<br>4: 21.7%<br>5: (most deprived): 21.8%<br>Unknown: 1.5% | 2,894,088 adult patients with COVID (3.3%) or another medical reason (96.7%)<br><br><u>Population</u>: COVID infected<br><br><u>Hospitalized</u>: 100%<br><br><u>Confirmation method</u>: ICD-10 codes<br><br><u>Severity</u>:<br>*Overall*<br>ICU: 10.4%<br>ICU with invasive procedures: 2.2%<br><br>*Hospitalized for COVID*<br>ICU: 14.1%<br>ICU with invasive procedures: 10.3%<br><br>*Hospitalized for other reason*<br>ICU: 10.3%<br>ICU with invasive procedures: 2%<br><br><u>Variant</u>: study conducted during the first wave of the SARS-CoV-2 pandemic | <u>MH service</u>:<br>Subsequent re-hospitalization (at least once) following discharge for the reference hospital stay for COVID or hospitalization for another reason in the adult general population:<br>• psychiatric disorder of any type<br>• psychotic disorders<br>• mood disorders<br>• anxiety disorders<br>• personality disorders<br><br>*Overall*<br>Psychiatric history: 11%<br><br>*Hospitalized for COVID*<br>Psychiatric history: 13.6%<br><br>*Hospitalized for other reason*<br>Psychiatric history: 11%<br><br><u>Ascertainment</u>:<br>France’s national administrative healthcare database<br><br><u>Control</u>:<br>Adults previously hospitalized for another medical reason other than COVID infection |
| | **Findings**<br><u>Outcome</u>:<br>Subsequent re-hospitalization (at least once) for:<br>1. Psychiatric disorder / Previously hospitalized for COVID vs. Psychiatric disorder / Previously hospitalized for reason other than COVID<br>2. Psychotic disorders / Previously hospitalized for COVID vs. Psychotic disorders / Previously hospitalized for reason other than COVID<br>3. Mood disorders / Previously hospitalized for COVID vs. Mood disorders / Previously hospitalized for reason other than COVID | | | |

| Author, Year, Country<br>Study design<br>Funding | Mean age (SD) yrs<br>Female, %<br>Race/ethnicity, %<br>Enrollment dates | Comorbidities<br>SES variables | COVID population description | MH service description and control group details |
|---|---|---|---|---|
| | 4. Anxiety disorders / Previously hospitalized for COVID vs. Anxiety disorders / Previously hospitalized for reason other than COVID<br>5. Personality disorders / Previously hospitalized for COVID vs. Personality disorders / Previously hospitalized for reason other than COVID<br><u>Timing</u>: During the 12 months following hospital discharge from a medical, surgical, or obstetrics ward<br><u>Results</u>:<br>1. 10,685 / 96,313 = 11.1% vs. 258,566 / 2,797,775 = 9.2%<br>2. 1,019 / 96,313 = 1.1% vs. 26,595 / 2,797,775 = 1%<br>3. 4,115 / 96,313 = 4.27% vs. 106,861 / 2,797,775 = 3.8%<br>4. 5,834 / 96,313 = 6.1% vs. 140,819 / 2,797,775 = 5%<br>5. 766 / 96,313 = 1% vs. 27,182 / 2,797,775 = 1%<br><u>Adjustments</u>: adjusted for socio-demographic characteristics, psychiatric history and characteristics of the reference hospitalization: duration of hospitalization (days) and level of clinical care<br>1. Un-adjusted OR: 1.20 (95% CI: 1.18, 1.23), p<0.0001; Adjusted OR: 0.93 (95% CI: 0.91, 0.95), p<0.0001<br>2. Un-adjusted OR: 1.11 (95% CI: 1.05, 1.19), p<0.0001; Adjusted OR: 1.06 (95% CI: 0.99, 1.14], p=0.09<br>3. Un-adjusted OR: 1.12 (95% CI: 1.09, 1.16], p<0.0001; Adjusted OR: 0.87 (95% CI: 0.84, 0.9), p<0.0001<br>4. Un-adjusted OR: 1.22 (95% CI: 1.18, 1.25], p<0.0001; Adjusted OR: 0.98 (95% CI: 0.95, 1.01), p=0.1536<br>Un-adjusted OR: 0.82(95% CI: 0.76, 0.88], p<0.0001; Adjusted OR: 0.82 (95% CI: 0.76, 0.88), p<0.0001 | | | |
| Formoso, 2023<br>Italy<br><br>Cohort<br>Industry funded: NR<br><br><u>Sample</u>: health database<br><br><u>Risk of bias</u>: High | 43 (NR); range (1-103)<br>51.3%<br><br><u>Race</u>: NR<br><br>September 2020 to May 2021 | <u>Comorbidity</u>:<br>Moderate comorbidity according to Charlson Index (>=2): 5.8%<br><br><u>SES</u>: NR | 36,036 diagnosed with COVID using outpatient service use<br><br><u>Population</u>: COVID infected<br><br><u>Hospitalized</u>: 2%<br><br><u>Confirmation method</u>: PCR 100%<br><br><u>Severity</u>:<br>At least one emergency room access: 9.4%<br><br><u>Vaccination</u>:<br>Received before testing positive: 3% | <u>MH service</u>: outpatient MH specialist visits (e.g., anxiety, depression, sleep problems)<br><br>Total visits: n=190 (n=117 first visit, n=57 >1 visit)<br><br><u>Ascertainment</u>:<br>Administrative database<br><br><u>Control</u>:<br>Matched sample from local health authority database with equal distribution of sex, age, and comorbidities<br>Hospitalized for any reason (at least once): 1.5%<br>Access to emergency room (at least once): 6.9% |
| | **Findings**<br><u>Outcome</u>:<br>1. Those with at least one visit to MH specialist / All patients in database with convalescent COVID vs. general population never infected with COVID | | | |

| Author, Year, Country<br>Study design<br>Funding | Mean age (SD) yrs<br>Female, %<br>Race/ethnicity, %<br>Enrollment dates | Comorbidities<br>SES variables | COVID population description | MH service description and control group details |
|---|---|---|---|---|
| | 2. Total first visits to outpatient MH specialist visit / all patients in database with convalescent COVID vs. general population never infected with COVID<br>Timing: median follow-up of 152 days (range 1–180)<br>Results:<br>1. 174 / 36036 = 0.5% vs. 175 / 36036 = 0.5%<br>2. 117 / 36036 = 0.3% vs. 104 / 36036 = 0.3%<br>Adjustments:<br>1. Hazard ratio (95% CI): 0.87 (0.72–1.07), adjusted for age, sex and occurrence of hospital admissions and/or accesses to emergency room over the 365 days before the index date | | | |
| Staples, 2023<br>Australia<br><br>Cohort<br>Industry funded: N<br><br>Sample: MH outpatient clinic<br><br>Risk of bias: Low/moderate | Recovered COVID (n=7468):<br>33.2 (12.1)<br>76.2%<br><br>Australian, non-Indigenous: 72%<br>Indigenous Australian: 4.3%<br>Born outside Australia: 23.7%<br><br>PCC (n=1873):<br>35.8 (13)<br>77.8%<br><br>Australian, non-Indigenous: 66.6%<br>Indigenous Australian: 5.8%<br>Born outside Australia: 27.6%<br><br>No COVID (n=6151):<br>38 (15.5)<br>71.2%<br><br>Australian, non-Indigenous: 69.2%<br>Indigenous Australian: 4.7% | Comorbidity: NR<br><br>SES<br>Recovered COVID:<br>*Education*<br>Secondary school or below: 32.6%<br>Trade certificate or diploma: 25.7%<br>University degree: 41.6%<br><br>*Income*<br>Employed full or part time: 73.3%<br>Unemployed, no income: 11.9%<br>Pension or benefit: 10.9%<br>Other source of income: 4%<br><br>PCC:<br>*Education*<br>Secondary school or below: 31.9%<br>Trade certificate or diploma: 30.8%<br>University degree: 37.3%<br><br>*Income*<br>Employed full or part time: 63.4%<br>Unemployed, no income: 14%<br>Pension or benefit: 19.4% | 15492 with assessment at MindSpot clinic, a national digital MH service:<br><br>Population:<br>• Recovered at 3 months: 48.2%<br>• PCC (WHO): 12.1%<br>• No COVID: 39.7%<br><br>Hospitalized: NR<br><br>Confirmation method: self-report | MH service:<br>Current MH professional use for those with COVID completing an assessment at national digital MH service<br><br>Ascertainment:<br>Self-report during assessment at MindSpot (digital MH service for symptoms of anxiety or depression)<br><br>Control:<br>General population (without COVID) completing an assessment at national digital MH service |

| Author, Year, Country<br>Study design<br>Funding | Mean age (SD) yrs<br>Female, %<br>Race/ethnicity, %<br>Enrollment dates | Comorbidities<br>SES variables | COVID population description | MH service description and control group details |
|---|---|---|---|---|
| | Born outside Australia: 26%<br><br>September 5, 2022 to May 8, 2023 | Other source of income: 3.2%<br><br><u>No COVID</u>:<br>*Education*<br>Secondary school or below: 35.9%<br>Trade certificate or diploma: 27.2%<br>University degree: 36.9%<br><br>*Income*<br>Employed full or part time: 57%<br>Unemployed, no income: 14.7%<br>Pension or benefit: 21.8%<br>Other source of income: 6.5% | | |
| | **Findings**<br><u>Outcome</u>:<br>Current MH professional use<br>1. With COVID (recovered and PCC) / With COVID (non-symptomatic and symptomatic after 3 months) vs. Without COVID / General population without COVID<br>2. With PCC / With PCC (symptomatic after 3 months) vs. Without COVID / General population without COVID<br><u>Timing</u>: At least 3 months after COVID infection<br><u>Results</u>:<br>1. 1862 / 9341 = 19.9% vs. 1318 / 6151 = 21.4%<br>2. 443 / 1873 = 23.7% vs. 1318 / 6151 = 21.4%<br><u>Adjustments</u>: None | | | |

### Table S4b: Study Characteristics for Mental Health Service Use without Control Groups

<table>
<tr><th>Author, Year, Country<br>Study design<br>Funding</th><th>Mean age (SD) yrs<br>Female, %<br>Race/ethnicity, %<br>Enrollment dates</th><th>Comorbidities<br>SES variables</th><th>COVID population description</th><th>MH service description</th></tr>
<tr><td rowspan="2">Ahmed, 2022<br>USA<br><br>Cross-sectional<br>Industry funded: N<br><br>Sample: post COVID clinic<br><br>Risk of bias:<br>High</td><td>53 (IQR: 21-89)<br>66.7%<br><br>Race:<br>Caucasian: 40.4%<br>Hispanic: 31.6%<br>Black: 21.1%<br><br>June 1, 2020 to December 1, 2020</td><td>Comorbidity: NR<br><br>SES: NR</td><td>1511 COVID positive patients<br>114/1511 (7.5%) attended post-COVID clinic for initial evaluation of long-term sequelae, early rehabilitation, and targeted specialist referrals<br><br>Population: COVID infected<br><br>Hospitalized: 87.7%<br><br>Confirmation method: NR<br><br>Severity:<br>ICU admission: 7.9%</td><td>MH service:<br>Psychiatry service use at the post-COVID clinic when presenting with depressed mood, hopelessness, anxiety, and/or fatigue<br><br>Ascertainment: Medical record</td></tr>
<tr><td colspan="4">Findings<br>Outcome:<br>Psychiatry service use<br>1. For those attending post-COVID clinic / All COVID positive patients<br>2. For those attending post-COVID clinic / All COVID positive patients referred to post COVID clinic for baseline evaluation<br>Timing: Over the initial 6 months follow-up, with any re-evaluations occurring around 1 month after initial evaluation<br>Results:<br>1. 22/1511 = 1.5%<br>2. 22/114 = 19.3%<br>Adjustments: None</td></tr>
<tr><td>Bek, 2023<br>The Netherlands<br><br>Cross-sectional<br><br>Industry funded: N<br><br>Sample: outpatient post-COVID clinic<br><br>Risk of bias:<br>Low/Moderate</td><td>59.7 (11.4)<br>31%<br><br>Race:<br>European: 73%<br>North African: 4%<br>Dutch Caribbean: 14%<br>Asian: 6%<br>Turkish: 4%<br><br>July 1, 2020 to June 13, 2022</td><td>Comorbidity:<br>≥1: 81%<br><br>Pre-COVID educational level:<br>Low [primary or secondary]: 35%<br>Middle [high school]: 35%<br>High [post-secondary or university]: 30%<br><br>Pre-COVID employment:<br>Unemployed: 15%</td><td>617 patients attending the COVID follow-up care path (CO-FLOW) after hospital discharge<br><br>Population: COVID infected<br><br>Hospitalized: 100%<br><br>Confirmation method: PCR or multidisciplinary team decision based on symptoms: 100%<br><br>Severity</td><td>MH service:<br>Attended psychologist session for anxiety or depression<br><br>Previous MH: 4.7% had mental disorder at baseline<br><br>Ascertainment:<br>Electronic data capture system</td></tr>
</table>

| **Author, Year, Country**<br>**Study design**<br>**Funding** | **Mean age (SD) yrs**<br>**Female, %**<br>**Race/ethnicity, %**<br>**Enrollment dates** | **Comorbidities**<br>**SES variables** | **COVID population description** | **MH service description** |
|---|---|---|---|---|
| | | Employed: 60%<br>Retirement: 25% | ICU admission: 41%<br>Oxygen supplementation: 97%<br>High flow nasal cannula: 33%<br>Invasive mechanical ventilation: 35% | |
| | Outcome: Consulted a psychologist / All patient after COVID infection<br>Timing: 3 months from hospital discharge for acute COVID infection<br>Results: 71/617 = 11.5%<br>Adjustments: None | | | |
| Benzakour, 2021<br>Switzerland<br><br>Cross-sectional<br>Industry funded: NR<br><br>Sample: outpatient clinic after hospitalization<br><br>Risk of bias: High | 56.8 (range: 18-86)<br>38.5%<br><br>Race: NR<br><br>March 30, 2020 to July 1, 2020 | Comorbidity: NR<br><br>SES: NR | 109 hospitalized for COVID and attending CoviCare program (a multidisciplinary strategy, including specialists in internal medicine, pulmonary disease, primary care, infectious diseases and psychiatry)<br><br>Population: COVID infected<br><br>Hospitalized: 100%<br><br>Confirmation method: NR<br><br>Severity:<br>ICU: 16.5% | MH service:<br>Psychiatric follow-up: the psychiatrist provided an intervention depending on patients' mental status at assessment. Patients whose questionnaire scores exceeded the established cut-off (PDEQ>15 or/and HADS-A>11 or/and HADS-D>11 or/and PCL5>31), were referred to a psychiatric consultation by the liaison psychiatry team, consisting of an adapted psychiatric and/or psychotherapeutic intervention<br><br>Ascertainment:<br>Questionnaire sent out by e-mail or postal mail, and phone call to follow-up by CoviCare team |
| | **Findings**<br>Outcome:<br>1. Patients taking psychiatric follow-up/ All COVID patients after hospitalization<br>2. Patients taking psychiatric follow-up/ All COVID patients attending CoviCare service after hospitalization<br><br>Timing: One month following hospitalization for COVID pneumonia<br>Results:<br>1. 26/109 = 23.9%<br>2. 26/64 = 40.6%<br>Adjustments: None | | | |
| Bonazza, 2020<br>Italy | 59 (13.3)<br>31.8% | Comorbidity: NR<br><br>SES: NR | 261 recovered COVID patients previously hospitalized and offered a multidisciplinary follow- | MH service:<br>Psychological service use (interview) provided by the hospital psychologist |

| Author, Year, Country<br>Study design<br>Funding | Mean age (SD) yrs<br>Female, %<br>Race/ethnicity, %<br>Enrollment dates | Comorbidities<br>SES variables | COVID population description | MH service description |
|---|---|---|---|---|
| Cross-sectional<br>Industry funded: NR<br><br>Sample: outpatient program<br><br>Risk of bias: Low/moderate | Race: NR<br><br>April 2020 to July 2020 (data collection) | | up screening program within 2 months<br><br>Population: COVID infected<br><br>Hospitalized: 100%<br><br>Confirmation method: NR<br><br>Severity:<br>Sub-ICU treatments: 27.7%<br>ICU treatment: 7.5% | when Hospital Anxiety and Depression scale (HADs) score was >11 for abnormal severe anxiety, and the Impact of Events Scale-Revised (IES-R) was >33 for possible ongoing acute stress disorder or PTSD<br><br>Psychologist service use, 8 free sessions after psychological interview by psychotherapists if the cut-off score for HADs and IES-R is above normal<br><br>Ascertainment: Face to face data collection interview and needed referral for psychologist |
| | **Findings**<br>Outcome:<br>1. Participants using psychological service (interview) provided by the hospital psychologist/ All COVID patients previously hospitalized<br>2. Participants using psychologist service (after psychological interview) by psychotherapists/ All COVID patients previously hospitalized<br>Timing: Within 2 months of discharge of hospital<br>Results:<br>1. 36/261 = 13.8%<br>2. 16/261 = 6.1%<br>Adjustments: None | | | |
| Brehon, 2022<br>Canada<br><br>Cross-sectional<br>Industry funded: N<br><br>Sample: post-COVID rehabilitation clinic<br><br>Risk of bias: High | 48.9 (10.5)<br>64%<br><br>Race: NR<br><br>March 2020 to mid-May 2021 | Comorbidity: NR<br><br>SES:<br>Employed at admission: 95%<br>Health-related occupations: 53 %<br>Employed at discharge: 94% | 81 workers discharged from post-COVID occupational rehabilitation program<br><br>Population: post-COVID with symptoms<br><br>Hospitalized: 17%<br><br>Confirmation method: NR | MH service:<br>Psychology services received prior to beginning the post-COVID rehabilitation program<br><br>Ascertainment:<br>Provincial databases managed by WCB-Alberta Health Care Strategy |
| | **Findings**<br>Outcome:<br>Those receiving psychology services / All workers participating in the occupational post-COVID rehabilitation program | | | |

| **Author, Year,<br>Country<br>Study design<br>Funding** | **Mean age (SD) yrs<br>Female, %<br>Race/ethnicity, %<br>Enrollment dates** | **Comorbidities<br>SES variables** | **COVID population description** | **MH service description** |
|---|---|---|---|---|
| | Timing: Mean (SD) days between symptom onset and program admission: 165.2 (73.0)<br>Results: 26 / 81 = 32%<br>Adjustments: None | | | |
| Chopra, 2021<br>USA<br><br>Cross-sectional<br>Industry funded: N<br><br>Sample:<br>hospitalized<br><br>Risk of bias:<br>High | Median (IQR): 62 (50-72)<br>48.2%<br><br>Race:<br>Black: 51.6%<br>White: 37.3%<br>Other/unknown: 11.1%<br><br>Ethnicity:<br>Hispanic: 4.4%<br>Non-Hispanic: 86.7%<br><br>March 16, 2020 to July 1, 2020 | Comorbidity: NR<br><br>SES:<br>Residence before hospitalization:<br>Home: 82.8%<br>Congregated living facility: 15.2%<br>Subacute rehabilitation facility: 0.6%<br>Other/unknown: 0.4% | 1250 hospitalized with COVID<br><br>Population: COVID infected<br><br>Hospitalized: 100%<br><br>Confirmation method: NR<br><br>Severity:<br>Treated in an ICU: 13.2%<br>Treated with invasive mechanical ventilation: 5.9%<br>Treated with supplemental oxygen: 69.2%<br>Discharged to a health care facility: 12.6% | MH service: Health care use related to MH<br><br>Ascertainment: Medical record and phone call to participants |
| | **Findings**<br>Outcome: Health care use related to MH<br>1. Among those surviving COVID hospitalization<br>2. Among those surviving COVID hospitalization and completing telephone survey<br>Timing: 60-day post discharge for outcome<br>Results:<br>1. 28 / 1250 = 2.2%<br>2. 28 / 488 = 5.7%<br>Adjustments: None | | | |
| Chow, 2023<br>USA<br><br>Cross-sectional<br>Industry funded: NR<br><br>Sample: outpatient clinic<br><br>Risk of bias:<br>High | 48.1 (12.9)<br>72%<br><br>Race:<br>Hispanic: 47%<br>White: 36%<br>Black: 8%<br>Asian: 4%<br>Other: 4%<br><br>March 2020 - April 18, 2022 | Comorbidity:<br>NR<br><br>SES: NR | 156 patients with PCC attending specialty outpatient clinic for recovery<br><br>Population: PCC (self-reported symptoms) 100%<br><br>Hospitalized: 20%<br><br>Confirmation method: NR<br><br>Severity: | MH service: Referral to psychiatrists<br><br>Previous MH:<br>Depression: 26%<br>Anxiety: 29%<br><br>Ascertainment: Electronic medical record |

| Author, Year, Country<br>Study design<br>Funding | Mean age (SD) yrs<br>Female, %<br>Race/ethnicity, %<br>Enrollment dates | Comorbidities<br>SES variables | COVID population description | MH service description |
|---|---|---|---|---|
| | | | Symptoms with no hospitalization: 80%<br>Symptoms hospitalized: 20% | |
| | Outcome: Referral to psychiatrists with new psychiatric symptoms after COVID / All patients see with PCC<br>Timing: NR (all with PCC)<br>Results: 44 / 156 = 28.2%<br>Adjustments: None | | | |
| Diem, 2022<br>Switzerland<br><br>Cross-sectional<br>Industry funded: N<br><br>Sample:<br>community and social media<br><br>Risk of bias:<br>High | 44.6 (NR); range 19-83<br>80.6%<br><br>Nationality:<br>Swiss: 95.1%<br>German: 1.9%<br>French: 1.9%<br>Belgian: 0.6%<br>Austrian: 0.3%<br><br>October 15, 2021 to December 12, 2021 | Comorbidity:<br>Yes: 36.2%<br><br>SES:<br>Education ≥13 years: 59.5% | 309 with PCC and persistent symptom over 3 months<br><br>Population: PCC (WHO) 100%<br><br>Hospitalized: 10.7%<br><br>Confirmation method: PCR, antigen: 82.8%<br><br>Severity:<br>Intubation: 2%<br>ICU without intubation: 2.6%<br>Oxygen requirement: 9.4%<br><br>Vaccination:<br>Yes: 82.9% (n=232)<br>Spikevax (Moderna): 50%<br>Comirnaty (Pfizer/Biotech): 49.1%<br>Janssen (Johnson & Johnson): 0.9% | MH service:<br>Consulted a psychiatrist/psychologist<br><br>Ascertainment: Self-report survey |
| | **Findings**<br>Outcome:<br>Consulted a psychiatrist or psychologist / all patients included in survey with PCC<br>Timing: survey completed after acute infection; mean = 13 months, range 0.2 - 24.5 months<br>Results:<br>113 / 309 = 36.5%<br>Adjustments: None | | | |
| Farooqi, 2022<br>USA<br><br>Cross-sectional<br>Industry funded: N | 51.3 (12.6)<br>87%<br><br>Race:<br>Caucasian: 53% | Comorbidity:<br>Medical comorbidities; mean (SD) = 1.13 (0.96)<br><br>SES: NR | 30 patients complaining of neuropsychiatric symptoms referred and assessed for psychiatric complications of COVID to outpatient department | MH service:<br>Recommendation for group or individual psychotherapeutic treatment |

| Author, Year, Country<br>Study design<br>Funding | Mean age (SD) yrs<br>Female, %<br>Race/ethnicity, %<br>Enrollment dates | Comorbidities<br>SES variables | COVID population description | MH service description |
|---|---|---|---|---|
| <br>Sample: outpatient department<br><br>Risk of bias:<br>High | Black: 13%<br>Hispanic: 13%<br>Asian: 3%<br>Other: 20%<br><br>December 1, 2020 to July 26, 2021 | <br>Employment:<br>Currently employed: 23%<br>Not employed/retired before pandemic: 17%<br>Significant interruption: 60% | of an academic Behavioral Health Center<br><br>Population: PCC (symptom evaluation) 100%<br><br>Hospitalized: 36.7%<br><br>Confirmation method: NR<br><br>Severity: Moderate illness constituting pulmonary symptoms, short-term hospitalization, or significant symptoms while at home: 33.3%<br>Significant illness requiring intubation for hypoxia and prolonged hospitalization: 3.3%<br><br>COVID diagnosis (variant):<br>February-March 2020: 30%<br>January-February 2021: 30%<br>January 2020-March 2021: 40% | Ascertainment:<br>Treatments recommended by Post COVID Recovery Program |
| | **Findings**<br>Outcome: recommended psychotherapeutic treatment for all COVID patients with MH symptoms previously treated at a post-COVID recovery program and referred to an outpatient's psychiatric department<br>Timing: Mean (SD) months from COVID diagnosis to timing of consult: 7.23 (4.89)<br>Results: 4 / 30 = 13.3%<br>Adjustments: None | | | |
| Frontera, 2022<br>USA<br><br>Cross-sectional<br>Industry funded: NR<br><br>Sample: hospitalized<br><br>Risk of bias: | Overall: (n=242)<br>65 (NR); IQR: 53-73<br>36%<br>Race: NR<br><br>PCC: (n=122/242)<br>64 (NR); IQR: 55-71<br>38%<br>Race: White: 50% | Comorbidity:<br>122 (50%) reported >=1 PCC symptom (median 3, IQR 1–5) lasting a median of 12-months (range 1–15) post-COVID diagnosis; according to CDC criteria as new or persistent symptoms occurring >=4 weeks after COVID infection | 242 hospitalized with COVID<br><br>Population: PCC (CDC) 50%<br><br>Hospitalized: 100%<br><br>Confirmation method: PCR 100%<br><br>Severity:<br>Invasive mechanical ventilation: 34% | MH service:<br>Psychological talk therapy to deal with prolonged COVID symptoms<br><br>Ascertainment: Self-report survey |

| Author, Year, Country<br>Study design<br>Funding | Mean age (SD) yrs<br>Female, %<br>Race/ethnicity, %<br>Enrollment dates | Comorbidities<br>SES variables | COVID population description | MH service description |
|---|---|---|---|---|
| Low/moderate | March 10, 2020 to May 20, 2020 (hospitalized between) | SES:<br>With PCC, Education level >12 years: 73.8% | Neurological complication during hospitalization: 47%<br>With PCC and mechanically ventilated: 47%<br><br>Vaccination:<br>Those with PASC: 67.2% | |
| | **Findings**<br>Outcome:<br>Use of psychological talk therapy<br>1. All with COVID / Total sample with COVID<br>2. All with COVID / Total sample with PCC<br>Timing: Follow-up interviews performed at 12-months (+/- 2 months) after initial COVID diagnosis (January 10 - July 20, 2021)<br>Results:<br>1. 11 / 242 = 4.5%<br>2. 11 / 122 = 9%<br>Adjustments: None | | | |
| Gramaglia, 2021<br>Italy<br><br>Cross-sectional<br>Industry funded: N<br><br>Sample:<br>hospitalized<br><br>Risk of bias:<br>Low/moderate | 61 (IQR: 50-71)<br>40.3%<br><br>March 1 to June 29, 2020 (discharged from hospital) | Comorbidity:<br>Total median (IQR): 2 (1-3)<br><br>SES: NR | 238 participants (or their caregivers) aged 18 years or older with COVID<br><br>Population: COVID infected<br>Hospitalized: 100%<br>Confirmation method: PCR 97.3%<br>Severity:<br>Oxygen via nasal cannula or Venturi masks: 42.9%<br>Non-invasive ventilation: 20.6%<br>Mechanical ventilation: 8.8%<br>ICU: 11.8% | MH service:<br>Referred to further psychiatric consultation provided by psychiatrist which was determined during face-to-face interview by an experienced psychiatrist<br><br>Ascertainment:<br>Face-to-face interview by an experienced psychiatrist and self-reported outcome during hospital stay and follow-up visits |
| | **Findings**<br>Outcome: Participants referred to further psychiatric consultation / All COVID patients previously hospitalized<br>Timing: 4 months, median time after discharge 131 (119–145) days<br>Results: 26/237 = 10.97%<br>Adjustments: None | | | |
| Heeney, 2023<br>Ireland<br><br>Cross-sectional<br>Industry funded: N | Median (IQR): 43 (31-53)<br>66%<br><br>Race:<br>NR | Comorbidity: At initial post-COVID clinic visit 221 of 311 patients (71%) had ongoing symptoms | 311 patients at first visit to outpatient post-COVID clinic after their acute illness and to screen for potential sequelae of COVID infection | MH service:<br>Patients referred to clinic psychology service and onward referrals to psychology. Onward referrals |

<table>
<tr><th>Author, Year, Country<br>Study design<br>Funding</th><th>Mean age (SD) yrs<br>Female, %<br>Race/ethnicity, %<br>Enrollment dates</th><th>Comorbidities<br>SES variables</th><th>COVID population description</th><th>MH service description</th></tr>
<tr><td rowspan="2"><u>Sample</u>: post-COVID clinic<br><br><u>Risk of bias</u>:<br>High</td><td>March 10 to June 14, 2020</td><td><u>SES</u>:<br>Healthcare staff: 62%</td><td><u>Population</u>: post-COVID with symptoms 71%<br><br><u>Hospitalized</u>: 44%<br><br><u>Confirmation method</u>: PCR 100%<br><br><u>Severity</u>:<br>Required admission to ICU or high dependency unit: 12%<br>Required supplemental oxygen therapy: 50%<br>Required invasive ventilation: 6.5%<br><br><u>Variant</u>:<br>Based on timing of recruitment we infected exclusively with the original Wuhan variant of COVID</td><td>required further follow-up beyond first clinic visit.<br><br><u>Ascertainment</u>:<br>In-person clinic visits with a member of the infectious diseases medical team</td></tr>
<tr><td colspan="4">Findings<br><u>Outcome</u>:<br>1. Patient referral to clinic psychology service / All patients attending outpatient PCC clinic<br>2. Patients with onward referrals to psychology / All patients with further follow-up arranged (not necessarily being referred) after attending outpatient post COVID clinic<br>3. Patients with onward referrals to psychology / All patients requiring onward referral to one of following: respiratory, cardiology, physio, psychology, neurology, or other after attending outpatient post COVID clinic<br>4. Patients with onward referrals to psychology / All patients attending outpatient post COVID clinic<br><u>Timing</u>: Median (IQR) days from positive PCR test to first clinic appointment: 95 (77–105.5)<br><u>Results</u>:<br>1. 7 / 311 = 2.3%<br>2. 13 / 62 = 21%<br>3. 13 / 56 = 23.2%<br>4. 13 / 311 = 4.2%<br><u>Adjustments</u>: None</td></tr>
<tr><td>Hodgson, 2021<br>Australia<br><br>Cross-sectional<br>Industry funded: N</td><td><u>Hospital cohort</u>: n=212<br>61 (NR) range 51-70<br>41.5%<br><br><u>6-mos follow-up</u>: n=160</td><td><u>Comorbidity</u>: NR<br><br><u>SES</u>:<br>Hospital cohort:</td><td>212 COVID patients admitted to ICU, 160/212 (75.5%) followed-up at 6 mos, and 115 alive at 6 mos and reporting persistent</td><td><u>MH service</u>:<br>Treatment for anxiety/depression at 6-month follow-up<br><br><u>Ascertainment</u>:</td></tr>
</table>

| Author, Year, Country<br>Study design<br>Funding | Mean age (SD) yrs<br>Female, %<br>Race/ethnicity, %<br>Enrollment dates | Comorbidities<br>SES variables | COVID population description | MH service description |
|---|---|---|---|---|
| <br>Sample: ICU admitted<br><br>Risk of bias: High | 62 (NR) range 55-71<br>39.4%<br><br>Alive and interviews at 6-mos: n=115<br>58 (NR) range 51-69<br>42.6%<br><br>Race: NR<br><br>March 06, 2020 to October 04, 2020 | Mean (range) years of education: 14 (11-16)<br>Healthcare worker: 11.7%<br><br>Hospital readmission:<br>Mean (range) years of education: 14 (11-16)<br>Healthcare worker: 9.8%<br><br>Alive and interviews at 6-mos:<br>Mean (range) years of education: 14 (11-16)<br>Healthcare worker: 13% | symptoms. 50/115 (43.5%) reporting three or more symptoms<br><br>Population:<br>COVID infected: 100%<br>Post-COVID with symptoms: 43.5%<br><br>Hospitalized: 100%<br><br>Confirmation method: PCR 100%<br><br>Severity:<br>Hospital cohort:<br>ICU mortality: 18.4%<br><br>6-mos follow-up: n=160<br>Hospital readmission: 16.7%<br>ICU mortality: 24.4%<br>Death or new disability at 6 months: 56.3%<br><br>Alive and interviews at 6-mos:<br>Hospital readmission: 17% | Surviving patients contacted prospectively by mail and invited to in telephone interviews at 6 mo after ICU admission<br><br>Anxiety in follow-up cohort at 6-months: 4.3% |
| | **Findings**<br>Outcome:<br>Treatment for anxiety/depression at 6-month follow-up<br>1. Among all assessed at 6 mo follow-up<br>2. Among all in ICU at baseline<br>Timing: Final 6-month follow-up was conducted on April 21, 2021<br>Median days (IQR) between symptoms - hospital admission: 6.1 (3.6-9.2)<br>Median days (IQR) between symptoms - ICU admission: 8.1 (5.3-11)<br>Results:<br>1. 16 / 109 = 14.7%<br>2. 16 / 212 = 7.5%<br>Adjustments: None | | | |
| Holland, 2023<br>Australia<br><br>Cross-sectional | 53 (NR), range (18-78)<br>65% | Comorbidity: NR<br><br>SES: NR | 726 people with COVID completing 8 weeks follow-up questionnaire | MH service:<br>Referral to neuropsychology<br><br>Ascertainment: |

| **Author, Year, Country<br>Study design<br>Funding** | **Mean age (SD) yrs<br>Female, %<br>Race/ethnicity, %<br>Enrollment dates** | **Comorbidities<br>SES variables** | **COVID population description** | **MH service description** |
|---|---|---|---|---|
| Industry funded: NR<br><br><u>Sample</u>: inpatients and those managed as home-based acute medical service<br><br><u>Risk of bias</u>: High | March 2020 to December 2022 | | <u>Population</u>: COVID infected<br><br><u>Hospitalized</u>: 90.1%<br><br><u>Confirmation method</u>: NR<br><br><u>Severity</u>: ICU: 2.9%<br><br><u>Reinfection</u>: In 2022: 1% | Online questionnaire with free text answers |
| | **Findings**<br><u>Outcome:</u><br>Participants who got a referral to neuropsychology/ COVID patients followed up at the Alfred Health Post-COVID service<br><u>Timing:</u> 8 weeks after the onset of COVID<br><u>Results:</u><br>92/726 = 12.7%<br><u>Adjustments:</u> None | | | |
| Huang, 2022<br>China<br><br>Cross-sectional<br>Industry funded: N<br><br><u>Sample</u>: hospitalized<br><br><u>Risk of bias</u>: Low/moderate | 57 (IQR: 48-65)<br>46%<br><br><u>Race</u>: NR<br><br>January 2020 to May 2020 | <u>Comorbidity</u>: NR<br><br><u>SES</u>:<br>Education:<br>College or higher: 28%<br>High school or lower: 72%<br><br>Work status before COVID:<br>Retired: 55%<br>Full-time or part-time job: 42%<br>Jobless: 4%<br>Homemaker: 0% | 1192 COVID survivors who were previously hospitalized and completed 3 follow-up visits<br><br><u>Population</u>: COVID infected<br>At 2 year follow up 650/1190 (54.6%) had PCC symptoms defined as at least one sequelae symptom<br><br><u>Hospitalized</u>: 100%<br><br><u>Confirmation method</u>: laboratory confirmed 100%<br><br><u>Severity</u>:<br>• ICU admission: 4%<br>• Hospitalization, requiring supplemental oxygen: 68%<br>• Hospitalization, requiring HFNC or non-IMV, or both: 7%<br>• Hospitalization, requiring ECMO or IMV, or both: 1% | <u>MH service</u>:<br>Use of psychological help as health-care use after discharge<br><br><u>Ascertainment</u>:<br>Self-reported questionnaire |
| | **Findings** | | | |

| Author, Year, Country<br>Study design<br>Funding | Mean age (SD) yrs<br>Female, %<br>Race/ethnicity, %<br>Enrollment dates | Comorbidities<br>SES variables | COVID population description | MH service description |
|---|---|---|---|---|
| | Outcome: Use of psychological help as health-care after discharge/ COVID survivors completed assessments for healthcare use<br>Timing: From symptom onset to 2-year follow-up, median days (IQR): 685 (675-698)<br>Results: 3/1187 = 0.25%<br>Adjustments: None | | | |
| Kalyani, 2022<br>India<br><br>Cross-sectional<br>Industry funded: NR<br><br>Sample: hospitalized<br><br>Risk of bias: High | 55.2 (6.1)<br>29.1%<br><br>Race: NR<br><br>June 2020 to November 2020 | Comorbidity: NR<br><br>Going to work:<br>Returned to work: 64%<br>Not returned to work: 36% | 86 of 2654 (3.2%) COVID infected reporting back with persistent symptoms (various complaints related to medical problems and ENT) at 3 months<br><br>Population: COVID infected (n=2654), post-COVID with symptoms (n=86/2654)<br><br>Hospitalized: 100%<br><br>Confirmation method: PCR 100%<br><br>Severity:<br>Admitted to ICU: 22.1% | MH service:<br>Previously hospitalized patients being readmitted to hospital with complaint of confusion<br><br>Ascertainment: Hospital records |
| | **Findings**<br>Outcome:<br>1. Patients readmitted for confusion / all patients who reporting back with any persistent symptoms<br>2. Patients readmitted for confusion / all patients admitted to hospital during same period for treatment of COVID<br>Timing: PCC symptoms developed 3 months after hospital discharge<br>Results:<br>1. 34 / 86 = 39.5%<br>2. 34 / 2654 = 1.3%<br>Adjustments: None | | | |
| Kisiel, 2023<br>Sweden<br><br>Cross-sectional<br>Industry funded: N<br><br>Sample: outpatient care<br><br>Risk of bias: Low/moderate | 43.5 (NR)<br>73.4%<br><br>Race: NR<br><br>March 10, 2020 - December 30, 2020 | Comorbidity:<br>0: 50.6%<br>1: 30.3%<br>≥2: 19.1%<br><br>Education level:<br>University: 55.9%<br>No university: 44.1%<br><br>Place of birth: | 359 symptomatic non-hospitalized healthcare workers and other occupational groups, receiving outpatient follow-up care<br><br>Population: post-COVID with symptoms<br><br>Hospitalized: 0%<br><br>Confirmation method: PCR 100% | MH service:<br>Newly introduced treatment for depression/anxiety after COVID (including psychological treatment or medication)<br><br>Ascertainment:<br>Survey administered within 45 to 51 weeks following diagnosis |

| Author, Year, Country<br>Study design<br>Funding | Mean age (SD) yrs<br>Female, %<br>Race/ethnicity, %<br>Enrollment dates | Comorbidities<br>SES variables | COVID population description | MH service description |
|---|---|---|---|---|
| | | Sweden and both parents: 70.2%<br>Sweden and one parent: 7.8%<br>Sweden, not parents: 2.6%<br>Not in Sweden: 19.4% | Self-reported severity:<br>Mild: 42%<br>Moderate: 39.1%<br>Severe: 18.9% | |
| | **Findings**<br>Outcome: Newly introduced treatment for depression or anxiety after COVID / all patients with COVID<br>Timing: 12-month follow-up after COVID<br>Results: 17/359 = 4.7%<br>Adjustments: None | | | |
| Lazzaroni, 2022<br>Italy<br><br>Cross-sectional<br>Industry funded: NR<br><br>Sample: outpatient clinic<br><br>Risk of bias: Low/moderate | All patients: n=523<br>60 (IQR 55-74)<br>34.2%<br><br>Those with MH symptoms: n=93<br>65 (IQR 55-75)<br>36.5%<br><br>Patient with 4-6 visits with psychologist: n=58<br>61 (IQR 55-69)<br>47.5%<br><br>Race: NR<br><br>March 2020 - November 2021 | Comorbidity: NR<br><br>SES: NR | 523 patients previously hospitalized with COVID, attending a COVID outpatient clinic after hospital discharge<br><br>Population: COVID infected<br><br>Hospitalized: 100%<br><br>Confirmation method: PCR 100%<br><br>All patients:<br>CPAP or ICU: 23.5%<br>Other oxygen treatment or none: 76.5%<br><br>Those with MH symptoms:<br>CPAP or ICU: 32%<br>Other oxygen treatment or none: 68%<br><br>Patient with 4-6 visits with psychologist:<br>CPAP or ICU: 51.8%<br>Other oxygen treatment or none: 48.2% | MH service:<br>Psychologist consultation; 4-6 visits among patients after COVID (previously hospitalized) that screened positive for PTSD or depression<br><br>Trauma-focused psychological support intervention was proposed to patients with positive scores aimed to facilitating the resumption of the daily life by reducing the impact of this recent traumatic experience<br><br>Ascertainment:<br>Electronic data capture system |
| | **Findings**<br>Outcome:<br>1. Patients agreeing to undergo psychologist consultation / All patients after COVID infection | | | |

| Author, Year, Country<br>Study design<br>Funding | Mean age (SD) yrs<br>Female, %<br>Race/ethnicity, %<br>Enrollment dates | Comorbidities<br>SES variables | COVID population description | MH service description |
|---|---|---|---|---|
| | 2. Patients being referred for psychologist consultation / All patients after COVID infection<br>3. Patients agreeing to undergo psychologist consultation / Patients being referred for psychologist consultation<br>Timing: NR (post-hospitalization, during outpatient COVID clinic visit)<br>Results:<br>1. 58 / 523 = 11.1%<br>2. 93 / 523 = 17.8%<br>3. 58 / 93 = 62.3%<br>Adjustments: None | | | |
| LeGoff, 2023<br>USA<br><br>Cross-sectional<br>Industry funded: NR<br><br>Sample: outpatient MH clinic<br><br>Risk of bias: High | 48.3 (NR)<br>54.7%<br><br>Race: NR<br><br>January 2021 to December 2022 (referral cases)<br><br>April 2020 to February 2022 (contracted COVID) | Comorbidity: NR<br><br>SES: NR<br><br>Employment:<br>Occupational Category:<br>Healthcare services: 50%<br>Corrections: 21.9%<br>First responders: 14.1%<br>Facilities maintenance: 14.1% | 67 California employees referred by occupational physician's outpatient MH provider panel for assessment and treatment due to diagnosis of PCC with neurocognitive complaints of "brain fog," including mental fatigue, forgetfulness/impaired short-term memory, difficulties concentrating, inattentiveness, diminished task focus, confusion, etc.<br><br>Population: PCC (neurocognitive screening) 100%<br><br>Hospitalized: NR<br><br>Confirmation method: NR | MH service:<br>Recommended (referred) for MH treatment after assessment by neurocognitive screening evaluation<br><br>Attended MH treatment after assessment by neurocognitive screening evaluation with an average number of therapy session of 2.4 (range 0-42)<br><br>MH Prevalence: 100%, All with neurocognitive complaints<br><br>Ascertainment:<br>Self-reported neurocognitive complaints after PCC diagnosis |
| | **Findings**<br>Outcome:<br>1. Referred to MH treatment after neurocognitive screening evaluation / Assessed by neurocognitive evaluation with MH symptoms<br>2. Attended MH treatment after assessment by neurocognitive screening evaluation / Assessed by neurocognitive evaluation with MH symptoms<br>Timing: The mean duration of illness before the referral for MH services was 11 months (341.5 days; SD, 178.3 days), with a range of 9 weeks (67.7 days) to 25 months (751.0 days), and a median of 315.5 days<br>Results:<br>1. 29/64 = 45.3%<br>2. 9/64 = 14.1%<br>Adjustments: None | | | |
| Lynch, 2024<br>USA | 43.5 (15)<br>70.7 | Comorbidity: | 75 undergoing neuropsychological, psychiatric | MH Service: |

| Author, Year, Country<br>Study design<br>Funding | Mean age (SD) yrs<br>Female, %<br>Race/ethnicity, %<br>Enrollment dates | Comorbidities<br>SES variables | COVID population description | MH service description |
|---|---|---|---|---|
| Cross-sectional<br>Industry funded: N<br><br>Sample: mix (outpatient & health practitioner)<br><br>Risk of bias: High | Race:<br>Black: 8.0%<br>Asian/South Asian: 8.0%<br>White: 64%<br>Hispanic/Latino: 17.3%<br>Other/Unknown: 2.7%<br><br>Enrollment: NR | Mean (SD) number: 1.5 (1.5)<br><br>SES:<br>Mean (SD) years of education: 16.0 (2.2) | and medical assessments. More than half referred from outpatient post-COVID recovery program offering treatment for various medical symptoms of PCC<br><br>Population: mixed; not specified<br><br>Hospitalized: 12%<br><br>Confirmation Method: 100% naso, antibody<br><br>Severity: acute COVID symptom severity was mostly moderate (score 11-20 on 0-33 scale) with an average length of acute illness 3 weeks<br>Peak COVID symptom score, mean (SD): 16.6 (6.2)<br>Required ICU or ventilator support: 0%<br><br>Vaccination:<br>At least 1 dose at study assessment: 42.3%<br>At least 1 dose by second assessment: 88.9% | Participants hospitalized psychiatrically, received neurocognitive rehabilitation, started seeing a psychiatrist or started psychotherapy<br><br>Based on interval history between initial visit and follow-up assessments after 6 months. At enrollment, participants excluded if currently unstable psychiatric symptoms, such as active symptoms of psychosis or active suicidal ideation. Patients currently engaged in psychiatric care were not excluded.<br><br>Ascertainment:<br>Self-reported; psychiatric and medical information collected by study assessors during in-person assessments |
| | **Findings**<br>Outcome:<br>Hospitalized psychiatrically<br>1. Among those with low (≥1) or extremely low score (≥2) on neuropsychological assessment at initial visit;<br>2. Among all undergoing neuropsychological assessment at initial visit<br>Neurocognitive rehabilitation<br>3. Among those with low (≥1) or extremely low score (≥2) on neuropsychological assessment at initial visit;<br>4. Among all undergoing neuropsychological assessment at initial visit<br>Started seeing a psychiatrist<br>5. Among those with low (≥1) or extremely low score (≥2) on neuropsychological assessment at initial visit;<br>6. Among all undergoing neuropsychological assessment at initial visit<br>Started psychotherapy<br>7. Among those with low (≥1) or extremely low score (≥2) on neuropsychological assessment at initial visit; | | | |

| Author, Year, Country<br>Study design<br>Funding | Mean age (SD) yrs<br>Female, %<br>Race/ethnicity, %<br>Enrollment dates | Comorbidities<br>SES variables | COVID population description | MH service description |
|---|---|---|---|---|
| | 8. Among all undergoing neuropsychological assessment at initial visit<br>Timing: On average 208 days after their initial assessment, or 430 days since their initial COVID infection<br>Results:<br>1. 0 / 37 = 0%<br>2. 0 / 75 = 0%<br>3. 2/ 37 = 5.4%<br>4. 2 / 75 = 2.7%<br>5. 4 / 37 = 10.8%<br>6. 4 / 75 = 5.3%<br>7. 12 / 37 = 34.4%<br>8. 12 / 75 = 16%<br>Adjustments: None | | | |
| Rival, 2023<br>France<br><br>Cross-sectional<br>Industry funded: N<br><br>Sample:<br>hospitalized in ICU or non-ICU units<br><br>Risk of bias:<br>High | 64 (13)<br>45%<br><br>Race: NR<br><br>March 12 to April 15, 2020 (admitted between) | Comorbidity: NR<br><br>SES: NR | 40 hospitalized for COVID, with and without ICU admission<br><br>Population: COVID infected<br><br>Hospitalized: 100%<br><br>Confirmation method: NR<br><br>Severity:<br>•Admitted to ICU: 45.0%<br>•Post-ICU neuromyopathy: 61.1%<br>•Mean invasive mechanical ventilation: 15 days<br><br>Variant: Hospitalization during the first wave of the pandemic (i.e. Wild type) | MH service:<br>Accepting psychological follow-up (N=1: agoraphobia, 2: PTSD,3: doubt PTSD, 10: anxiety, 22: doubt PTSD, 38: PTSD). A psychological or psychiatric consultation was offered to patients with a positive Post-Traumatic Stress Checklist-5 (PCL-5) screening result (score of ≥33/80 or 4/4 positive clusters)<br><br>Ascertainment: NR |
| | **Findings**<br>Outcome: Accepting psychological follow-up / All undergoing PTSD assessments<br>Timing: Approximately 6 months post-discharge; mean time between COVID hospitalization and the follow-up consultation: 196±34 days<br>Results: 6 / 40 = 15%<br>Adjustments: None | | | |
| Umbrello, 2022<br>Italy<br><br>Cross-sectional | Median (IQR)<br>63 (57-71)<br>16%<br><br>Race: NR | Comorbidity:<br>Pre-existing: 52%<br>More than one: 23%<br><br>SES: | 79 Italian-speaking patients admitted to ICU (>24 h)<br><br>Population: COVID infected | MH service:<br>Met and unmet needs for psychologist based on information on health service utilization and need for care after ICU discharge |

| Author, Year, Country<br>Study design<br>Funding | Mean age (SD) yrs<br>Female, %<br>Race/ethnicity, %<br>Enrollment dates | Comorbidities<br>SES variables | COVID population description | MH service description |
|---|---|---|---|---|
| Industry funded: NR<br><br>Sample: ICU admissions<br><br>Risk of bias: High | March 5 to April 30, 2020 | Employment status<br>Active worker: 42.0%<br>Retired: 54.0%<br><br>Monthly income (€)-<br>≤ 1500: 54.0%<br>> 1500: 46.0%<br><br>Education (International Standard Classification of Education - ISCED):<br>0–2: 34%<br>> 2: 65% | Hospitalized: 100%<br><br>Confirmation method: NR<br><br>Severity:<br>•Required invasive mechanical ventilation: 87.3%<br>•Non-invasive ventilation with PEEP ≥ 5 cmH20: 12.6%<br>SOFA score on ICU admission, median (IQR): 6 (4-7) | Ascertainment:<br>Self-reported in interviews performed on the telephone, by an experienced ICU physician previously involved in patient clinical management |
| | **Findings**<br>Outcome: Need for psychologist / All COVD ICU survivors<br>Timing: 6 months after ICU discharge (time from symptoms to hospital admission, median [IQR]: 7 [5–10] days)<br>Results: 11 / 79 = 13.9%<br>Adjustments: None | | | |
| Tager, 2023<br>Argentina<br><br>Cross-sectional<br>Industry funded: NR<br><br>Sample: occupational<br><br>Risk of bias: High | 47.8 (11.8)<br>64.2%<br><br>Race:<br>All recruited from Spanish-speaking social network (all Latin American countries) | Comorbidity: NR<br><br>SES:<br>All healthcare professionals<br>Physicians: 67.5%<br>Nurses: 11.5% | 4673 Latin-American health professionals with COVID; some reported that the symptoms had not lasted more than one month since COVID episode<br><br>Population: COVID infected<br><br>Hospitalized: 11.7%<br><br>Confirmation method: PCR 100% | MH service:<br>Required new consultation for psychotherapy<br><br>Ascertainment:<br>Self-report; based on questionnaire response |
| | **Findings**<br>Outcome: Requirement for psychotherapy / All patients with COVID<br>Timing: At least 1 month after initial COVID infection<br>Results: 672 / 4673 = 14.4%<br>Adjustments: None | | | |
| Van Wambeke, 2023<br>France<br><br>Cross-sectional<br>Industry funded: N | 49.6 (11.2)<br>62.2%<br><br>Race: NR | Comorbidity:<br>Only six subjects (14%) were fully healthy, i.e., without comorbidity before<br><br>SES: NR | 45 COVID patients receiving consultations with study team, including follow-up through the first and second phone call. | MH service:<br>Psychological support therapy attendance<br><br>Patients contacted the post-COVID coordination team and received |

| Author, Year, Country<br>Study design<br>Funding | Mean age (SD) yrs<br>Female, %<br>Race/ethnicity, %<br>Enrollment dates | Comorbidities<br>SES variables | COVID population description | MH service description |
|---|---|---|---|---|
| <br>Sample: community<br><br>Risk of bias: High | September 2021 to March 2022 | | Population: post-COVID with symptoms<br><br>Hospitalized: 0%<br><br>Confirmation method: NR | phone consultations with an advanced practice nurse (45-65 minutes), who proposed therapeutic follow-up focused on patients' main symptoms. Outcome reported as those who had the opportunity to benefit from the psychological support therapy<br><br>Ascertainment: Telephone interview |
| | **Findings**<br>Outcome: Those attending psychological support therapy among those with COVID who contacted coordination team for assessment (attended two telephone consultations)<br>Timing: COVID occurrence and the first phone call delay: 15.1 ± 7.8 months; COVID occurrence and the second phone call 22.6 ± 7.7 months (last follow-up)<br>Results: 11 / 45 = 24.4%<br>Adjustments: None | | | |
| Wander, 2023<br>USA<br><br>Cross-sectional<br>Industry funded: N<br><br>Sample: health database<br><br>Risk of bias: High | Total with COVID (n=388980)<br>61.4 (16.1)<br>12.7%<br><br>Race:<br>American Indian or Alaska Native: 0.8%<br>Asian: 1.4%<br>Black or African American: 20.7%<br>Native Hawaiian or Pacific Islander: 1%<br>White: 67.8%<br><br>Ethnicity:<br>Hispanic or Latino: 9.3%<br>Not Hispanic or Latino: 84.7%<br><br>U09.9 Encounters (n=18587)<br>61.4 (NR) | Comorbidity:<br>All COVID<br>CCI 0: 39.6%<br>CCI 1: 20.3%<br>CCI 2: 14.8%<br>CCI 3: 8.6%<br>CCI 4: 5.9%<br>CCI 5-6: 6.4%<br>CCI 7-8: 3.0%<br>CCI >=9: 1.5%<br><br>PCC<br>CCI 0: 33.7%<br>CCI 1: 20.9%<br>CCI 2: 15.4%<br>CCI 3: 9.8%<br>CCI 4: 6.9%<br>CCI 5-6: 7.7%<br>CCI 7-8: 3.8%<br>CCI >=9: 1.9% | 388980 with COVID in the national US Veterans Affairs (VA) health care system<br><br>18587 (4.8%) documentation of ICD-10 code U09.9 in the VA health care system<br><br>Population: 18587 PCC (ICD) 100%<br><br>Hospitalized:<br>All with COVID 10.3%<br>PCC 24.3%<br><br>Confirmation method: PCR 100%<br><br>Severity: Mechanical ventilation<br>All with COVID 1%<br>PCC 2.4%<br><br>Vaccination:<br>Primary | MH service:<br>MH clinic encounters<br><br>Ascertainment:<br>Identified this code in records for all outpatient and inpatient encounters in the VA health care system using the Corporate Data Warehouse. To get the most complete ascertainment of U09.9 documentation possible, we also identified whether this code was documented in 2 non-VA health care sources<br><br>Previous MH:<br>Depression at baseline:<br>All with COVID: 35.9%<br>PCC: 37.8%<br><br>PTSD at baseline:<br>All with COVID: 27%<br>PCC: 27.1% |

| Author, Year, Country<br>Study design<br>Funding | Mean age (SD) yrs<br>Female, %<br>Race/ethnicity, %<br>Enrollment dates | Comorbidities<br>SES variables | COVID population description | MH service description |
|---|---|---|---|---|
| | 13%<br><br>Race:<br>American Indian or Alaska Native: 1%<br>Asian: 1%<br>Black or African American: 14.8%<br>Native Hawaiian or Pacific Islander: 1.1%<br>White: 72.9%<br><br>Ethnicity:<br>Hispanic or Latino: 14.5%<br>Not Hispanic or Latino: 79.8%<br><br>October 1, 2021 to January 31, 2023 | SES: NR | All with COVID 34.6%<br>PCC 38.6%<br><br>Booster<br>All with COVID 35.5%<br>PCC 30.9%<br><br>Number of acute symptoms at presentation with acute infection:<br><br>All with COVID<br>0: (44.9%<br>1-2: 23%<br>3-4: 15.4%<br>≥5: 16.6%<br><br>PCC<br>0: 39.4%<br>1-2: 24%<br>3-4: 17.4%<br>≥5: 19.1% | |
| | **Findings**<br>Outcome:<br>MH U09.9 clinic encounters<br>1. Among those with U09.9 documentation<br>2. Among all VA patients COVID positive<br>Timing: clinical notes were reviewed up to 6 months after acute COVID infection, and only new-onset symptoms present for 30 days or more after acute COVID infection were captured in accordance with the Centers for Disease Control and Prevention definition of post–COVID conditions<br>Results:<br>1. 804 / 35875 = 2.2%<br>2. 804 / 388980 = 0.2%<br>Adjustments: None | | | |

### Supplementary Figure 1: Labour Force by Risk of Bias

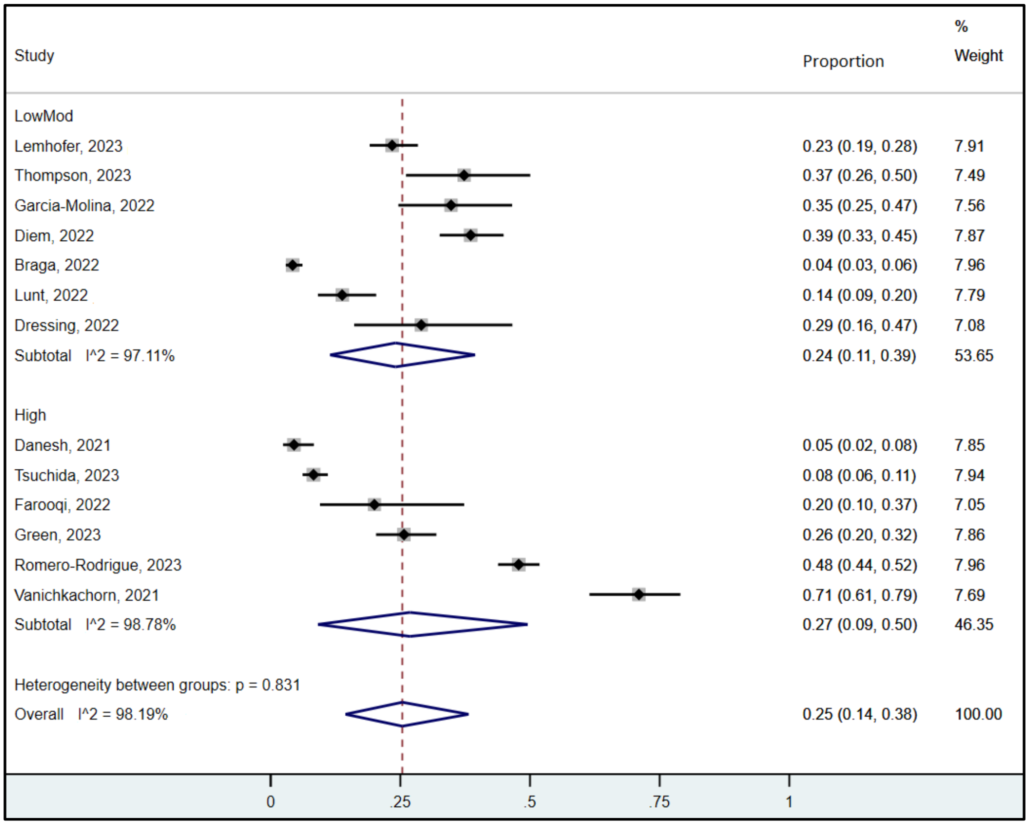


### Supplementary Figure 2: Labour Force Long-Term Timing by Risk of Bias

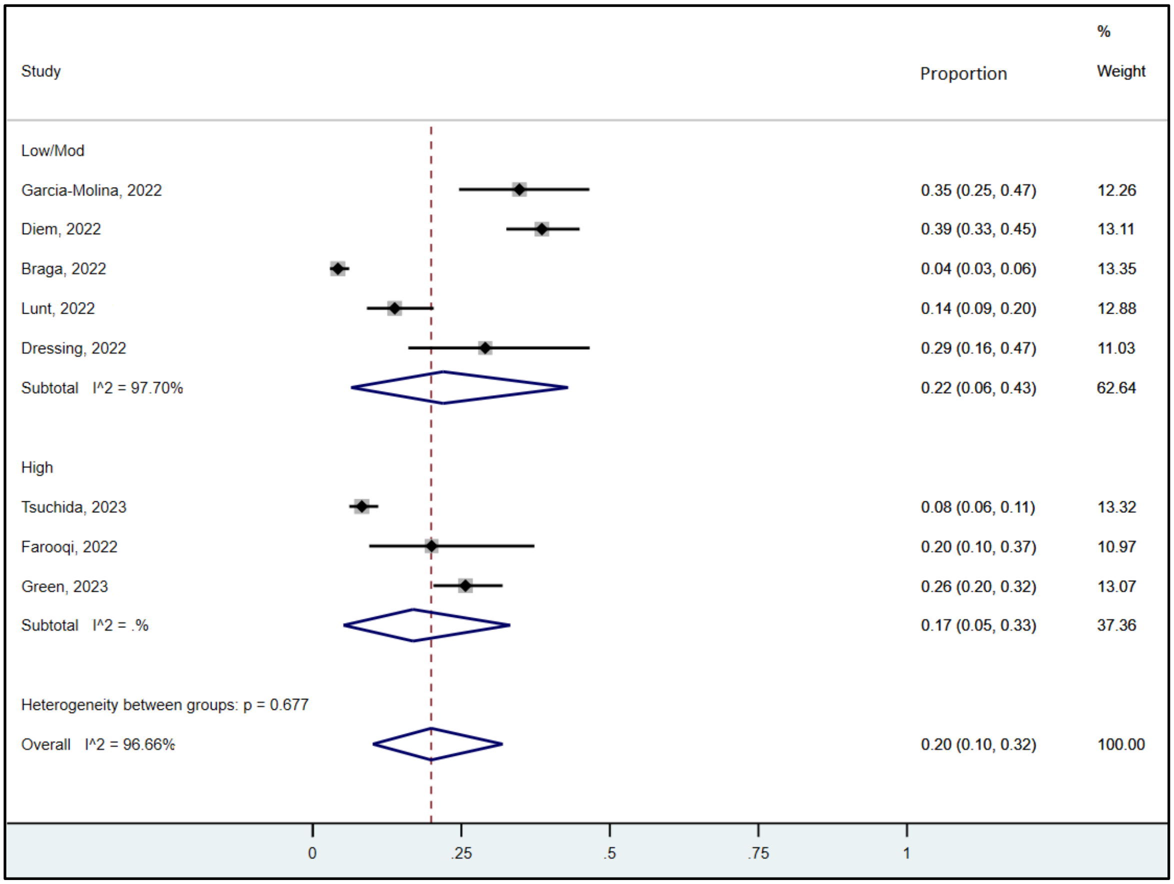

### Supplementary Figure 3: Labour Force MH Symptoms Alone vs. MH and Other Symptoms

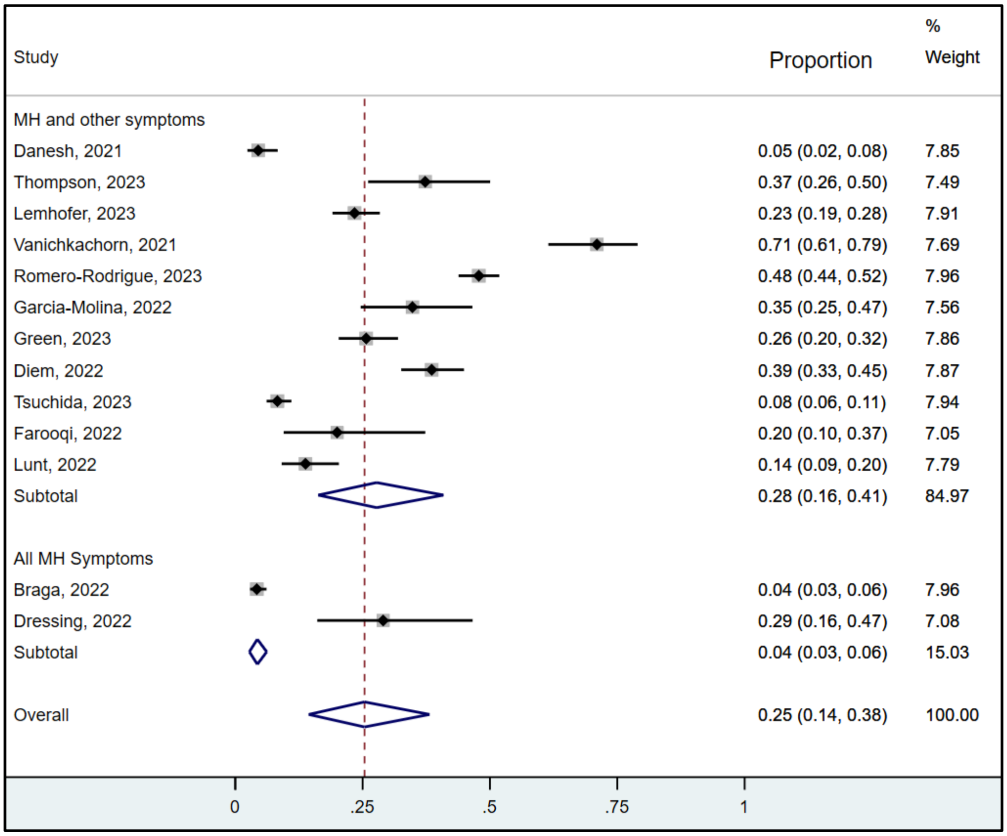


### Suppementary Figure 4: Labour Force by Narrow vs. Broad Spectrum of Symptoms

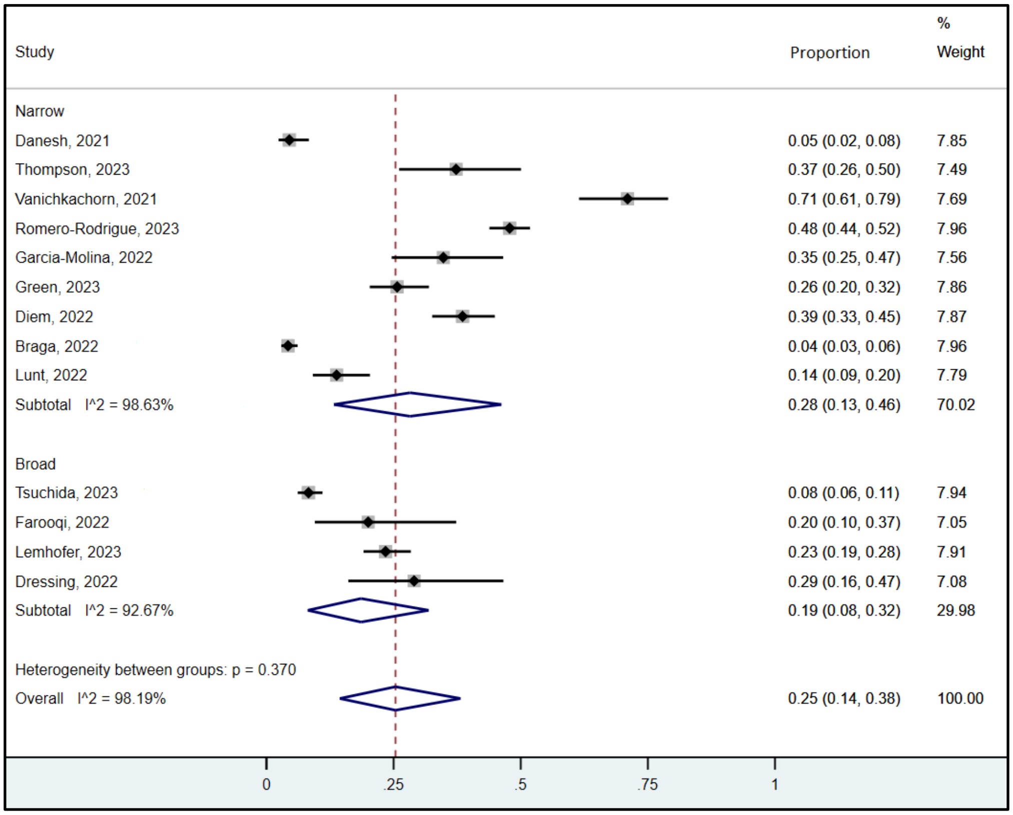

### Supplementary Figure 5: Mental health service, use by type of professional

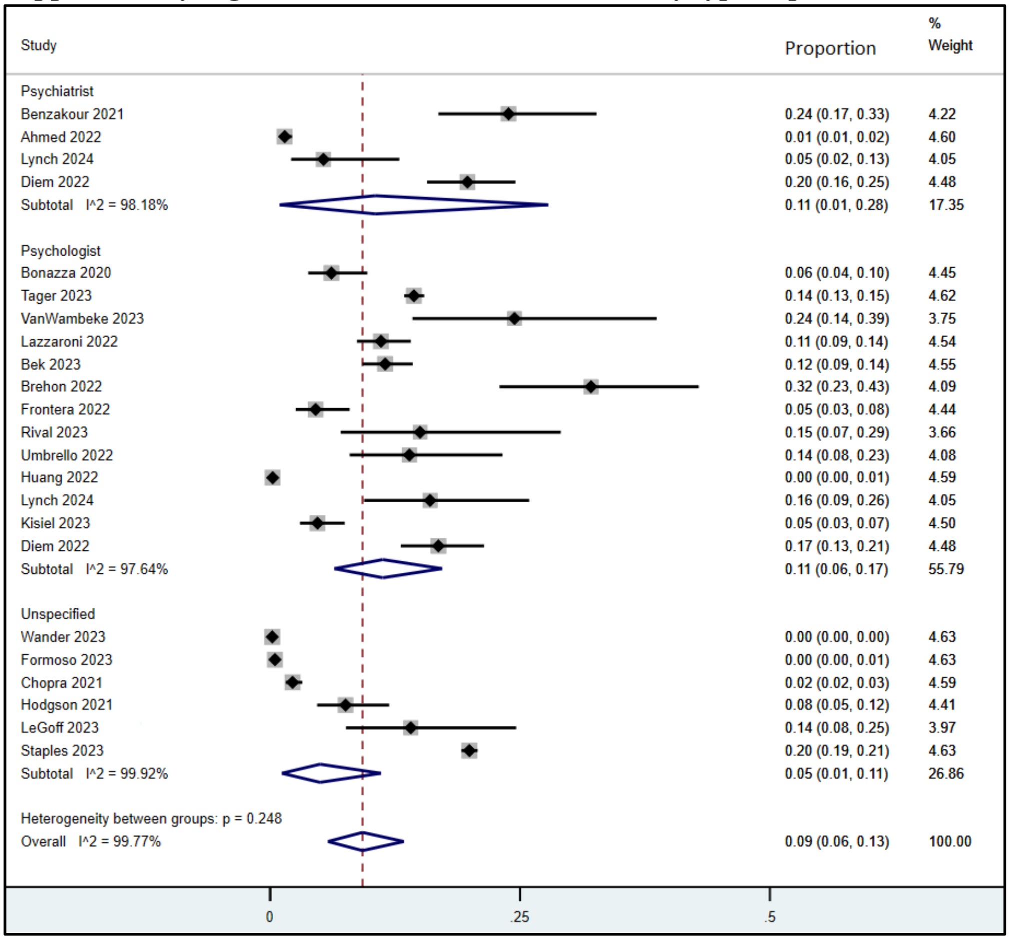

**Supplementary Figure 6: Mental Health Service Use by Severity**

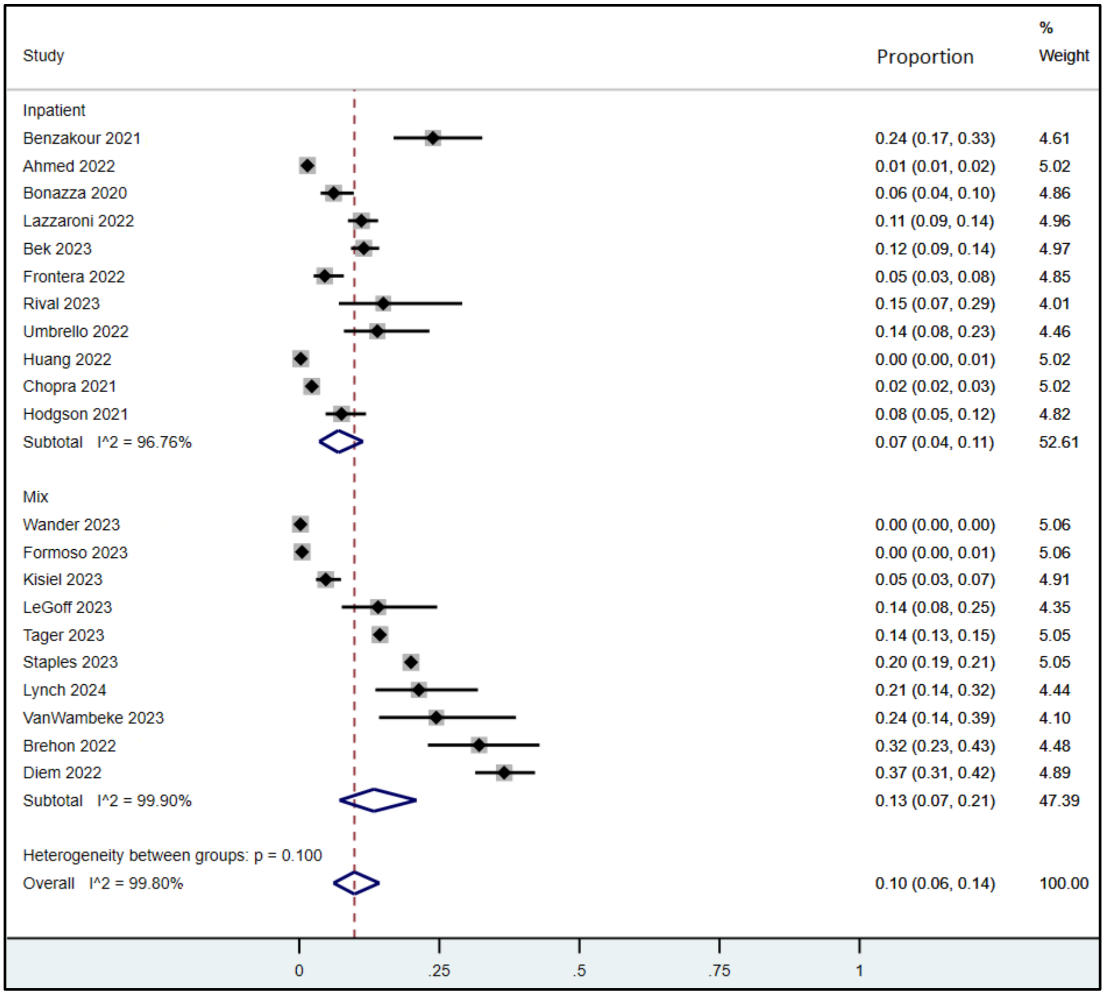

**Supplementary Figure 7: Mental Health Service Use by Severity and Variant**

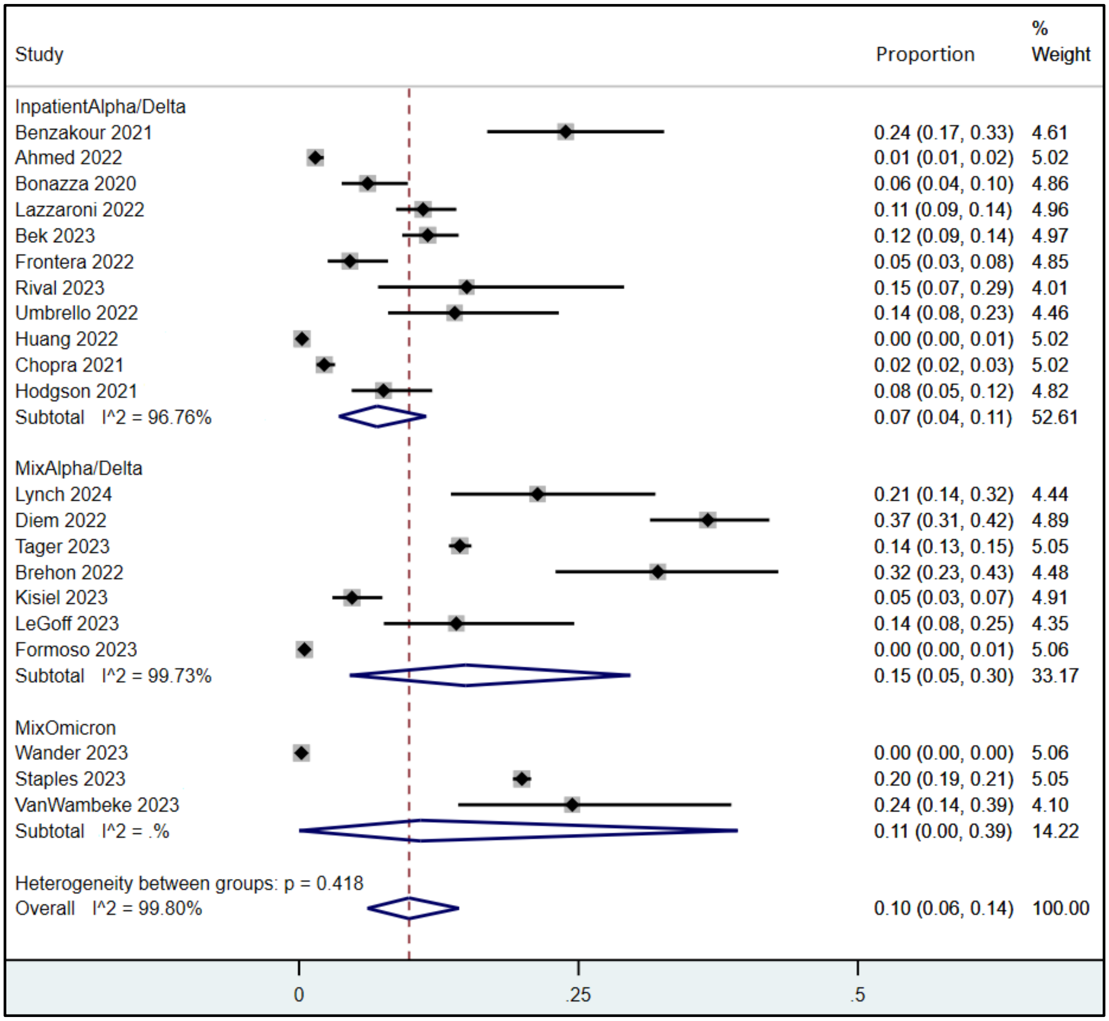

**Supplementary Figure 8: Mental Health Service Use by Risk of Bias**

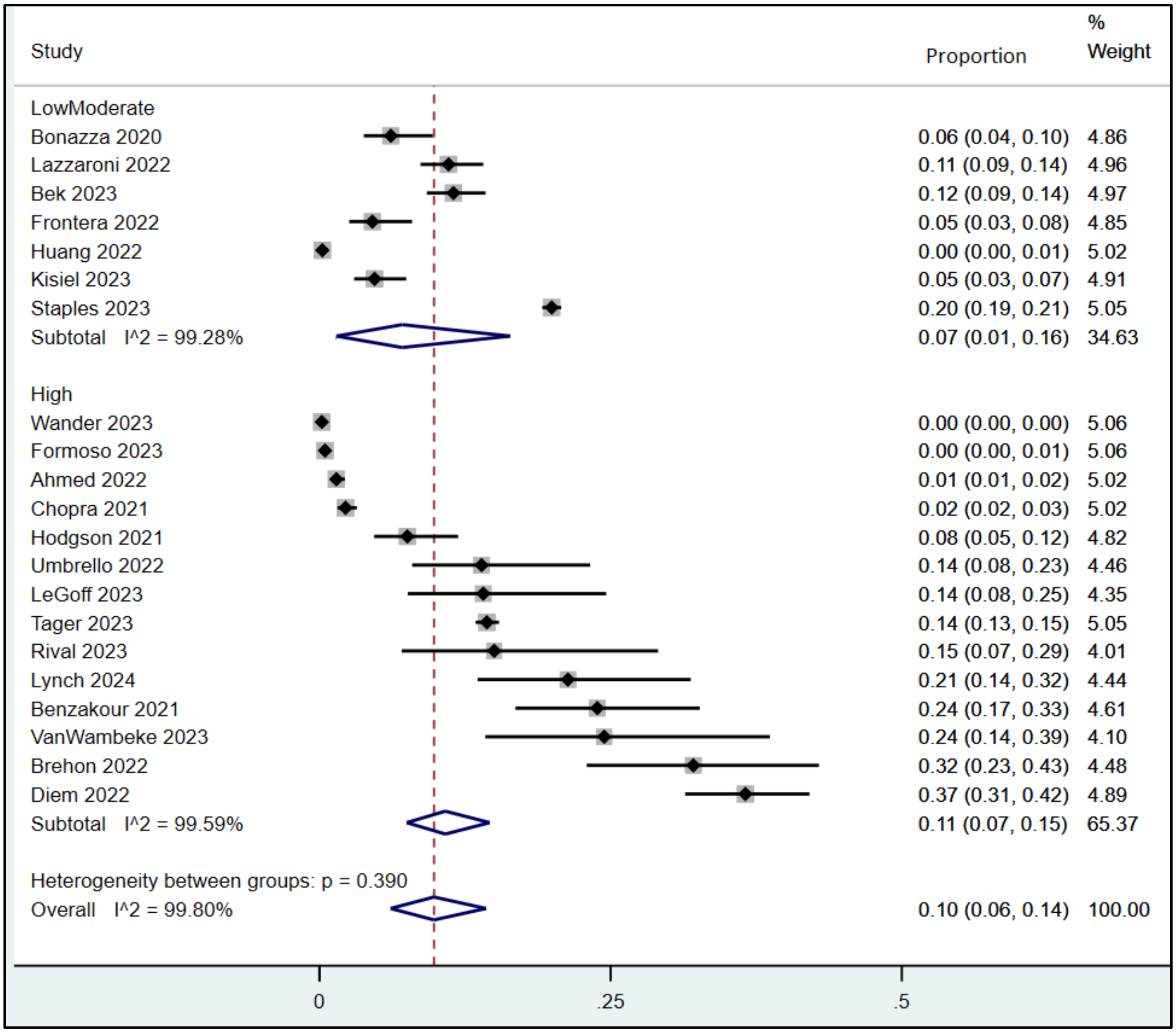
